# The role of COVID-19 vaccines in preventing post COVID-19 thromboembolic and cardiovascular complications: a multinational cohort study

**DOI:** 10.1101/2023.06.28.23291997

**Authors:** Núria Mercadé-Besora, Xintong Li, Raivo Kolde, Nhung TH Trinh, Maria T. Sanchez-Santos, Wai Yi Man, Elena Roel, Carlen Reyes, Antonella Delmestri, Hedvig ME Nordeng, Anneli Uusküla, Talita Duarte-Salles, Clara Prats, Daniel Prieto-Alhambra, Annika M Jödicke, Martí Català

**Author notes:** MC and AMJ share senior authorship. Corresponding author: Prof. Daniel Prieto-Alhambra Botnar Research Centre, Windmill Road, OX3 7LD, Oxford, United Kingdomlll⍰.

## Abstract

**Importance:** The overall effects of vaccination on the risk of cardiac, and venous and arterial thromboembolic complications following COVID-19 remain unclear.

**Objective:** We studied the association between COVID-19 vaccination and the risk of acute and subacute COVID-19 cardiac and thromboembolic complications.

**Design:** Multinational staggered cohort study, based on national vaccination campaign rollouts.

**Setting:** Network study using electronic health records from primary care records from the UK, primary care data linked to hospital data from Spain, and national insurance claims from Estonia.

**Participants:** All adults with a prior medical history of ≥180 days, with no history of COVID-19 or previous COVID-19 vaccination at the beginning of vaccine rollout were eligible.

**Exposure:** Vaccination status was used as a time-varying exposure. Vaccinated individuals were classified by vaccine brand according to the first dose received.

**Main Outcomes:** Post COVID-19 complications including myocarditis, pericarditis, arrhythmia, heart failure (HF), venous (VTE) and arterial thromboembolism (ATE) up to 1 year after SARS-CoV-2 infection.

**Measures:** Propensity Score overlap weighting and empirical calibration based on negative control outcomes were used to minimise bias due to observed and unobserved confounding, respectively. Fine-Gray models were fitted to estimate sub-distribution Hazard Ratios (sHR) for each outcome according to vaccination status. Random effect meta-analyses were conducted across staggered cohorts and databases.

**Results:** Overall, 10.17 million vaccinated and 10.39 million unvaccinated people were included. Vaccination was consistently associated with reduced risks of acute (30-day) and subacute post COVID-19 VTE and HF: e.g., meta-analytic sHR 0.34 (95%CI, 0.27-0.44) and 0.59 (0.50-0.70) respectively for 0-30 days, sHR 0.58 (0.48 - 0.69) and 0.71 (0.59 - 0.85) respectively for 90-180 days post COVID-19. Additionally, reduced risks of ATE, myocarditis/pericarditis and arrhythmia were seen, but mostly in the acute phase (0-30 days post COVID-19).

**Conclusions:** COVID-19 vaccination reduced the risk of post COVID-19 complications, including cardiac and thromboembolic outcomes. These effects were more pronounced for acute (1-month) post COVID-19 outcomes, consistent with known reductions in disease severity following breakthrough vs unvaccinated SARS-CoV-2 infection.

**Relevance:** These findings highlight the importance of COVID-19 vaccination to prevent cardiovascular outcomes after COVID-19, beyond respiratory disease.

**Key Points:** *Question:* What is the impact of COVID-19 vaccination to prevent cardiac complications and thromboembolic events following a SARS-CoV-2 infection?

*Findings:* Results from this multinational cohort study showed that COVID-19 vaccination reduced risk for acute and subacute COVID-19 heart failure, as well as venous and arterial thromboembolic events following SARS-CoV-2 infection.

*Meaning:* These findings highlight yet another benefit of vaccination against COVID-19, and support the recommendations for COVID-19 vaccination even in people at high cardiovascular risk.

## Introduction

COVID-19 vaccines were approved under emergency authorisation as from December 2020. They showed high effectiveness against SARS-CoV-2 infection, COVID-19 related hospitalisation and death (1–5). Nonetheless, concerns were raised after research findings suggested a potential association between adenovirus -based COVID-19 vaccines and unusual thromboembolic events (6–10). More recently, mRNA -based vaccines were found to be associated with risk of rare myocarditis events, especially among male teenagers and young adults (11–16).

On the other hand, it is well-known that SARS-CoV-2 infection can trigger cardiac and thromboembolic complications (17–19). Previous studies have showed that while slowly decreasing over time, risk for serious complications remain high for up to a year after infection (20,21). While acute and subacute cardiac and thromboembolic complications following SARS-CoV-19 are rare, they present a substantial burden to the affected patients. Given the global extension of the pandemic, the absolute number of cases could become substantial.

Recent studies suggest that COVID-19 vaccination could potentially protect against cardiac and thromboembolic complications attributable to COVID-19 (22–24). However, most of those focussed on relatively short-term complications (up to 30 (22) and 180 (23) days after infection) and were conducted in specific populations (ambulatory infections (22) and US Veterans (23)).

Evidence is still scarce to whether the additive effect of COVID-19 vaccines protecting 1) against SARS-CoV-2 infection and 2) reducing post COVID-19 cardiac and thromboembolic outcomes, outweighs any risks of these complications potentially associated with vaccination.

We leveraged large representative data sources from 3 European countries to assess the overall effect of COVID-19 vaccines on the risk of acute and subacute post COVID-19 complications including venous thromboembolism (VTE), arterial thromboembolism (ATE), and cardiac events. Additionally, we studied the comparative effects of ChAdOx1 (adenovirus -based) vs BNT162b2 (mRNA -based) on the risk of these same outcomes.

## Methods

### Data Sources

We used four routinely-collected population-based healthcare datasets from three European countries: the UK, Spain, and Estonia.

For the UK, we utilized data from two primary care databases, namely Clinical Practice Research Datalink, CPRD AURUM (25) and CPRD GOLD (26), which currently cover 3.1 million and 13 million active participants, respectively. Additionally, Spanish data was provided by the Information System for Research in Primary Care (SIDIAP) (27)which encompasses primary care records from around 75% of the population in the region of Catalonia, and it was linked to hospital admissions data (Conjunt Mínim Bàsic de Dades d’Alta Hospitalària). Finally, the CORIVA cohort was retrieved from the Estonian national health insurance claims database, covering around 440 thousand participants consisting of a random sample of the population plus all COVID-19 cases identified during the first year of the pandemic. CORIVA was linked to death registry and all COVID-19 testing from the national health information system.

Databases included socio-demographic information, diagnoses, measurements, prescriptions, and secondary care referrals. Further, all databases were linked to vaccine registries, including records of all administered vaccines from all healthcare settings. Data availability for CPRD GOLD ended in 12/2021, AURUM in 01/2022, SIDIAP in 06/2022, and CORIVA in 12/2022.

All four databases were mapped to the Observational Medical Outcomes Partnership (OMOP) Common Data Model (CDM) (28) to facilitate federated analytics.

### Multinational network staggered cohort study: Study design and participants

The study considered vaccination as a time-varying exposure using a staggered cohort design. Vaccine rollout phases were grouped in four different stages, each one with country-specific dates and inclusion criteria.

Source population comprised all adults registered in the respective database for at least 180 days at the start of the study (04/01/2021 for CPRD GOLD and AURUM, 20/02/2021 for SIDIAP, and 28/01/2021 for CORIVA). Subsequently, each staggered cohort included individuals eligible for vaccination during the enrolment period who were not previously vaccinated against or infected with SARS-CoV-2. Cohort 1 enrolled elderly population, whereas cohort 2 comprised individuals at risk for severe COVID-19. Cohort 3 comprised people aged ≥40, and cohort 4 included predominantly younger people (≥ 18).

People receiving a first vaccine dose within enrolment period were allocated to the vaccinated cohort (VC), and their index date was the date of the first vaccine dose. Individuals who did not receive a vaccine dose comprised the unvaccinated cohort (UVC), and their index date was sorted within enrolment period, based on the distribution of index dates in the VC. Individuals infected with SARS-CoV-2 before index date were excluded.

Follow-up of individuals was until the earliest of: end of available data, death, change in exposure status (first vaccine dose for UVC), or outcome of interest.

Supplementary Figures S1-2 and Table S1 provide more details on cohort definitions, enrolment periods and inclusion criteria for each country. The proposed staggered cohort study design was used in previous work and is published in detail elsewhere (29).

### COVID-19 vaccination

All vaccines approved within study periods from 01/2021 to 07/2021, namely ChAdOx1 [Oxford/AstraZeneca], BNT162b2 [BioNTech/Pfizer], Ad26.COV2.S [Janssen], and mRNA-1273 [Moderna] were included for this study.

### Post COVID-19 outcomes of interest

Outcomes of interest in the study were defined as a SARS-CoV-2 infection followed by a record of a pre-defined thromboembolic or cardiac event of interest within a year after infection, in those individuals with no record of the same clinical event in the 6 months before COVID-19. Outcome date was set as the corresponding SARS-CoV-2 infection date. COVID-19 disease was identified from either a positive SARS-CoV-2 test (PCR/ antigen), or a clinical COVID-19 diagnosis, with no record of COVID-19 in the previous 6 weeks. This wash-out period was imposed to identify “new” infections and exclude re-recordings of the same COVID-19 episode.

Events of interest after COVID-19 were selected based on previous studies (21,22,30). These were ischemic stroke (IS), haemorrhagic stroke (HS), transient ischemic attack (TIA), ventricular arrhythmia or cardiac arrest (VACA), myocarditis or pericarditis (MP), myocardial infarction (MI), heart failure (HF), pulmonary embolism (PE) and deep vein thrombosis (DVT). We used two composite outcomes: 1) VTE, as an aggregate of PE and DVT, and 2) ATE, as a composite of IS, TIA, and MI. To avoid re-recording of the same complication we imposed a wash-out period of 90 days between records. Phenotypes for these complications were based on previously published studies (31,32).

Further, outcomes were ascertained in four different time periods following SARS-CoV-2 infection: the first period described the acute infection phase, i.e. 0 to 30 days after COVID-19, whereas the later periods, i.e. 31 to 90 days, 91 to 180 days, and 181 to 365 days illustrate the subacute phase (Figure 1).

**Figure 1:**
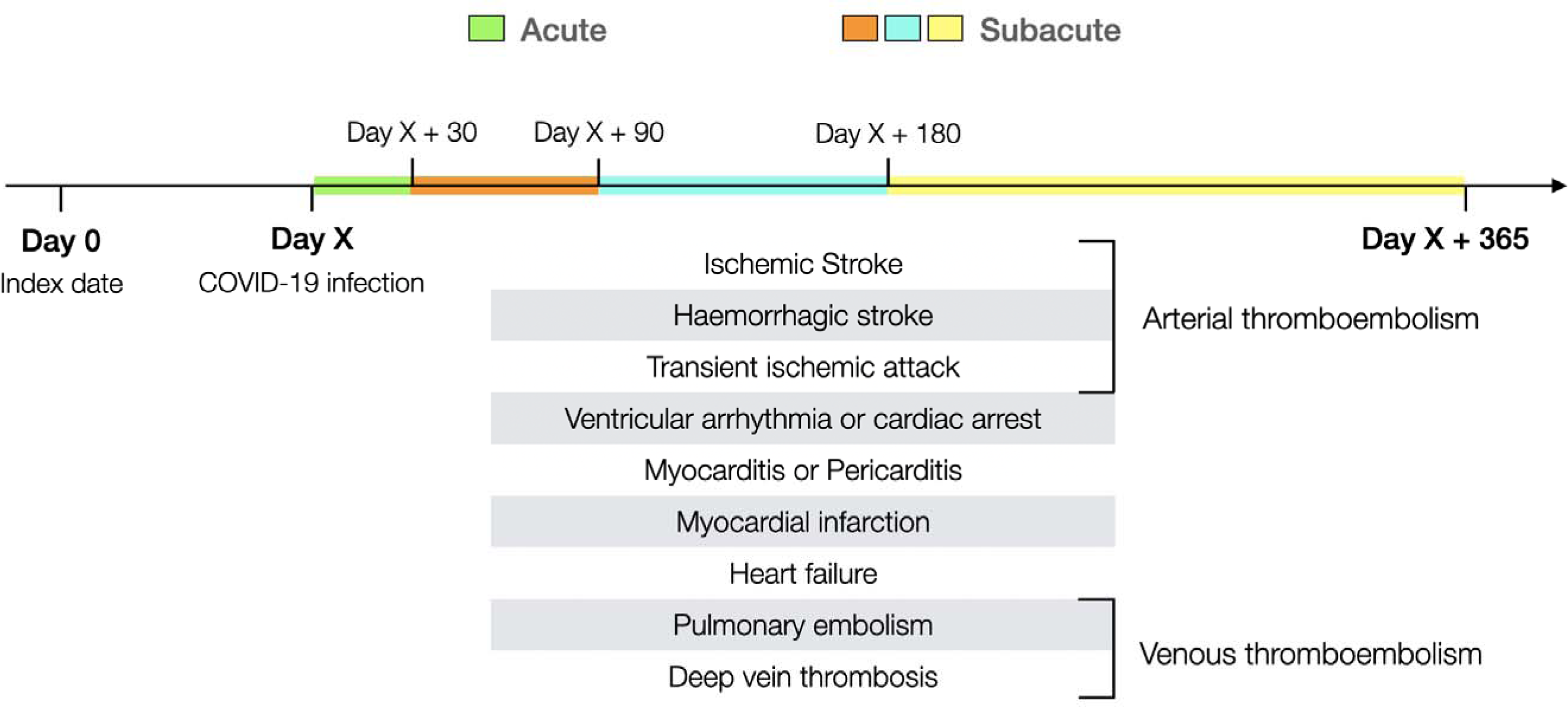
Study outcome design. Study outcomes of interest are defined as a COVID-19 infection followed by one of the complications in the figure, within year after infection. Outcomes were ascertained in four different time windows after SARS-CoV-2 infection: 0-30 days (namely the acute phase), 31-90 days, 91-180 days, and 181-365 days (these last three comprise the subacute phase).

### Negative Control Outcomes

Negative control outcomes (NCO) refer to outcomes which investigators believe they are not associated (causal relation) with the exposure, and share the same causal or bias structure with the exposure and outcome of interest (33). The expected occurrence of events should be similar in the compared cohorts, otherwise it may indicate a source of unobserved bias. The study used 43 different NCO from previous work assessing vaccine effectiveness (34,35) to detect residual confounding.

### Statistical analysis

#### Federated network analyses

A template for an analytical script was developed, and subsequently tailored to include the country-specific aspects (e.g. dates, priority groups) for the vaccination rollout. The specific R code was then executed locally in each database. Only aggregated data was shared and person counts <5 were clouded.

#### Propensity Score Weighting

Large-Scale Propensity Scores (PS) were calculated to estimate the likelihood of a person to receive the vaccine based on their demographic and health-related (e.g. conditions, medication) characteristics prior to the index date. PS were then used to minimise observed confounding by creating a weighted population (based on Overlap Weights (36)) in which individuals contributed with a different weight based on their PS and vaccination status. Variables included in the PS comprised age, sex, location, index date, prior observation time in the database, number of previous outpatient visits, and previous SARS-CoV-2 PCR/antigen tests. Regional vaccination, testing and COVID-19 incidence rates were also forced into PS equation for CPRD AURUM and GOLD (37) and SIDIAP (38). In addition, conditions and prescriptions within 0-30 days, 31-180 days, and 181-anytime (latter window only for conditions) before index date, with a prevalence of >0.5% and predictive of vaccination based on LASSO regression, were included to the PS model.

PS were estimated for each staggered cohort and analysis separately. We considered that covariate balance was achieved if Absolute Standardised Mean Differences (ASMD) were ≤0.1 after weighting (39). Baseline characteristics (e.g. age, sex, prior history, general health conditions) were reported.

#### Effect estimation

To account for the competing risk of death associated with COVID-19, Fine-and-Gray models (40) were utilized to calculate subdistribution Hazard Ratios (sHR). Subsequently, sHR and confidence intervals were empirically calibrated from NCO estimates (41) to account for unmeasured confounding.

For each outcome, treatment estimates from the four staggered studies were pooled using random effect meta-analysis (42), both separately for each database and across all four databases.

### Sensitivity analysis

Sensitivity analyses were used to test the effect of two approaches in the main analysis: first, we censored follow-up for vaccinated people at the time when they received their second vaccine dose. Second, we captured only the first post COVID-19 outcome within the year after infection. Supplementary Figure S3 depicts follow-up for the analyses.

Further, we compared vaccinated groups by vaccine brand, namely, BNT162b2 vs. ChAdOx1.

### Data and code availability

All analytic code for the study is available at (https://github.com/oxford-pharmacoepi/vaccineEffectOnPostCovidCardiacThromboembolicEvents). The repository also contains code lists for vaccines, COVID-19 tests and diagnoses, cardiac and thromboembolic events, NCO, and health conditions to prioritize patients for vaccination in each country. Analyses were conducted using R 4.2.3.

## Results

All aggregated results are available via a web application (https://dpa-pde-oxford.shinyapps.io/PostCovidComplications/).

We included over 10.17 million vaccinated individuals (1,618,395 from CPRD GOLD; 5,729,800 from CPRD AURUM; 2,744,821 from SIDIAP and 77,603 from CORIVA), and 10.39 million unvaccinated (1,640,371; 5,860,564; 2,588,518 and 302,267, respectively). Supplementary Figures S4-7 illustrate flowcharts in each database.

Adequate covariate balance was achieved after PS weighting in most studies: CORIVA (all cohorts) and SIDIAP (cohorts 1 and 4) did not contribute to ChAdOx1 sub-analysis due to sample size and covariate unbalance. ASMD results are accessible in the web application.

NCO analyses suggested residual bias despite PS weighting, with a majority of NCO associated positively with vaccination. Therefore, calibrated estimates are reported in this manuscript. Uncalibrated effect estimates and NCO analyses are available in the web interface.

### Population characteristics

Table 1 presents baseline characteristics for the weighted populations in CPRD AURUM, for illustrative purposes. Supplementary Tables S2-26 summarise baseline characteristics for weighted and unweighted populations for each database and comparison. Across databases and cohorts, populations followed similar patterns: cohort 1 represented the eldest population (around 80 years old) with predominant female population (60%). Age median lowered to 30-40 years in cohort 4, which was more balanced regarding sex. Overall, weighted populations showed adequate balance (ASMD < 0.1) in all baseline characteristics.

**Table 1:** Characteristics of weighted populations in CPRD AURUM, database, stratified by staggered cohort and exposure status. Exposure is any COVID-19 vaccine.

|  | Cohort 1 |  |  | Cohort 2 |  |  | Cohort 3 |  |  | Cohort 4 |  |  |
| --- | --- | --- | --- | --- | --- | --- | --- | --- | --- | --- | --- | --- |
|  | Unvaccinated | Vaccinated | ASMD | Unvaccinated | Vaccinated | ASMD | Unvaccinated | Vaccinated | ASMD | Unvaccinated | Vaccinated | ASMD |
| **N (individuals)** | 154,864 | 154,245 |  | 420,707 | 420,931 |  | 463,495 | 462,463 |  | 818,917 | 827,124 |  |
| **Age, median [Q25-Q75]** | 80 [76-84] | 80 [76-84] | 0.000 | 58 [44-67] | 58 [44-67] | 0.005 | 50 [41-58] | 52 [40-58] | 0.003 | 34 [26-42] | 34 [26-42] | 0.004 |
| **Sex: Female, N(%)** | 88,349 (57%) | 87,639 (57%) | 0.005 | 248,156 (59%) | 249,561 (59%) | 0.006 | 245,248 (53%) | 245,600 (53%) | 0.004 | 351,435 (43%) | 358,688 (43%) | 0.009 |
| **Years of prior history<sup>a</sup>, median [Q25-Q75]** | 24 [10-35] | 24 [10-36] | 0.006 | 18 [8-29] | 18 [8-29] | 0.003 | 14 [6-24] | 14 [7-24] | 0.008 | 8 [4-17] | 7 [3-18] | 0.001 |
| **Number visits, median [Q25-Q75]** | 10 [5-18] | 10 [6-17] |  | 8 [3-15] | 8 [5-14] |  | 4 [1-11] | 6 [3-11] |  | 2 [0-6] | 2 [0-6] |  |
| **Number pcrs, median [Q25-Q75]** | 0[0-0] | 0[0-0] |  | 0[0-0] | 0[0-0] |  | 0[0-0] | 0[0-0] |  | 0[0-0] | 0[0-0] |  |
| **Comorbidities<sup>b</sup>, N(%)** |  |  |  |  |  |  |  |  |  |  |  |  |
| Anxiety | 23,200 (15%) | 22,789 (15%) | 0.006 | 94,390 (22%) | 91,644 (22%) | 0.016 | 92,820 (20%) | 90,807 (20%) | 0.010 | 123,055 (15%) | 125,202 (15%) | 0.003 |
| Asthma | 16,978 (11%) | 16,663 (11%) | 0.005 | 95,770 (23%) | 94,550 (22%) | 0.007 | 79,642 (17%) | 78,266 (17%) | 0.007 | 63,687 (8%) | 61,472 (7%) | 0.013 |
| Chronic kidney disease | 36,149 (23%) | 36,046 (23%) | 0.001 | 28,181 (7%) | 29,756 (7%) | 0.015 | 10,283 (2%) | 10,577 (2%) | 0.005 | 3,840 (0%) | 3,572 (0%) | 0.006 |
| COPD | 13,385 (9%) | 13,181 (9%) | 0.003 | 17,447 (4%) | 17,999 (4%) | 0.006 | 6,062 (1%) | 5,754 (1%) | 0.006 | 1,901 (0%) | 1,918 (0%) | 0.000 |
| Dementia | 9,483 (6%) | 8,517 (6%) | 0.026 | 4,182 (1%) | 3,879 (1%) | 0.007 | 1,361 (0%) | 1,392 (0%) | 0.001 | 276 (0%) | 495 (0%) | 0.012 |
| Depressive disorder | 18,632 (12%) | 18,547 (12%) | 0.000 | 85,280 (20%) | 81,945 (19%) | 0.020 | 81,891 (18%) | 79,804 (17%) | 0.011 | 94,373 (12%) | 97,053 (12%) | 0.007 |
| Diabetes | 29,365 (19%) | 28,831 (19%) | 0.007 | 49,408 (12%) | 48,562 (12%) | 0.006 | 26,616 (6%) | 28,628 (6%) | 0.019 | 12,787 (2%) | 12,539 (2%) | 0.004 |
|  | Unvaccinated | Vaccinated | ASMD | Unvaccinated | Vaccinated | ASMD | Unvaccinated | Vaccinated | ASMD | Unvaccinated | Vaccinated | ASMD |
| GERD | 8,718 (6%) | 8,515 (6%) | 0.005 | 19,907 (5%) | 18,924 (4%) | 0.011 | 15,646 (3%) | 14,982 (3%) | 0.008 | 13,882 (2%) | 13,893 (2%) | 0.001 |
| Heart failure | 9,349 (6%) | 8,851 (6%) | 0.013 | 7,284 (2%) | 6,502 (2%) | 0.015 | 2,660 (1%) | 2,470 (1%) | 0.005 | 930 (0%) | 816 (0%) | 0.005 |
| Hypertension | 81,563 (53%) | 80,806 (52%) | 0.006 | 97,707 (23%) | 98,193 (23%) | 0.002 | 54,649 (12%) | 55,798 (12%) | 0.008 | 22,925 (3%) | 24,450 (3%) | 0.009 |
| Hypothyroidism | 15,125 (10%) | 15,098 (10%) | 0.001 | 25,579 (6%) | 25,962 (6%) | 0.004 | 17,162 (4%) | 17,580 (4%) | 0.005 | 12,427 (2%) | 12,641 (2%) | 0.001 |
| Malignant neoplastic disease | 33,467 (22%) | 33,024 (21%) | 0.005 | 30,194 (7%) | 35,085 (8%) | 0.043 | 14,815 (3%) | 14,140 (3%) | 0.008 | 6,447 (1%) | 5,766 (1%) | 0.011 |
| Myocardial infarction | 7,824 (5%) | 7,731 (5%) | 0.002 | 9,964 (2%) | 11,319 (3%) | 0.020 | 3,787 (1%) | 3,664 (1%) | 0.003 | 1,315 (0%) | 1,069 (0%) | 0.008 |
| Osteoporosis | 15,275 (10%) | 15,373 (10%) | 0.003 | 10,626 (3%) | 10,718 (3%) | 0.001 | 4,113 (1%) | 4,131 (1%) | 0.001 | 1,376 (0%) | 1,472 (0%) | 0.002 |
| Pneumonia | 8,573 (6%) | 7,621 (5%) | 0.027 | 11,355 (3%) | 10,691 (3%) | 0.010 | 6,651 (1%) | 6,545 (1%) | 0.002 | 5,144 (1%) | 5,151 (1%) | 0.001 |
| Rheumatoid arthritis | 3,066 (2%) | 3,092 (2%) | 0.002 | 6,198 (1%) | 6,570 (2%) | 0.007 | 2,355 (1%) | 3,111 (1%) | 0.021 | 1,201 (0%) | 859 (0%) | 0.012 |
| Stroke | 7,667 (5%) | 7,047 (5%) | 0.018 | 8,041 (2%) | 8,794 (2%) | 0.013 | 3,518 (1%) | 3,293 (1%) | 0.006 | 1,496 (0%) | 1,305 (0%) | 0.006 |
| Venous thromboembolism | 9,589 (6%) | 9,241 (6%) | 0.008 | 11,836 (3%) | 12,475 (3%) | 0.009 | 6,503 (1%) | 8,075 (2%) | 0.028 | 4,661 (1%) | 2,441 (0%) | 0.042 |

The 4 cohorts represent vaccine rollout periods.
<sup>a</sup>Calculated as the days of previous observation in the database before index date.
<sup>b</sup>Assessed anytime before index date. ASMD = Absolute standardized mean difference, COPD = Chronic obstructive pulmonary disease, GERD = Gastro-Esophageal reflux disease

### COVID-19 vaccination and post COVID-19 complications

Table 2 shows the incidence of post COVID-19 VTE, ATE and HF, the 3 most common post COVID-19 conditions among the studied. Outcome counts are presented separately for the 0-30, 31-90, 91-180, and 181-365 days after SARS-CoV-2 infection. Supplementary Tables S27-37 include all studied complications, also for the different sensitivity and sub-analyses. Similar incidence patterns were observed across all analyses: higher rates in the elderly populations (cohort 1) and decreasing frequency with time after infection in all cohorts.

**Table 2:**
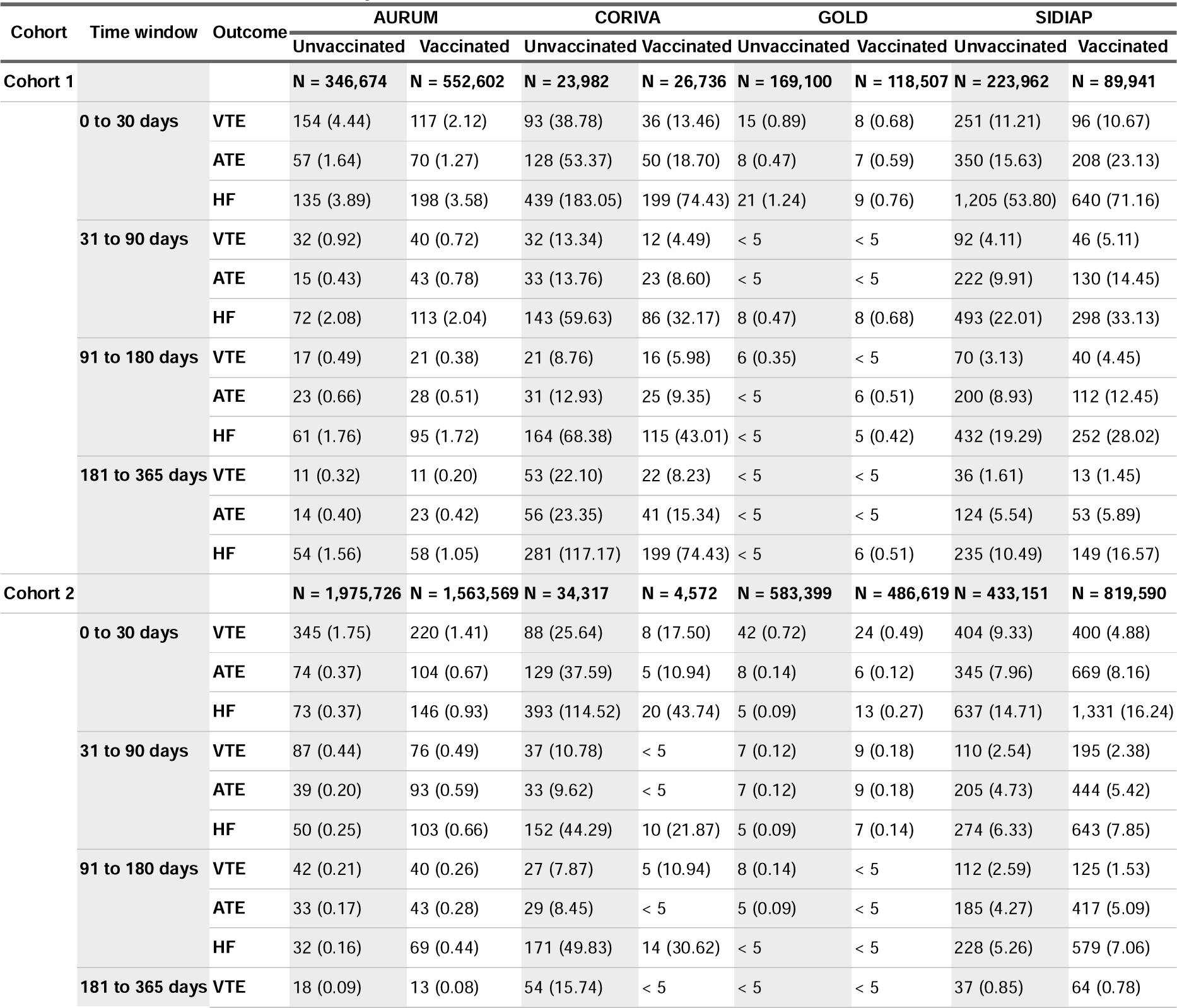
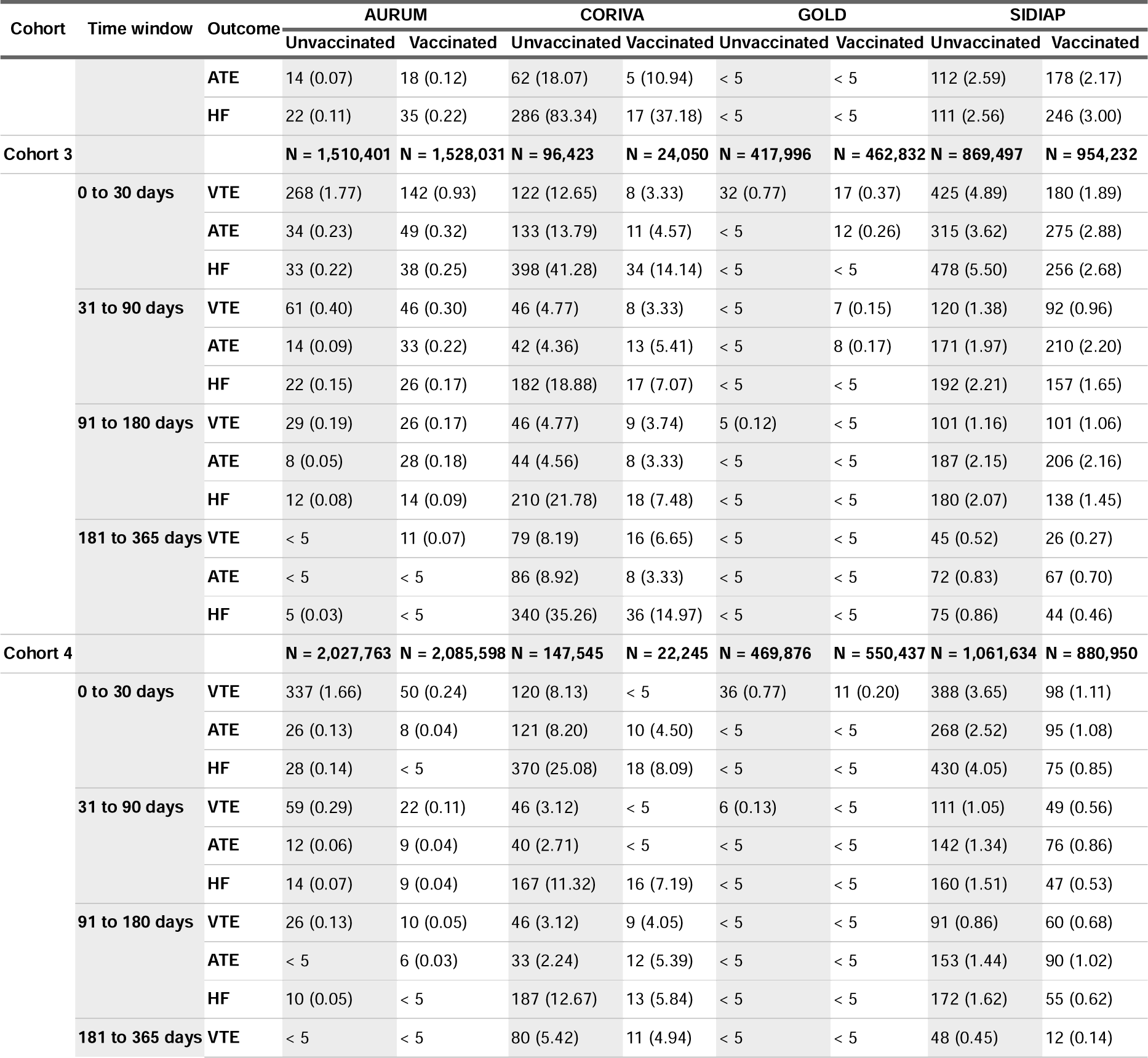
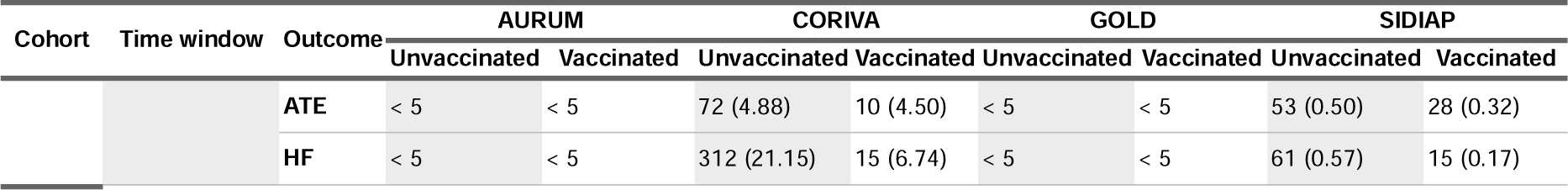
Number of records (and risk per 10,000 individuals) for acute and subacute COVID-19 cardiac and thromboembolic complications, across cohorts and databases for any COVID-19 vaccination.

Results from calibrated estimates pooled in meta-analysis across cohorts and databases are shown in Figure 2.

**Figure 2:**
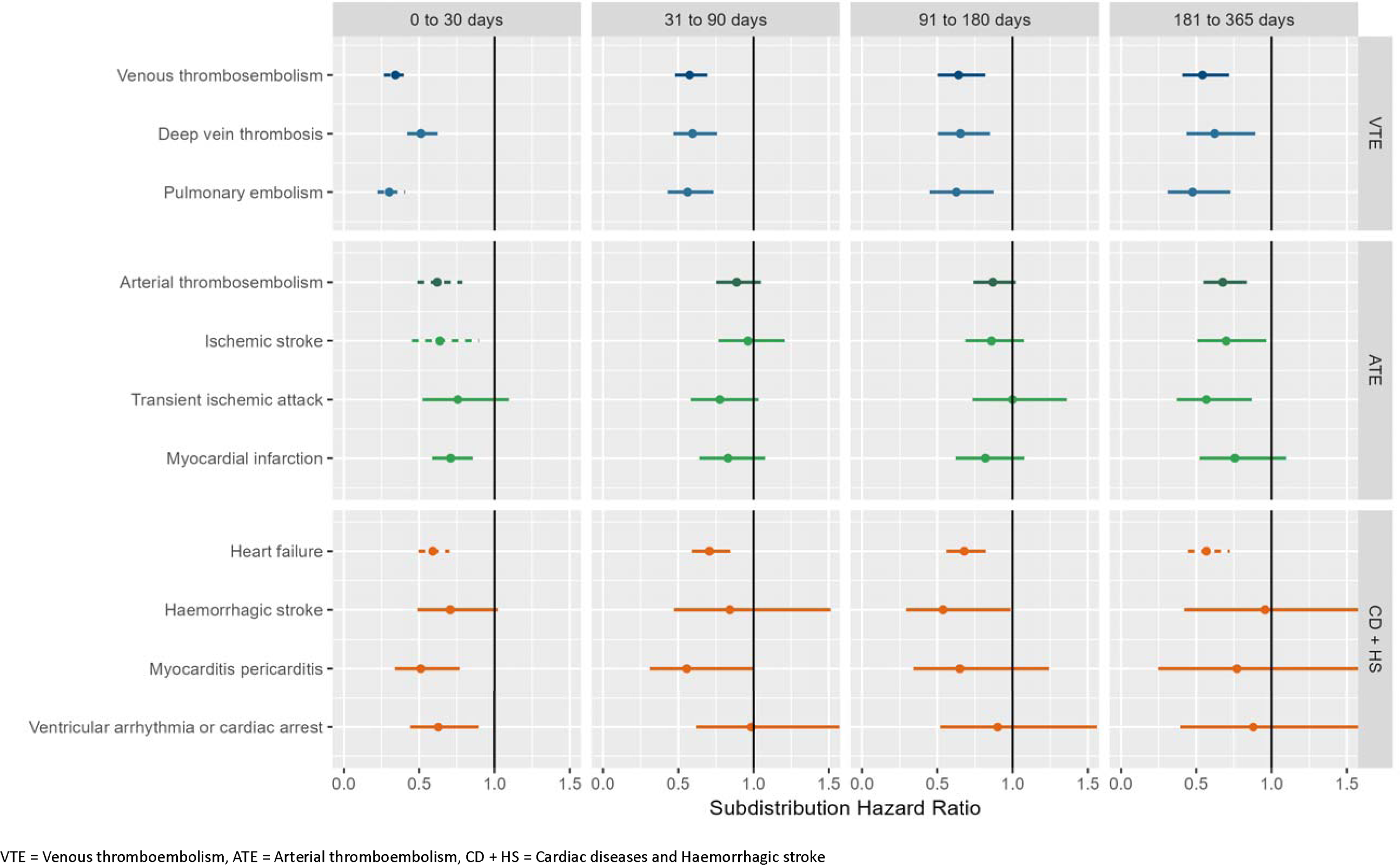
Forest plots for vaccine effectiveness (any COVID-19 vaccine), meta-analysis across cohorts and databases. Dashed line represents a level of heterogeneity I2 > 0.4.

Protective effects of vaccination are observed against acute post COVID-19 VTE, DVT, and PE: meta-analytic sHR of 0.34 (95% CI, 0.27-0.44); 0.51 (0.42-0.62); and 0.30 (0.22-0.41) respectively. The protective effects persisted for subacute VTE: sHR of 0.58 (0.48-0.69), 0.64 (0.50-0.82) and 0.54 (0.41-0.72) for 31-90, 91-180, and 181-365 days post COVID-19, respectively. Protection effects of vaccine against these outcomes were observed across databases and cohorts, see Supplementary Figures S16-24.

Similarly, the risk of ATE, IS and MI in the acute phase after infection was reduced in the vaccinated, sHR of 0.62 (0.49-0.77), 0.63 (0.45-0.90), and 0.71 (0.58-0.85) respectively. Protection persisted for subacute ATE and IS in the 181-365 days post COVID-19 (sHR of 0.68 (0.55-0.84) and 0.70 (0.51-0.96)), and no effect was seen in the 30-180 days window. Vaccination effect on post COVID-19 TIA was only seen in the 181-365 days, sHR of 0.56 (0.37-0.86). Supplementary Figures S25-33 show database- and cohort-specific estimates for ATE related complications.

Risk of post COVID-19 cardiac complications was reduced in vaccinated individuals. Meta-analytic estimates in the acute phase showed sHR of 0.58 (0.50-0.70) for HF, 0.51 (0.34-0.77) for MP, and 0.63 (0.44, 0.89) for VACA. Protective effects persisted for subacute post COVID-19 HF: sHR 0.71 (0.59-0.85) for 31-90 days, 0.68 (0.56-0.82) for 91-180 days, and 0.57 (0.44-0.72) for 181-365 days. For subacute MP, risk was only lowered in the 31-90 days post infection, sHR 0.55 (0.31-0.99), and no effect was seen for subacute VACA. Protection against post COVID-19 HS was only seen in the 91-180 days after infection, s, of 0.54 (0.29-0.99). Database- and cohort-specific results for these cardiac diseases are in Supplementary Figures S34-42.

Stratified analyses by vaccine brand showed similar associations, except for ChAdOx1 which was not associated with reduced VTE risk in the subacute window. Sensitivity analyses were consistent with main results, see Supplementary Figures S8-15.

Figure 3 shows the results of comparative effects of BNT162b2 vs ChAdOx1, based on UK data. Meta-analytic estimates favoured BNT162b2 (sHR of 0.66 (0.46, 0.93)) for VTE in the 0-30 days after infection, but no differences were seen for subacute VTE or for any of the other outcomes. Results from sensitivity analyses, database- and cohort-specific estimates were in line with the main approach, see Supplementary Figures S43-53.

**Figure 3:**
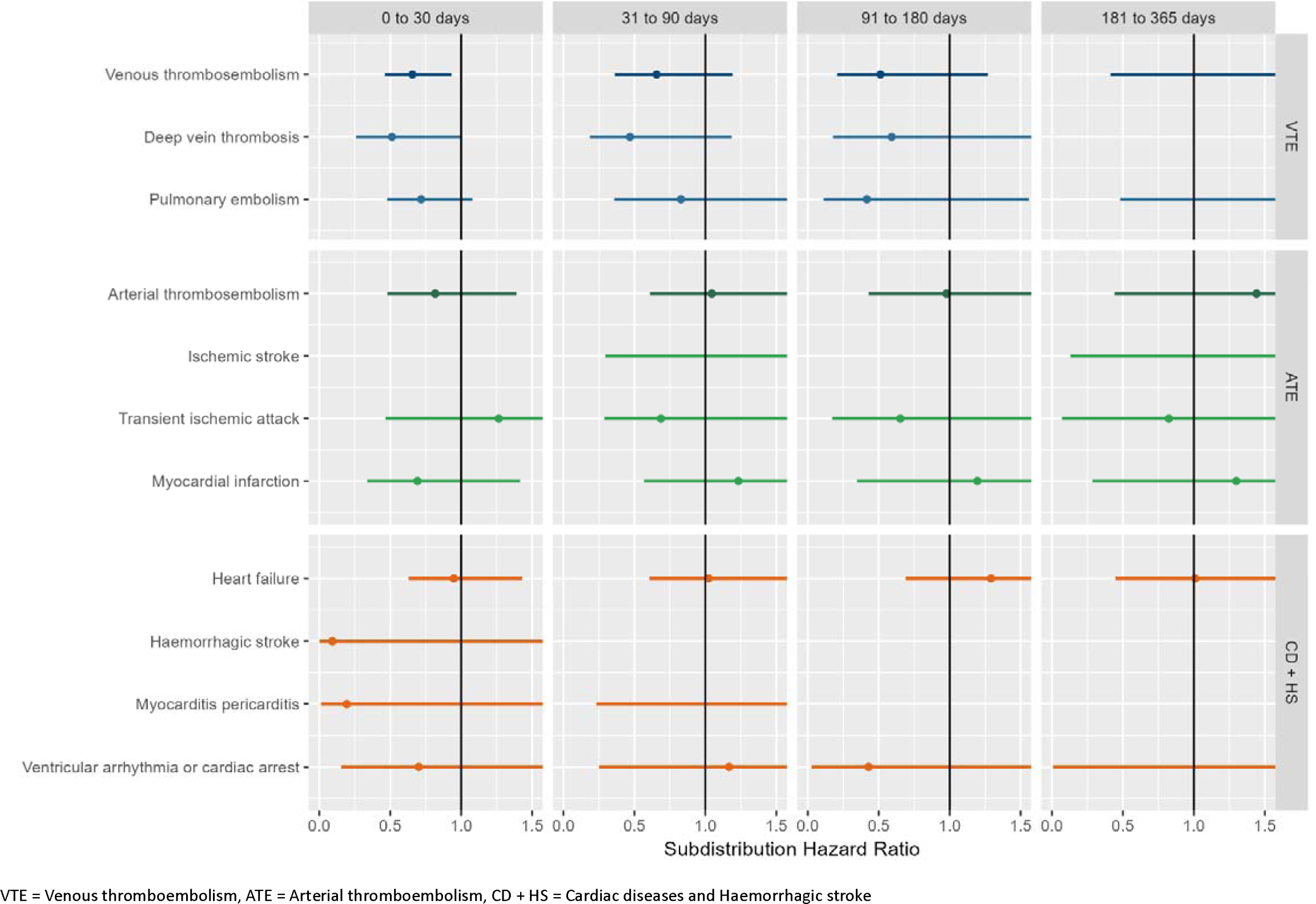
Forest plots for comparative vaccine effect (BNT162b2 vs. ChAdOx1), meta-analysis across cohorts and databases.

## Discussion

### Key findings

Our analyses showed a substantial reduction of risk (38-66%) for thromboembolic and cardiac events in the acute phase of COVID-19 associated with vaccination. This finding was consistent across four databases and three different European countries. Risks for subacute post COVID-19 VTE and HF were also reduced, but to a lesser extent (29-46%).

The risks of post COVID-19 ATE, MP, and VACA were also reduced in vaccinated persons, but only in the acute phase.

### Results in context

The relationship between SARS-CoV-2 infection, COVID-19 vaccines, and thromboembolic and/or cardiac complications is tangled. Some large studies have found an increased risk of VTE and ATE following both ChAdOx1 and BNT162b2 vaccination (17,43). However, other studies have not identified such risk (44). It is worth noting that an elevated risk of VTE has also been reported among patients with COVID-19, and its occurrence can lead to poor prognosis and mortality (45–47). Similarly, several observational studies have found an association between COVID-19 mRNA vaccination and a short-term increased risk of myocarditis, particularly among younger male individuals (13–16,48–50). For instance, a self-controlled case series study conducted in England revealed about 30% increased risk of hospital admission due to myocarditis within 28 days following both ChAdOx1 and BNT162b2 vaccines. However, this same study also found a 9-fold higher risk for myocarditis following a positive SARS-CoV-2 test, clearly offsetting the observed post-vaccine risk.

COVID-19 vaccines have demonstrated high efficacy and effectiveness in preventing infection and reducing the severity of acute-phase infection. However, with the emergence of newer variants of the virus, such as the Omicron variant (51), and the waning protective effect of the vaccine over time (52), there is a growing interest in understanding whether the vaccine can also reduce the risk of complications after breakthrough infections. Our study demonstrates that vaccination may help mitigate the risk of thromboembolic and cardiac events associated with SARS-CoV-2, including breakthrough infections.

Recent studies suggested that COVID-19 vaccination could potentially protect against acute post COVID-19 cardiac and thromboembolic events (22–24). For example, in a large prospective cohort study, Xie et. al (2022) (22) found the risk of VTE after SARS-CoV-2 infection was largely attenuated in the fully vaccinated ambulatory patients (about 21-fold and 6-fold after non-vaccinated and breakthrough infection, respectively).

While the effect of vaccination on subacute post COVID-19 conditions has been studied, (23,24,53–59), there has been limited reporting on the condition-specific risks associated with COVID-19, even though the prognosis for different complications can vary significantly.

In line with previous studies, our findings suggest a potential benefit of vaccination in reducing the risk of post COVID-19 thromboembolic and cardiac complications. We included broader populations, estimated the risk in both acute and subacute infection phases and replicated these using four large independent observational databases. By pooling results across different settings, we provided the most up-to-date and robust evidence on this topic.

### Strengths and limitations

The study has several strengths. Our multinational study covering different healthcare systems and setting showed consistent results across all databases, which highlights the robustness and replicability of our findings. All databases had complete recordings of vaccination status (date and brand). Algorithms to identify study outcomes were used in previous published network studies, including regulatory-funded research (6,8,19,31). The staggered cohort design and methods used are yet another strength of our study, as recent research has demonstrated the validity of this design to minimise bias (35).

Our study also has limitations. First, the use of real-world data comes with inherent limitations including data quality issues and risk of confounding (60). To address these limitations, we employed state-of-the-art methods, including large-scale propensity score weighting and calibration of effect estimates using NCO (35,41). A recent study (61) has demonstrated that methodologically sound observational studies based on routinely collected data can produce similar results to clinical trials. Another limitation is potential misclassification: Asymptomatic and mild COVID-19 infections may have not been recorded and post COVID-19 outcomes of interest may be under-recorded in primary care databases (CPRD AURUM and GOLD) without hospital linkage. However, results in SIDIAP and CORIVA which include secondary care data were similar.

## Conclusions

Vaccination against SARS-CoV-2 substantially reduced the risk of acute post COVID-19 thromboembolic and cardiac complications, likely through a reduction in the severity of COVID-19 disease due to vaccine-induced immunity. These protective effects lasted for up to one year for post COVID-19 VTE, but not clearly for other complications. Further research is needed on the possible waning of these protective effects over time, and on the impact of booster vaccination to regain these potential benefits.

## Data Availability

CPRD: CPRD data were obtained under the CPRD multi-study license held by the University of Oxford after Research Data Governance (RDG) approval. Direct data sharing is not allowed.
SIDIAP: In accordance with current European and national law, the data used in this study is only available for the researchers participating in this study. Thus, we are not allowed to distribute or make publicly available the data to other parties. However, researchers from public institutions can request data from SIDIAP if they comply with certain requirements. Further information is available online (https://www.sidiap.org/index.php/menu-solicitudesen/application-proccedure) or by contacting SIDIAP.
CORIVA: CORIVA data were obtained under the approval of Research Ethics Committee of the University of Tartu and the patient level data sharing is not allowed.

## Article Information

### Corresponding author

Prof. Daniel Prieto-Alhambra

Botnar Research Centre, Windmill Road, OX3 7LD, Oxford, United Kingdom

### Author Contributions

DPA and AMJ led the conceptualisation of the study with contributions from MC and NMB. AMJ, TDS, ER, AU and NT adapted the study design with respect to the local vaccine rollouts. AD and WM mapped and curated CPRD data. MC and NMB developed code with methodological contributions advice from MTSS and CP. DPA, MC, NT, TDS, HE, XL, CR, and AMJ clinically interpreted the results. NMB, XL, AMJ and DPA wrote the first draft of the manuscript, and all authors read, revised, and approved the last version. DPA and AMJ obtained the funding for this research.

### Ethics

The study was approved by the CPRD’s Research Data Governance Process, Protocol No 21_000557 and the Clinical Research Ethics committee of Fundació Institut Universitari per a la recerca a l’Atenció Primària de Salut Jordi Gol i Gurina (IDIAPJGol) (approval number 4R22/133) and the Research Ethics Committee of the University of Tartu (approval No. 330/T-10).

### Funding/Support

The research was supported by the National Institute for Health and Care Research (NIHR) Oxford Biomedical Research Centre (BRC).

DPA is funded through a NIHR Senior Research Fellowship (Grant number SRF-2018–11-ST2-004).

Funding to perform the study in the SIDIAP database was provided by the Real World Epidemiology (RWEpi) research group at IDIAPJGol.

Costs of databases mapping to OMOP CDM were covered by the European Health Data and Evidence Network (EHDEN).

### Data Sharing Statement

<u>CPRD</u>: CPRD data were obtained under the CPRD multi-study license held by the University of Oxford after Research Data Governance (RDG) approval. Direct data sharing is not allowed.

<u>SIDIAP</u>: In accordance with current European and national law, the data used in this study is only available for the researchers participating in this study. Thus, we are not allowed to distribute or make publicly available the data to other parties. However, researchers from public institutions can request data from SIDIAP if they comply with certain requirements. Further information is available online (https://www.sidiap.org/index.php/menu-solicitudesen/application-proccedure) or by contacting SIDIAP.

<u>CORIVA:</u> CORIVA data were obtained under the approval of Research Ethics Committee of the University of Tartu and the patient level data sharing is not allowed.

## Acknowledgements

This study is based in part on data from the Clinical Practice Research Datalink obtained under license from the UK Medicines and Healthcare products Regulatory Agency. We thank the patients who provided these data, and the NHS who collected the data as part of their care and support. All interpretations, conclusions and views expressed in this publication are those of the authors alone and not necessarily those of CPRD.

We would also like to thank the healthcare professionals in the Catalan healthcare system involved in the management of COVID-19 during these challenging times, from primary care to intensive care units; the Institut de Català de la Salut and the Program d’Analítica de Dades per a la Recerca i la Innovació en Salut for providing access to the different data sources accessible through SIDIAP.

