## Supplementary material for "The role of COVID-19 vaccines in preventing post COVID-19 thromboembolic and cardiovascular complications: a multinational cohort study"

### SUPPLEMENT

The effectiveness of COVID-19 vaccines to prevent post COVID cardiac and thromboembolic complications: staggered cohort analyses of data from Spain, Estonia, and the UK.

**Figure S17: Forest plots for vaccine effectiveness (any COVID-19 vaccine) on preventing venous thromboembolism complications,** for each cohort and database (meta-analysis estimates across cohorts in the last panel).Follow-up ends at first vaccine dose after index date. Dashed line represents a level of hetereogeneity I2 > 0.4. 119

**Figure S18: Forest plots for vaccine effectiveness (any COVID-19 vaccine) on preventing venous thromboembolism complications,** for each cohort and database (meta-analysis estimates across cohorts in the last panel).Only first outcome after COVID-19 captured. Dashed line represents a level of hetereogeneity I2 > 0.4. 120

**Figure S19: Forest plots for vaccine effectiveness (ChAdOx1 vaccine) on preventing venous thromboembolism complications,** for each cohort and database (meta-analysis estimates across cohorts in the last panel). Dashed line represents a level of hetereogeneity I2 > 0.4. 121

**Figure S20: Forest plots for vaccine effectiveness (ChAdOx1 vaccine) on preventing venous thromboembolism complications,** for each cohort and database (meta-analysis estimates across cohorts in the last panel).Follow-up ends at first vaccine dose after index date. Dashed line represents a level of hetereogeneity I2 > 0.4. 122

**Figure S21: Forest plots for vaccine effectiveness (ChAdOx1 vaccine) on preventing venous thromboembolism complications,** for each cohort and database (meta-analysis estimates across cohorts in the last panel).Only first outcome after COVID-19 captured. Dashed line represents a level of hetereogeneity I2 > 0.4. 123

**Figure S22: Forest plots for vaccine effectiveness (BNT162b2 vaccine) on preventing venous thromboembolism complications,** for each cohort and database (meta-analysis estimates across cohorts in the last panel). Dashed line represents a level of hetereogeneity I2 > 0.4. 124

**Figure S23: Forest plots for vaccine effectiveness (BNT162b2 vaccine) on preventing venous thromboembolism complications,** for each cohort and database (meta-analysis estimates across cohorts in the last panel).Follow-up ends at first vaccine dose after index date. Dashed line represents a level of hetereogeneity I2 > 0.4. 125

**Figure S24: Forest plots for vaccine effectiveness (BNT162b2 vaccine) on preventing venous thromboembolism complications,** for each cohort and database (meta-analysis estimates across cohorts in the last panel).Only first outcome after COVID-19 captured. Dashed line represents a level of hetereogeneity I2 > 0.4. 126

**Figure S25: Forest plots for vaccine effectiveness (any COVID-19 vaccine) on preventing arterial thromboembolism complications,** for each cohort and database (meta-analysis estimates across cohorts in the last panel). Dashed line represents a level of hetereogeneity I2 > 0.4. 127

**Figure S26: Forest plots for vaccine effectiveness (any COVID-19 vaccine) on preventing arterial thromboembolism complications,** for each cohort and database (meta-analysis estimates across cohorts in the last panel).Follow-up ends at first vaccine dose after index date. Dashed line represents a level of hetereogeneity I2 > 0.4. 128

**Figure S27: Forest plots for vaccine effectiveness (any COVID-19 vaccine) on preventing arterial thromboembolism complications,** for each cohort and database (meta-analysis estimates across cohorts in the last panel).Only first outcome after COVID-19 captured. Dashed line represents a level of hetereogeneity I2 > 0.4. 129

**Figure S28: Forest plots for vaccine effectiveness (ChAdOx1 vaccine) on preventing arterial thromboembolism complications,** for each cohort and database (meta-analysis estimates across cohorts in the last panel). Dashed line represents a level of hetereogeneity I2 > 0.4. 130

**Figure S29: Forest plots for vaccine effectiveness (ChAdOx1 vaccine) on preventing arterial thromboembolism complications,** for each cohort and database (meta-analysis estimates across cohorts in the last panel).Follow-up ends at first vaccine dose after index date. Dashed line represents a level of hetereogeneity I2 > 0.4. 131

#### Figure S1: Details of the country-specific vaccination rollout and study-specific enrolment periods and definition of priority groups for vaccination

| | CPRD (AURUM and GOLD) | | | SPAIN (SIDIAP) | | | ESTONIA (CORIVA) | | |
|---|---|---|---|---|---|---|---|---|---|
| | **Priority group*** | **Cohort inclusion criteria** | **Enrolment windows** | **Priority group*** | **Cohort inclusion criteria** | **Enrolment windows** | **Priority group*** | **Cohort inclusion criteria** | **Enrolment windows** |
| | 1) Residents in a care home for older adults and staff working in care homes for older adults | | excluded as probably vaccinated before 04/01/2021 | 1) Residents and workers of nursing homes<br>2) First line healthcare workers<br>3) Second line healthcare workers<br>4) Severely disabled | | excluded as probably vaccinated before 20/02/2021 | Healthcare workers in hospitals, emergency rooms and family medicine centers | | excluded as probably vaccinated before 28/01/2021 |
| **Study 1** | 2) All those 80 years of age and over and frontline health and social care workers<br>3) All those 75 years of age and over | Age ≥ 75 | 04/01/2021-27/01/2021 | 5A) > 80 years | Age ≥ 80 | 20/02/2021 - 22/03/2021 | People in their 80s, then 70-80-year-olds with chronic diseases<br>All people 70+ | Age ≥ 70 | 28/01/2021 - 20/04/2021 |
| **Study 2** | 4) All those 70 years of age and over and clinically extremely vulnerable individuals (not including pregnant women and those under 16 years of age)<br>5) All those 65 years of age and over<br>6) Adults aged 16 to 65 years in an at-risk group | Age ≥ 65 and clinically extremely vulnerable, at risk people | 28/01/2021-28/02/2021 | 5 B) 70-79 years<br>5C) 66-69 years<br>6) Essential workers**<br>7) People >16 years with high risk factors*<br>8) 60-65 years | Age ≥ 60 and clinically extremely vulnerable | 24/03/2021 - 04/05/2021 | All people 65+ and people in the risk group < 65 years old | Age ≥ 65 and ≥ 18 with high risk | 21/04/2021 - 01/05/2021 |
| **Study 3** | 7) All those 60 years of age and over<br>8) All those 55 years of age and over<br>9) All those 50 years of age and over | Age ≥ 50 | 01/03/2021 – 13/04/2021 | 9) 50-59 years<br>10) 40-49 years | Age ≥ 40 | 05/05/2021 - 16/06/2021 | Starting vaccination for the age group 60+<br>Age group 50-59 starting vaccination<br>Age group 40-49 starting vaccination | Age ≥ 40 | 02/05/2021 - 12/06/2021 |
| **Study 4** | 10) Rest of the population >=18 | Age ≥ 18 | 14/04/2021 – 31/07/2021 | 11) 30-39 years<br>12) 16-29 years | Age ≥ 18 | 17/06/2021 - 01/08/2021 | Age group 30-39 starting vaccination<br>Age group 19-99 starting vaccination | Age ≥ 19 | 13/06/2021 - 01/08/2021 |

*Priority groups are country-specific and originally defined for the planning of the respective national vaccination rollout strategy by the respective governmental agency.
**Essential workers, frontline healthcare workers, social care workers could not be identified from the data. Therefore, people were included based on their age and records of conditions relevant for "high risk" and "at risk" categories for priorisation.

**Table S1: Details of the country-specific vaccination rollout Country-specific lists of conditions characterising people as “clinically extremely vulnerable/high risk” or “at risk” for severe COVID-19.**

| Categories | UK [1] | Spain [2] | Estonia [3] |
|---|---|---|---|
| | **Conditions for “high risk”/ ”clinically extremely vulnerable”** | | |
| *Solid organ transplant* | Solid organ transplant recipients who remain on long term immune suppression therapy | Solid organ transplant recipient (or in waiting list for transplant): lung, renal, pancreatic, cardiac, hepatic, or intestinal | organ transplantation (performed up to 2 years ago) |
| *Bone marrow/stem cell transplant* | People who have had bone marrow or stem cell transplants in the last 6 months, or still taking immunosuppression drugs | Hematopoietic stem cell transplant recipient in the last 2 years OR >50 years of age AND/OR <80% Karnofsky index | bone marrow transplantation (performed up to 2 years ago) |
| *Immunodeficiencies* | | Primary immunodeficiencies, excluding IgA deficiency and antibody formation defect | primary immunodeficiency |
| | | HIV infection with <200c/ml in the last 6 months | |
| *Immunosuppressive therapy* | People on immunosuppression therapies sufficient to significantly increase risk of infection | | people who have received immunosuppressive therapy in the last 5 years with hematological, rheumatological, gastroenterological, neurological diseases |
| *Blood cancer* | People with cancers of the blood or bone marrow such as leukaemia, lymphoma, or myeloma at any stage of treatment | Blood cancer in the last 5 years OR uncontrolled cancer AND/OR ECOG 3-4 AND/OR severe neutropenia (<500 neutrophiles/mm3) regardless of years since diagnosis | malignant tumors of lymphoid and hematopoietic tissue (diagnosed up to 5 years ago) |
| *Other active cancer* | Cancer undergoing active chemo/ radiotherapy or radical radiotherapy for lung cancer. | Solid organ cancer in treatment with chemotherapy or metastatic, OR receiving radiotherapy for tumours in the thorax with risks of pneumonitis (oesophageal tumour, radiotherapy on lung metastases, etc.) | other tumors (diagnosed up to 1 year ago) |
| *Neurological/muscular disease* | | | demyelinating diseases of the central nervous system |
| | | | amyotrophic lateral sclerosis; a stroke within the last year and residual symptoms of a stroke; |
| | | | dementia; Parkinson's disease |
| *Down Syndrome* | | Individuals with Down syndrome aged >40 years | |
| *Kidney failure + Dialysis* | | Haemodialysis or peritoneal dialysis | Kidney failure |
| *Severe respiratory disease* | Cystic Fibrosis, idiopathic pulmonary fibrosis, etc. | Cystic fibrosis –*added on 22 June 2021, so can be omitted the purpose of the study* | cystic fibrosis |
| | People with severe respiratory conditions including severe asthma and severe COPD | | |
| *Rare diseases* | People with rare diseases and inborn errors of metabolism that significantly increase the risk of infections (such as SCID, homozygous sickle cell) | | |

| | |
|---|---|
| *Pregnancy + Heart disease* | People who are pregnant with significant congenital heart disease |

| Categories | Conditions for "at risk"/ "underlying conditions" | | |
|---|---|---|---|
| *Severe Chronic Kidney disease* | unresolved CKD stage 3-5 diagnosis | | |
| *Severe Chronic Liver disease* | chronic liver disease diagnosis | | |
| *Immunosuppression* | immunosuppressive drug treatment within the 12 month period leading up to and including the reporting period end date. | | |
| | expiring immunosuppression diagnosis within the 12 months leading up to and including the reporting period end date which is not resolved | | |
| | unresolved 'persisting' immunosuppression diagnosis | | |
| *Diabetes* | unresolved diabetes diagnosis. | | diabetes |
| *Chronic and severe respiratory disease* | chronic respiratory disease diagnosis | | chronic bronchitis, emphysema, COPD; severe asthma (oral corticosteroid therapy in the last 5 years, biological therapy), bronchiectasis disease |
| | Asthma treatment within the 12 month period leading up to the reporting period end date OR Emergency hospital admission due to asthma. | | |
| *Obesity* | explicit high BMI/ BMI >=40 | | obesity (BMI ≥40) |
| *Neurological disease* | chronic neurological disease diagnosis | | |
| *Severe cardiovascular disease* | chronic heart disease diagnosis | | cardiological diseases |
| *cerebrovascular disease* | TIA diagnosis. | | |
| *Pregnancy* | pregnant in the last 9 months | | |
| *Other* | 'requires influenza virus vaccination' | | |
| | learning disability diagnosis | | |

Sources for overview table:

[1] NHS Digital. COVID-19 – high risk shielded patient list identification methodology- Rule logic. Update from 8 December 2021 — General Practice Specification and Extraction Service (GPSES), NHS Digital: "Business Rules for Patient-level Data Extracts 2019/20 COVID19 At-risk patients", Version: 1.1, Version Date: 17/03/2020

[2] https://beteve.cat/societat/vacunacio-grups-risc-malalties-greus-simultaniament-franja-70-79-anys/

[3] https://vaktsineeri.ee/wp-content/uploads/2021/06/covid-19_vaktsineerimise_plaan_19.01_0.pdf

**Figure S2: Study Design using UK vaccination rollout (CPRD AURUM and GOLD) as illustrative example.**

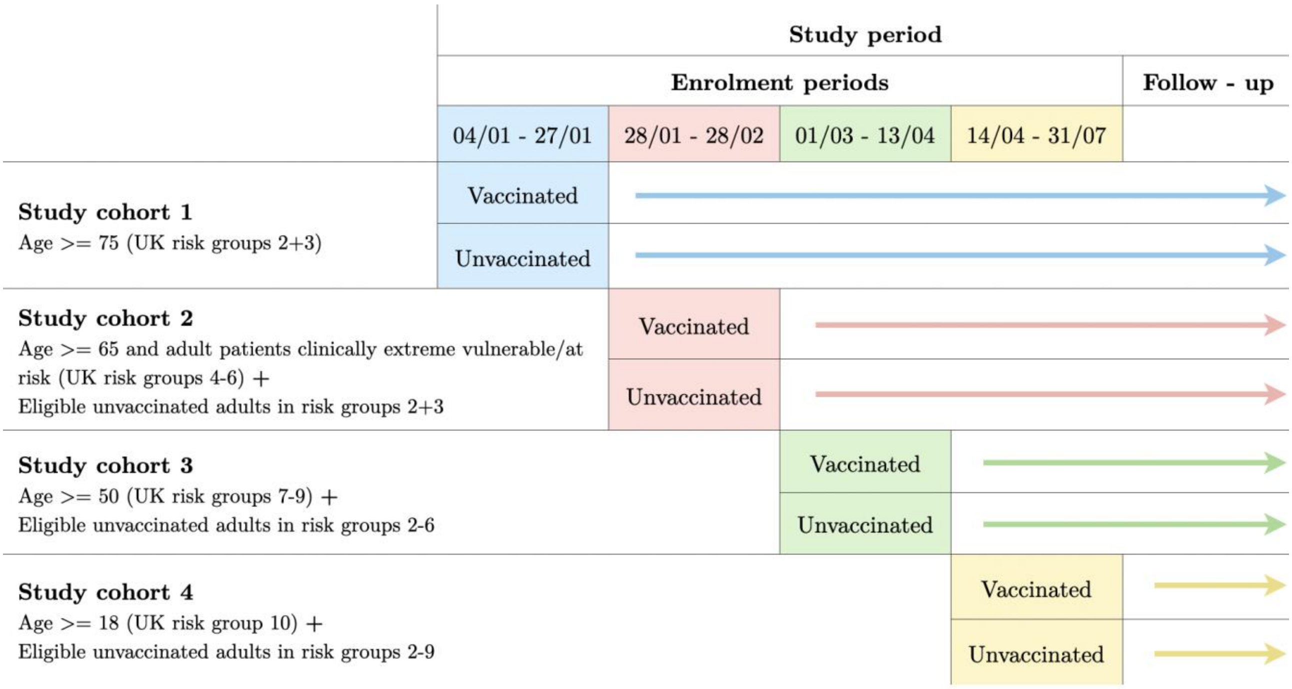

**Figure S3: Follow-up in vaccinated and unvaccinated cohorts.**

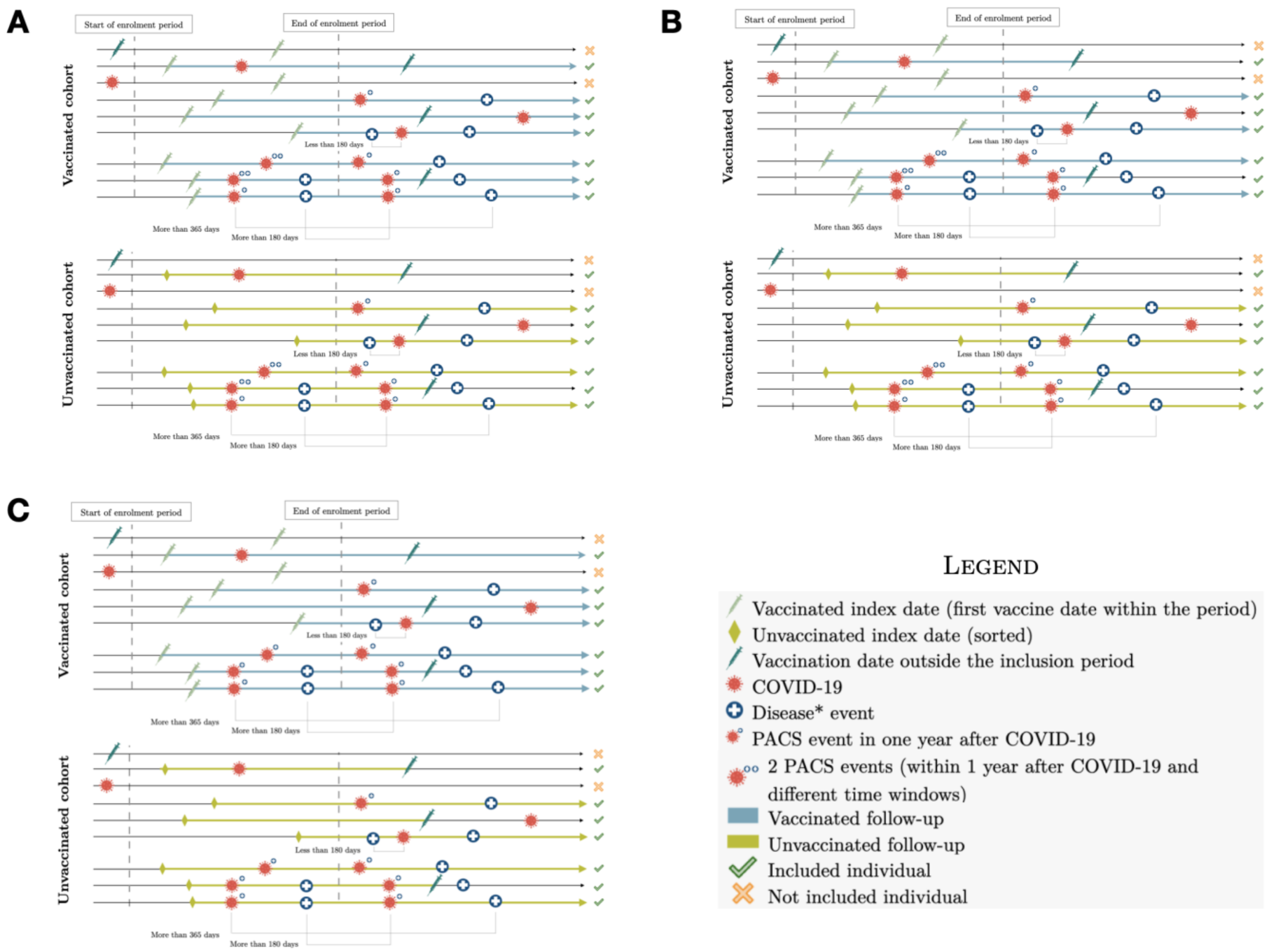

(A) Main analysis. (B) Follow-up ends at first vaccine dose after index date. (C) Post COVID outcomes refer to the first event within a year after COVID-19 infection.

#### Figure S4: Study Inclusion Flowchart CPRD AURUM

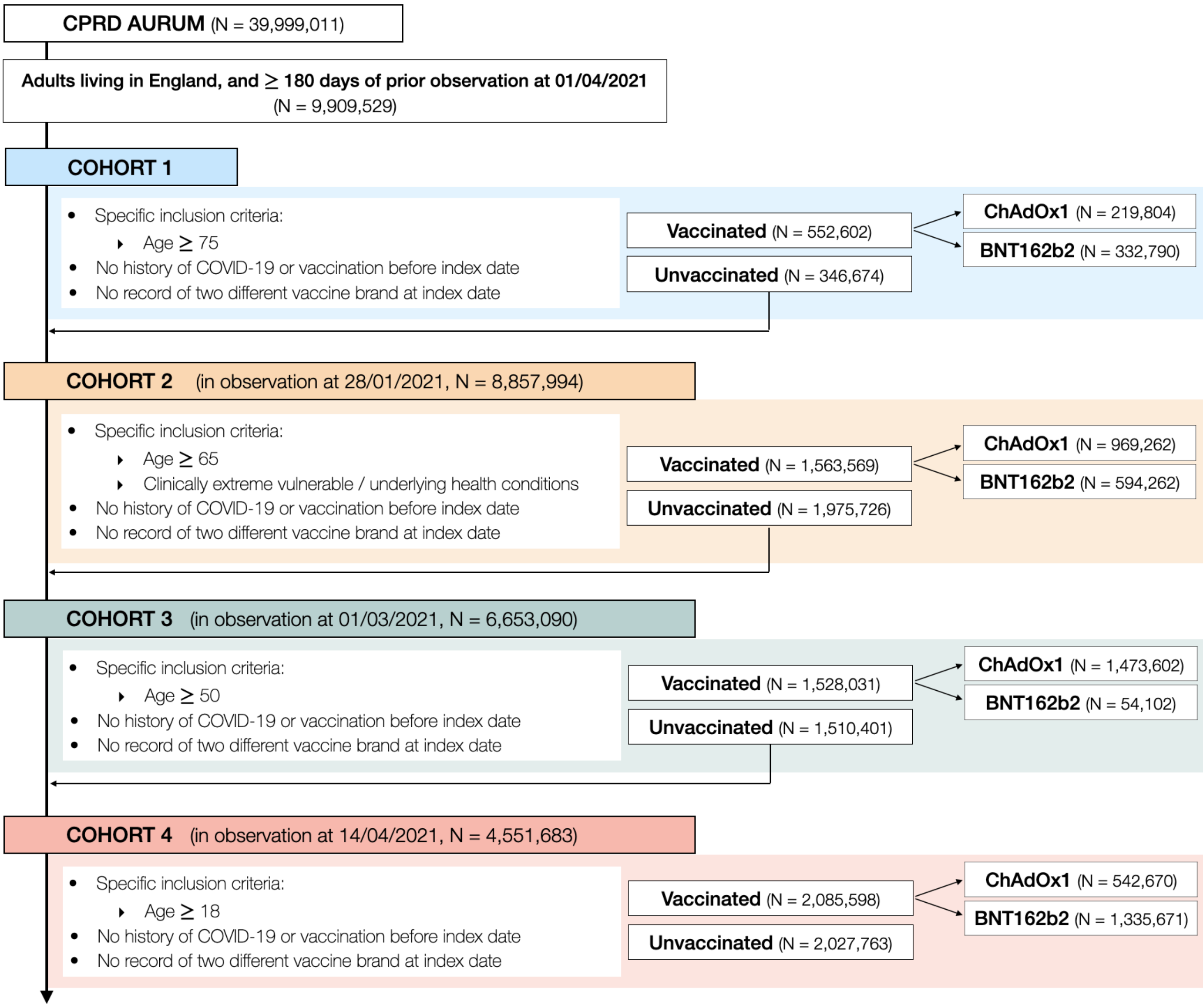

**Figure S5: Study Inclusion Flowchart CPRD GOLD.**

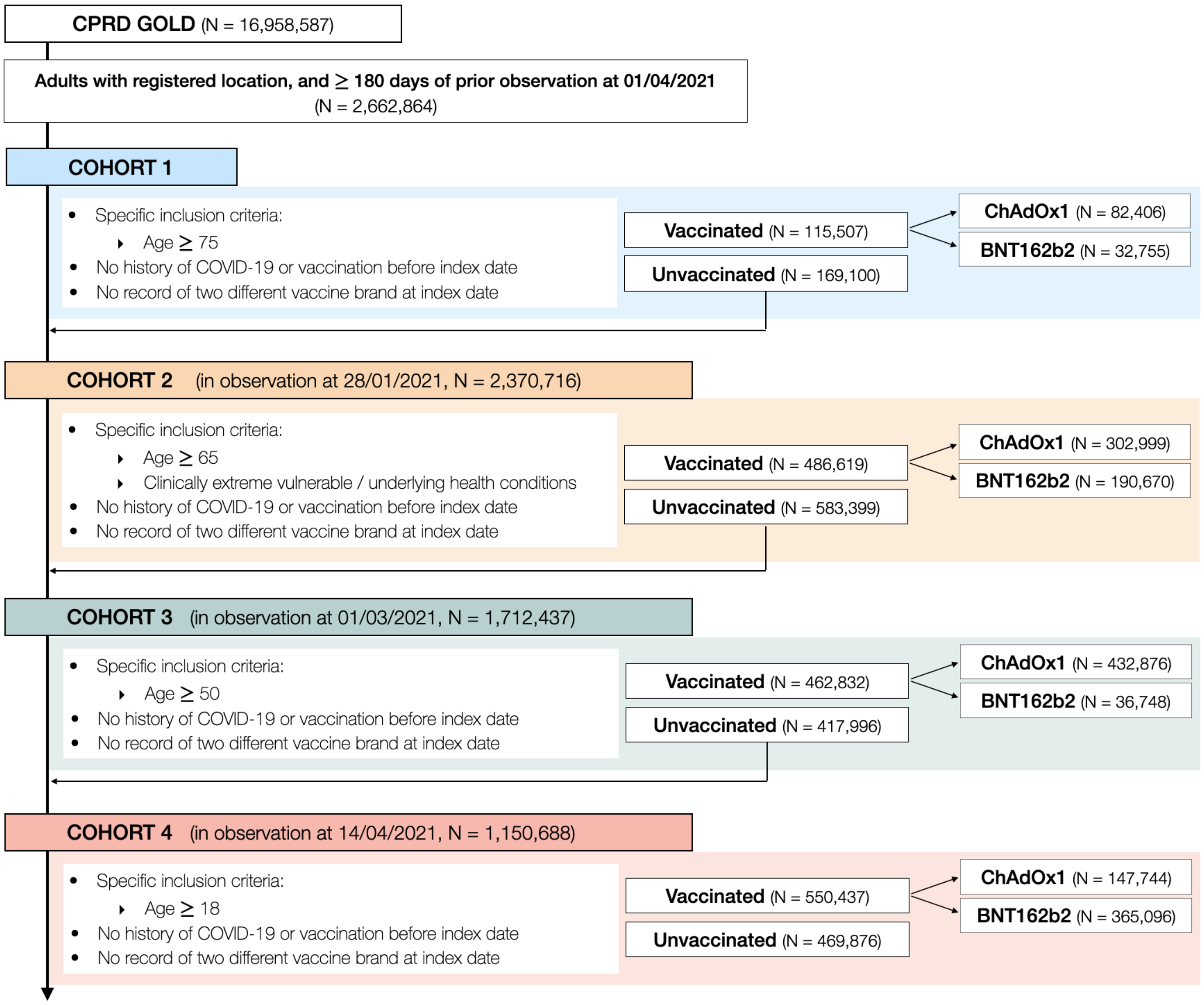

**Figure S6: Study Inclusion Flowchart SIDIAP.**

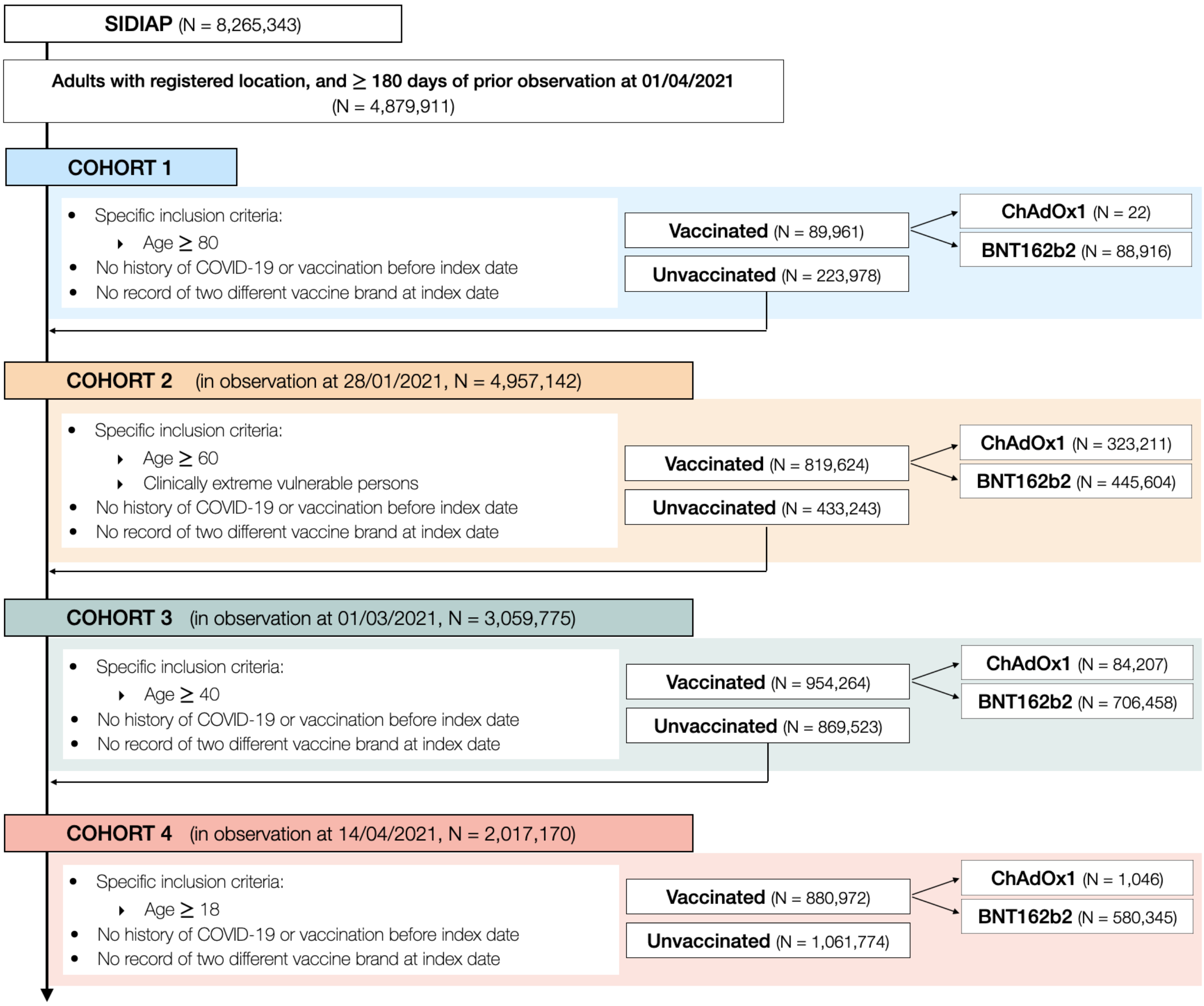

**Figure S7: Study Inclusion Flowchart CORIVA.**

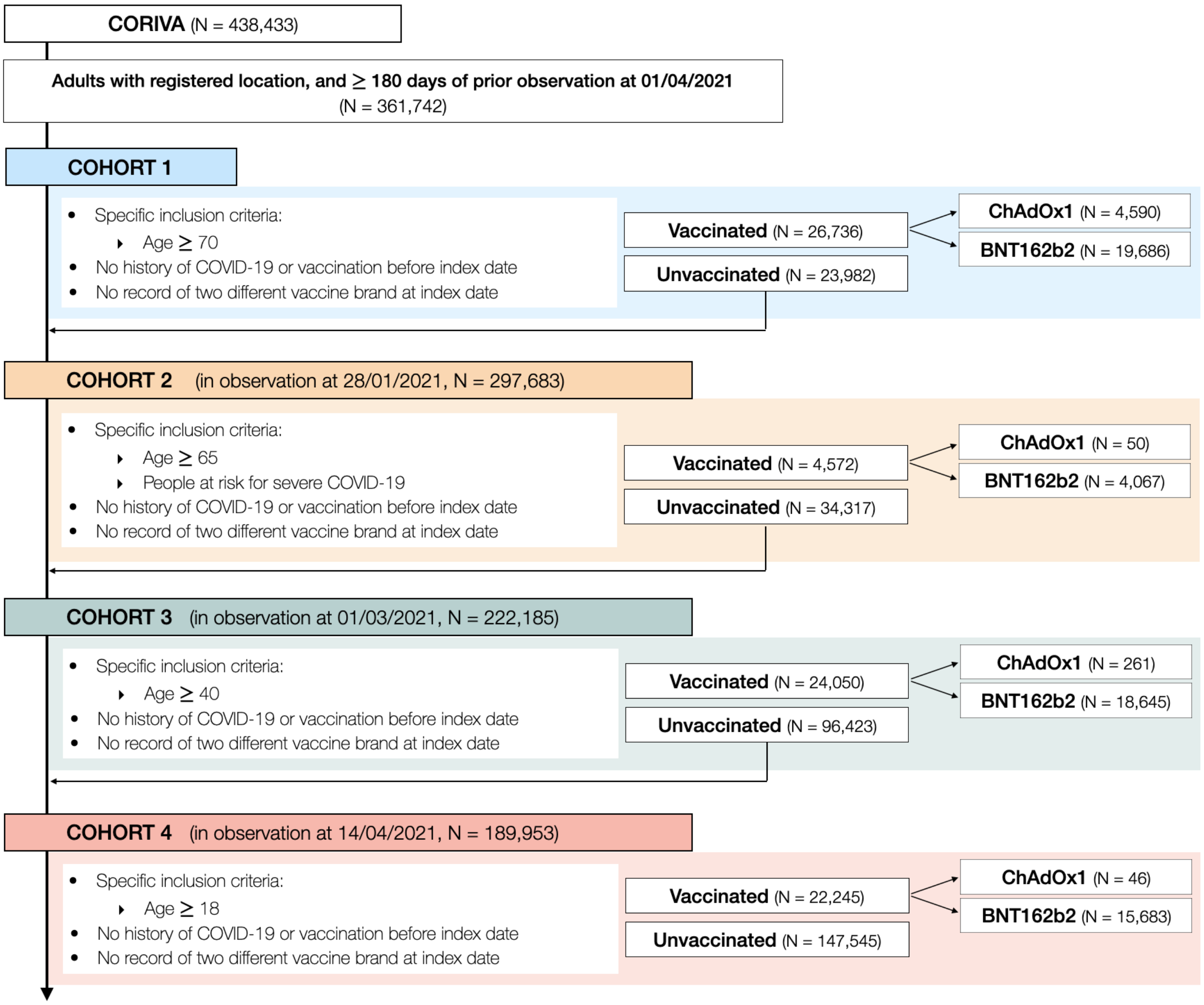

**Table S2: Characteristics of unweighted populations in CPRD AURUM,** database, stratified by staggered cohort and exposure status. Exposure is any COVID-19 vaccine.

| | Cohort 1 | | | Cohort 2 | | | Cohort 3 | | | Cohort 4 | | |
|---|---|---|---|---|---|---|---|---|---|---|---|---|
| | **Unvaccinated** | **Vaccinated** | **ASMD** | **Unvaccinated** | **Vaccinated** | **ASMD** | **Unvaccinated** | **Vaccinated** | **ASMD** | **Unvaccinated** | **Vaccinated** | **ASMD** |
| **N (individuals)** | 346,674 | 552,602 | | 1,975,726 | 1,563,569 | | 1,510,401 | 1,528,031 | | 2,027,763 | 2,085,598 | |
| **Age, median [Q25-Q75]** | 78 [76-82] | 81 [78-86] | 0.319 | 42 [32-55] | 68 [60-73] | 0.995 | 41 [31-53] | 55 [51-60] | 0.610 | 35 [27-45] | 35 [27-43] | 0.134 |
| **Sex: Female, N(%)** | 192,771 (56%) | 314,141 (57%) | 0.025 | 1,244,636 (63%) | 852,976 (55%) | 0.172 | 880,404 (58%) | 771,929 (51%) | 0.157 | 862,676 (43%) | 958,124 (46%) | 0.068 |
| **Years of prior history*, median [Q25-Q75]** | 24 [10-35] | 25 [11-37] | 0.056 | 15 [7-25] | 21 [9-32] | 0.278 | 13 [6-22] | 17 [8-27] | 0.207 | 8 [4-16] | 8 [3-18] | 0.032 |
| **Number visits, median [Q25-Q75]** | 10 [5-17] | 11 [6-19] | | 6 [2-11] | 10 [6-17] | | 4 [1-10] | 7 [3-12] | | 2 [0-5] | 3 [1-7] | |
| **Number pcrs, median [Q25-Q75]** | 0[0-0] | 0[0-0] | | 0[0-0] | 0[0-0] | | 0[0-0] | 0[0-0] | | 0[0-0] | 0[0-0] | |
| **Comorbidities**, N(%)** | | | | | | | | | | | | |
| Anxiety | 51,067 (15%) | 82,765 (15%) | 0.007 | 435,770 (22%) | 309,567 (20%) | 0.056 | 289,563 (19%) | 292,336 (19%) | 0.001 | 265,627 (13%) | 338,058 (16%) | 0.088 |
| Asthma | 36,474 (11%) | 61,899 (11%) | 0.022 | 501,209 (25%) | 275,768 (18%) | 0.189 | 294,123 (19%) | 209,708 (14%) | 0.155 | 138,317 (7%) | 173,056 (8%) | 0.056 |
| Chronic kidney disease | 71,083 (21%) | 137,953 (25%) | 0.107 | 55,104 (3%) | 154,263 (10%) | 0.294 | 24,340 (2%) | 30,896 (2%) | 0.031 | 18,625 (1%) | 5,796 (0%) | 0.083 |
| COPD | 28,665 (8%) | 47,422 (9%) | 0.011 | 29,729 (2%) | 107,297 (7%) | 0.270 | 13,433 (1%) | 15,685 (1%) | 0.014 | 10,543 (1%) | 2,535 (0%) | 0.071 |
| Dementia | 16,610 (5%) | 32,934 (6%) | 0.052 | 6,692 (0%) | 15,828 (1%) | 0.082 | 3,425 (0%) | 3,219 (0%) | 0.003 | 2,691 (0%) | 575 (0%) | 0.037 |
| Depressive disorder | 42,145 (12%) | 64,840 (12%) | 0.013 | 367,463 (19%) | 283,390 (18%) | 0.012 | 236,584 (16%) | 265,876 (17%) | 0.047 | 210,644 (10%) | 253,317 (12%) | 0.056 |
| Diabetes | 63,154 (18%) | 101,682 (18%) | 0.005 | 107,425 (5%) | 253,026 (16%) | 0.351 | 60,932 (4%) | 84,224 (6%) | 0.069 | 49,154 (2%) | 22,968 (1%) | 0.101 |
| GERD | 18,336 (5%) | 32,360 (6%) | 0.025 | 68,070 (3%) | 84,985 (5%) | 0.097 | 39,594 (3%) | 58,961 (4%) | 0.070 | 31,119 (2%) | 38,612 (2%) | 0.025 |
| Heart failure | 17,882 (5%) | 33,578 (6%) | 0.040 | 12,275 (1%) | 37,396 (2%) | 0.146 | 5,641 (0%) | 7,204 (0%) | 0.015 | 4,611 (0%) | 1,163 (0%) | 0.046 |
| Hypertension | 172,799 (50%) | 301,294 (55%) | 0.094 | 206,105 (10%) | 545,782 (35%) | 0.611 | 110,912 (7%) | 240,120 (16%) | 0.264 | 86,435 (4%) | 54,357 (3%) | 0.091 |
| Hypothyroidism | 31,444 (9%) | 56,367 (10%) | 0.038 | 78,934 (4%) | 118,453 (8%) | 0.154 | 43,892 (3%) | 67,686 (4%) | 0.081 | 30,993 (2%) | 35,364 (2%) | 0.013 |

| | Cohort 1 | | | Cohort 2 | | | Cohort 3 | | | Cohort 4 | | |
|---|---|---|---|---|---|---|---|---|---|---|---|---|
| | **Unvaccinated** | **Vaccinated** | **ASMD** | **Unvaccinated** | **Vaccinated** | **ASMD** | **Unvaccinated** | **Vaccinated** | **ASMD** | **Unvaccinated** | **Vaccinated** | **ASMD** |
| Malignant neoplastic disease | 68,984 (20%) | 131,635 (24%) | 0.095 | 60,318 (3%) | 207,240 (13%) | 0.379 | 29,689 (2%) | 61,923 (4%) | 0.122 | 22,701 (1%) | 13,590 (1%) | 0.050 |
| Myocardial infarction | 16,027 (5%) | 29,525 (5%) | 0.033 | 20,259 (1%) | 56,161 (4%) | 0.172 | 7,396 (0%) | 12,703 (1%) | 0.042 | 5,764 (0%) | 1,621 (0%) | 0.049 |
| Osteoporosis | 29,909 (9%) | 59,521 (11%) | 0.072 | 18,263 (1%) | 64,108 (4%) | 0.204 | 8,846 (1%) | 15,089 (1%) | 0.045 | 6,922 (0%) | 2,535 (0%) | 0.046 |
| Pneumonia | 16,992 (5%) | 28,398 (5%) | 0.011 | 31,746 (2%) | 50,739 (3%) | 0.107 | 17,785 (1%) | 22,064 (1%) | 0.023 | 13,305 (1%) | 13,282 (1%) | 0.002 |
| Rheumatoid arthritis | 6,596 (2%) | 11,237 (2%) | 0.009 | 14,811 (1%) | 33,173 (2%) | 0.116 | 5,548 (0%) | 9,130 (1%) | 0.033 | 3,710 (0%) | 1,816 (0%) | 0.026 |
| Stroke | 15,249 (4%) | 26,478 (5%) | 0.019 | 16,942 (1%) | 39,907 (3%) | 0.131 | 7,422 (0%) | 10,828 (1%) | 0.028 | 6,030 (0%) | 2,291 (0%) | 0.042 |
| Venous thromboembolism | 19,607 (6%) | 35,332 (6%) | 0.031 | 28,876 (1%) | 63,048 (4%) | 0.158 | 13,608 (1%) | 26,299 (2%) | 0.072 | 12,945 (1%) | 4,787 (0%) | 0.062 |

The 4 cohorts represent vaccine rollout periods. *Calculated as the days of previous observation in the database before index date. **Assessed anytime before index date. ASMD = Absolute standardized mean difference, COPD = Chronic obstructive pulmonary disease, GERD = Gastro-Esophageal reflux disease

**Table S3: Characteristics of weighted populations in CPRD AURUM,** database, stratified by staggered cohort and exposure status. Exposure is ChAdOx1 vaccine.

| | Cohort 1 | | | Cohort 2 | | | Cohort 3 | | | Cohort 4 | | |
|---|---|---|---|---|---|---|---|---|---|---|---|---|
| | **Unvaccinated** | **Vaccinated** | **ASMD** | **Unvaccinated** | **Vaccinated** | **ASMD** | **Unvaccinated** | **Vaccinated** | **ASMD** | **Unvaccinated** | **Vaccinated** | **ASMD** |
| **N (individuals)** | 100,517 | 99,985 | | 308,608 | 304,995 | | 448,766 | 445,397 | | 242,955 | 243,847 | |
| **Age, median [Q25-Q75]** | 78 [76-84] | 78 [76-84] | 0.001 | 62 [47-69] | 62 [47-69] | 0.007 | 50 [41-57] | 52 [41-57] | 0.001 | 44 [41-48] | 44 [41-48] | 0.003 |
| **Sex: Female, N(%)** | 57,756 (57%) | 57,174 (57%) | 0.006 | 179,574 (58%) | 176,871 (58%) | 0.004 | 236,260 (53%) | 234,320 (53%) | 0.001 | 101,805 (42%) | 100,846 (41%) | 0.011 |
| **Years of prior history*, median [Q25-Q75]** | 24 [10-36] | 24 [10-36] | 0.004 | 18 [8-29] | 18 [8-29] | 0.002 | 14 [6-24] | 14 [6-24] | 0.002 | 10 [5-18] | 9 [5-18] | 0.004 |
| **Number visits, median [Q25-Q75]** | 10 [5-18] | 10 [6-18] | | 8 [3-15] | 8 [5-15] | | 4 [1-11] | 6 [2-10] | | 2 [0-7] | 2 [1-7] | |
| **Number pcrs, median [Q25-Q75]** | 0[0-0] | 0[0-0] | | 0[0-0] | 0[0-0] | | 0[0-0] | 0[0-0] | | 0[0-0] | 0[0-0] | |
| **Comorbidities**, N(%)** | | | | | | | | | | | | |
| Anxiety | 15,457 (15%) | 15,069 (15%) | 0.009 | 67,125 (22%) | 64,408 (21%) | 0.015 | 90,806 (20%) | 87,243 (20%) | 0.016 | 37,475 (15%) | 38,040 (16%) | 0.005 |
| Asthma | 10,993 (11%) | 10,863 (11%) | 0.002 | 65,274 (21%) | 62,424 (20%) | 0.017 | 76,417 (17%) | 75,841 (17%) | 0.000 | 17,944 (7%) | 17,468 (7%) | 0.009 |
| Chronic kidney disease | 23,223 (23%) | 22,872 (23%) | 0.005 | 23,022 (7%) | 24,288 (8%) | 0.019 | 8,460 (2%) | 8,356 (2%) | 0.001 | 2,419 (1%) | 2,043 (1%) | 0.017 |
| COPD | 8,669 (9%) | 8,770 (9%) | 0.005 | 14,411 (5%) | 15,226 (5%) | 0.015 | 5,747 (1%) | 4,794 (1%) | 0.019 | 1,263 (1%) | 1,250 (1%) | 0.001 |
| Dementia | 6,529 (6%) | 6,431 (6%) | 0.003 | 3,880 (1%) | 3,541 (1%) | 0.009 | 898 (0%) | 1,004 (0%) | 0.006 | 203 (0%) | 399 (0%) | 0.023 |
| Depressive disorder | 12,559 (12%) | 12,267 (12%) | 0.007 | 60,745 (20%) | 57,903 (19%) | 0.018 | 80,264 (18%) | 77,017 (17%) | 0.016 | 32,304 (13%) | 33,302 (14%) | 0.011 |
| Diabetes | 18,750 (19%) | 18,602 (19%) | 0.001 | 39,262 (13%) | 38,979 (13%) | 0.002 | 25,334 (6%) | 26,121 (6%) | 0.009 | 6,679 (3%) | 6,315 (3%) | 0.010 |
| GERD | 5,590 (6%) | 5,471 (5%) | 0.004 | 14,645 (5%) | 14,339 (5%) | 0.002 | 14,758 (3%) | 14,823 (3%) | 0.002 | 5,727 (2%) | 6,021 (2%) | 0.007 |
| Heart failure | 6,181 (6%) | 5,763 (6%) | 0.016 | 6,098 (2%) | 5,685 (2%) | 0.008 | 2,241 (0%) | 1,983 (0%) | 0.008 | 610 (0%) | 545 (0%) | 0.006 |
| Hypertension | 52,737 (52%) | 51,796 (52%) | 0.013 | 78,623 (25%) | 79,827 (26%) | 0.016 | 51,054 (11%) | 51,059 (11%) | 0.003 | 15,350 (6%) | 15,670 (6%) | 0.004 |
| Hypothyroidism | 9,956 (10%) | 9,690 (10%) | 0.007 | 19,720 (6%) | 19,562 (6%) | 0.001 | 16,480 (4%) | 16,361 (4%) | 0.000 | 5,735 (2%) | 5,984 (2%) | 0.006 |

| | Cohort 1 | | | Cohort 2 | | | Cohort 3 | | | Cohort 4 | | |
|---|---|---|---|---|---|---|---|---|---|---|---|---|
| | **Unvaccinated** | **Vaccinated** | **ASMD** | **Unvaccinated** | **Vaccinated** | **ASMD** | **Unvaccinated** | **Vaccinated** | **ASMD** | **Unvaccinated** | **Vaccinated** | **ASMD** |
| Malignant neoplastic disease | 21,691 (22%) | 21,468 (21%) | 0.003 | 25,191 (8%) | 27,892 (9%) | 0.035 | 13,140 (3%) | 12,870 (3%) | 0.002 | 4,003 (2%) | 3,621 (1%) | 0.013 |
| Myocardial infarction | 5,182 (5%) | 4,988 (5%) | 0.008 | 7,777 (3%) | 8,594 (3%) | 0.018 | 3,415 (1%) | 3,228 (1%) | 0.004 | 933 (0%) | 636 (0%) | 0.022 |
| Osteoporosis | 9,975 (10%) | 10,018 (10%) | 0.003 | 9,052 (3%) | 9,423 (3%) | 0.009 | 3,285 (1%) | 3,499 (1%) | 0.006 | 840 (0%) | 934 (0%) | 0.006 |
| Pneumonia | 5,770 (6%) | 5,268 (5%) | 0.021 | 9,096 (3%) | 8,541 (3%) | 0.009 | 6,196 (1%) | 6,077 (1%) | 0.001 | 1,923 (1%) | 2,040 (1%) | 0.005 |
| Rheumatoid arthritis | 2,002 (2%) | 2,063 (2%) | 0.005 | 4,856 (2%) | 4,971 (2%) | 0.004 | 2,266 (1%) | 2,795 (1%) | 0.016 | 665 (0%) | 478 (0%) | 0.016 |
| Stroke | 5,085 (5%) | 4,791 (5%) | 0.012 | 6,650 (2%) | 6,616 (2%) | 0.001 | 3,150 (1%) | 2,942 (1%) | 0.005 | 908 (0%) | 773 (0%) | 0.010 |
| Venous thromboembolism | 6,342 (6%) | 6,069 (6%) | 0.010 | 9,208 (3%) | 9,850 (3%) | 0.014 | 6,193 (1%) | 7,393 (2%) | 0.023 | 2,532 (1%) | 1,239 (1%) | 0.061 |

The 4 cohorts represent vaccine rollout periods. *Calculated as the days of previous observation in the database before index date. **Assessed anytime before index date. ASMD = Absolute standardized mean difference, COPD = Chronic obstructive pulmonary disease, GERD = Gastro-Esophageal reflux disease

**Table S4: Characteristics of unweighted populations in CPRD AURUM,** database, stratified by staggered cohort and exposure status. Exposure is ChAdOx1 vaccine.

| | Cohort 1 | | | Cohort 2 | | | Cohort 3 | | | Cohort 4 | | |
|---|---|---|---|---|---|---|---|---|---|---|---|---|
| | **Unvaccinated** | **Vaccinated** | **ASMD** | **Unvaccinated** | **Vaccinated** | **ASMD** | **Unvaccinated** | **Vaccinated** | **ASMD** | **Unvaccinated** | **Vaccinated** | **ASMD** |
| **N (individuals)** | 344,687 | 219,804 | | 1,975,770 | 969,262 | | 1,510,493 | 1,473,602 | | 2,066,318 | 542,670 | |
| **Age, median [Q25-Q75]** | 78 [76-82] | 80 [77-85] | 0.240 | 42 [32-55] | 69 [63-73] | 1.059 | 41 [31-53] | 55 [51-60] | 0.605 | 34 [27-45] | 45 [42-48] | 0.507 |
| **Sex: Female, N(%)** | 191,649 (56%) | 127,656 (58%) | 0.050 | 1,244,532 (63%) | 528,692 (55%) | 0.172 | 880,468 (58%) | 739,444 (50%) | 0.163 | 880,225 (43%) | 242,758 (45%) | 0.043 |
| **Years of prior history*, median [Q25-Q75]** | 24 [10-35] | 24 [9-36] | 0.018 | 15 [7-25] | 21 [9-33] | 0.288 | 13 [6-22] | 17 [8-27] | 0.203 | 7 [4-16] | 10 [5-18] | 0.143 |
| **Number visits, median [Q25-Q75]** | 10 [5-17] | 12 [6-19] | | 6 [2-11] | 10 [6-17] | | 4 [1-10] | 7 [3-12] | | 2 [0-5] | 3 [1-8] | |
| **Number pcrs, median [Q25-Q75]** | 0[0-0] | 0[0-0] | | 0[0-0] | 0[0-0] | | 0[0-0] | 0[0-0] | | 0[0-0] | 0[0-0] | |
| **Comorbidities**, N(%)** | | | | | | | | | | | | |
| Anxiety | 50,786 (15%) | 33,355 (15%) | 0.012 | 435,737 (22%) | 187,469 (19%) | 0.067 | 289,618 (19%) | 281,846 (19%) | 0.001 | 268,451 (13%) | 86,805 (16%) | 0.085 |
| Asthma | 36,259 (11%) | 24,009 (11%) | 0.013 | 501,234 (25%) | 159,162 (16%) | 0.221 | 294,156 (19%) | 200,995 (14%) | 0.157 | 140,178 (7%) | 41,393 (8%) | 0.033 |
| Chronic kidney disease | 70,604 (20%) | 51,053 (23%) | 0.066 | 55,103 (3%) | 97,339 (10%) | 0.299 | 24,361 (2%) | 24,198 (2%) | 0.002 | 18,798 (1%) | 2,879 (1%) | 0.045 |
| COPD | 28,448 (8%) | 19,105 (9%) | 0.016 | 29,730 (2%) | 67,700 (7%) | 0.274 | 13,436 (1%) | 13,621 (1%) | 0.004 | 10,647 (1%) | 1,510 (0%) | 0.038 |
| Dementia | 16,327 (5%) | 16,795 (8%) | 0.121 | 6,689 (0%) | 12,106 (1%) | 0.103 | 3,424 (0%) | 1,896 (0%) | 0.023 | 2,729 (0%) | 447 (0%) | 0.015 |
| Depressive disorder | 41,905 (12%) | 27,009 (12%) | 0.004 | 367,500 (19%) | 170,626 (18%) | 0.026 | 236,628 (16%) | 256,789 (17%) | 0.047 | 212,626 (10%) | 74,210 (14%) | 0.104 |
| Diabetes | 62,733 (18%) | 39,751 (18%) | 0.003 | 107,449 (5%) | 154,348 (16%) | 0.344 | 60,968 (4%) | 78,394 (5%) | 0.061 | 49,223 (2%) | 9,729 (2%) | 0.041 |
| GERD | 18,253 (5%) | 12,156 (6%) | 0.010 | 68,089 (3%) | 52,694 (5%) | 0.097 | 39,619 (3%) | 56,444 (4%) | 0.068 | 31,367 (2%) | 13,807 (3%) | 0.073 |
| Heart failure | 17,716 (5%) | 13,147 (6%) | 0.037 | 12,277 (1%) | 24,276 (3%) | 0.152 | 5,638 (0%) | 5,673 (0%) | 0.002 | 4,625 (0%) | 681 (0%) | 0.024 |
| Hypertension | 171,846 (50%) | 114,922 (52%) | 0.049 | 206,105 (10%) | 345,014 (36%) | 0.626 | 110,975 (7%) | 224,543 (15%) | 0.251 | 87,010 (4%) | 31,330 (6%) | 0.072 |
| Hypothyroidism | 31,256 (9%) | 21,855 (10%) | 0.030 | 78,921 (4%) | 73,992 (8%) | 0.156 | 43,887 (3%) | 64,026 (4%) | 0.077 | 31,315 (2%) | 13,920 (3%) | 0.074 |
| Malignant neoplastic disease | 68,598 (20%) | 49,023 (22%) | 0.059 | 60,355 (3%) | 131,135 (14%) | 0.387 | 29,693 (2%) | 55,601 (4%) | 0.108 | 22,873 (1%) | 7,505 (1%) | 0.025 |

| | Cohort 1 | | | Cohort 2 | | | Cohort 3 | | | Cohort 4 | | |
|---|---|---|---|---|---|---|---|---|---|---|---|---|
| | **Unvaccinated** | **Vaccinated** | **ASMD** | **Unvaccinated** | **Vaccinated** | **ASMD** | **Unvaccinated** | **Vaccinated** | **ASMD** | **Unvaccinated** | **Vaccinated** | **ASMD** |
| Myocardial infarction | 15,938 (5%) | 11,236 (5%) | 0.023 | 20,262 (1%) | 33,142 (3%) | 0.163 | 7,401 (0%) | 11,237 (1%) | 0.035 | 5,781 (0%) | 861 (0%) | 0.026 |
| Osteoporosis | 29,704 (9%) | 22,986 (10%) | 0.063 | 18,255 (1%) | 42,319 (4%) | 0.216 | 8,856 (1%) | 12,455 (1%) | 0.031 | 6,980 (0%) | 1,380 (0%) | 0.015 |
| Pneumonia | 16,837 (5%) | 12,101 (6%) | 0.028 | 31,706 (2%) | 33,170 (3%) | 0.116 | 17,787 (1%) | 20,399 (1%) | 0.018 | 13,425 (1%) | 4,297 (1%) | 0.017 |
| Rheumatoid arthritis | 6,549 (2%) | 4,544 (2%) | 0.012 | 14,813 (1%) | 20,294 (2%) | 0.114 | 5,536 (0%) | 8,463 (1%) | 0.030 | 3,723 (0%) | 821 (0%) | 0.007 |
| Stroke | 15,118 (4%) | 10,876 (5%) | 0.027 | 16,944 (1%) | 24,851 (3%) | 0.132 | 7,424 (0%) | 9,547 (1%) | 0.021 | 6,059 (0%) | 1,153 (0%) | 0.016 |
| Venous thromboembolism | 19,455 (6%) | 13,782 (6%) | 0.026 | 28,887 (1%) | 39,415 (4%) | 0.159 | 13,609 (1%) | 24,312 (2%) | 0.067 | 13,021 (1%) | 1,936 (0%) | 0.039 |

The 4 cohorts represent vaccine rollout periods. *Calculated as the days of previous observation in the database before index date. **Assessed anytime before index date. ASMD = Absolute standardized mean difference, COPD = Chronic obstructive pulmonary disease, GERD = Gastro-Esophageal reflux disease

**Table S5: Characteristics of weighted populations in CPRD AURUM,** database, stratified by staggered cohort and exposure status. Exposure is BNT162b2 vaccine.

| | Cohort 1 | | | Cohort 2 | | | Cohort 3 | | | Cohort 4 | | |
|---|---|---|---|---|---|---|---|---|---|---|---|---|
| | **Unvaccinated** | **Vaccinated** | **ASMD** | **Unvaccinated** | **Vaccinated** | **ASMD** | **Unvaccinated** | **Vaccinated** | **ASMD** | **Unvaccinated** | **Vaccinated** | **ASMD** |
| **N (individuals)** | 105,853 | 106,684 | | 255,400 | 253,164 | | 37,236 | 37,227 | | 630,449 | 633,628 | |
| **Age, median [Q25-Q75]** | 80 [77-85] | 80 [77-85] | 0.003 | 58 [46-67] | 58 [46-68] | 0.006 | 54 [45-64] | 54 [45-64] | 0.016 | 32 [25-37] | 32 [25-37] | 0.001 |
| **Sex: Female, N(%)** | 60,612 (57%) | 60,891 (57%) | 0.004 | 147,020 (58%) | 146,572 (58%) | 0.007 | 22,049 (59%) | 22,144 (59%) | 0.005 | 278,649 (44%) | 282,744 (45%) | 0.009 |
| **Years of prior history*, median [Q25-Q75]** | 24 [11-36] | 24 [11-35] | 0.005 | 18 [8-29] | 18 [8-29] | 0.007 | 16 [7-27] | 17 [7-27] | 0.007 | 7 [4-16] | 7 [3-18] | 0.006 |
| **Number visits, median [Q25-Q75]** | 10 [5-18] | 10 [6-17] | | 8 [3-15] | 10 [5-15] | | 6 [2-15] | 8 [4-14] | | 2 [0-6] | 2 [0-6] | |
| **Number pcrs, median [Q25-Q75]** | 0[0-0] | 0[0-0] | | 0[0-0] | 0[0-0] | | 0[0-0] | 0[0-0] | | 0[0-0] | 0[0-0] | |
| **Comorbidities**, N(%)** | | | | | | | | | | | | |
| Anxiety | 15,706 (15%) | 15,578 (15%) | 0.007 | 57,951 (23%) | 55,594 (22%) | 0.018 | 7,672 (21%) | 7,363 (20%) | 0.021 | 96,918 (15%) | 96,528 (15%) | 0.004 |
| Asthma | 11,563 (11%) | 11,733 (11%) | 0.002 | 60,265 (24%) | 58,010 (23%) | 0.016 | 6,397 (17%) | 6,215 (17%) | 0.013 | 50,849 (8%) | 48,590 (8%) | 0.015 |
| Chronic kidney disease | 25,741 (24%) | 26,243 (25%) | 0.007 | 17,219 (7%) | 18,864 (7%) | 0.028 | 2,790 (7%) | 2,621 (7%) | 0.017 | 2,035 (0%) | 1,846 (0%) | 0.006 |
| COPD | 9,240 (9%) | 8,999 (8%) | 0.011 | 11,625 (5%) | 12,410 (5%) | 0.017 | 1,224 (3%) | 886 (2%) | 0.055 | 859 (0%) | 807 (0%) | 0.002 |
| Dementia | 6,683 (6%) | 5,727 (5%) | 0.040 | 2,225 (1%) | 1,309 (1%) | 0.043 | 618 (2%) | 529 (1%) | 0.019 | 112 (0%) | 107 (0%) | 0.001 |
| Depressive disorder | 12,529 (12%) | 12,450 (12%) | 0.005 | 52,883 (21%) | 50,765 (20%) | 0.016 | 6,588 (18%) | 6,431 (17%) | 0.011 | 71,832 (11%) | 71,139 (11%) | 0.005 |
| Diabetes | 20,227 (19%) | 20,182 (19%) | 0.005 | 32,557 (13%) | 32,989 (13%) | 0.008 | 3,153 (8%) | 3,212 (9%) | 0.006 | 8,183 (1%) | 7,675 (1%) | 0.008 |
| GERD | 6,148 (6%) | 5,937 (6%) | 0.011 | 12,436 (5%) | 12,160 (5%) | 0.003 | 1,552 (4%) | 1,509 (4%) | 0.006 | 9,586 (2%) | 10,173 (2%) | 0.007 |
| Heart failure | 6,643 (6%) | 6,329 (6%) | 0.014 | 4,618 (2%) | 3,987 (2%) | 0.018 | 767 (2%) | 552 (1%) | 0.044 | 474 (0%) | 344 (0%) | 0.008 |
| Hypertension | 56,732 (54%) | 57,023 (53%) | 0.003 | 62,628 (25%) | 63,500 (25%) | 0.013 | 7,802 (21%) | 7,697 (21%) | 0.007 | 11,603 (2%) | 12,098 (2%) | 0.005 |
| Hypothyroidism | 10,528 (10%) | 10,633 (10%) | 0.001 | 16,084 (6%) | 16,368 (6%) | 0.007 | 2,046 (5%) | 2,032 (5%) | 0.002 | 8,722 (1%) | 8,355 (1%) | 0.006 |
| Malignant neoplastic disease | 23,367 (22%) | 23,618 (22%) | 0.002 | 19,432 (8%) | 22,945 (9%) | 0.053 | 2,799 (8%) | 2,742 (7%) | 0.006 | 3,432 (1%) | 2,913 (0%) | 0.012 |

| | Cohort 1 | | | Cohort 2 | | | Cohort 3 | | | Cohort 4 | | |
|---|---|---|---|---|---|---|---|---|---|---|---|---|
| | **Unvaccinated** | **Vaccinated** | **ASMD** | **Unvaccinated** | **Vaccinated** | **ASMD** | **Unvaccinated** | **Vaccinated** | **ASMD** | **Unvaccinated** | **Vaccinated** | **ASMD** |
| Myocardial infarction | 5,451 (5%) | 5,473 (5%) | 0.001 | 6,811 (3%) | 7,856 (3%) | 0.026 | 704 (2%) | 637 (2%) | 0.013 | 607 (0%) | 569 (0%) | 0.002 |
| Osteoporosis | 10,923 (10%) | 10,997 (10%) | 0.000 | 6,664 (3%) | 6,208 (2%) | 0.010 | 1,166 (3%) | 1,014 (3%) | 0.024 | 653 (0%) | 702 (0%) | 0.002 |
| Pneumonia | 5,814 (5%) | 5,303 (5%) | 0.023 | 7,059 (3%) | 6,337 (3%) | 0.016 | 991 (3%) | 831 (2%) | 0.028 | 3,698 (1%) | 3,579 (1%) | 0.003 |
| Rheumatoid arthritis | 2,115 (2%) | 2,076 (2%) | 0.004 | 4,061 (2%) | 4,486 (2%) | 0.014 | 330 (1%) | 382 (1%) | 0.014 | 723 (0%) | 523 (0%) | 0.010 |
| Stroke | 5,304 (5%) | 4,911 (5%) | 0.019 | 5,139 (2%) | 5,592 (2%) | 0.014 | 657 (2%) | 560 (2%) | 0.021 | 828 (0%) | 669 (0%) | 0.007 |
| Venous thromboembolism | 6,564 (6%) | 6,603 (6%) | 0.000 | 7,760 (3%) | 7,984 (3%) | 0.007 | 980 (3%) | 965 (3%) | 0.003 | 2,860 (0%) | 1,602 (0%) | 0.034 |

The 4 cohorts represent vaccine rollout periods. *Calculated as the days of previous observation in the database before index date. **Assessed anytime before index date. ASMD = Absolute standardized mean difference, COPD = Chronic obstructive pulmonary disease, GERD = Gastro-Esophageal reflux disease

**Table S6: Characteristics of unweighted populations in CPRD AURUM,** database, stratified by staggered cohort and exposure status. Exposure is BNT162b2 vaccine.

| | Cohort 1 | | | Cohort 2 | | | Cohort 3 | | | Cohort 4 | | |
|---|---|---|---|---|---|---|---|---|---|---|---|---|
| | **Unvaccinated** | **Vaccinated** | **ASMD** | **Unvaccinated** | **Vaccinated** | **ASMD** | **Unvaccinated** | **Vaccinated** | **ASMD** | **Unvaccinated** | **Vaccinated** | **ASMD** |
| **N (individuals)** | 348,052 | 332,790 | | 1,976,163 | 594,262 | | 1,510,323 | 54,102 | | 2,014,161 | 1,335,671 | |
| **Age, median [Q25-Q75]** | 78 [76-82] | 82 [79-86] | 0.375 | 42 [32-55] | 67 [56-73] | 0.899 | 41 [31-53] | 58 [50-82] | 0.777 | 35 [27-45] | 31 [25-37] | 0.377 |
| **Sex: Female, N(%)** | 193,520 (56%) | 186,481 (56%) | 0.009 | 1,244,798 (63%) | 324,259 (55%) | 0.172 | 880,418 (58%) | 32,310 (60%) | 0.029 | 856,596 (43%) | 625,195 (47%) | 0.086 |
| **Years of prior history*, median [Q25-Q75]** | 23 [10-35] | 26 [13-38] | 0.081 | 15 [7-25] | 21 [9-32] | 0.261 | 13 [6-22] | 19 [8-30] | 0.308 | 8 [4-16] | 7 [3-19] | 0.012 |
| **Number visits, median [Q25-Q75]** | 10 [5-17] | 11 [6-18] | | 6 [2-11] | 11 [6-17] | | 4 [1-10] | 10 [5-17] | | 2 [0-5] | 3 [1-7] | |
| **Number pcrs, median [Q25-Q75]** | 0[0-0] | 0[0-0] | | 0[0-0] | 0[0-0] | | 0[0-0] | 0[0-0] | | 0[0-0] | 0[0-0] | |
| **Comorbidities**, N(%)** | | | | | | | | | | | | |
| Anxiety | 51,245 (15%) | 49,410 (15%) | 0.003 | 435,705 (22%) | 122,087 (21%) | 0.037 | 289,623 (19%) | 10,441 (19%) | 0.003 | 264,577 (13%) | 218,144 (16%) | 0.090 |
| Asthma | 36,625 (11%) | 37,889 (11%) | 0.028 | 501,289 (25%) | 116,596 (20%) | 0.138 | 294,101 (19%) | 8,668 (16%) | 0.090 | 137,548 (7%) | 114,378 (9%) | 0.065 |
| Chronic kidney disease | 71,413 (21%) | 86,899 (26%) | 0.133 | 55,136 (3%) | 56,923 (10%) | 0.285 | 24,372 (2%) | 6,692 (12%) | 0.431 | 18,579 (1%) | 2,664 (0%) | 0.097 |
| COPD | 28,829 (8%) | 28,317 (9%) | 0.008 | 29,793 (2%) | 39,595 (7%) | 0.263 | 13,430 (1%) | 2,063 (4%) | 0.194 | 10,501 (1%) | 942 (0%) | 0.083 |
| Dementia | 16,828 (5%) | 16,138 (5%) | 0.001 | 6,731 (0%) | 3,720 (1%) | 0.041 | 3,424 (0%) | 1,323 (2%) | 0.194 | 2,673 (0%) | 119 (0%) | 0.047 |
| Depressive disorder | 42,297 (12%) | 37,831 (11%) | 0.024 | 367,534 (19%) | 112,753 (19%) | 0.010 | 236,577 (16%) | 9,046 (17%) | 0.029 | 209,905 (10%) | 155,037 (12%) | 0.038 |
| Diabetes | 63,483 (18%) | 61,929 (19%) | 0.010 | 107,471 (5%) | 98,676 (17%) | 0.362 | 60,945 (4%) | 5,817 (11%) | 0.259 | 49,143 (2%) | 12,040 (1%) | 0.120 |
| GERD | 18,404 (5%) | 20,204 (6%) | 0.034 | 68,110 (3%) | 32,288 (5%) | 0.097 | 39,607 (3%) | 2,506 (5%) | 0.108 | 31,062 (2%) | 21,599 (2%) | 0.006 |
| Heart failure | 17,980 (5%) | 20,430 (6%) | 0.042 | 12,311 (1%) | 13,120 (2%) | 0.134 | 5,627 (0%) | 1,529 (3%) | 0.197 | 4,614 (0%) | 448 (0%) | 0.054 |
| Hypertension | 173,422 (50%) | 186,367 (56%) | 0.124 | 206,136 (10%) | 200,762 (34%) | 0.586 | 110,969 (7%) | 15,536 (29%) | 0.579 | 86,231 (4%) | 19,649 (1%) | 0.169 |
| Hypothyroidism | 31,595 (9%) | 34,512 (10%) | 0.044 | 78,935 (4%) | 44,456 (7%) | 0.150 | 43,876 (3%) | 3,642 (7%) | 0.179 | 30,913 (2%) | 18,516 (1%) | 0.012 |
| Malignant neoplastic disease | 69,247 (20%) | 82,610 (25%) | 0.118 | 60,370 (3%) | 76,103 (13%) | 0.367 | 29,693 (2%) | 6,311 (12%) | 0.392 | 22,630 (1%) | 5,104 (0%) | 0.086 |

| | Cohort 1 | | | Cohort 2 | | | Cohort 3 | | | Cohort 4 | | |
|---|---|---|---|---|---|---|---|---|---|---|---|---|
| | Unvaccinated | Vaccinated | ASMD | Unvaccinated | Vaccinated | ASMD | Unvaccinated | Vaccinated | ASMD | Unvaccinated | Vaccinated | ASMD |
| Myocardial infarction | 16,111 (5%) | 18,289 (5%) | 0.040 | 20,304 (1%) | 23,019 (4%) | 0.185 | 7,409 (0%) | 1,464 (3%) | 0.177 | 5,738 (0%) | 715 (0%) | 0.056 |
| Osteoporosis | 30,053 (9%) | 36,535 (11%) | 0.079 | 18,282 (1%) | 21,789 (4%) | 0.184 | 8,852 (1%) | 2,632 (5%) | 0.265 | 6,904 (0%) | 1,028 (0%) | 0.058 |
| Pneumonia | 17,134 (5%) | 16,297 (5%) | 0.001 | 31,724 (2%) | 17,567 (3%) | 0.091 | 17,791 (1%) | 1,658 (3%) | 0.131 | 13,263 (1%) | 7,793 (1%) | 0.010 |
| Rheumatoid arthritis | 6,613 (2%) | 6,693 (2%) | 0.008 | 14,823 (1%) | 12,878 (2%) | 0.118 | 5,533 (0%) | 667 (1%) | 0.097 | 3,692 (0%) | 891 (0%) | 0.033 |
| Stroke | 15,325 (4%) | 15,601 (5%) | 0.014 | 16,962 (1%) | 15,056 (3%) | 0.130 | 7,431 (0%) | 1,281 (2%) | 0.158 | 6,019 (0%) | 1,022 (0%) | 0.051 |
| Venous thromboembolism | 19,679 (6%) | 21,549 (6%) | 0.034 | 28,882 (1%) | 23,632 (4%) | 0.155 | 13,639 (1%) | 1,980 (4%) | 0.185 | 12,941 (1%) | 2,541 (0%) | 0.070 |

The 4 cohorts represent vaccine rollout periods. *Calculated as the days of previous observation in the database before index date. **Assessed anytime before index date. ASMD = Absolute standardized mean difference, COPD = Chronic obstructive pulmonary disease, GERD = Gastro-Esophageal reflux disease

**Table S7: Characteristics of weighted populations in CPRD GOLD,** database, stratified by staggered cohort and exposure status. Exposure is any COVID-19 vaccine.

| | Cohort 1 | | | Cohort 2 | | | Cohort 3 | | | Cohort 4 | | |
|---|---|---|---|---|---|---|---|---|---|---|---|---|
| | **Unvaccinated** | **Vaccinated** | **ASMD** | **Unvaccinated** | **Vaccinated** | **ASMD** | **Unvaccinated** | **Vaccinated** | **ASMD** | **Unvaccinated** | **Vaccinated** | **ASMD** |
| **N (individuals)** | 40,084 | 40,014 | | 111,674 | 111,393 | | 124,883 | 124,955 | | 196,235 | 195,302 | |
| **Age, median [Q25-Q75]** | 82 [78-86] | 82 [78-86] | 0.004 | 62 [50-69] | 62 [50-69] | 0.005 | 50 [41-58] | 52 [41-58] | 0.000 | 34 [26-42] | 34 [26-43] | 0.004 |
| **Sex: Female, N(%)** | 23,206 (58%) | 23,175 (58%) | 0.000 | 63,290 (57%) | 63,079 (57%) | 0.001 | 63,572 (51%) | 63,955 (51%) | 0.006 | 84,762 (43%) | 85,083 (44%) | 0.007 |
| **Years of prior history*, median [Q25-Q75]** | 17 [13-20] | 18 [13-20] | 0.001 | 17 [11-19] | 17 [11-19] | 0.005 | 17 [9-19] | 17 [9-19] | 0.000 | 13 [6-18] | 14 [5-18] | 0.006 |
| **Number visits, median [Q25-Q75]** | 12 [7-20] | 12 [8-20] | | 8 [3-15] | 10 [5-15] | | 4 [0-10] | 4 [1-10] | | 0 [0-5] | 2 [0-5] | |
| **Number pcrs, median [Q25-Q75]** | 0[0-0] | 0[0-0] | | 0[0-0] | 0[0-0] | | 0[0-0] | 0[0-0] | | 0[0-0] | 0[0-0] | |
| **Comorbidities**, N(%)** | | | | | | | | | | | | |
| Anxiety | 4,933 (12%) | 4,920 (12%) | 0.000 | 22,884 (20%) | 22,299 (20%) | 0.012 | 24,993 (20%) | 24,695 (20%) | 0.006 | 31,360 (16%) | 30,689 (16%) | 0.007 |
| Asthma | 3,911 (10%) | 3,931 (10%) | 0.002 | 18,514 (17%) | 18,209 (16%) | 0.006 | 17,431 (14%) | 17,388 (14%) | 0.001 | 14,817 (8%) | 14,596 (7%) | 0.003 |
| Chronic kidney disease | 8,998 (22%) | 8,953 (22%) | 0.002 | 6,848 (6%) | 7,431 (7%) | 0.022 | 2,120 (2%) | 2,287 (2%) | 0.010 | 922 (0%) | 807 (0%) | 0.009 |
| COPD | 3,439 (9%) | 3,341 (8%) | 0.008 | 5,972 (5%) | 6,496 (6%) | 0.021 | 2,089 (2%) | 1,938 (2%) | 0.010 | 734 (0%) | 688 (0%) | 0.004 |
| Dementia | 2,634 (7%) | 2,332 (6%) | 0.031 | 1,383 (1%) | 865 (1%) | 0.046 | 283 (0%) | 313 (0%) | 0.005 | 78 (0%) | 134 (0%) | 0.013 |
| Depressive disorder | 3,783 (9%) | 3,705 (9%) | 0.006 | 20,874 (19%) | 20,246 (18%) | 0.013 | 22,015 (18%) | 21,682 (17%) | 0.007 | 23,010 (12%) | 22,630 (12%) | 0.004 |
| Diabetes | 5,928 (15%) | 5,802 (14%) | 0.008 | 11,000 (10%) | 11,657 (10%) | 0.020 | 5,345 (4%) | 5,651 (5%) | 0.012 | 2,586 (1%) | 2,366 (1%) | 0.009 |
| GERD | 1,930 (5%) | 1,970 (5%) | 0.005 | 4,521 (4%) | 4,479 (4%) | 0.001 | 3,343 (3%) | 3,346 (3%) | 0.000 | 2,657 (1%) | 2,614 (1%) | 0.001 |
| Heart failure | 2,417 (6%) | 2,205 (6%) | 0.022 | 2,002 (2%) | 2,029 (2%) | 0.002 | 639 (1%) | 614 (0%) | 0.003 | 250 (0%) | 207 (0%) | 0.006 |
| Hypertension | 14,288 (36%) | 14,282 (36%) | 0.001 | 22,488 (20%) | 22,695 (20%) | 0.006 | 11,108 (9%) | 11,313 (9%) | 0.006 | 4,318 (2%) | 4,619 (2%) | 0.011 |
| Hypothyroidism | 3,151 (8%) | 3,155 (8%) | 0.001 | 6,321 (6%) | 6,361 (6%) | 0.002 | 4,134 (3%) | 4,055 (3%) | 0.004 | 2,475 (1%) | 2,621 (1%) | 0.007 |
| Malignant neoplastic disease | 8,302 (21%) | 8,283 (21%) | 0.000 | 8,661 (8%) | 10,177 (9%) | 0.050 | 3,813 (3%) | 3,777 (3%) | 0.002 | 1,614 (1%) | 1,676 (1%) | 0.004 |

| | Cohort 1 | | | Cohort 2 | | | Cohort 3 | | | Cohort 4 | | |
|---|---|---|---|---|---|---|---|---|---|---|---|---|
| | **Unvaccinated** | **Vaccinated** | **ASMD** | **Unvaccinated** | **Vaccinated** | **ASMD** | **Unvaccinated** | **Vaccinated** | **ASMD** | **Unvaccinated** | **Vaccinated** | **ASMD** |
| Myocardial infarction | 2,119 (5%) | 1,984 (5%) | 0.015 | 3,308 (3%) | 3,740 (3%) | 0.023 | 1,193 (1%) | 1,267 (1%) | 0.006 | 430 (0%) | 343 (0%) | 0.010 |
| Osteoporosis | 3,702 (9%) | 3,670 (9%) | 0.002 | 3,154 (3%) | 3,179 (3%) | 0.002 | 1,038 (1%) | 1,030 (1%) | 0.001 | 363 (0%) | 375 (0%) | 0.002 |
| Pneumonia | 1,654 (4%) | 1,452 (4%) | 0.026 | 2,424 (2%) | 2,211 (2%) | 0.013 | 1,401 (1%) | 1,314 (1%) | 0.007 | 1,035 (1%) | 1,055 (1%) | 0.002 |
| Rheumatoid arthritis | 637 (2%) | 624 (2%) | 0.002 | 1,618 (1%) | 1,818 (2%) | 0.015 | 551 (0%) | 760 (1%) | 0.023 | 280 (0%) | 157 (0%) | 0.019 |
| Stroke | 2,103 (5%) | 1,866 (5%) | 0.027 | 2,468 (2%) | 2,512 (2%) | 0.003 | 916 (1%) | 981 (1%) | 0.006 | 385 (0%) | 356 (0%) | 0.003 |
| Venous thromboembolism | 1,732 (4%) | 1,648 (4%) | 0.010 | 2,639 (2%) | 2,596 (2%) | 0.002 | 1,466 (1%) | 1,638 (1%) | 0.012 | 959 (0%) | 518 (0%) | 0.036 |

The 4 cohorts represent vaccine rollout periods. *Calculated as the days of previous observation in the database before index date. **Assessed anytime before index date. ASMD = Absolute standardized mean difference, COPD = Chronic obstructive pulmonary disease, GERD = Gastro-Esophageal reflux disease

**Table S8: Characteristics of unweighted populations in CPRD GOLD,** database, stratified by staggered cohort and exposure status. Exposure is any COVID-19 vaccine.

| | **Cohort 1** | | | **Cohort 2** | | | **Cohort 3** | | | **Cohort 4** | | |
|---|---|---|---|---|---|---|---|---|---|---|---|---|
| | **Unvaccinated** | **Vaccinated** | **ASMD** | **Unvaccinated** | **Vaccinated** | **ASMD** | **Unvaccinated** | **Vaccinated** | **ASMD** | **Unvaccinated** | **Vaccinated** | **ASMD** |
| **N (individuals)** | 169,100 | 118,507 | | 583,399 | 486,619 | | 417,996 | 462,832 | | 469,876 | 550,437 | |
| **Age, median [Q25-Q75]** | 78 [76-82] | 84 [81-87] | 0.550 | 45 [33-58] | 71 [66-75] | 1.117 | 42 [31-55] | 56 [52-61] | 0.546 | 37 [28-49] | 35 [27-43] | 0.250 |
| **Sex: Female, N(%)** | 93,719 (55%) | 68,258 (58%) | 0.044 | 344,747 (59%) | 262,113 (54%) | 0.106 | 230,567 (55%) | 230,957 (50%) | 0.105 | 208,184 (44%) | 249,603 (45%) | 0.021 |
| **Years of prior history*, median [Q25-Q75]** | 17 [13-20] | 18 [14-20] | 0.023 | 17 [10-19] | 17 [13-20] | 0.085 | 17 [9-19] | 17 [10-19] | 0.055 | 13 [6-18] | 14 [6-18] | 0.020 |
| **Number visits, median [Q25-Q75]** | 11 [6-18] | 14 [9-21] | | 5 [1-11] | 11 [6-17] | | 3 [0-8] | 6 [2-12] | | 0 [0-4] | 2 [0-6] | |
| **Number pcrs, median [Q25-Q75]** | 0[0-0] | 0[0-0] | | 0[0-0] | 0[0-0] | | 0[0-0] | 0[0-0] | | 0[0-0] | 0[0-0] | |
| **Comorbidities**, N(%)** | | | | | | | | | | | | |
| Anxiety | 20,097 (12%) | 14,878 (13%) | 0.020 | 132,282 (23%) | 78,442 (16%) | 0.166 | 83,022 (20%) | 83,629 (18%) | 0.046 | 65,306 (14%) | 88,512 (16%) | 0.061 |
| Asthma | 15,573 (9%) | 12,090 (10%) | 0.034 | 119,520 (20%) | 61,821 (13%) | 0.210 | 68,746 (16%) | 52,312 (11%) | 0.149 | 30,811 (7%) | 46,774 (8%) | 0.074 |
| Chronic kidney disease | 29,343 (17%) | 29,934 (25%) | 0.194 | 15,484 (3%) | 49,550 (10%) | 0.311 | 6,676 (2%) | 8,145 (2%) | 0.013 | 4,746 (1%) | 1,376 (0%) | 0.096 |
| COPD | 14,248 (8%) | 9,602 (8%) | 0.012 | 12,247 (2%) | 39,371 (8%) | 0.275 | 5,114 (1%) | 6,699 (1%) | 0.020 | 3,716 (1%) | 953 (0%) | 0.089 |
| Dementia | 6,678 (4%) | 6,860 (6%) | 0.086 | 1,986 (0%) | 6,557 (1%) | 0.110 | 1,188 (0%) | 636 (0%) | 0.032 | 774 (0%) | 159 (0%) | 0.044 |
| Depressive disorder | 16,339 (10%) | 10,720 (9%) | 0.021 | 111,568 (19%) | 72,063 (15%) | 0.115 | 66,705 (16%) | 77,560 (17%) | 0.022 | 50,341 (11%) | 64,215 (12%) | 0.030 |
| Diabetes | 23,579 (14%) | 17,895 (15%) | 0.033 | 29,125 (5%) | 66,688 (14%) | 0.303 | 12,407 (3%) | 25,465 (6%) | 0.126 | 9,225 (2%) | 5,012 (1%) | 0.089 |
| GERD | 7,610 (5%) | 6,174 (5%) | 0.033 | 18,053 (3%) | 22,884 (5%) | 0.083 | 8,864 (2%) | 15,586 (3%) | 0.076 | 5,920 (1%) | 8,482 (2%) | 0.024 |
| Heart failure | 7,475 (4%) | 7,118 (6%) | 0.071 | 4,013 (1%) | 12,939 (3%) | 0.154 | 1,732 (0%) | 2,410 (1%) | 0.016 | 1,274 (0%) | 293 (0%) | 0.054 |
| Hypertension | 56,298 (33%) | 44,709 (38%) | 0.093 | 58,662 (10%) | 139,644 (29%) | 0.485 | 24,457 (6%) | 64,451 (14%) | 0.273 | 16,440 (3%) | 11,991 (2%) | 0.080 |
| Hypothyroidism | 12,126 (7%) | 9,825 (8%) | 0.042 | 22,026 (4%) | 31,923 (7%) | 0.126 | 10,616 (3%) | 19,001 (4%) | 0.087 | 6,676 (1%) | 7,790 (1%) | 0.000 |
| Malignant neoplastic disease | 30,687 (18%) | 26,725 (23%) | 0.110 | 19,781 (3%) | 65,779 (14%) | 0.370 | 8,870 (2%) | 19,474 (4%) | 0.119 | 5,916 (1%) | 4,164 (1%) | 0.050 |

| | Cohort 1 | | | Cohort 2 | | | Cohort 3 | | | Cohort 4 | | |
|---|---|---|---|---|---|---|---|---|---|---|---|---|
| | **Unvaccinated** | **Vaccinated** | **ASMD** | **Unvaccinated** | **Vaccinated** | **ASMD** | **Unvaccinated** | **Vaccinated** | **ASMD** | **Unvaccinated** | **Vaccinated** | **ASMD** |
| Myocardial infarction | 7,707 (5%) | 6,298 (5%) | 0.035 | 8,409 (1%) | 20,200 (4%) | 0.165 | 2,665 (1%) | 5,556 (1%) | 0.059 | 2,003 (0%) | 501 (0%) | 0.066 |
| Osteoporosis | 12,736 (8%) | 11,677 (10%) | 0.082 | 5,836 (1%) | 23,843 (5%) | 0.232 | 2,580 (1%) | 4,797 (1%) | 0.046 | 1,855 (0%) | 637 (0%) | 0.055 |
| Pneumonia | 5,401 (3%) | 4,381 (4%) | 0.028 | 7,250 (1%) | 12,120 (2%) | 0.092 | 3,822 (1%) | 4,889 (1%) | 0.014 | 2,522 (1%) | 2,797 (1%) | 0.004 |
| Rheumatoid arthritis | 2,565 (2%) | 1,833 (2%) | 0.002 | 4,013 (1%) | 9,000 (2%) | 0.104 | 1,294 (0%) | 2,566 (1%) | 0.037 | 877 (0%) | 316 (0%) | 0.037 |
| Stroke | 7,214 (4%) | 5,843 (5%) | 0.032 | 5,886 (1%) | 14,921 (3%) | 0.146 | 2,170 (1%) | 3,998 (1%) | 0.042 | 1,591 (0%) | 620 (0%) | 0.048 |
| Venous thromboembolism | 6,188 (4%) | 5,096 (4%) | 0.033 | 7,342 (1%) | 14,077 (3%) | 0.115 | 3,067 (1%) | 6,732 (1%) | 0.069 | 2,611 (1%) | 1,018 (0%) | 0.061 |

The 4 cohorts represent vaccine rollout periods. *Calculated as the days of previous observation in the database before index date. **Assessed anytime before index date. ASMD = Absolute standardized mean difference, COPD = Chronic obstructive pulmonary disease, GERD = Gastro-Esophageal reflux disease

**Table S9: Characteristics of weighted populations in CPRD GOLD,** database, stratified by staggered cohort and exposure status. Exposure is ChAdOx1 vaccine.

| | **Cohort 1** | | | **Cohort 2** | | | **Cohort 3** | | | **Cohort 4** | | |
|---|---|---|---|---|---|---|---|---|---|---|---|---|
| | **Unvaccinated** | **Vaccinated** | **ASMD** | **Unvaccinated** | **Vaccinated** | **ASMD** | **Unvaccinated** | **Vaccinated** | **ASMD** | **Unvaccinated** | **Vaccinated** | **ASMD** |
| **N (individuals)** | 29,163 | 29,082 | | 82,091 | 82,226 | | 116,090 | 115,912 | | 70,312 | 70,823 | |
| **Age, median [Q25-Q75]** | 82 [79-87] | 82 [80-87] | 0.002 | 64 [53-71] | 64 [53-70] | 0.001 | 52 [42-58] | 52 [42-58] | 0.003 | 42 [39-48] | 42 [38-48] | 0.000 |
| **Sex: Female, N(%)** | 17,126 (59%) | 17,076 (59%) | 0.000 | 45,269 (55%) | 45,487 (55%) | 0.003 | 58,599 (50%) | 58,799 (51%) | 0.005 | 29,480 (42%) | 29,458 (42%) | 0.007 |
| **Years of prior history*, median [Q25-Q75]** | 17 [13-20] | 17 [14-20] | 0.001 | 17 [11-19] | 17 [11-19] | 0.002 | 17 [9-19] | 17 [8-19] | 0.001 | 14 [7-18] | 14 [6-18] | 0.001 |
| **Number visits, median [Q25-Q75]** | 12 [7-21] | 12 [8-20] | | 8 [3-16] | 10 [5-16] | | 4 [0-10] | 4 [1-10] | | 0 [0-6] | 2 [0-6] | |
| **Number pcrs, median [Q25-Q75]** | 0[0-0] | 0[0-0] | | 0[0-0] | 0[0-0] | | 0[0-0] | 0[0-0] | | 0[0-0] | 0[0-0] | |
| **Comorbidities**, N(%)** | | | | | | | | | | | | |
| Anxiety | 3,481 (12%) | 3,496 (12%) | 0.003 | 16,202 (20%) | 15,970 (19%) | 0.008 | 23,083 (20%) | 22,695 (20%) | 0.008 | 12,154 (17%) | 11,947 (17%) | 0.011 |
| Asthma | 2,863 (10%) | 2,870 (10%) | 0.002 | 13,358 (16%) | 13,192 (16%) | 0.006 | 16,057 (14%) | 16,029 (14%) | 0.000 | 4,767 (7%) | 4,744 (7%) | 0.003 |
| Chronic kidney disease | 6,959 (24%) | 6,928 (24%) | 0.001 | 5,850 (7%) | 6,384 (8%) | 0.024 | 2,031 (2%) | 2,104 (2%) | 0.005 | 616 (1%) | 517 (1%) | 0.016 |
| COPD | 2,556 (9%) | 2,485 (9%) | 0.008 | 5,069 (6%) | 5,523 (7%) | 0.022 | 1,969 (2%) | 1,874 (2%) | 0.006 | 539 (1%) | 478 (1%) | 0.011 |
| Dementia | 2,042 (7%) | 1,794 (6%) | 0.034 | 1,319 (2%) | 826 (1%) | 0.053 | 263 (0%) | 265 (0%) | 0.000 | 59 (0%) | 111 (0%) | 0.021 |
| Depressive disorder | 2,641 (9%) | 2,608 (9%) | 0.003 | 14,875 (18%) | 14,558 (18%) | 0.011 | 20,497 (18%) | 20,298 (18%) | 0.004 | 10,492 (15%) | 10,218 (14%) | 0.014 |
| Diabetes | 4,346 (15%) | 4,273 (15%) | 0.006 | 8,933 (11%) | 9,524 (12%) | 0.022 | 5,139 (4%) | 5,350 (5%) | 0.009 | 1,459 (2%) | 1,421 (2%) | 0.005 |
| GERD | 1,366 (5%) | 1,380 (5%) | 0.003 | 3,514 (4%) | 3,414 (4%) | 0.006 | 3,153 (3%) | 3,136 (3%) | 0.001 | 1,258 (2%) | 1,297 (2%) | 0.003 |
| Heart failure | 1,844 (6%) | 1,699 (6%) | 0.020 | 1,793 (2%) | 1,720 (2%) | 0.006 | 613 (1%) | 601 (1%) | 0.001 | 177 (0%) | 151 (0%) | 0.008 |
| Hypertension | 10,466 (36%) | 10,581 (36%) | 0.010 | 17,843 (22%) | 18,115 (22%) | 0.007 | 10,489 (9%) | 10,714 (9%) | 0.007 | 3,119 (4%) | 3,233 (5%) | 0.006 |
| Hypothyroidism | 2,338 (8%) | 2,360 (8%) | 0.004 | 4,857 (6%) | 4,837 (6%) | 0.001 | 3,897 (3%) | 3,742 (3%) | 0.007 | 1,432 (2%) | 1,412 (2%) | 0.003 |
| Malignant neoplastic disease | 6,210 (21%) | 6,164 (21%) | 0.002 | 7,090 (9%) | 8,270 (10%) | 0.049 | 3,588 (3%) | 3,590 (3%) | 0.000 | 1,059 (2%) | 1,117 (2%) | 0.006 |
| Myocardial infarction | 1,569 (5%) | 1,500 (5%) | 0.010 | 2,720 (3%) | 3,083 (4%) | 0.024 | 1,155 (1%) | 1,202 (1%) | 0.004 | 319 (0%) | 242 (0%) | 0.018 |

| | Cohort 1 | | | Cohort 2 | | | Cohort 3 | | | Cohort 4 | | |
|---|---|---|---|---|---|---|---|---|---|---|---|---|
| | **Unvaccinated** | **Vaccinated** | **ASMD** | **Unvaccinated** | **Vaccinated** | **ASMD** | **Unvaccinated** | **Vaccinated** | **ASMD** | **Unvaccinated** | **Vaccinated** | **ASMD** |
| Osteoporosis | 2,838 (10%) | 2,846 (10%) | 0.002 | 2,715 (3%) | 2,668 (3%) | 0.004 | 944 (1%) | 986 (1%) | 0.004 | 252 (0%) | 281 (0%) | 0.006 |
| Pneumonia | 1,287 (4%) | 1,147 (4%) | 0.024 | 1,997 (2%) | 1,883 (2%) | 0.009 | 1,307 (1%) | 1,245 (1%) | 0.005 | 487 (1%) | 473 (1%) | 0.003 |
| Rheumatoid arthritis | 464 (2%) | 445 (2%) | 0.005 | 1,280 (2%) | 1,491 (2%) | 0.020 | 514 (0%) | 743 (1%) | 0.027 | 184 (0%) | 120 (0%) | 0.020 |
| Stroke | 1,600 (5%) | 1,421 (5%) | 0.027 | 2,073 (3%) | 2,122 (3%) | 0.003 | 873 (1%) | 963 (1%) | 0.009 | 263 (0%) | 257 (0%) | 0.002 |
| Venous thromboembolism | 1,339 (5%) | 1,191 (4%) | 0.024 | 2,112 (3%) | 2,219 (3%) | 0.008 | 1,407 (1%) | 1,572 (1%) | 0.013 | 590 (1%) | 297 (0%) | 0.053 |

The 4 cohorts represent vaccine rollout periods. *Calculated as the days of previous observation in the database before index date. **Assessed anytime before index date. ASMD = Absolute standardized mean difference, COPD = Chronic obstructive pulmonary disease, GERD = Gastro-Esophageal reflux disease

**Table S10: Characteristics of unweighted populations in CPRD GOLD,** database, stratified by staggered cohort and exposure status. Exposure is ChAdOx1 vaccine.

| | Cohort 1 | | | Cohort 2 | | | Cohort 3 | | | Cohort 4 | | |
|---|---|---|---|---|---|---|---|---|---|---|---|---|
| | **Unvaccinated** | **Vaccinated** | **ASMD** | **Unvaccinated** | **Vaccinated** | **ASMD** | **Unvaccinated** | **Vaccinated** | **ASMD** | **Unvaccinated** | **Vaccinated** | **ASMD** |
| **N (individuals)** | 168,972 | 82,406 | | 582,791 | 302,999 | | 418,184 | 423,876 | | 485,154 | 147,744 | |
| **Age, median [Q25-Q75]** | 78 [76-82] | 84 [82-88] | 0.657 | 45 [33-58] | 71 [66-76] | 1.153 | 42 [31-55] | 56 [52-61] | 0.560 | 37 [28-49] | 44 [41-48] | 0.262 |
| **Sex: Female, N(%)** | 93,648 (55%) | 47,915 (58%) | 0.055 | 344,408 (59%) | 162,753 (54%) | 0.109 | 230,680 (55%) | 210,283 (50%) | 0.111 | 215,080 (44%) | 64,999 (44%) | 0.007 |
| **Years of prior history*, median [Q25-Q75]** | 17 [13-20] | 17 [14-20] | 0.016 | 17 [10-19] | 17 [13-19] | 0.090 | 17 [9-19] | 17 [10-19] | 0.047 | 13 [6-18] | 14 [7-18] | 0.054 |
| **Number visits, median [Q25-Q75]** | 11 [6-18] | 14 [9-21] | | 5 [1-11] | 11 [7-18] | | 3 [0-8] | 6 [2-12] | | 0 [0-4] | 3 [0-7] | |
| **Number pcrs, median [Q25-Q75]** | 0[0-0] | 0[0-0] | | 0[0-0] | 0[0-0] | | 0[0-0] | 0[0-0] | | 0[0-0] | 0[0-0] | |
| **Comorbidities**, N(%)** | | | | | | | | | | | | |
| Anxiety | 20,087 (12%) | 9,923 (12%) | 0.005 | 132,145 (23%) | 49,010 (16%) | 0.165 | 83,054 (20%) | 75,258 (18%) | 0.054 | 66,918 (14%) | 24,601 (17%) | 0.080 |
| Asthma | 15,567 (9%) | 8,491 (10%) | 0.037 | 119,442 (20%) | 39,988 (13%) | 0.196 | 68,792 (16%) | 48,034 (11%) | 0.148 | 31,538 (7%) | 10,517 (7%) | 0.025 |
| Chronic kidney disease | 29,280 (17%) | 21,564 (26%) | 0.216 | 15,435 (3%) | 34,168 (11%) | 0.344 | 6,675 (2%) | 7,553 (2%) | 0.014 | 4,947 (1%) | 739 (1%) | 0.060 |
| COPD | 14,234 (8%) | 6,837 (8%) | 0.005 | 12,242 (2%) | 27,472 (9%) | 0.307 | 5,122 (1%) | 6,334 (1%) | 0.023 | 3,834 (1%) | 640 (0%) | 0.046 |
| Dementia | 6,664 (4%) | 4,773 (6%) | 0.086 | 1,998 (0%) | 5,124 (2%) | 0.135 | 1,201 (0%) | 548 (0%) | 0.035 | 831 (0%) | 126 (0%) | 0.024 |
| Depressive disorder | 16,333 (10%) | 6,998 (8%) | 0.041 | 111,526 (19%) | 45,154 (15%) | 0.113 | 66,763 (16%) | 70,703 (17%) | 0.019 | 51,687 (11%) | 21,711 (15%) | 0.122 |
| Diabetes | 23,547 (14%) | 12,387 (15%) | 0.031 | 29,089 (5%) | 43,647 (14%) | 0.322 | 12,392 (3%) | 24,219 (6%) | 0.135 | 9,455 (2%) | 2,254 (2%) | 0.032 |
| GERD | 7,613 (5%) | 4,054 (5%) | 0.020 | 18,044 (3%) | 14,294 (5%) | 0.084 | 8,869 (2%) | 14,447 (3%) | 0.079 | 6,051 (1%) | 2,883 (2%) | 0.056 |
| Heart failure | 7,468 (4%) | 5,058 (6%) | 0.077 | 4,016 (1%) | 9,180 (3%) | 0.174 | 1,738 (0%) | 2,308 (1%) | 0.019 | 1,316 (0%) | 191 (0%) | 0.032 |
| Hypertension | 56,262 (33%) | 31,217 (38%) | 0.096 | 58,578 (10%) | 88,833 (29%) | 0.499 | 24,450 (6%) | 59,554 (14%) | 0.277 | 16,971 (3%) | 6,761 (5%) | 0.055 |
| Hypothyroidism | 12,132 (7%) | 7,042 (9%) | 0.051 | 21,999 (4%) | 20,521 (7%) | 0.134 | 10,622 (3%) | 17,382 (4%) | 0.087 | 6,848 (1%) | 3,245 (2%) | 0.059 |
| Malignant neoplastic disease | 30,670 (18%) | 18,950 (23%) | 0.120 | 19,707 (3%) | 42,687 (14%) | 0.386 | 8,893 (2%) | 17,903 (4%) | 0.120 | 6,143 (1%) | 2,192 (1%) | 0.019 |

| | Cohort 1 | | | Cohort 2 | | | Cohort 3 | | | Cohort 4 | | |
|---|---|---|---|---|---|---|---|---|---|---|---|---|
| | **Unvaccinated** | **Vaccinated** | **ASMD** | **Unvaccinated** | **Vaccinated** | **ASMD** | **Unvaccinated** | **Vaccinated** | **ASMD** | **Unvaccinated** | **Vaccinated** | **ASMD** |
| Myocardial infarction | 7,704 (5%) | 4,500 (5%) | 0.041 | 8,413 (1%) | 13,456 (4%) | 0.178 | 2,660 (1%) | 5,280 (1%) | 0.063 | 2,057 (0%) | 330 (0%) | 0.035 |
| Osteoporosis | 12,732 (8%) | 8,564 (10%) | 0.100 | 5,847 (1%) | 16,117 (5%) | 0.249 | 2,576 (1%) | 4,330 (1%) | 0.045 | 1,923 (0%) | 408 (0%) | 0.021 |
| Pneumonia | 5,401 (3%) | 3,231 (4%) | 0.039 | 7,241 (1%) | 8,604 (3%) | 0.113 | 3,814 (1%) | 4,527 (1%) | 0.016 | 2,615 (1%) | 921 (1%) | 0.011 |
| Rheumatoid arthritis | 2,559 (2%) | 1,257 (2%) | 0.001 | 4,017 (1%) | 5,999 (2%) | 0.113 | 1,286 (0%) | 2,405 (1%) | 0.039 | 918 (0%) | 191 (0%) | 0.015 |
| Stroke | 7,200 (4%) | 4,193 (5%) | 0.039 | 5,885 (1%) | 10,213 (3%) | 0.162 | 2,167 (1%) | 3,789 (1%) | 0.045 | 1,625 (0%) | 369 (0%) | 0.016 |
| Venous thromboembolism | 6,185 (4%) | 3,550 (4%) | 0.033 | 7,335 (1%) | 9,466 (3%) | 0.128 | 3,072 (1%) | 6,339 (1%) | 0.073 | 2,712 (1%) | 460 (0%) | 0.038 |

The 4 cohorts represent vaccine rollout periods. *Calculated as the days of previous observation in the database before index date. **Assessed anytime before index date. ASMD = Absolute standardized mean difference, COPD = Chronic obstructive pulmonary disease, GERD = Gastro-Esophageal reflux disease

**Table S11: Characteristics of weighted populations in CPRD GOLD,** database, stratified by staggered cohort and exposure status. Exposure is BNT162b2 vaccine.

| | Cohort 1 | | | Cohort 2 | | | Cohort 3 | | | Cohort 4 | | |
|---|---|---|---|---|---|---|---|---|---|---|---|---|
| | **Unvaccinated** | **Vaccinated** | **ASMD** | **Unvaccinated** | **Vaccinated** | **ASMD** | **Unvaccinated** | **Vaccinated** | **ASMD** | **Unvaccinated** | **Vaccinated** | **ASMD** |
| **N (individuals)** | 15,295 | 15,304 | | 60,813 | 61,113 | | 24,008 | 24,157 | | 151,713 | 152,100 | |
| **Age, median [Q25-Q75]** | 80 [77-85] | 80 [77-85] | 0.000 | 64 [53-70] | 66 [52-70] | 0.008 | 52 [44-58] | 52 [44-58] | 0.004 | 32 [24-38] | 32 [24-38] | 0.003 |
| **Sex: Female, N(%)** | 8,644 (57%) | 8,657 (57%) | 0.001 | 34,612 (57%) | 34,776 (57%) | 0.000 | 12,846 (54%) | 12,993 (54%) | 0.006 | 66,947 (44%) | 67,555 (44%) | 0.006 |
| **Years of prior history*, median [Q25-Q75]** | 17 [12-20] | 18 [12-21] | 0.001 | 17 [12-19] | 17 [11-19] | 0.003 | 17 [11-19] | 17 [11-19] | 0.004 | 13 [6-18] | 14 [5-18] | 0.001 |
| **Number visits, median [Q25-Q75]** | 12 [7-20] | 12 [8-20] | | 8 [3-15] | 8 [4-15] | | 4 [0-10] | 4 [1-10] | | 0 [0-5] | 2 [0-5] | |
| **Number pcrs, median [Q25-Q75]** | 0[0-0] | 0[0-0] | | 0[0-0] | 0[0-0] | | 0[0-0] | 0[0-0] | | 0[0-0] | 0[0-0] | |
| **Comorbidities**, N(%)** | | | | | | | | | | | | |
| Anxiety | 1,969 (13%) | 1,940 (13%) | 0.006 | 11,790 (19%) | 11,914 (19%) | 0.003 | 5,342 (22%) | 5,235 (22%) | 0.014 | 24,455 (16%) | 23,928 (16%) | 0.011 |
| Asthma | 1,498 (10%) | 1,488 (10%) | 0.002 | 9,126 (15%) | 9,093 (15%) | 0.004 | 2,926 (12%) | 2,985 (12%) | 0.005 | 12,195 (8%) | 11,929 (8%) | 0.007 |
| Chronic kidney disease | 3,112 (20%) | 3,057 (20%) | 0.009 | 3,563 (6%) | 3,764 (6%) | 0.013 | 398 (2%) | 407 (2%) | 0.002 | 468 (0%) | 346 (0%) | 0.016 |
| COPD | 1,190 (8%) | 1,151 (8%) | 0.010 | 3,222 (5%) | 3,237 (5%) | 0.000 | 293 (1%) | 274 (1%) | 0.008 | 315 (0%) | 250 (0%) | 0.010 |
| Dementia | 1,087 (7%) | 864 (6%) | 0.060 | 646 (1%) | 355 (1%) | 0.053 | 55 (0%) | 80 (0%) | 0.019 | 21 (0%) | 28 (0%) | 0.004 |
| Depressive disorder | 1,527 (10%) | 1,496 (10%) | 0.007 | 10,709 (18%) | 10,912 (18%) | 0.006 | 4,382 (18%) | 4,343 (18%) | 0.007 | 17,112 (11%) | 16,594 (11%) | 0.012 |
| Diabetes | 2,193 (14%) | 2,173 (14%) | 0.004 | 6,057 (10%) | 6,199 (10%) | 0.006 | 883 (4%) | 858 (4%) | 0.007 | 1,609 (1%) | 1,271 (1%) | 0.023 |
| GERD | 880 (6%) | 872 (6%) | 0.003 | 2,440 (4%) | 2,505 (4%) | 0.004 | 659 (3%) | 706 (3%) | 0.011 | 1,935 (1%) | 1,961 (1%) | 0.001 |
| Heart failure | 857 (6%) | 748 (5%) | 0.032 | 1,097 (2%) | 933 (2%) | 0.022 | 119 (0%) | 77 (0%) | 0.027 | 107 (0%) | 77 (0%) | 0.008 |
| Hypertension | 5,338 (35%) | 5,312 (35%) | 0.004 | 12,828 (21%) | 12,901 (21%) | 0.000 | 2,597 (11%) | 2,645 (11%) | 0.004 | 2,260 (1%) | 2,369 (2%) | 0.006 |
| Hypothyroidism | 1,129 (7%) | 1,067 (7%) | 0.016 | 3,476 (6%) | 3,416 (6%) | 0.005 | 935 (4%) | 954 (4%) | 0.003 | 1,772 (1%) | 1,653 (1%) | 0.008 |
| Malignant neoplastic disease | 3,126 (20%) | 3,097 (20%) | 0.005 | 5,084 (8%) | 5,529 (9%) | 0.024 | 864 (4%) | 922 (4%) | 0.011 | 877 (1%) | 902 (1%) | 0.002 |
| Myocardial infarction | 764 (5%) | 681 (4%) | 0.026 | 1,797 (3%) | 1,853 (3%) | 0.005 | 230 (1%) | 186 (1%) | 0.020 | 189 (0%) | 128 (0%) | 0.013 |

| | Cohort 1 | | | Cohort 2 | | | Cohort 3 | | | Cohort 4 | | |
|---|---|---|---|---|---|---|---|---|---|---|---|---|
| | **Unvaccinated** | **Vaccinated** | **ASMD** | **Unvaccinated** | **Vaccinated** | **ASMD** | **Unvaccinated** | **Vaccinated** | **ASMD** | **Unvaccinated** | **Vaccinated** | **ASMD** |
| Osteoporosis | 1,312 (9%) | 1,225 (8%) | 0.021 | 1,775 (3%) | 1,805 (3%) | 0.002 | 251 (1%) | 282 (1%) | 0.012 | 154 (0%) | 135 (0%) | 0.004 |
| Pneumonia | 573 (4%) | 475 (3%) | 0.035 | 1,232 (2%) | 1,107 (2%) | 0.016 | 256 (1%) | 244 (1%) | 0.005 | 744 (0%) | 698 (0%) | 0.005 |
| Rheumatoid arthritis | 248 (2%) | 234 (2%) | 0.008 | 854 (1%) | 945 (2%) | 0.012 | 113 (0%) | 102 (0%) | 0.007 | 165 (0%) | 62 (0%) | 0.025 |
| Stroke | 739 (5%) | 650 (4%) | 0.028 | 1,291 (2%) | 1,297 (2%) | 0.000 | 172 (1%) | 129 (1%) | 0.023 | 204 (0%) | 138 (0%) | 0.013 |
| Venous thromboembolism | 640 (4%) | 622 (4%) | 0.006 | 1,355 (2%) | 1,347 (2%) | 0.002 | 273 (1%) | 256 (1%) | 0.008 | 604 (0%) | 288 (0%) | 0.039 |

The 4 cohorts represent vaccine rollout periods. *Calculated as the days of previous observation in the database before index date. **Assessed anytime before index date. ASMD = Absolute standardized mean difference, COPD = Chronic obstructive pulmonary disease, GERD = Gastro-Esophageal reflux disease

**Table S12: Characteristics of unweighted populations in CPRD GOLD,** database, stratified by staggered cohort and exposure status. Exposure is BNT162b2 vaccine.

| | Cohort 1 | | | Cohort 2 | | | Cohort 3 | | | Cohort 4 | | |
|---|---|---|---|---|---|---|---|---|---|---|---|---|
| | **Unvaccinated** | **Vaccinated** | **ASMD** | **Unvaccinated** | **Vaccinated** | **ASMD** | **Unvaccinated** | **Vaccinated** | **ASMD** | **Unvaccinated** | **Vaccinated** | **ASMD** |
| **N (individuals)** | 169,459 | 32,755 | | 584,309 | 180,670 | | 416,549 | 36,748 | | 465,326 | 365,096 | |
| **Age, median [Q25-Q75]** | 78 [76-82] | 81 [79-86] | 0.306 | 45 [33-58] | 70 [66-74] | 1.059 | 42 [31-55] | 54 [50-60] | 0.407 | 37 [28-49] | 31 [24-37] | 0.445 |
| **Sex: Female, N(%)** | 93,939 (55%) | 18,415 (56%) | 0.016 | 345,249 (59%) | 97,747 (54%) | 0.101 | 229,807 (55%) | 19,649 (53%) | 0.034 | 206,080 (44%) | 167,510 (46%) | 0.032 |
| **Years of prior history*, median [Q25-Q75]** | 17 [13-20] | 18 [13-21] | 0.017 | 17 [10-19] | 17 [12-19] | 0.076 | 17 [9-19] | 18 [12-20] | 0.153 | 13 [6-18] | 15 [6-18] | 0.018 |
| **Number visits, median [Q25-Q75]** | 11 [6-18] | 14 [9-21] | | 5 [1-11] | 10 [6-16] | | 3 [0-8] | 6 [2-11] | | 0 [0-4] | 2 [0-6] | |
| **Number pcrs, median [Q25-Q75]** | 0[0-0] | 0[0-0] | | 0[0-0] | 0[0-0] | | 0[0-0] | 0[0-0] | | 0[0-0] | 0[0-0] | |
| **Comorbidities**, N(%)** | | | | | | | | | | | | |
| Anxiety | 20,145 (12%) | 4,336 (13%) | 0.041 | 132,397 (23%) | 28,697 (16%) | 0.172 | 82,752 (20%) | 8,004 (22%) | 0.047 | 64,849 (14%) | 59,094 (16%) | 0.063 |
| Asthma | 15,611 (9%) | 3,250 (10%) | 0.024 | 119,679 (20%) | 21,396 (12%) | 0.236 | 68,556 (16%) | 4,053 (11%) | 0.158 | 30,579 (7%) | 33,240 (9%) | 0.094 |
| Chronic kidney disease | 29,378 (17%) | 7,430 (23%) | 0.134 | 15,504 (3%) | 15,033 (8%) | 0.251 | 6,636 (2%) | 566 (2%) | 0.004 | 4,692 (1%) | 578 (0%) | 0.112 |
| COPD | 14,265 (8%) | 2,417 (7%) | 0.039 | 12,276 (2%) | 11,545 (6%) | 0.214 | 5,085 (1%) | 348 (1%) | 0.026 | 3,657 (1%) | 285 (0%) | 0.108 |
| Dementia | 6,705 (4%) | 1,895 (6%) | 0.085 | 2,008 (0%) | 1,375 (1%) | 0.056 | 1,168 (0%) | 87 (0%) | 0.009 | 746 (0%) | 29 (0%) | 0.053 |
| Depressive disorder | 16,385 (10%) | 3,310 (10%) | 0.015 | 111,708 (19%) | 26,294 (15%) | 0.122 | 66,510 (16%) | 6,569 (18%) | 0.051 | 49,969 (11%) | 39,181 (11%) | 0.000 |
| Diabetes | 23,614 (14%) | 4,882 (15%) | 0.028 | 29,200 (5%) | 22,552 (12%) | 0.267 | 12,338 (3%) | 1,191 (3%) | 0.016 | 9,143 (2%) | 2,507 (1%) | 0.112 |
| GERD | 7,655 (5%) | 2,012 (6%) | 0.072 | 18,069 (3%) | 8,525 (5%) | 0.084 | 8,839 (2%) | 1,093 (3%) | 0.054 | 5,864 (1%) | 5,213 (1%) | 0.015 |
| Heart failure | 7,502 (4%) | 1,782 (5%) | 0.047 | 4,026 (1%) | 3,669 (2%) | 0.116 | 1,715 (0%) | 100 (0%) | 0.024 | 1,236 (0%) | 98 (0%) | 0.063 |
| Hypertension | 56,411 (33%) | 11,995 (37%) | 0.070 | 58,749 (10%) | 49,763 (28%) | 0.459 | 24,301 (6%) | 4,679 (13%) | 0.239 | 16,363 (4%) | 4,776 (1%) | 0.144 |
| Hypothyroidism | 12,163 (7%) | 2,373 (7%) | 0.003 | 22,061 (4%) | 11,106 (6%) | 0.109 | 10,565 (3%) | 1,553 (4%) | 0.094 | 6,617 (1%) | 4,180 (1%) | 0.025 |
| Malignant neoplastic disease | 30,728 (18%) | 6,972 (21%) | 0.079 | 19,832 (3%) | 22,617 (13%) | 0.342 | 8,827 (2%) | 1,522 (4%) | 0.116 | 5,804 (1%) | 1,796 (0%) | 0.081 |

| | Cohort 1 | | | Cohort 2 | | | Cohort 3 | | | Cohort 4 | | |
|---|---|---|---|---|---|---|---|---|---|---|---|---|
| | **Unvaccinated** | **Vaccinated** | **ASMD** | **Unvaccinated** | **Vaccinated** | **ASMD** | **Unvaccinated** | **Vaccinated** | **ASMD** | **Unvaccinated** | **Vaccinated** | **ASMD** |
| Myocardial infarction | 7,716 (5%) | 1,603 (5%) | 0.016 | 8,437 (1%) | 6,621 (4%) | 0.141 | 2,644 (1%) | 271 (1%) | 0.012 | 1,985 (0%) | 154 (0%) | 0.080 |
| Osteoporosis | 12,776 (8%) | 2,770 (8%) | 0.034 | 5,867 (1%) | 7,531 (4%) | 0.200 | 2,558 (1%) | 458 (1%) | 0.066 | 1,841 (0%) | 205 (0%) | 0.072 |
| Pneumonia | 5,415 (3%) | 1,032 (3%) | 0.003 | 7,266 (1%) | 3,418 (2%) | 0.052 | 3,790 (1%) | 345 (1%) | 0.003 | 2,512 (1%) | 1,696 (0%) | 0.011 |
| Rheumatoid arthritis | 2,566 (2%) | 512 (2%) | 0.004 | 4,018 (1%) | 2,929 (2%) | 0.087 | 1,276 (0%) | 154 (0%) | 0.019 | 881 (0%) | 111 (0%) | 0.048 |
| Stroke | 7,203 (4%) | 1,490 (5%) | 0.015 | 5,907 (1%) | 4,632 (3%) | 0.117 | 2,152 (1%) | 199 (1%) | 0.003 | 1,578 (0%) | 225 (0%) | 0.062 |
| Venous thromboembolism | 6,198 (4%) | 1,389 (4%) | 0.030 | 7,333 (1%) | 4,490 (2%) | 0.091 | 3,056 (1%) | 370 (1%) | 0.029 | 2,585 (1%) | 496 (0%) | 0.072 |

The 4 cohorts represent vaccine rollout periods. *Calculated as the days of previous observation in the database before index date. **Assessed anytime before index date. ASMD = Absolute standardized mean difference, COPD = Chronic obstructive pulmonary disease, GERD = Gastro-Esophageal reflux disease

**Table S13: Characteristics of weighted populations in SIDIAP,** database, stratified by staggered cohort and exposure status. Exposure is any COVID-19 vaccine.

| | Cohort 1 | | | Cohort 2 | | | Cohort 3 | | | Cohort 4 | | |
|---|---|---|---|---|---|---|---|---|---|---|---|---|
| | **Unvaccinated** | **Vaccinated** | **ASMD** | **Unvaccinated** | **Vaccinated** | **ASMD** | **Unvaccinated** | **Vaccinated** | **ASMD** | **Unvaccinated** | **Vaccinated** | **ASMD** |
| **N (individuals)** | 58,342 | 58,535 | | 235,543 | 235,780 | | 344,520 | 342,678 | | 413,428 | 412,893 | |
| **Age, median [Q25-Q75]** | 86 [83-89] | 86 [82-89] | 0.002 | 68 [63-73] | 68 [63-74] | 0.003 | 52 [46-58] | 52 [46-59] | 0.000 | 36 [28-44] | 36 [28-44] | 0.002 |
| **Sex: Female, N(%)** | 36,115 (62%) | 36,214 (62%) | 0.001 | 128,806 (55%) | 129,053 (55%) | 0.001 | 164,316 (48%) | 163,188 (48%) | 0.001 | 188,426 (46%) | 188,471 (46%) | 0.001 |
| **Years of prior history*, median [Q25-Q75]** | 15 [15-15] | 15 [15-15] | 0.001 | 15 [15-15] | 15 [15-15] | 0.000 | 15 [15-15] | 15 [15-15] | 0.003 | 15 [13-16] | 15 [13-16] | 0.004 |
| **Number visits, median [Q25-Q75]** | 8 [3-15] | 8 [4-15] | | 4 [0-9] | 4 [1-9] | | 0 [0-6] | 2 [0-6] | | 0 [0-4] | 0 [0-5] | |
| **Number pcrs, median [Q25-Q75]** | 0[0-0] | 0[0-0] | | 0[0-0] | 0[0-0] | | 0[0-0] | 0[0-0] | | 0[0-0] | 0[0-0] | |
| **Comorbidities**, N(%)** | | | | | | | | | | | | |
| Anxiety | 11,384 (20%) | 11,467 (20%) | 0.002 | 52,314 (22%) | 52,262 (22%) | 0.001 | 81,514 (24%) | 80,775 (24%) | 0.002 | 84,856 (21%) | 83,145 (20%) | 0.010 |
| Asthma | 3,708 (6%) | 3,627 (6%) | 0.007 | 10,973 (5%) | 10,703 (5%) | 0.006 | 14,915 (4%) | 14,492 (4%) | 0.005 | 19,969 (5%) | 19,628 (5%) | 0.004 |
| Chronic kidney disease | 18,755 (32%) | 18,803 (32%) | 0.001 | 21,147 (9%) | 21,249 (9%) | 0.001 | 7,411 (2%) | 7,335 (2%) | 0.001 | 3,293 (1%) | 2,846 (1%) | 0.012 |
| COPD | 7,008 (12%) | 6,983 (12%) | 0.002 | 17,160 (7%) | 16,784 (7%) | 0.006 | 8,852 (3%) | 8,622 (3%) | 0.003 | 3,210 (1%) | 3,215 (1%) | 0.000 |
| Dementia | 5,686 (10%) | 5,812 (10%) | 0.006 | 4,628 (2%) | 4,425 (2%) | 0.006 | 1,247 (0%) | 1,229 (0%) | 0.001 | 673 (0%) | 689 (0%) | 0.001 |
| Depressive disorder | 10,960 (19%) | 11,020 (19%) | 0.001 | 34,232 (15%) | 34,571 (15%) | 0.004 | 34,243 (10%) | 32,427 (9%) | 0.016 | 23,679 (6%) | 23,024 (6%) | 0.007 |
| Diabetes | 14,529 (25%) | 14,269 (24%) | 0.012 | 39,335 (17%) | 39,133 (17%) | 0.003 | 25,964 (8%) | 26,135 (8%) | 0.003 | 14,136 (3%) | 14,091 (3%) | 0.000 |
| GERD | 6,898 (12%) | 6,871 (12%) | 0.003 | 21,088 (9%) | 21,212 (9%) | 0.002 | 16,959 (5%) | 17,024 (5%) | 0.002 | 10,569 (3%) | 10,903 (3%) | 0.005 |
| Heart failure | 9,320 (16%) | 9,141 (16%) | 0.010 | 10,251 (4%) | 10,196 (4%) | 0.001 | 4,001 (1%) | 3,743 (1%) | 0.007 | 1,606 (0%) | 1,565 (0%) | 0.002 |
| Hypertension | 38,140 (65%) | 37,917 (65%) | 0.013 | 96,384 (41%) | 96,118 (41%) | 0.003 | 58,596 (17%) | 57,241 (17%) | 0.008 | 22,160 (5%) | 22,739 (6%) | 0.006 |
| Hypothyroidism | 7,350 (13%) | 7,236 (12%) | 0.007 | 24,054 (10%) | 24,260 (10%) | 0.003 | 21,956 (6%) | 21,573 (6%) | 0.003 | 17,452 (4%) | 17,758 (4%) | 0.004 |
| Malignant neoplastic disease | 15,402 (26%) | 15,230 (26%) | 0.009 | 37,052 (16%) | 38,490 (16%) | 0.016 | 19,765 (6%) | 19,599 (6%) | 0.001 | 9,413 (2%) | 8,865 (2%) | 0.009 |

| | Cohort 1 | | | Cohort 2 | | | Cohort 3 | | | Cohort 4 | | |
|---|---|---|---|---|---|---|---|---|---|---|---|---|
| | **Unvaccinated** | **Vaccinated** | **ASMD** | **Unvaccinated** | **Vaccinated** | **ASMD** | **Unvaccinated** | **Vaccinated** | **ASMD** | **Unvaccinated** | **Vaccinated** | **ASMD** |
| Myocardial infarction | 2,258 (4%) | 2,235 (4%) | 0.003 | 5,972 (3%) | 5,812 (2%) | 0.005 | 3,615 (1%) | 3,448 (1%) | 0.004 | 1,362 (0%) | 1,364 (0%) | 0.000 |
| Osteoporosis | 9,270 (16%) | 9,151 (16%) | 0.007 | 18,870 (8%) | 19,036 (8%) | 0.002 | 7,126 (2%) | 7,039 (2%) | 0.001 | 2,254 (1%) | 2,040 (0%) | 0.007 |
| Pneumonia | 6,643 (11%) | 6,563 (11%) | 0.005 | 15,630 (7%) | 15,557 (7%) | 0.002 | 14,821 (4%) | 14,580 (4%) | 0.002 | 14,171 (3%) | 14,083 (3%) | 0.001 |
| Rheumatoid arthritis | 765 (1%) | 727 (1%) | 0.006 | 2,274 (1%) | 2,286 (1%) | 0.000 | 1,544 (0%) | 1,592 (0%) | 0.002 | 754 (0%) | 646 (0%) | 0.006 |
| Stroke | 5,119 (9%) | 5,043 (9%) | 0.006 | 8,750 (4%) | 7,706 (3%) | 0.024 | 4,343 (1%) | 4,147 (1%) | 0.005 | 1,828 (0%) | 1,840 (0%) | 0.001 |
| Venous thromboembolism | 2,469 (4%) | 2,418 (4%) | 0.005 | 4,745 (2%) | 4,419 (2%) | 0.010 | 3,365 (1%) | 2,982 (1%) | 0.011 | 1,931 (0%) | 1,777 (0%) | 0.005 |

The 4 cohorts represent vaccine rollout periods. *Calculated as the days of previous observation in the database before index date. **Assessed anytime before index date. ASMD = Absolute standardized mean difference, COPD = Chronic obstructive pulmonary disease, GERD = Gastro-Esophageal reflux disease

**Table S14: Characteristics of unweighted populations in SIDIAP,** database, stratified by staggered cohort and exposure status. Exposure is any COVID-19 vaccine.

| | Cohort 1 | | | Cohort 2 | | | Cohort 3 | | | Cohort 4 | | |
|---|---|---|---|---|---|---|---|---|---|---|---|---|
| | **Unvaccinated** | **Vaccinated** | **ASMD** | **Unvaccinated** | **Vaccinated** | **ASMD** | **Unvaccinated** | **Vaccinated** | **ASMD** | **Unvaccinated** | **Vaccinated** | **ASMD** |
| **N (individuals)** | 223,962 | 89,941 | | 433,151 | 819,590 | | 869,497 | 954,232 | | 1,061,634 | 880,950 | |
| **Age, median [Q25-Q75]** | 85 [82-88] | 87 [83-90] | 0.253 | 67 [62-72] | 72 [65-78] | 0.391 | 48 [43-57] | 54 [49-58] | 0.192 | 38 [28-51] | 37 [29-43] | 0.246 |
| **Sex: Female, N(%)** | 136,888 (61%) | 56,235 (63%) | 0.029 | 237,494 (55%) | 443,019 (54%) | 0.016 | 409,207 (47%) | 462,599 (48%) | 0.028 | 488,499 (46%) | 408,555 (46%) | 0.007 |
| **Years of prior history*, median [Q25-Q75]** | 15 [15-15] | 15 [15-15] | 0.051 | 15 [15-15] | 15 [15-15] | 0.185 | 15 [15-15] | 15 [15-15] | 0.246 | 15 [10-16] | 15 [15-16] | 0.195 |
| **Number visits, median [Q25-Q75]** | 7 [2-13] | 10 [5-18] | | 3 [0-8] | 6 [2-11] | | 1 [0-5] | 2 [0-7] | | 0 [0-4] | 2 [0-6] | |
| **Number pcrs, median [Q25-Q75]** | 0[0-0] | 0[0-1] | | 0[0-0] | 0[0-1] | | 0[0-0] | 0[0-1] | | 0[0-0] | 0[0-1] | |
| **Comorbidities**, N(%)** | | | | | | | | | | | | |
| Anxiety | 41,908 (19%) | 18,117 (20%) | 0.036 | 97,857 (23%) | 181,286 (22%) | 0.011 | 197,922 (23%) | 233,123 (24%) | 0.039 | 203,019 (19%) | 186,562 (21%) | 0.051 |
| Asthma | 13,371 (6%) | 5,841 (6%) | 0.022 | 19,809 (5%) | 41,966 (5%) | 0.025 | 36,811 (4%) | 42,221 (4%) | 0.009 | 45,374 (4%) | 46,776 (5%) | 0.049 |
| Chronic kidney disease | 63,041 (28%) | 31,132 (35%) | 0.140 | 32,450 (7%) | 101,792 (12%) | 0.165 | 22,454 (3%) | 20,303 (2%) | 0.030 | 20,368 (2%) | 5,314 (1%) | 0.118 |
| COPD | 24,459 (11%) | 11,292 (13%) | 0.051 | 28,780 (7%) | 67,795 (8%) | 0.062 | 19,020 (2%) | 27,451 (3%) | 0.044 | 15,415 (1%) | 5,836 (1%) | 0.077 |
| Dementia | 16,743 (7%) | 10,455 (12%) | 0.142 | 7,404 (2%) | 19,153 (2%) | 0.045 | 6,009 (1%) | 2,787 (0%) | 0.057 | 5,866 (1%) | 1,310 (0%) | 0.068 |
| Depressive disorder | 38,148 (17%) | 18,040 (20%) | 0.078 | 61,308 (14%) | 124,929 (15%) | 0.031 | 77,039 (9%) | 98,723 (10%) | 0.050 | 66,864 (6%) | 49,608 (6%) | 0.028 |
| Diabetes | 51,964 (23%) | 22,968 (26%) | 0.054 | 65,572 (15%) | 155,362 (19%) | 0.102 | 60,254 (7%) | 78,356 (8%) | 0.048 | 49,331 (5%) | 29,768 (3%) | 0.065 |
| GERD | 25,376 (11%) | 10,728 (12%) | 0.019 | 34,715 (8%) | 87,626 (11%) | 0.092 | 37,828 (4%) | 53,565 (6%) | 0.058 | 31,600 (3%) | 23,657 (3%) | 0.018 |
| Heart failure | 28,538 (13%) | 16,202 (18%) | 0.147 | 16,697 (4%) | 42,924 (5%) | 0.066 | 11,909 (1%) | 10,469 (1%) | 0.025 | 10,738 (1%) | 2,776 (0%) | 0.086 |
| Hypertension | 138,631 (62%) | 60,128 (67%) | 0.104 | 157,290 (36%) | 390,610 (48%) | 0.231 | 125,239 (14%) | 188,390 (20%) | 0.142 | 97,835 (9%) | 43,351 (5%) | 0.168 |

| | Cohort 1 | | | Cohort 2 | | | Cohort 3 | | | Cohort 4 | | |
|---|---|---|---|---|---|---|---|---|---|---|---|---|
| | **Unvaccinated** | **Vaccinated** | **ASMD** | **Unvaccinated** | **Vaccinated** | **ASMD** | **Unvaccinated** | **Vaccinated** | **ASMD** | **Unvaccinated** | **Vaccinated** | **ASMD** |
| Hypothyroidism | 26,875 (12%) | 11,473 (13%) | 0.023 | 42,245 (10%) | 89,496 (11%) | 0.038 | 50,396 (6%) | 64,873 (7%) | 0.041 | 49,279 (5%) | 38,477 (4%) | 0.013 |
| Malignant neoplastic disease | 54,714 (24%) | 24,173 (27%) | 0.056 | 81,394 (19%) | 151,745 (19%) | 0.007 | 48,971 (6%) | 59,566 (6%) | 0.026 | 37,755 (4%) | 18,021 (2%) | 0.092 |
| Myocardial infarction | 7,886 (4%) | 3,642 (4%) | 0.028 | 9,894 (2%) | 22,773 (3%) | 0.031 | 7,570 (1%) | 11,483 (1%) | 0.033 | 6,171 (1%) | 2,459 (0%) | 0.046 |
| Osteoporosis | 32,239 (14%) | 14,846 (17%) | 0.058 | 29,512 (7%) | 81,707 (10%) | 0.114 | 16,393 (2%) | 21,679 (2%) | 0.027 | 14,373 (1%) | 3,343 (0%) | 0.105 |
| Pneumonia | 22,254 (10%) | 10,959 (12%) | 0.072 | 27,837 (6%) | 61,325 (7%) | 0.042 | 34,513 (4%) | 44,236 (5%) | 0.033 | 36,413 (3%) | 32,657 (4%) | 0.015 |
| Rheumatoid arthritis | 2,666 (1%) | 1,204 (1%) | 0.013 | 3,771 (1%) | 8,890 (1%) | 0.022 | 3,239 (0%) | 4,770 (0%) | 0.019 | 2,706 (0%) | 1,281 (0%) | 0.024 |
| Stroke | 16,690 (7%) | 8,542 (9%) | 0.073 | 14,256 (3%) | 32,612 (4%) | 0.037 | 10,877 (1%) | 12,068 (1%) | 0.001 | 9,400 (1%) | 3,384 (0%) | 0.063 |
| Venous thromboembolism | 8,014 (4%) | 4,126 (5%) | 0.051 | 8,172 (2%) | 17,846 (2%) | 0.021 | 7,747 (1%) | 8,920 (1%) | 0.005 | 6,632 (1%) | 3,768 (0%) | 0.027 |

The 4 cohorts represent vaccine rollout periods. *Calculated as the days of previous observation in the database before index date. **Assessed anytime before index date. ASMD = Absolute standardized mean difference, COPD = Chronic obstructive pulmonary disease, GERD = Gastro-Esophageal reflux disease

**Table S15: Characteristics of weighted populations in SIDIAP,** database, stratified by staggered cohort and exposure status. Exposure is BNT162b2 vaccine.

| | Cohort 1 | | | Cohort 2 | | | Cohort 3 | | | Cohort 4 | | |
|---|---|---|---|---|---|---|---|---|---|---|---|---|
| | Unvaccinated | Vaccinated | ASMD | Unvaccinated | Vaccinated | ASMD | Unvaccinated | Vaccinated | ASMD | Unvaccinated | Vaccinated | ASMD |
| **N (individuals)** | 57,981 | 57,843 | | 99,838 | 99,488 | | 276,241 | 273,593 | | 312,492 | 313,346 | |
| **Age, median [Q25-Q75]** | 86 [83-89] | 86 [82-89] | 0.000 | 74 [71-81] | 74 [72-81] | 0.006 | 50 [45-56] | 50 [45-56] | 0.003 | 36 [29-43] | 36 [29-43] | 0.004 |
| **Sex: Female, N(%)** | 35,828 (62%) | 35,895 (62%) | 0.005 | 58,343 (58%) | 57,844 (58%) | 0.006 | 130,375 (47%) | 129,701 (47%) | 0.004 | 145,270 (46%) | 144,384 (46%) | 0.008 |
| **Years of prior history*, median [Q25-Q75]** | 15 [15-15] | 15 [15-15] | 0.002 | 15 [15-15] | 15 [15-15] | 0.005 | 15 [15-15] | 15 [15-15] | 0.001 | 15 [13-16] | 15 [14-16] | 0.004 |
| **Number visits, median [Q25-Q75]** | 8 [3-15] | 8 [4-15] | | 4 [0-10] | 6 [2-10] | | 0 [0-5] | 2 [0-6] | | 0 [0-4] | 0 [0-5] | |
| **Number pcrs, median [Q25-Q75]** | 0[0-0] | 0[0-0] | | 0[0-0] | 0[0-0] | | 0[0-0] | 0[0-0] | | 0[0-0] | 0[0-0] | |
| **Comorbidities**, N(%)** | | | | | | | | | | | | |
| Anxiety | 11,327 (20%) | 11,359 (20%) | 0.003 | 20,281 (20%) | 20,181 (20%) | 0.001 | 67,000 (24%) | 63,263 (23%) | 0.027 | 65,060 (21%) | 64,616 (21%) | 0.005 |
| Asthma | 3,734 (6%) | 3,583 (6%) | 0.010 | 5,213 (5%) | 5,179 (5%) | 0.001 | 12,220 (4%) | 11,760 (4%) | 0.006 | 15,491 (5%) | 15,211 (5%) | 0.005 |
| Chronic kidney disease | 18,610 (32%) | 18,576 (32%) | 0.000 | 15,548 (16%) | 15,607 (16%) | 0.003 | 5,583 (2%) | 5,399 (2%) | 0.003 | 1,999 (1%) | 2,008 (1%) | 0.000 |
| COPD | 6,980 (12%) | 6,907 (12%) | 0.003 | 9,163 (9%) | 9,045 (9%) | 0.003 | 6,020 (2%) | 6,047 (2%) | 0.002 | 1,998 (1%) | 2,000 (1%) | 0.000 |
| Dementia | 5,678 (10%) | 5,771 (10%) | 0.006 | 4,152 (4%) | 3,716 (4%) | 0.022 | 1,037 (0%) | 980 (0%) | 0.003 | 502 (0%) | 506 (0%) | 0.000 |
| Depressive disorder | 10,920 (19%) | 10,923 (19%) | 0.001 | 15,653 (16%) | 15,532 (16%) | 0.002 | 26,419 (10%) | 24,357 (9%) | 0.023 | 17,262 (6%) | 17,507 (6%) | 0.003 |
| Diabetes | 14,416 (25%) | 14,096 (24%) | 0.011 | 20,461 (20%) | 20,645 (21%) | 0.006 | 19,302 (7%) | 18,543 (7%) | 0.008 | 10,119 (3%) | 10,440 (3%) | 0.005 |
| GERD | 6,784 (12%) | 6,826 (12%) | 0.003 | 10,187 (10%) | 10,088 (10%) | 0.002 | 12,811 (5%) | 12,805 (5%) | 0.002 | 7,750 (2%) | 8,224 (3%) | 0.009 |
| Heart failure | 9,390 (16%) | 8,983 (16%) | 0.018 | 7,563 (8%) | 7,296 (7%) | 0.009 | 3,044 (1%) | 2,699 (1%) | 0.011 | 1,069 (0%) | 1,049 (0%) | 0.001 |
| Hypertension | 37,879 (65%) | 37,465 (65%) | 0.012 | 51,262 (51%) | 50,608 (51%) | 0.010 | 42,266 (15%) | 41,291 (15%) | 0.006 | 15,063 (5%) | 15,323 (5%) | 0.003 |
| Hypothyroidism | 7,310 (13%) | 7,166 (12%) | 0.007 | 11,611 (12%) | 11,249 (11%) | 0.010 | 16,597 (6%) | 16,509 (6%) | 0.001 | 13,155 (4%) | 13,617 (4%) | 0.007 |
| Malignant neoplastic disease | 15,302 (26%) | 14,971 (26%) | 0.012 | 19,393 (19%) | 18,719 (19%) | 0.015 | 14,311 (5%) | 13,981 (5%) | 0.003 | 6,555 (2%) | 6,398 (2%) | 0.004 |
| Myocardial infarction | 2,286 (4%) | 2,182 (4%) | 0.009 | 3,130 (3%) | 2,924 (3%) | 0.011 | 2,569 (1%) | 2,451 (1%) | 0.004 | 875 (0%) | 880 (0%) | 0.000 |

| | Cohort 1 | | | Cohort 2 | | | Cohort 3 | | | Cohort 4 | | |
|---|---|---|---|---|---|---|---|---|---|---|---|---|
| | **Unvaccinated** | **Vaccinated** | **ASMD** | **Unvaccinated** | **Vaccinated** | **ASMD** | **Unvaccinated** | **Vaccinated** | **ASMD** | **Unvaccinated** | **Vaccinated** | **ASMD** |
| Osteoporosis | 9,170 (16%) | 9,085 (16%) | 0.003 | 11,182 (11%) | 11,201 (11%) | 0.002 | 4,764 (2%) | 4,639 (2%) | 0.002 | 1,344 (0%) | 1,306 (0%) | 0.002 |
| Pneumonia | 6,588 (11%) | 6,476 (11%) | 0.005 | 7,945 (8%) | 7,918 (8%) | 0.000 | 11,759 (4%) | 11,273 (4%) | 0.007 | 10,730 (3%) | 10,798 (3%) | 0.001 |
| Rheumatoid arthritis | 755 (1%) | 718 (1%) | 0.006 | 1,064 (1%) | 1,111 (1%) | 0.005 | 1,113 (0%) | 1,026 (0%) | 0.004 | 536 (0%) | 461 (0%) | 0.006 |
| Stroke | 5,070 (9%) | 5,006 (9%) | 0.003 | 5,363 (5%) | 5,086 (5%) | 0.012 | 3,121 (1%) | 3,043 (1%) | 0.002 | 1,199 (0%) | 1,253 (0%) | 0.003 |
| Venous thromboembolism | 2,452 (4%) | 2,378 (4%) | 0.006 | 2,660 (3%) | 2,526 (3%) | 0.008 | 2,527 (1%) | 2,211 (1%) | 0.012 | 1,394 (0%) | 1,327 (0%) | 0.003 |

The 4 cohorts represent vaccine rollout periods. *Calculated as the days of previous observation in the database before index date. **Assessed anytime before index date. ASMD = Absolute standardized mean difference, COPD = Chronic obstructive pulmonary disease, GERD = Gastro-Esophageal reflux disease

**Table S16: Characteristics of unweighted populations in SIDIAP,** database, stratified by staggered cohort and exposure status. Exposure is BNT162b2 vaccine.

| | Cohort 1 | | | Cohort 2 | | | Cohort 3 | | | Cohort 4 | | |
|---|---|---|---|---|---|---|---|---|---|---|---|---|
| | **Unvaccinated** | **Vaccinated** | **ASMD** | **Unvaccinated** | **Vaccinated** | **ASMD** | **Unvaccinated** | **Vaccinated** | **ASMD** | **Unvaccinated** | **Vaccinated** | **ASMD** |
| **N (individuals)** | 223,960 | 88,896 | | 433,111 | 445,581 | | 869,109 | 706,435 | | 1,068,043 | 580,329 | |
| **Age, median [Q25-Q75]** | 85 [82-88] | 87 [84-90] | 0.258 | 67 [62-72] | 78 [74-83] | 0.943 | 48 [43-57] | 53 [48-57] | 0.127 | 38 [28-51] | 38 [31-43] | 0.231 |
| **Sex: Female, N(%)** | 136,891 (61%) | 55,704 (63%) | 0.032 | 237,440 (55%) | 254,378 (57%) | 0.046 | 409,076 (47%) | 341,289 (48%) | 0.025 | 491,528 (46%) | 274,150 (47%) | 0.024 |
| **Years of prior history*, median [Q25-Q75]** | 15 [15-15] | 15 [15-15] | 0.051 | 15 [15-15] | 15 [15-15] | 0.206 | 15 [15-15] | 15 [15-15] | 0.244 | 15 [10-16] | 15 [15-16] | 0.211 |
| **Number visits, median [Q25-Q75]** | 7 [2-13] | 10 [5-18] | | 3 [0-8] | 7 [3-12] | | 1 [0-5] | 2 [0-7] | | 0 [0-4] | 2 [0-6] | |
| **Number pcrs, median [Q25-Q75]** | 0[0-0] | 0[0-1] | | 0[0-0] | 0[0-1] | | 0[0-0] | 0[0-1] | | 0[0-0] | 0[0-1] | |
| **Comorbidities**, N(%)** | | | | | | | | | | | | |
| Anxiety | 41,914 (19%) | 17,962 (20%) | 0.038 | 97,901 (23%) | 91,620 (21%) | 0.050 | 197,933 (23%) | 172,434 (24%) | 0.039 | 204,108 (19%) | 125,466 (22%) | 0.062 |
| Asthma | 13,370 (6%) | 5,796 (7%) | 0.023 | 19,824 (5%) | 25,291 (6%) | 0.050 | 36,788 (4%) | 31,586 (4%) | 0.012 | 45,747 (4%) | 30,865 (5%) | 0.048 |
| Chronic kidney disease | 63,046 (28%) | 30,745 (35%) | 0.139 | 32,447 (7%) | 83,968 (19%) | 0.341 | 22,494 (3%) | 13,920 (2%) | 0.041 | 20,416 (2%) | 3,171 (1%) | 0.124 |
| COPD | 24,471 (11%) | 11,169 (13%) | 0.051 | 28,773 (7%) | 44,207 (10%) | 0.119 | 19,028 (2%) | 17,815 (3%) | 0.022 | 15,424 (1%) | 3,218 (1%) | 0.090 |
| Dementia | 16,748 (7%) | 10,393 (12%) | 0.143 | 7,389 (2%) | 17,606 (4%) | 0.136 | 6,003 (1%) | 2,033 (0%) | 0.058 | 5,878 (1%) | 856 (0%) | 0.068 |
| Depressive disorder | 38,161 (17%) | 17,894 (20%) | 0.079 | 61,309 (14%) | 72,447 (16%) | 0.059 | 77,050 (9%) | 69,517 (10%) | 0.033 | 67,070 (6%) | 32,476 (6%) | 0.029 |
| Diabetes | 51,948 (23%) | 22,682 (26%) | 0.054 | 65,583 (15%) | 99,692 (22%) | 0.186 | 60,242 (7%) | 53,330 (8%) | 0.024 | 49,433 (5%) | 19,502 (3%) | 0.065 |
| GERD | 25,370 (11%) | 10,611 (12%) | 0.019 | 34,704 (8%) | 53,110 (12%) | 0.131 | 37,842 (4%) | 37,707 (5%) | 0.046 | 31,662 (3%) | 15,544 (3%) | 0.017 |
| Heart failure | 28,525 (13%) | 16,041 (18%) | 0.147 | 16,686 (4%) | 34,932 (8%) | 0.171 | 11,920 (1%) | 7,057 (1%) | 0.034 | 10,734 (1%) | 1,642 (0%) | 0.090 |
| Hypertension | 138,655 (62%) | 59,414 (67%) | 0.103 | 157,291 (36%) | 252,365 (57%) | 0.416 | 125,235 (14%) | 128,859 (18%) | 0.104 | 97,944 (9%) | 26,383 (5%) | 0.184 |

| | Cohort 1 | | | Cohort 2 | | | Cohort 3 | | | Cohort 4 | | |
|---|---|---|---|---|---|---|---|---|---|---|---|---|
| | **Unvaccinated** | **Vaccinated** | **ASMD** | **Unvaccinated** | **Vaccinated** | **ASMD** | **Unvaccinated** | **Vaccinated** | **ASMD** | **Unvaccinated** | **Vaccinated** | **ASMD** |
| Hypothyroidism | 26,868 (12%) | 11,363 (13%) | 0.024 | 42,216 (10%) | 53,488 (12%) | 0.073 | 50,384 (6%) | 46,214 (7%) | 0.031 | 49,424 (5%) | 25,961 (4%) | 0.007 |
| Malignant neoplastic disease | 54,729 (24%) | 23,784 (27%) | 0.053 | 81,399 (19%) | 96,204 (22%) | 0.070 | 48,967 (6%) | 40,510 (6%) | 0.004 | 37,874 (4%) | 11,535 (2%) | 0.095 |
| Myocardial infarction | 7,890 (4%) | 3,590 (4%) | 0.027 | 9,892 (2%) | 14,140 (3%) | 0.055 | 7,573 (1%) | 7,753 (1%) | 0.023 | 6,182 (1%) | 1,405 (0%) | 0.053 |
| Osteoporosis | 32,250 (14%) | 14,701 (17%) | 0.059 | 29,528 (7%) | 56,885 (13%) | 0.201 | 16,385 (2%) | 14,126 (2%) | 0.008 | 14,380 (1%) | 1,876 (0%) | 0.113 |
| Pneumonia | 22,249 (10%) | 10,811 (12%) | 0.071 | 27,831 (6%) | 38,127 (9%) | 0.081 | 34,511 (4%) | 31,972 (5%) | 0.028 | 36,645 (3%) | 21,355 (4%) | 0.013 |
| Rheumatoid arthritis | 2,666 (1%) | 1,189 (1%) | 0.013 | 3,769 (1%) | 5,143 (1%) | 0.028 | 3,242 (0%) | 2,843 (0%) | 0.005 | 2,700 (0%) | 812 (0%) | 0.026 |
| Stroke | 16,693 (7%) | 8,475 (10%) | 0.075 | 14,244 (3%) | 24,307 (5%) | 0.106 | 10,868 (1%) | 8,114 (1%) | 0.009 | 9,394 (1%) | 2,033 (0%) | 0.068 |
| Venous thromboembolism | 8,023 (4%) | 4,072 (5%) | 0.050 | 8,162 (2%) | 12,564 (3%) | 0.062 | 7,738 (1%) | 6,066 (1%) | 0.003 | 6,658 (1%) | 2,434 (0%) | 0.028 |

The 4 cohorts represent vaccine rollout periods. *Calculated as the days of previous observation in the database before index date. **Assessed anytime before index date. ASMD = Absolute standardized mean difference, COPD = Chronic obstructive pulmonary disease, GERD = Gastro-Esophageal reflux disease

**Table S17: Characteristics of weighted populations in SIDIAP,** database, stratified by staggered cohort and exposure status. Exposure is ChAdOx1 vaccine.

| | Cohort 2 | | | Cohort 3 | | |
|---|---|---|---|---|---|---|
| | Unvaccinated | Vaccinated | ASMD | Unvaccinated | Vaccinated | ASMD |
| **N (individuals)** | 120,307 | 120,802 | | 44,033 | 43,901 | |
| **Age, median [Q25-Q75]** | 64 [61-66] | 64 [61-66] | 0.000 | 64 [61-66] | 64 [61-67] | 0.002 |
| **Sex: Female, N(%)** | 62,434 (52%) | 62,425 (52%) | 0.004 | 23,009 (52%) | 22,980 (52%) | 0.002 |
| **Years of prior history*, median [Q25-Q75]** | 15 [15-15] | 15 [15-15] | 0.003 | 15 [15-15] | 15 [15-15] | 0.005 |
| **Number visits, median [Q25-Q75]** | 2 [0-7] | 4 [1-8] | | 2 [0-7] | 2 [0-7] | |
| **Number pcrs, median [Q25-Q75]** | 0[0-0] | 0[0-0] | | 0[0-0] | 0[0-0] | |
| **Comorbidities**, N(%)** | | | | | | |
| Anxiety | 28,213 (23%) | 27,989 (23%) | 0.007 | 10,333 (23%) | 9,717 (22%) | 0.032 |
| Asthma | 4,909 (4%) | 4,785 (4%) | 0.006 | 1,664 (4%) | 1,639 (4%) | 0.002 |
| Chronic kidney disease | 4,390 (4%) | 4,526 (4%) | 0.005 | 1,556 (4%) | 1,597 (4%) | 0.006 |
| COPD | 6,702 (6%) | 6,846 (6%) | 0.004 | 2,405 (5%) | 2,350 (5%) | 0.005 |
| Dementia | 470 (0%) | 397 (0%) | 0.010 | 170 (0%) | 171 (0%) | 0.000 |
| Depressive disorder | 16,483 (14%) | 16,258 (13%) | 0.007 | 5,940 (13%) | 5,813 (13%) | 0.007 |
| Diabetes | 16,127 (13%) | 16,418 (14%) | 0.005 | 5,698 (13%) | 5,705 (13%) | 0.002 |
| GERD | 9,580 (8%) | 9,569 (8%) | 0.002 | 3,206 (7%) | 3,216 (7%) | 0.002 |
| Heart failure | 2,268 (2%) | 2,236 (2%) | 0.003 | 844 (2%) | 847 (2%) | 0.001 |
| Hypertension | 39,422 (33%) | 39,551 (33%) | 0.001 | 13,589 (31%) | 13,737 (31%) | 0.009 |
| Hypothyroidism | 10,965 (9%) | 11,173 (9%) | 0.005 | 4,002 (9%) | 3,830 (9%) | 0.013 |
| Malignant neoplastic disease | 12,621 (10%) | 12,418 (10%) | 0.007 | 4,304 (10%) | 3,910 (9%) | 0.030 |
| Myocardial infarction | 2,553 (2%) | 2,500 (2%) | 0.004 | 888 (2%) | 868 (2%) | 0.003 |
| Osteoporosis | 6,687 (6%) | 6,679 (6%) | 0.001 | 2,155 (5%) | 2,200 (5%) | 0.005 |
| Pneumonia | 6,452 (5%) | 6,392 (5%) | 0.003 | 2,243 (5%) | 2,167 (5%) | 0.007 |
| Rheumatoid arthritis | 1,009 (1%) | 931 (1%) | 0.008 | 337 (1%) | 259 (1%) | 0.021 |
| Stroke | 2,693 (2%) | 2,582 (2%) | 0.007 | 965 (2%) | 1,000 (2%) | 0.006 |
| Venous thromboembolism | 1,686 (1%) | 1,469 (1%) | 0.016 | 659 (1%) | 541 (1%) | 0.023 |

The 4 cohorts represent vaccine rollout periods. *Calculated as the days of previous observation in the database before index date. **Assessed anytime before index date. ASMD = Absolute standardized mean difference, COPD = Chronic obstructive pulmonary disease, GERD = Gastro-Esophageal reflux disease

**Table S18: Characteristics of unweighted populations in SIDIAP,** database, stratified by staggered cohort and exposure status. Exposure is ChAdOx1 vaccine.

| | Cohort 2 | | | Cohort 3 | | |
|---|---|---|---|---|---|---|
| | Unvaccinated | Vaccinated | ASMD | Unvaccinated | Vaccinated | ASMD |
| **N (individuals)** | 433,636 | 323,204 | | 873,400 | 84,204 | |
| **Age, median [Q25-Q75]** | 67 [62-72] | 64 [62-67] | 0.241 | 48 [43-57] | 64 [61-67] | 0.986 |
| **Sex: Female, N(%)** | 237,722 (55%) | 162,269 (50%) | 0.093 | 411,107 (47%) | 44,374 (53%) | 0.113 |
| **Years of prior history*, median [Q25-Q75]** | 15 [15-15] | 15 [15-15] | 0.158 | 15 [15-15] | 15 [15-15] | 0.281 |
| **Number visits, median [Q25-Q75]** | 3 [0-8] | 4 [1-9] | | 1 [0-5] | 4 [1-8] | |
| **Number pcrs, median [Q25-Q75]** | 0[0-0] | 0[0-1] | | 0[0-0] | 0[0-0] | |
| **Comorbidities**, N(%)** | | | | | | |
| Anxiety | 97,950 (23%) | 77,793 (24%) | 0.035 | 198,282 (23%) | 19,626 (23%) | 0.014 |
| Asthma | 19,861 (5%) | 13,894 (4%) | 0.014 | 36,915 (4%) | 3,283 (4%) | 0.017 |
| Chronic kidney disease | 32,497 (7%) | 12,206 (4%) | 0.162 | 22,570 (3%) | 3,199 (4%) | 0.069 |
| COPD | 28,826 (7%) | 18,658 (6%) | 0.036 | 19,103 (2%) | 4,996 (6%) | 0.191 |
| Dementia | 7,407 (2%) | 1,009 (0%) | 0.140 | 6,041 (1%) | 345 (0%) | 0.038 |
| Depressive disorder | 61,377 (14%) | 44,463 (14%) | 0.011 | 77,249 (9%) | 11,874 (14%) | 0.166 |
| Diabetes | 65,666 (15%) | 45,453 (14%) | 0.031 | 60,383 (7%) | 12,142 (14%) | 0.245 |
| GERD | 34,755 (8%) | 28,610 (9%) | 0.030 | 37,840 (4%) | 6,754 (8%) | 0.154 |
| Heart failure | 16,741 (4%) | 5,636 (2%) | 0.129 | 11,981 (1%) | 1,703 (2%) | 0.050 |
| Hypertension | 157,484 (36%) | 114,179 (35%) | 0.021 | 125,527 (14%) | 28,314 (34%) | 0.463 |
| Hypothyroidism | 42,257 (10%) | 30,350 (9%) | 0.012 | 50,476 (6%) | 7,763 (9%) | 0.131 |
| Malignant neoplastic disease | 81,510 (19%) | 36,352 (11%) | 0.212 | 49,066 (6%) | 7,842 (9%) | 0.141 |
| Myocardial infarction | 9,910 (2%) | 7,242 (2%) | 0.003 | 7,561 (1%) | 1,784 (2%) | 0.103 |
| Osteoporosis | 29,546 (7%) | 19,864 (6%) | 0.027 | 16,434 (2%) | 4,382 (5%) | 0.180 |
| Pneumonia | 27,865 (6%) | 18,329 (6%) | 0.032 | 34,617 (4%) | 4,445 (5%) | 0.063 |
| Rheumatoid arthritis | 3,762 (1%) | 2,557 (1%) | 0.008 | 3,246 (0%) | 535 (1%) | 0.037 |
| Stroke | 14,284 (3%) | 6,369 (2%) | 0.083 | 10,884 (1%) | 1,915 (2%) | 0.078 |
| Venous thromboembolism | 8,163 (2%) | 3,613 (1%) | 0.063 | 7,746 (1%) | 1,110 (1%) | 0.041 |

The 4 cohorts represent vaccine rollout periods. *Calculated as the days of previous observation in the database before index date. **Assessed anytime before index date. ASMD = Absolute standardized mean difference, COPD = Chronic obstructive pulmonary disease, GERD = Gastro-Esophageal reflux disease

**Table S19: Characteristics of weighted populations in CORIVA,** database, stratified by staggered cohort and exposure status. Exposure is any COVID-19 vaccine.

| | Cohort 1 | | | Cohort 2 | | | Cohort 3 | | | Cohort 4 | | |
|---|---|---|---|---|---|---|---|---|---|---|---|---|
| | **Unvaccinated** | **Vaccinated** | **ASMD** | **Unvaccinated** | **Vaccinated** | **ASMD** | **Unvaccinated** | **Vaccinated** | **ASMD** | **Unvaccinated** | **Vaccinated** | **ASMD** |
| **N (individuals)** | 10,630 | 10,630 | | 3,863 | 3,863 | | 17,929 | 17,929 | | 18,740 | 18,740 | |
| **Age, median [Q25-Q75]** | 78 [72-83] | 78 [72-83] | 0.000 | 70 [65-75] | 70 [65-75] | 0.000 | 52 [45-59] | 50 [45-59] | 0.000 | 40 [30-51] | 40 [30-51] | 0.000 |
| **Sex: Female, N(%)** | 7,165 (67%) | 7,165 (67%) | 0.000 | 2,439 (63%) | 2,439 (63%) | 0.000 | 9,054 (50%) | 9,054 (50%) | 0.000 | 8,493 (45%) | 8,493 (45%) | 0.000 |
| **Years of prior history*, median [Q25-Q75]** | 4 [4-4] | 4 [4-4] | 0.000 | 4 [4-4] | 4 [4-4] | 0.000 | 4 [4-4] | 4 [4-4] | 0.000 | 5 [4-5] | 5 [4-5] | 0.000 |
| **Number visits, median [Q25-Q75]** | 14 [4-24] | 14 [6-23] | | 12 [3-23] | 12 [5-22] | | 2 [0-11] | 4 [1-10] | | 2 [0-8] | 2 [0-8] | |
| **Number pcrs, median [Q25-Q75]** | 0[0-0] | 0[0-0] | | 0[0-0] | 0[0-0] | | 0[0-0] | 0[0-0] | | 0[0-0] | 0[0-0] | |
| **Comorbidities**, N(%)** | | | | | | | | | | | | |
| Anxiety | 1,060 (10%) | 1,076 (10%) | 0.005 | 405 (10%) | 415 (11%) | 0.008 | 1,545 (9%) | 1,500 (8%) | 0.009 | 1,552 (8%) | 1,495 (8%) | 0.011 |
| Asthma | 996 (9%) | 917 (9%) | 0.026 | 372 (10%) | 365 (9%) | 0.006 | 936 (5%) | 894 (5%) | 0.011 | 845 (5%) | 782 (4%) | 0.016 |
| Chronic kidney disease | 1,015 (10%) | 999 (9%) | 0.005 | 336 (9%) | 309 (8%) | 0.025 | 347 (2%) | 339 (2%) | 0.003 | 212 (1%) | 194 (1%) | 0.009 |
| COPD | 757 (7%) | 723 (7%) | 0.012 | 264 (7%) | 258 (7%) | 0.006 | 442 (2%) | 417 (2%) | 0.009 | 318 (2%) | 249 (1%) | 0.030 |
| Dementia | 438 (4%) | 412 (4%) | 0.013 | 106 (3%) | 90 (2%) | 0.027 | 120 (1%) | 131 (1%) | 0.008 | 71 (0%) | 70 (0%) | 0.001 |
| Depressive disorder | 1,076 (10%) | 1,083 (10%) | 0.002 | 462 (12%) | 433 (11%) | 0.023 | 1,787 (10%) | 1,764 (10%) | 0.004 | 1,679 (9%) | 1,739 (9%) | 0.011 |
| Diabetes | 1,861 (18%) | 1,833 (17%) | 0.007 | 640 (17%) | 588 (15%) | 0.037 | 1,012 (6%) | 963 (5%) | 0.012 | 814 (4%) | 764 (4%) | 0.014 |
| GERD | 1,416 (13%) | 1,458 (14%) | 0.011 | 523 (14%) | 531 (14%) | 0.006 | 1,562 (9%) | 1,600 (9%) | 0.007 | 1,166 (6%) | 1,172 (6%) | 0.001 |
| Heart failure | 3,898 (37%) | 3,839 (36%) | 0.011 | 1,003 (26%) | 970 (25%) | 0.020 | 1,348 (8%) | 1,324 (7%) | 0.005 | 781 (4%) | 737 (4%) | 0.012 |
| Hypertension | 8,016 (75%) | 8,221 (77%) | 0.045 | 2,569 (67%) | 2,641 (68%) | 0.040 | 5,521 (31%) | 5,643 (31%) | 0.015 | 3,359 (18%) | 3,538 (19%) | 0.025 |
| Hypothyroidism | 1,218 (11%) | 1,160 (11%) | 0.017 | 419 (11%) | 428 (11%) | 0.007 | 981 (5%) | 948 (5%) | 0.008 | 654 (3%) | 675 (4%) | 0.006 |
| Malignant neoplastic disease | 1,655 (16%) | 1,732 (16%) | 0.020 | 640 (17%) | 659 (17%) | 0.013 | 771 (4%) | 782 (4%) | 0.003 | 455 (2%) | 453 (2%) | 0.001 |
| Myocardial infarction | 269 (3%) | 264 (2%) | 0.003 | 86 (2%) | 71 (2%) | 0.026 | 127 (1%) | 116 (1%) | 0.008 | 81 (0%) | 75 (0%) | 0.005 |
| Osteoporosis | 675 (6%) | 685 (6%) | 0.004 | 187 (5%) | 176 (5%) | 0.013 | 233 (1%) | 230 (1%) | 0.001 | 131 (1%) | 146 (1%) | 0.009 |
| Pneumonia | 771 (7%) | 768 (7%) | 0.001 | 249 (6%) | 244 (6%) | 0.005 | 657 (4%) | 670 (4%) | 0.004 | 567 (3%) | 562 (3%) | 0.002 |
| Rheumatoid arthritis | 268 (3%) | 273 (3%) | 0.003 | 139 (4%) | 122 (3%) | 0.024 | 209 (1%) | 187 (1%) | 0.012 | 130 (1%) | 116 (1%) | 0.009 |

| | Cohort 1 | | | Cohort 2 | | | Cohort 3 | | | Cohort 4 | | |
|---|---|---|---|---|---|---|---|---|---|---|---|---|
| | **Unvaccinated** | **Vaccinated** | **ASMD** | **Unvaccinated** | **Vaccinated** | **ASMD** | **Unvaccinated** | **Vaccinated** | **ASMD** | **Unvaccinated** | **Vaccinated** | **ASMD** |
| Stroke | 460 (4%) | 461 (4%) | 0.001 | 147 (4%) | 144 (4%) | 0.004 | 179 (1%) | 189 (1%) | 0.006 | 112 (1%) | 116 (1%) | 0.003 |
| Venous thromboembolism | 469 (4%) | 472 (4%) | 0.001 | 171 (4%) | 123 (3%) | 0.064 | 306 (2%) | 301 (2%) | 0.002 | 219 (1%) | 201 (1%) | 0.009 |

The 4 cohorts represent vaccine rollout periods. *Calculated as the days of previous observation in the database before index date. **Assessed anytime before index date. ASMD = Absolute standardized mean difference, COPD = Chronic obstructive pulmonary disease, GERD = Gastro-Esophageal reflux disease

**Table S20: Characteristics of unweighted populations in CORIVA,** database, stratified by staggered cohort and exposure status. Exposure is any COVID-19 vaccine.

| | Cohort 1 | | | Cohort 2 | | | Cohort 3 | | | Cohort 4 | | |
|---|---|---|---|---|---|---|---|---|---|---|---|---|
| | **Unvaccinated** | **Vaccinated** | **ASMD** | **Unvaccinated** | **Vaccinated** | **ASMD** | **Unvaccinated** | **Vaccinated** | **ASMD** | **Unvaccinated** | **Vaccinated** | **ASMD** |
| **N (individuals)** | 23,982 | 26,736 | | 34,317 | 4,572 | | 96,423 | 24,050 | | 147,545 | 22,245 | |
| **Age, median [Q25-Q75]** | 79 [73-85] | 77 [73-82] | 0.127 | 72 [67-81] | 70 [66-75] | 0.177 | 54 [46-66] | 51 [45-59] | 0.227 | 42 [32-58] | 40 [30-51] | 0.186 |
| **Sex: Female, N(%)** | 16,532 (69%) | 17,342 (65%) | 0.087 | 21,862 (64%) | 2,891 (63%) | 0.010 | 48,984 (51%) | 12,233 (51%) | 0.001 | 72,057 (49%) | 10,026 (45%) | 0.076 |
| **Years of prior history*, median [Q25-Q75]** | 4 [4-4] | 4 [4-4] | 0.019 | 4 [4-4] | 4 [4-4] | 0.002 | 4 [4-4] | 4 [4-4] | 0.002 | 5 [4-5] | 5 [4-5] | 0.003 |
| **Number visits, median [Q25-Q75]** | 11 [1-22] | 16 [8-26] | | 9 [1-20] | 13 [6-23] | | 2 [0-10] | 5 [1-11] | | 1 [0-6] | 3 [1-8] | |
| **Number pcrs, median [Q25-Q75]** | 0[0-0] | 0[0-0] | | 0[0-0] | 0[0-0] | | 0[0-0] | 0[0-0] | | 0[0-0] | 0[0-0] | |
| **Comorbidities**, N(%)** | | | | | | | | | | | | |
| Anxiety | 1,966 (8%) | 3,238 (12%) | 0.130 | 2,817 (8%) | 513 (11%) | 0.102 | 6,406 (7%) | 2,162 (9%) | 0.087 | 9,287 (6%) | 1,847 (8%) | 0.077 |
| Asthma | 1,986 (8%) | 2,668 (10%) | 0.059 | 2,653 (8%) | 447 (10%) | 0.072 | 4,385 (5%) | 1,250 (5%) | 0.030 | 5,405 (4%) | 968 (4%) | 0.035 |
| Chronic kidney disease | 2,270 (9%) | 2,507 (9%) | 0.003 | 2,939 (9%) | 367 (8%) | 0.019 | 2,523 (3%) | 415 (2%) | 0.061 | 2,335 (2%) | 221 (1%) | 0.052 |
| COPD | 1,569 (7%) | 1,976 (7%) | 0.033 | 2,015 (6%) | 312 (7%) | 0.039 | 2,743 (3%) | 529 (2%) | 0.041 | 2,558 (2%) | 294 (1%) | 0.034 |
| Dementia | 1,152 (5%) | 819 (3%) | 0.090 | 1,297 (4%) | 100 (2%) | 0.094 | 1,147 (1%) | 151 (1%) | 0.059 | 1,067 (1%) | 76 (0%) | 0.052 |
| Depressive disorder | 2,066 (9%) | 3,233 (12%) | 0.114 | 3,189 (9%) | 535 (12%) | 0.079 | 7,286 (8%) | 2,561 (11%) | 0.108 | 10,034 (7%) | 2,152 (10%) | 0.105 |
| Diabetes | 3,971 (17%) | 4,734 (18%) | 0.030 | 4,818 (14%) | 712 (16%) | 0.043 | 6,065 (6%) | 1,228 (5%) | 0.051 | 6,594 (4%) | 901 (4%) | 0.021 |
| GERD | 2,533 (11%) | 4,585 (17%) | 0.192 | 3,364 (10%) | 667 (15%) | 0.147 | 6,287 (7%) | 2,322 (10%) | 0.115 | 7,455 (5%) | 1,427 (6%) | 0.059 |
| Heart failure | 8,724 (36%) | 9,642 (36%) | 0.007 | 9,570 (28%) | 1,139 (25%) | 0.068 | 9,982 (10%) | 1,607 (7%) | 0.132 | 9,182 (6%) | 828 (4%) | 0.115 |
| Hypertension | 16,651 (69%) | 21,677 (81%) | 0.272 | 20,654 (60%) | 3,167 (69%) | 0.191 | 29,974 (31%) | 7,555 (31%) | 0.007 | 27,992 (19%) | 4,172 (19%) | 0.006 |
| Hypothyroidism | 2,496 (10%) | 3,094 (12%) | 0.037 | 3,198 (9%) | 515 (11%) | 0.064 | 4,725 (5%) | 1,309 (5%) | 0.025 | 5,180 (4%) | 799 (4%) | 0.004 |
| Malignant neoplastic disease | 3,194 (13%) | 5,044 (19%) | 0.151 | 5,061 (15%) | 782 (17%) | 0.064 | 4,587 (5%) | 1,011 (4%) | 0.027 | 4,134 (3%) | 523 (2%) | 0.028 |
| Myocardial infarction | 548 (2%) | 743 (3%) | 0.031 | 696 (2%) | 86 (2%) | 0.011 | 840 (1%) | 146 (1%) | 0.031 | 777 (1%) | 87 (0%) | 0.020 |
| Osteoporosis | 1,273 (5%) | 2,015 (8%) | 0.091 | 1,446 (4%) | 212 (5%) | 0.021 | 1,545 (2%) | 289 (1%) | 0.034 | 1,408 (1%) | 164 (1%) | 0.024 |
| Pneumonia | 1,611 (7%) | 2,046 (8%) | 0.036 | 2,081 (6%) | 291 (6%) | 0.012 | 3,342 (3%) | 898 (4%) | 0.014 | 3,919 (3%) | 683 (3%) | 0.025 |

| | Cohort 1 | | | Cohort 2 | | | Cohort 3 | | | Cohort 4 | | |
|---|---|---|---|---|---|---|---|---|---|---|---|---|
| | **Unvaccinated** | **Vaccinated** | **ASMD** | **Unvaccinated** | **Vaccinated** | **ASMD** | **Unvaccinated** | **Vaccinated** | **ASMD** | **Unvaccinated** | **Vaccinated** | **ASMD** |
| Rheumatoid arthritis | 493 (2%) | 817 (3%) | 0.063 | 891 (3%) | 153 (3%) | 0.044 | 1,022 (1%) | 250 (1%) | 0.002 | 987 (1%) | 137 (1%) | 0.007 |
| Stroke | 1,137 (5%) | 1,011 (4%) | 0.048 | 1,450 (4%) | 169 (4%) | 0.027 | 1,359 (1%) | 232 (1%) | 0.041 | 1,256 (1%) | 133 (1%) | 0.030 |
| Venous thromboembolism | 1,058 (4%) | 1,207 (5%) | 0.005 | 1,349 (4%) | 149 (3%) | 0.036 | 1,808 (2%) | 387 (2%) | 0.020 | 1,788 (1%) | 235 (1%) | 0.015 |

The 4 cohorts represent vaccine rollout periods. *Calculated as the days of previous observation in the database before index date. **Assessed anytime before index date. ASMD = Absolute standardized mean difference, COPD = Chronic obstructive pulmonary disease, GERD = Gastro-Esophageal reflux disease

**Table S21: Characteristics of weighted populations in CORIVA,** database, stratified by staggered cohort and exposure status. Exposure is BNT162b2 vaccine.

| | **Cohort 1** | | | **Cohort 2** | | | **Cohort 3** | | | **Cohort 4** | | |
|---|---|---|---|---|---|---|---|---|---|---|---|---|
| | **Unvaccinated** | **Vaccinated** | **ASMD** | **Unvaccinated** | **Vaccinated** | **ASMD** | **Unvaccinated** | **Vaccinated** | **ASMD** | **Unvaccinated** | **Vaccinated** | **ASMD** |
| **N (individuals)** | 9,118 | 9,118 | | 3,465 | 3,465 | | 14,414 | 14,414 | | 13,759 | 13,759 | |
| **Age, median [Q25-Q75]** | 78 [73-83] | 78 [73-83] | 0.000 | 70 [65-75] | 70 [65-75] | 0.000 | 50 [45-59] | 50 [45-59] | 0.000 | 38 [30-50] | 38 [30-50] | 0.000 |
| **Sex: Female, N(%)** | 6,206 (68%) | 6,206 (68%) | 0.000 | 2,197 (63%) | 2,197 (63%) | 0.000 | 7,324 (51%) | 7,324 (51%) | 0.000 | 6,552 (48%) | 6,552 (48%) | 0.000 |
| **Years of prior history*, median [Q25-Q75]** | 4 [4-4] | 4 [4-4] | 0.000 | 4 [4-4] | 4 [4-4] | 0.000 | 4 [4-4] | 4 [4-4] | 0.000 | 5 [4-5] | 5 [4-5] | 0.000 |
| **Number visits, median [Q25-Q75]** | 14 [4-25] | 14 [7-24] | | 12 [3-23] | 12 [5-22] | | 2 [0-10] | 4 [1-10] | | 2 [0-8] | 2 [0-8] | |
| **Number pcrs, median [Q25-Q75]** | 0[0-0] | 0[0-0] | | 0[0-0] | 0[0-0] | | 0[0-0] | 0[0-0] | | 0[0-0] | 0[0-0] | |
| **Comorbidities**, N(%)** | | | | | | | | | | | | |
| Anxiety | 935 (10%) | 956 (10%) | 0.007 | 367 (11%) | 373 (11%) | 0.006 | 1,284 (9%) | 1,278 (9%) | 0.001 | 1,206 (9%) | 1,170 (9%) | 0.009 |
| Asthma | 891 (10%) | 822 (9%) | 0.026 | 340 (10%) | 333 (10%) | 0.007 | 747 (5%) | 718 (5%) | 0.009 | 644 (5%) | 615 (4%) | 0.010 |
| Chronic kidney disease | 939 (10%) | 905 (10%) | 0.012 | 302 (9%) | 278 (8%) | 0.025 | 225 (2%) | 220 (2%) | 0.003 | 144 (1%) | 132 (1%) | 0.008 |
| COPD | 666 (7%) | 639 (7%) | 0.011 | 238 (7%) | 227 (7%) | 0.013 | 333 (2%) | 316 (2%) | 0.008 | 223 (2%) | 171 (1%) | 0.032 |
| Dementia | 403 (4%) | 377 (4%) | 0.014 | 91 (3%) | 81 (2%) | 0.018 | 75 (1%) | 72 (0%) | 0.002 | 46 (0%) | 38 (0%) | 0.012 |
| Depressive disorder | 960 (11%) | 966 (11%) | 0.002 | 420 (12%) | 411 (12%) | 0.008 | 1,472 (10%) | 1,485 (10%) | 0.003 | 1,311 (10%) | 1,367 (10%) | 0.014 |
| Diabetes | 1,647 (18%) | 1,624 (18%) | 0.007 | 576 (17%) | 531 (15%) | 0.036 | 747 (5%) | 731 (5%) | 0.005 | 596 (4%) | 565 (4%) | 0.011 |
| GERD | 1,240 (14%) | 1,268 (14%) | 0.009 | 476 (14%) | 473 (14%) | 0.003 | 1,283 (9%) | 1,322 (9%) | 0.009 | 892 (6%) | 900 (7%) | 0.002 |
| Heart failure | 3,527 (39%) | 3,470 (38%) | 0.013 | 911 (26%) | 877 (25%) | 0.022 | 920 (6%) | 899 (6%) | 0.006 | 519 (4%) | 484 (4%) | 0.014 |
| Hypertension | 7,040 (77%) | 7,234 (79%) | 0.051 | 2,327 (67%) | 2,384 (69%) | 0.036 | 4,296 (30%) | 4,340 (30%) | 0.007 | 2,391 (17%) | 2,479 (18%) | 0.017 |
| Hypothyroidism | 1,080 (12%) | 1,035 (11%) | 0.015 | 376 (11%) | 388 (11%) | 0.011 | 770 (5%) | 743 (5%) | 0.009 | 500 (4%) | 503 (4%) | 0.001 |
| Malignant neoplastic disease | 1,485 (16%) | 1,565 (17%) | 0.023 | 579 (17%) | 594 (17%) | 0.012 | 582 (4%) | 606 (4%) | 0.008 | 324 (2%) | 317 (2%) | 0.003 |
| Myocardial infarction | 248 (3%) | 234 (3%) | 0.010 | 78 (2%) | 68 (2%) | 0.020 | 97 (1%) | 88 (1%) | 0.008 | 52 (0%) | 52 (0%) | 0.000 |
| Osteoporosis | 615 (7%) | 632 (7%) | 0.007 | 170 (5%) | 167 (5%) | 0.003 | 167 (1%) | 161 (1%) | 0.004 | 97 (1%) | 107 (1%) | 0.008 |
| Pneumonia | 698 (8%) | 696 (8%) | 0.001 | 222 (6%) | 222 (6%) | 0.001 | 543 (4%) | 505 (4%) | 0.014 | 418 (3%) | 429 (3%) | 0.005 |
| Rheumatoid arthritis | 240 (3%) | 239 (3%) | 0.001 | 128 (4%) | 110 (3%) | 0.028 | 167 (1%) | 148 (1%) | 0.013 | 97 (1%) | 88 (1%) | 0.008 |
| Stroke | 419 (5%) | 417 (5%) | 0.001 | 134 (4%) | 131 (4%) | 0.004 | 121 (1%) | 130 (1%) | 0.007 | 74 (1%) | 82 (1%) | 0.008 |

| | Cohort 1 | | | Cohort 2 | | | Cohort 3 | | | Cohort 4 | | |
|---|---|---|---|---|---|---|---|---|---|---|---|---|
| | Unvaccinated | Vaccinated | ASMD | Unvaccinated | Vaccinated | ASMD | Unvaccinated | Vaccinated | ASMD | Unvaccinated | Vaccinated | ASMD |
| Venous thromboembolism | 438 (5%) | 416 (5%) | 0.011 | 153 (4%) | 117 (3%) | 0.053 | 220 (2%) | 214 (1%) | 0.003 | 159 (1%) | 138 (1%) | 0.015 |

The 4 cohorts represent vaccine rollout periods. *Calculated as the days of previous observation in the database before index date. **Assessed anytime before index date. ASMD = Absolute standardized mean difference, COPD = Chronic obstructive pulmonary disease, GERD = Gastro-Esophageal reflux disease

**Table S22: Characteristics of unweighted populations in CORIVA,** database, stratified by staggered cohort and exposure status. Exposure is BNT162b2 vaccine.

| | Cohort 1 | | | Cohort 2 | | | Cohort 3 | | | Cohort 4 | | |
|---|---|---|---|---|---|---|---|---|---|---|---|---|
| | **Unvaccinated** | **Vaccinated** | **ASMD** | **Unvaccinated** | **Vaccinated** | **ASMD** | **Unvaccinated** | **Vaccinated** | **ASMD** | **Unvaccinated** | **Vaccinated** | **ASMD** |
| **N (individuals)** | 24,073 | 19,686 | | 34,320 | 4,067 | | 96,471 | 18,645 | | 147,553 | 15,683 | |
| **Age, median [Q25-Q75]** | 79 [73-85] | 79 [74-83] | 0.040 | 72 [67-81] | 70 [66-75] | 0.174 | 54 [46-66] | 51 [45-59] | 0.257 | 42 [32-58] | 38 [30-50] | 0.216 |
| **Sex: Female, N(%)** | 16,582 (69%) | 13,022 (66%) | 0.058 | 21,867 (64%) | 2,582 (63%) | 0.005 | 49,023 (51%) | 9,570 (51%) | 0.010 | 72,056 (49%) | 7,500 (48%) | 0.020 |
| **Years of prior history*, median [Q25-Q75]** | 4 [4-4] | 4 [4-4] | 0.026 | 4 [4-4] | 4 [4-4] | 0.000 | 4 [4-4] | 4 [4-4] | 0.000 | 5 [4-5] | 5 [4-5] | 0.003 |
| **Number visits, median [Q25-Q75]** | 11 [1-22] | 17 [9-27] | | 9 [1-20] | 13 [6-23] | | 2 [0-10] | 5 [1-11] | | 1 [0-6] | 4 [1-9] | |
| **Number pcrs, median [Q25-Q75]** | 0[0-0] | 0[0-0] | | 0[0-0] | 0[0-0] | | 0[0-0] | 0[0-0] | | 0[0-0] | 0[0-0] | |
| **Comorbidities**, N(%)** | | | | | | | | | | | | |
| Anxiety | 1,966 (8%) | 2,429 (12%) | 0.138 | 2,816 (8%) | 458 (11%) | 0.103 | 6,416 (7%) | 1,780 (10%) | 0.106 | 9,291 (6%) | 1,384 (9%) | 0.096 |
| Asthma | 2,004 (8%) | 2,029 (10%) | 0.068 | 2,653 (8%) | 404 (10%) | 0.078 | 4,393 (5%) | 969 (5%) | 0.030 | 5,403 (4%) | 730 (5%) | 0.050 |
| Chronic kidney disease | 2,284 (9%) | 2,041 (10%) | 0.029 | 2,943 (9%) | 326 (8%) | 0.020 | 2,537 (3%) | 263 (1%) | 0.087 | 2,339 (2%) | 146 (1%) | 0.059 |
| COPD | 1,587 (7%) | 1,501 (8%) | 0.040 | 2,013 (6%) | 274 (7%) | 0.036 | 2,746 (3%) | 388 (2%) | 0.049 | 2,560 (2%) | 193 (1%) | 0.042 |
| Dementia | 1,162 (5%) | 688 (3%) | 0.067 | 1,301 (4%) | 90 (2%) | 0.093 | 1,136 (1%) | 81 (0%) | 0.083 | 1,062 (1%) | 40 (0%) | 0.067 |
| Depressive disorder | 2,070 (9%) | 2,449 (12%) | 0.125 | 3,188 (9%) | 502 (12%) | 0.098 | 7,303 (8%) | 2,075 (11%) | 0.122 | 10,051 (7%) | 1,623 (10%) | 0.127 |
| Diabetes | 3,964 (16%) | 3,599 (18%) | 0.048 | 4,814 (14%) | 638 (16%) | 0.047 | 6,071 (6%) | 900 (5%) | 0.064 | 6,594 (4%) | 644 (4%) | 0.018 |
| GERD | 2,532 (11%) | 3,307 (17%) | 0.184 | 3,364 (10%) | 587 (14%) | 0.142 | 6,296 (7%) | 1,858 (10%) | 0.125 | 7,446 (5%) | 1,050 (7%) | 0.070 |
| Heart failure | 8,750 (36%) | 7,743 (39%) | 0.062 | 9,570 (28%) | 1,026 (25%) | 0.060 | 10,000 (10%) | 1,057 (6%) | 0.174 | 9,185 (6%) | 525 (3%) | 0.135 |
| Hypertension | 16,718 (69%) | 16,325 (83%) | 0.321 | 20,659 (60%) | 2,838 (70%) | 0.202 | 29,995 (31%) | 5,624 (30%) | 0.020 | 27,979 (19%) | 2,798 (18%) | 0.029 |
| Hypothyroidism | 2,526 (10%) | 2,352 (12%) | 0.046 | 3,197 (9%) | 464 (11%) | 0.069 | 4,732 (5%) | 988 (5%) | 0.018 | 5,173 (4%) | 576 (4%) | 0.009 |
| Malignant neoplastic disease | 3,216 (13%) | 3,873 (20%) | 0.171 | 5,060 (15%) | 700 (17%) | 0.067 | 4,587 (5%) | 764 (4%) | 0.032 | 4,139 (3%) | 352 (2%) | 0.036 |
| Myocardial infarction | 539 (2%) | 591 (3%) | 0.048 | 696 (2%) | 82 (2%) | 0.001 | 850 (1%) | 108 (1%) | 0.035 | 779 (1%) | 58 (0%) | 0.024 |
| Osteoporosis | 1,283 (5%) | 1,628 (8%) | 0.117 | 1,447 (4%) | 200 (5%) | 0.034 | 1,550 (2%) | 197 (1%) | 0.048 | 1,403 (1%) | 117 (1%) | 0.022 |
| Pneumonia | 1,621 (7%) | 1,624 (8%) | 0.058 | 2,080 (6%) | 264 (6%) | 0.018 | 3,353 (3%) | 672 (4%) | 0.007 | 3,920 (3%) | 499 (3%) | 0.031 |

| | Cohort 1 | | | Cohort 2 | | | Cohort 3 | | | Cohort 4 | | |
|---|---|---|---|---|---|---|---|---|---|---|---|---|
| | **Unvaccinated** | **Vaccinated** | **ASMD** | **Unvaccinated** | **Vaccinated** | **ASMD** | **Unvaccinated** | **Vaccinated** | **ASMD** | **Unvaccinated** | **Vaccinated** | **ASMD** |
| Rheumatoid arthritis | 491 (2%) | 603 (3%) | 0.065 | 892 (3%) | 137 (3%) | 0.045 | 1,025 (1%) | 190 (1%) | 0.004 | 990 (1%) | 99 (1%) | 0.005 |
| Stroke | 1,129 (5%) | 834 (4%) | 0.022 | 1,447 (4%) | 152 (4%) | 0.025 | 1,370 (1%) | 155 (1%) | 0.056 | 1,254 (1%) | 90 (1%) | 0.033 |
| Venous thromboembolism | 1,067 (4%) | 972 (5%) | 0.024 | 1,353 (4%) | 140 (3%) | 0.027 | 1,808 (2%) | 266 (1%) | 0.035 | 1,785 (1%) | 156 (1%) | 0.021 |

The 4 cohorts represent vaccine rollout periods. *Calculated as the days of previous observation in the database before index date. **Assessed anytime before index date. ASMD = Absolute standardized mean difference, COPD = Chronic obstructive pulmonary disease, GERD = Gastro-Esophageal reflux disease

**Table S23: Characteristics of weighted populations in CPRD AURUM** database, stratified by staggered cohort and vaccine brand.

| | Cohort 1 | | | Cohort 2 | | | Cohort 3 | | | Cohort 4 | | |
|---|---|---|---|---|---|---|---|---|---|---|---|---|
| | ChAdOx1 | BNT162b2 | ASMD | ChAdOx1 | BNT162b2 | ASMD | ChAdOx1 | BNT162b2 | ASMD | ChAdOx1 | BNT162b2 | ASMD |
| **N (individuals)** | 48,138 | 48,432 | | 306,114 | 307,972 | | 35,517 | 35,840 | | 117,220 | 116,722 | |
| **Age, median [Q25-Q75]** | 80 [77-85] | 80 [77-85] | 0.003 | 66 [57-72] | 66 [57-72] | 0.003 | 54 [46-60] | 54 [46-60] | 0.000 | 42 [39-46] | 42 [39-46] | 0.004 |
| **Sex: Female, N(%)** | 27,920 (58%) | 27,962 (58%) | 0.005 | 167,623 (55%) | 168,473 (55%) | 0.001 | 21,299 (60%) | 21,484 (60%) | 0.000 | 54,350 (46%) | 54,304 (47%) | 0.003 |
| **Years of prior history*, median [Q25-Q75]** | 24 [9-36] | 25 [11-36] | 0.011 | 21 [9-32] | 21 [9-32] | 0.000 | 17 [8-27] | 17 [7-27] | 0.007 | 9 [4-17] | 9 [4-17] | 0.001 |
| **Number visits, median [Q25-Q75]** | 12 [6-19] | 12 [6-19] | | 10 [5-16] | 10 [6-16] | | 8 [4-14] | 8 [4-14] | | 4 [1-8] | 4 [1-8] | |
| **Number pcrs, median [Q25-Q75]** | 0[0-0] | 0[0-0] | | 0[0-0] | 0[0-0] | | 0[0-0] | 0[0-0] | | 0[0-0] | 0[0-0] | |
| **Comorbidities**, N(%)** | | | | | | | | | | | | |
| Anxiety | 7,101 (15%) | 7,442 (15%) | 0.017 | 62,230 (20%) | 61,892 (20%) | 0.006 | 7,419 (21%) | 7,362 (21%) | 0.009 | 19,086 (16%) | 19,543 (17%) | 0.012 |
| Asthma | 5,241 (11%) | 5,396 (11%) | 0.008 | 57,043 (19%) | 56,551 (18%) | 0.007 | 6,365 (18%) | 6,097 (17%) | 0.024 | 9,186 (8%) | 9,059 (8%) | 0.003 |
| Chronic kidney disease | 11,403 (24%) | 11,809 (24%) | 0.016 | 28,219 (9%) | 29,790 (10%) | 0.016 | 1,565 (4%) | 1,700 (5%) | 0.016 | 793 (1%) | 897 (1%) | 0.011 |
| COPD | 4,080 (8%) | 4,014 (8%) | 0.007 | 20,765 (7%) | 20,456 (7%) | 0.006 | 661 (2%) | 669 (2%) | 0.000 | 424 (0%) | 471 (0%) | 0.007 |
| Dementia | 3,667 (8%) | 3,510 (7%) | 0.014 | 2,733 (1%) | 2,137 (1%) | 0.022 | 419 (1%) | 288 (1%) | 0.038 | 109 (0%) | 60 (0%) | 0.016 |
| Depressive disorder | 5,666 (12%) | 5,885 (12%) | 0.012 | 56,642 (19%) | 57,377 (19%) | 0.003 | 6,508 (18%) | 6,556 (18%) | 0.001 | 16,027 (14%) | 16,191 (14%) | 0.006 |
| Diabetes | 8,849 (18%) | 9,020 (19%) | 0.006 | 50,019 (16%) | 50,218 (16%) | 0.001 | 2,560 (7%) | 2,850 (8%) | 0.028 | 3,231 (3%) | 3,106 (3%) | 0.006 |
| GERD | 2,637 (5%) | 2,694 (6%) | 0.004 | 16,666 (5%) | 16,658 (5%) | 0.002 | 1,479 (4%) | 1,443 (4%) | 0.007 | 3,087 (3%) | 3,046 (3%) | 0.001 |
| Heart failure | 2,843 (6%) | 2,904 (6%) | 0.004 | 6,822 (2%) | 6,936 (2%) | 0.002 | 443 (1%) | 361 (1%) | 0.023 | 204 (0%) | 202 (0%) | 0.000 |
| Hypertension | 25,422 (53%) | 26,106 (54%) | 0.022 | 103,545 (34%) | 105,521 (34%) | 0.009 | 6,295 (18%) | 6,514 (18%) | 0.012 | 6,692 (6%) | 6,522 (6%) | 0.005 |
| Hypothyroidism | 4,806 (10%) | 4,978 (10%) | 0.010 | 22,667 (7%) | 23,290 (8%) | 0.006 | 1,845 (5%) | 1,886 (5%) | 0.003 | 2,985 (3%) | 3,025 (3%) | 0.003 |
| Malignant neoplastic disease | 10,958 (23%) | 11,284 (23%) | 0.013 | 38,507 (13%) | 40,499 (13%) | 0.017 | 2,019 (6%) | 2,152 (6%) | 0.014 | 1,504 (1%) | 1,576 (1%) | 0.006 |
| Myocardial infarction | 2,481 (5%) | 2,516 (5%) | 0.002 | 11,083 (4%) | 11,197 (4%) | 0.001 | 425 (1%) | 521 (1%) | 0.022 | 256 (0%) | 349 (0%) | 0.016 |
| Osteoporosis | 5,117 (11%) | 5,335 (11%) | 0.012 | 11,759 (4%) | 11,858 (4%) | 0.000 | 719 (2%) | 667 (2%) | 0.012 | 344 (0%) | 372 (0%) | 0.004 |
| Pneumonia | 2,576 (5%) | 2,527 (5%) | 0.006 | 9,543 (3%) | 9,423 (3%) | 0.003 | 808 (2%) | 692 (2%) | 0.024 | 979 (1%) | 904 (1%) | 0.007 |

| | Cohort 1 | | | Cohort 2 | | | Cohort 3 | | | Cohort 4 | | |
|---|---|---|---|---|---|---|---|---|---|---|---|---|
| | ChAdOx1 | BNT162b2 | ASMD | ChAdOx1 | BNT162b2 | ASMD | ChAdOx1 | BNT162b2 | ASMD | ChAdOx1 | BNT162b2 | ASMD |
| Rheumatoid arthritis | 935 (2%) | 1,005 (2%) | 0.010 | 6,476 (2%) | 6,640 (2%) | 0.003 | 284 (1%) | 344 (1%) | 0.017 | 216 (0%) | 258 (0%) | 0.008 |
| Stroke | 2,314 (5%) | 2,418 (5%) | 0.009 | 7,284 (2%) | 7,949 (3%) | 0.013 | 446 (1%) | 414 (1%) | 0.009 | 294 (0%) | 336 (0%) | 0.007 |
| Venous thromboembolism | 3,001 (6%) | 3,101 (6%) | 0.007 | 11,833 (4%) | 12,315 (4%) | 0.007 | 812 (2%) | 853 (2%) | 0.006 | 601 (1%) | 765 (1%) | 0.019 |

The 4 cohorts represent vaccine rollout periods. *Calculated as the days of previous observation in the database before index date. **Assessed anytime before index date. ASMD = Absolute standardized mean difference, COPD = Chronic obstructive pulmonary disease, GERD = Gastro-Esophageal reflux disease

**Table S24: Characteristics of unweighted populations in CPRD AURUM** database, stratified by staggered cohort and vaccine brand.

| | Cohort 1 | | | Cohort 2 | | | Cohort 3 | | | Cohort 4 | | |
|---|---|---|---|---|---|---|---|---|---|---|---|---|
| | ChAdOx1 | BNT162b2 | ASMD | ChAdOx1 | BNT162b2 | ASMD | ChAdOx1 | BNT162b2 | ASMD | ChAdOx1 | BNT162b2 | ASMD |
| **N (individuals)** | 219,804 | 332,790 | | 969,262 | 594,262 | | 1,473,602 | 54,102 | | 542,670 | 1,335,671 | |
| **Age, median [Q25-Q75]** | 80 [77-85] | 82 [79-86] | 0.129 | 69 [63-73] | 67 [56-73] | 0.148 | 55 [51-60] | 58 [50-82] | 0.381 | 45 [42-48] | 31 [25-37] | 1.319 |
| **Sex: Female, N(%)** | 127,656 (58%) | 186,481 (56%) | 0.041 | 528,692 (55%) | 324,259 (55%) | 0.000 | 739,444 (50%) | 32,310 (60%) | 0.193 | 242,758 (45%) | 625,195 (47%) | 0.042 |
| **Years of prior history*, median [Q25-Q75]** | 24 [9-36] | 26 [13-38] | 0.059 | 21 [9-33] | 21 [9-32] | 0.028 | 17 [8-27] | 19 [8-30] | 0.116 | 10 [5-18] | 7 [3-19] | 0.146 |
| **Number visits, median [Q25-Q75]** | 12 [6-19] | 11 [6-18] | | 10 [6-17] | 11 [6-17] | | 7 [3-12] | 10 [5-17] | | 3 [1-8] | 3 [1-7] | |
| **Number pcrs, median [Q25-Q75]** | 0[0-0] | 0[0-0] | | 0[0-0] | 0[0-0] | | 0[0-0] | 0[0-0] | | 0[0-0] | 0[0-0] | |
| **Comorbidities**, N(%)** | | | | | | | | | | | | |
| Anxiety | 33,355 (15%) | 49,410 (15%) | 0.009 | 187,469 (19%) | 122,087 (21%) | 0.030 | 281,846 (19%) | 10,441 (19%) | 0.004 | 86,805 (16%) | 218,144 (16%) | 0.009 |
| Asthma | 24,009 (11%) | 37,889 (11%) | 0.015 | 159,162 (16%) | 116,596 (20%) | 0.083 | 200,995 (14%) | 8,668 (16%) | 0.067 | 41,393 (8%) | 114,378 (9%) | 0.034 |
| Chronic kidney disease | 51,053 (23%) | 86,899 (26%) | 0.067 | 97,339 (10%) | 56,923 (10%) | 0.016 | 24,198 (2%) | 6,692 (12%) | 0.430 | 2,879 (1%) | 2,664 (0%) | 0.055 |
| COPD | 19,105 (9%) | 28,317 (9%) | 0.007 | 67,700 (7%) | 39,595 (7%) | 0.013 | 13,621 (1%) | 2,063 (4%) | 0.191 | 1,510 (0%) | 942 (0%) | 0.050 |
| Dementia | 16,795 (8%) | 16,138 (5%) | 0.116 | 12,106 (1%) | 3,720 (1%) | 0.065 | 1,896 (0%) | 1,323 (2%) | 0.207 | 447 (0%) | 119 (0%) | 0.034 |
| Depressive disorder | 27,009 (12%) | 37,831 (11%) | 0.028 | 170,626 (18%) | 112,753 (19%) | 0.035 | 256,789 (17%) | 9,046 (17%) | 0.019 | 74,210 (14%) | 155,037 (12%) | 0.062 |
| Diabetes | 39,751 (18%) | 61,929 (19%) | 0.014 | 154,348 (16%) | 98,676 (17%) | 0.018 | 78,394 (5%) | 5,817 (11%) | 0.201 | 9,729 (2%) | 12,040 (1%) | 0.077 |
| GERD | 12,156 (6%) | 20,204 (6%) | 0.023 | 52,694 (5%) | 32,288 (5%) | 0.000 | 56,444 (4%) | 2,506 (5%) | 0.040 | 13,807 (3%) | 21,599 (2%) | 0.065 |
| Heart failure | 13,147 (6%) | 20,430 (6%) | 0.007 | 24,276 (3%) | 13,120 (2%) | 0.020 | 5,673 (0%) | 1,529 (3%) | 0.195 | 681 (0%) | 448 (0%) | 0.033 |
| Hypertension | 114,922 (52%) | 186,367 (56%) | 0.075 | 345,014 (36%) | 200,762 (34%) | 0.038 | 224,543 (15%) | 15,536 (29%) | 0.330 | 31,330 (6%) | 19,649 (1%) | 0.232 |
| Hypothyroidism | 21,855 (10%) | 34,512 (10%) | 0.014 | 73,992 (8%) | 44,456 (7%) | 0.006 | 64,026 (4%) | 3,642 (7%) | 0.104 | 13,920 (3%) | 18,516 (1%) | 0.085 |
| Malignant neoplastic disease | 49,023 (22%) | 82,610 (25%) | 0.059 | 131,135 (14%) | 76,103 (13%) | 0.021 | 55,601 (4%) | 6,311 (12%) | 0.299 | 7,505 (1%) | 5,104 (0%) | 0.107 |
| Myocardial infarction | 11,236 (5%) | 18,289 (5%) | 0.017 | 33,142 (3%) | 23,019 (4%) | 0.024 | 11,237 (1%) | 1,464 (3%) | 0.149 | 861 (0%) | 715 (0%) | 0.032 |
| Osteoporosis | 22,986 (10%) | 36,535 (11%) | 0.017 | 42,319 (4%) | 21,789 (4%) | 0.036 | 12,455 (1%) | 2,632 (5%) | 0.243 | 1,380 (0%) | 1,028 (0%) | 0.044 |

| | Cohort 1 | | | Cohort 2 | | | Cohort 3 | | | Cohort 4 | | |
|---|---|---|---|---|---|---|---|---|---|---|---|---|
| | ChAdOx1 | BNT162b2 | ASMD | ChAdOx1 | BNT162b2 | ASMD | ChAdOx1 | BNT162b2 | ASMD | ChAdOx1 | BNT162b2 | ASMD |
| Pneumonia | 12,101 (6%) | 16,297 (5%) | 0.027 | 33,170 (3%) | 17,567 (3%) | 0.027 | 20,399 (1%) | 1,658 (3%) | 0.114 | 4,297 (1%) | 7,793 (1%) | 0.025 |
| Rheumatoid arthritis | 4,544 (2%) | 6,693 (2%) | 0.004 | 20,294 (2%) | 12,878 (2%) | 0.005 | 8,463 (1%) | 667 (1%) | 0.070 | 821 (0%) | 891 (0%) | 0.026 |
| Stroke | 10,876 (5%) | 15,601 (5%) | 0.012 | 24,851 (3%) | 15,056 (3%) | 0.002 | 9,547 (1%) | 1,281 (2%) | 0.141 | 1,153 (0%) | 1,022 (0%) | 0.036 |
| Venous thromboembolism | 13,782 (6%) | 21,549 (6%) | 0.008 | 39,415 (4%) | 23,632 (4%) | 0.005 | 24,312 (2%) | 1,980 (4%) | 0.125 | 1,936 (0%) | 2,541 (0%) | 0.032 |

The 4 cohorts represent vaccine rollout periods. *Calculated as the days of previous observation in the database before index date. **Assessed anytime before index date. ASMD = Absolute standardized mean difference, COPD = Chronic obstructive pulmonary disease, GERD = Gastro-Esophageal reflux disease

**Table S25: Characteristics of weighted populations in CPRD GOLD** database, stratified by staggered cohort and vaccine brand.

| | Cohort 1 | | | Cohort 2 | | | Cohort 3 | | | Cohort 4 | | |
|---|---|---|---|---|---|---|---|---|---|---|---|---|
| | **ChAdOx1** | **BNT162b2** | **ASMD** | **ChAdOx1** | **BNT162b2** | **ASMD** | **ChAdOx1** | **BNT162b2** | **ASMD** | **ChAdOx1** | **BNT162b2** | **ASMD** |
| **N (individuals)** | 7,490 | 7,490 | | 77,012 | 76,978 | | 24,590 | 24,597 | | 49,538 | 49,577 | |
| **Age, median [Q25-Q75]** | 82 [78-86] | 80 [78-86] | 0.000 | 70 [65-74] | 70 [65-74] | 0.004 | 54 [49-59] | 54 [49-59] | 0.009 | 42 [37-46] | 42 [37-47] | 0.003 |
| **Sex: Female, N(%)** | 4,266 (57%) | 4,266 (57%) | 0.000 | 41,394 (54%) | 41,607 (54%) | 0.006 | 13,052 (53%) | 13,078 (53%) | 0.002 | 22,308 (45%) | 22,520 (45%) | 0.008 |
| **Years of prior history*, median [Q25-Q75]** | 18 [12-21] | 18 [11-21] | 0.000 | 17 [12-20] | 17 [12-20] | 0.005 | 17 [12-20] | 17 [12-20] | 0.004 | 14 [6-18] | 14 [6-18] | 0.005 |
| **Number visits, median [Q25-Q75]** | 14 [9-21] | 14 [9-21] | | 10 [6-16] | 10 [6-17] | | 6 [2-11] | 6 [2-11] | | 2 [0-6] | 2 [0-7] | |
| **Number pcrs, median [Q25-Q75]** | 0[0-0] | 0[0-0] | | 0[0-0] | 0[0-0] | | 0[0-0] | 0[0-0] | | 0[0-0] | 0[0-0] | |
| **Comorbidities**, N(%)** | | | | | | | | | | | | |
| Anxiety | 1,008 (13%) | 1,015 (14%) | 0.002 | 12,765 (17%) | 12,897 (17%) | 0.005 | 5,218 (21%) | 5,293 (22%) | 0.007 | 8,501 (17%) | 8,578 (17%) | 0.004 |
| Asthma | 783 (10%) | 774 (10%) | 0.004 | 9,867 (13%) | 10,028 (13%) | 0.006 | 2,993 (12%) | 2,826 (11%) | 0.021 | 3,700 (7%) | 3,568 (7%) | 0.010 |
| Chronic kidney disease | 1,711 (23%) | 1,694 (23%) | 0.005 | 7,209 (9%) | 7,045 (9%) | 0.007 | 530 (2%) | 411 (2%) | 0.035 | 193 (0%) | 202 (0%) | 0.003 |
| COPD | 568 (8%) | 583 (8%) | 0.008 | 5,739 (7%) | 5,725 (7%) | 0.001 | 321 (1%) | 274 (1%) | 0.017 | 170 (0%) | 165 (0%) | 0.002 |
| Dementia | 543 (7%) | 495 (7%) | 0.025 | 726 (1%) | 737 (1%) | 0.001 | 55 (0%) | 74 (0%) | 0.015 | 28 (0%) | 21 (0%) | 0.006 |
| Depressive disorder | 733 (10%) | 729 (10%) | 0.002 | 11,461 (15%) | 11,746 (15%) | 0.011 | 4,478 (18%) | 4,427 (18%) | 0.005 | 7,038 (14%) | 7,119 (14%) | 0.004 |
| Diabetes | 1,114 (15%) | 1,107 (15%) | 0.003 | 10,260 (13%) | 10,130 (13%) | 0.005 | 1,051 (4%) | 963 (4%) | 0.018 | 768 (2%) | 654 (1%) | 0.019 |
| GERD | 451 (6%) | 447 (6%) | 0.002 | 3,691 (5%) | 3,753 (5%) | 0.004 | 763 (3%) | 770 (3%) | 0.001 | 975 (2%) | 978 (2%) | 0.000 |
| Heart failure | 395 (5%) | 411 (5%) | 0.009 | 1,763 (2%) | 1,819 (2%) | 0.005 | 125 (1%) | 81 (0%) | 0.028 | 50 (0%) | 54 (0%) | 0.002 |
| Hypertension | 2,761 (37%) | 2,754 (37%) | 0.002 | 21,627 (28%) | 21,686 (28%) | 0.002 | 3,184 (13%) | 3,261 (13%) | 0.009 | 1,842 (4%) | 1,975 (4%) | 0.014 |
| Hypothyroidism | 579 (8%) | 566 (8%) | 0.007 | 4,932 (6%) | 4,869 (6%) | 0.003 | 1,064 (4%) | 1,065 (4%) | 0.000 | 979 (2%) | 1,028 (2%) | 0.007 |
| Malignant neoplastic disease | 1,632 (22%) | 1,744 (23%) | 0.036 | 10,682 (14%) | 9,986 (13%) | 0.026 | 1,007 (4%) | 1,089 (4%) | 0.017 | 598 (1%) | 660 (1%) | 0.011 |
| Myocardial infarction | 330 (4%) | 325 (4%) | 0.004 | 2,919 (4%) | 2,943 (4%) | 0.002 | 239 (1%) | 217 (1%) | 0.010 | 90 (0%) | 85 (0%) | 0.002 |
| Osteoporosis | 665 (9%) | 638 (9%) | 0.013 | 3,388 (4%) | 3,496 (5%) | 0.007 | 323 (1%) | 314 (1%) | 0.003 | 103 (0%) | 92 (0%) | 0.005 |

| | Cohort 1 | | | Cohort 2 | | | Cohort 3 | | | Cohort 4 | | |
|---|---|---|---|---|---|---|---|---|---|---|---|---|
| | **ChAdOx1** | **BNT162b2** | **ASMD** | **ChAdOx1** | **BNT162b2** | **ASMD** | **ChAdOx1** | **BNT162b2** | **ASMD** | **ChAdOx1** | **BNT162b2** | **ASMD** |
| Pneumonia | 286 (4%) | 258 (3%) | 0.020 | 1,807 (2%) | 1,724 (2%) | 0.007 | 263 (1%) | 252 (1%) | 0.005 | 271 (1%) | 265 (1%) | 0.002 |
| Rheumatoid arthritis | 111 (1%) | 111 (1%) | 0.001 | 1,437 (2%) | 1,460 (2%) | 0.002 | 123 (1%) | 115 (0%) | 0.005 | 58 (0%) | 40 (0%) | 0.011 |
| Stroke | 325 (4%) | 321 (4%) | 0.003 | 2,134 (3%) | 2,072 (3%) | 0.005 | 207 (1%) | 148 (1%) | 0.028 | 96 (0%) | 91 (0%) | 0.002 |
| Venous thromboembolism | 306 (4%) | 341 (5%) | 0.023 | 2,125 (3%) | 2,153 (3%) | 0.002 | 288 (1%) | 285 (1%) | 0.001 | 146 (0%) | 198 (0%) | 0.018 |

The 4 cohorts represent vaccine rollout periods. *Calculated as the days of previous observation in the database before index date. **Assessed anytime before index date. ASMD = Absolute standardized mean difference, COPD = Chronic obstructive pulmonary disease, GERD = Gastro-Esophageal reflux disease

**Table S26: Characteristics of unweighted populations in CPRD GOLD** database, stratified by staggered cohort and vaccine brand.

| | Cohort 1 | | | Cohort 2 | | | Cohort 3 | | | Cohort 4 | | |
|---|---|---|---|---|---|---|---|---|---|---|---|---|
| | **ChAdOx1** | **BNT162b2** | **ASMD** | **ChAdOx1** | **BNT162b2** | **ASMD** | **ChAdOx1** | **BNT162b2** | **ASMD** | **ChAdOx1** | **BNT162b2** | **ASMD** |
| **N (individuals)** | 82,406 | 32,755 | | 302,999 | 180,670 | | 423,876 | 36,748 | | 147,744 | 365,096 | |
| **Age, median [Q25-Q75]** | 84 [82-88] | 81 [79-86] | 0.351 | 71 [66-76] | 70 [66-74] | 0.132 | 56 [52-61] | 54 [50-60] | 0.171 | 44 [41-48] | 31 [24-37] | 1.090 |
| **Sex: Female, N(%)** | 47,915 (58%) | 18,415 (56%) | 0.039 | 162,753 (54%) | 97,747 (54%) | 0.008 | 210,283 (50%) | 19,649 (53%) | 0.077 | 64,999 (44%) | 167,510 (46%) | 0.038 |
| **Years of prior history*, median [Q25-Q75]** | 17 [14-20] | 18 [13-21] | 0.001 | 17 [13-19] | 17 [12-19] | 0.014 | 17 [10-19] | 18 [12-20] | 0.107 | 14 [7-18] | 15 [6-18] | 0.017 |
| **Number visits, median [Q25-Q75]** | 14 [9-21] | 14 [9-21] | | 11 [7-18] | 10 [6-16] | | 6 [2-12] | 6 [2-11] | | 3 [0-7] | 2 [0-6] | |
| **Number pcrs, median [Q25-Q75]** | 0[0-0] | 0[0-0] | | 0[0-0] | 0[0-0] | | 0[0-0] | 0[0-0] | | 0[0-0] | 0[0-0] | |
| **Comorbidities**, N(%)** | | | | | | | | | | | | |
| Anxiety | 9,923 (12%) | 4,336 (13%) | 0.036 | 49,010 (16%) | 28,697 (16%) | 0.008 | 75,258 (18%) | 8,004 (22%) | 0.101 | 24,601 (17%) | 59,094 (16%) | 0.013 |
| Asthma | 8,491 (10%) | 3,250 (10%) | 0.013 | 39,988 (13%) | 21,396 (12%) | 0.041 | 48,034 (11%) | 4,053 (11%) | 0.010 | 10,517 (7%) | 33,240 (9%) | 0.073 |
| Chronic kidney disease | 21,564 (26%) | 7,430 (23%) | 0.081 | 34,168 (11%) | 15,033 (8%) | 0.100 | 7,553 (2%) | 566 (2%) | 0.019 | 739 (1%) | 578 (0%) | 0.060 |
| COPD | 6,837 (8%) | 2,417 (7%) | 0.034 | 27,472 (9%) | 11,545 (6%) | 0.100 | 6,334 (1%) | 348 (1%) | 0.050 | 640 (0%) | 285 (0%) | 0.070 |
| Dementia | 4,773 (6%) | 1,895 (6%) | 0.000 | 5,124 (2%) | 1,375 (1%) | 0.085 | 548 (0%) | 87 (0%) | 0.025 | 126 (0%) | 29 (0%) | 0.036 |
| Depressive disorder | 6,998 (8%) | 3,310 (10%) | 0.056 | 45,154 (15%) | 26,294 (15%) | 0.010 | 70,703 (17%) | 6,569 (18%) | 0.032 | 21,711 (15%) | 39,181 (11%) | 0.119 |
| Diabetes | 12,387 (15%) | 4,882 (15%) | 0.004 | 43,647 (14%) | 22,552 (12%) | 0.056 | 24,219 (6%) | 1,191 (3%) | 0.120 | 2,254 (2%) | 2,507 (1%) | 0.080 |
| GERD | 4,054 (5%) | 2,012 (6%) | 0.054 | 14,294 (5%) | 8,525 (5%) | 0.000 | 14,447 (3%) | 1,093 (3%) | 0.025 | 2,883 (2%) | 5,213 (1%) | 0.041 |
| Heart failure | 5,058 (6%) | 1,782 (5%) | 0.030 | 9,180 (3%) | 3,669 (2%) | 0.064 | 2,308 (1%) | 100 (0%) | 0.043 | 191 (0%) | 98 (0%) | 0.037 |
| Hypertension | 31,217 (38%) | 11,995 (37%) | 0.026 | 88,833 (29%) | 49,763 (28%) | 0.039 | 59,554 (14%) | 4,679 (13%) | 0.039 | 6,761 (5%) | 4,776 (1%) | 0.194 |
| Hypothyroidism | 7,042 (9%) | 2,373 (7%) | 0.048 | 20,521 (7%) | 11,106 (6%) | 0.025 | 17,382 (4%) | 1,553 (4%) | 0.006 | 3,245 (2%) | 4,180 (1%) | 0.082 |
| Malignant neoplastic disease | 18,950 (23%) | 6,972 (21%) | 0.041 | 42,687 (14%) | 22,617 (13%) | 0.046 | 17,903 (4%) | 1,522 (4%) | 0.004 | 2,192 (1%) | 1,796 (0%) | 0.100 |
| Myocardial infarction | 4,500 (5%) | 1,603 (5%) | 0.026 | 13,456 (4%) | 6,621 (4%) | 0.039 | 5,280 (1%) | 271 (1%) | 0.051 | 330 (0%) | 154 (0%) | 0.050 |
| Osteoporosis | 8,564 (10%) | 2,770 (8%) | 0.066 | 16,117 (5%) | 7,531 (4%) | 0.054 | 4,330 (1%) | 458 (1%) | 0.021 | 408 (0%) | 205 (0%) | 0.054 |

| | Cohort 1 | | | Cohort 2 | | | Cohort 3 | | | Cohort 4 | | |
|---|---|---|---|---|---|---|---|---|---|---|---|---|
| | ChAdOx1 | BNT162b2 | ASMD | ChAdOx1 | BNT162b2 | ASMD | ChAdOx1 | BNT162b2 | ASMD | ChAdOx1 | BNT162b2 | ASMD |
| Pneumonia | 3,231 (4%) | 1,032 (3%) | 0.042 | 8,604 (3%) | 3,418 (2%) | 0.062 | 4,527 (1%) | 345 (1%) | 0.013 | 921 (1%) | 1,696 (0%) | 0.022 |
| Rheumatoid arthritis | 1,257 (2%) | 512 (2%) | 0.003 | 5,999 (2%) | 2,929 (2%) | 0.027 | 2,405 (1%) | 154 (0%) | 0.021 | 191 (0%) | 111 (0%) | 0.035 |
| Stroke | 4,193 (5%) | 1,490 (5%) | 0.025 | 10,213 (3%) | 4,632 (3%) | 0.048 | 3,789 (1%) | 199 (1%) | 0.042 | 369 (0%) | 225 (0%) | 0.048 |
| Venous thromboembolism | 3,550 (4%) | 1,389 (4%) | 0.003 | 9,466 (3%) | 4,490 (2%) | 0.039 | 6,339 (1%) | 370 (1%) | 0.044 | 460 (0%) | 496 (0%) | 0.037 |

The 4 cohorts represent vaccine rollout periods. *Calculated as the days of previous observation in the database before index date. **Assessed anytime before index date. ASMD = Absolute standardized mean difference, COPD = Chronic obstructive pulmonary disease, GERD = Gastro-Esophageal reflux disease

**Table S27: Number of records (and risk per 10,000 individuals) for post COVID-19 cardiac and thromboembolic complications,** across cohorts and databases, stratified by exposure status (any COVID-19 vaccine). Follow-up ends at first vaccine dose after index date.

| Cohort | Time window | Outcome | AURUM | | CORIVA | | GOLD | | SIDIAP | |
|---|---|---|---|---|---|---|---|---|---|---|
| | | | Unvaccinated | Vaccinated | Unvaccinated | Vaccinated | Unvaccinated | Vaccinated | Unvaccinated | Vaccinated |
| **Cohort 1** | | | **N = 346,674** | **N = 552,602** | **N = 23,982** | **N = 26,736** | **N = 169,100** | **N = 118,507** | **N = 223,962** | **N = 89,941** |
| | **0 to 30 days** | **VTE** | 93 (2.68) | 45 (0.81) | 74 (30.86) | 5 (1.87) | 8 (0.47) | < 5 | 75 (3.35) | < 5 |
| | | **DVT** | 22 (0.63) | 10 (0.18) | 11 (4.59) | < 5 | < 5 | < 5 | 19 (0.85) | < 5 |
| | | **PE** | 75 (2.16) | 37 (0.67) | 66 (27.52) | < 5 | 7 (0.41) | < 5 | 60 (2.68) | < 5 |
| | | **ATE** | 22 (0.63) | 28 (0.51) | 92 (38.36) | 5 (1.87) | 6 (0.35) | < 5 | 75 (3.35) | 6 (0.67) |
| | | **IS** | 8 (0.23) | < 5 | 52 (21.68) | < 5 | < 5 | < 5 | 42 (1.88) | < 5 |
| | | **TIA** | < 5 | 6 (0.11) | 19 (7.92) | < 5 | < 5 | < 5 | 7 (0.31) | < 5 |
| | | **MI** | 11 (0.32) | 20 (0.36) | 29 (12.09) | 5 (1.87) | < 5 | < 5 | 28 (1.25) | < 5 |
| | | **HF** | 59 (1.70) | 73 (1.32) | 349 (145.53) | 22 (8.23) | 10 (0.59) | < 5 | 305 (13.62) | 23 (2.56) |
| | | **HS** | < 5 | < 5 | < 5 | < 5 | < 5 | < 5 | 7 (0.31) | < 5 |
| | | **MP** | < 5 | < 5 | < 5 | < 5 | < 5 | < 5 | < 5 | < 5 |
| | **31 to 90 days** | **VTE** | 19 (0.55) | 20 (0.36) | 23 (9.59) | < 5 | < 5 | < 5 | 16 (0.71) | < 5 |
| | | **DVT** | 10 (0.29) | 7 (0.13) | 12 (5.00) | < 5 | < 5 | < 5 | 10 (0.45) | < 5 |
| | | **PE** | 9 (0.26) | 13 (0.24) | 14 (5.84) | < 5 | < 5 | < 5 | 6 (0.27) | < 5 |
| | | **ATE** | 5 (0.14) | 16 (0.29) | 20 (8.34) | < 5 | < 5 | < 5 | 42 (1.88) | 5 (0.56) |
| | | **IS** | < 5 | < 5 | 11 (4.59) | < 5 | < 5 | < 5 | 20 (0.89) | < 5 |
| | | **TIA** | < 5 | 6 (0.11) | 6 (2.50) | < 5 | < 5 | < 5 | 14 (0.63) | < 5 |
| | | **MI** | < 5 | 7 (0.13) | 9 (3.75) | < 5 | < 5 | < 5 | 11 (0.49) | < 5 |
| | | **HF** | 30 (0.87) | 56 (1.01) | 109 (45.45) | 19 (7.11) | < 5 | < 5 | 87 (3.88) | 8 (0.89) |
| | | **HS** | < 5 | < 5 | < 5 | < 5 | < 5 | < 5 | < 5 | < 5 |
| | | **MP** | < 5 | < 5 | < 5 | < 5 | < 5 | < 5 | < 5 | < 5 |
| | **91 to 180 days** | **VTE** | 10 (0.29) | 9 (0.16) | 15 (6.25) | < 5 | < 5 | < 5 | 20 (0.89) | < 5 |
| | | **DVT** | 5 (0.14) | < 5 | 9 (3.75) | < 5 | < 5 | < 5 | 9 (0.40) | < 5 |
| | | **PE** | 5 (0.14) | 5 (0.09) | 8 (3.34) | < 5 | < 5 | < 5 | 11 (0.49) | < 5 |
| | | **ATE** | 11 (0.32) | 14 (0.25) | 21 (8.76) | < 5 | < 5 | < 5 | 30 (1.34) | < 5 |
| | | **IS** | < 5 | < 5 | 11 (4.59) | < 5 | < 5 | < 5 | 16 (0.71) | < 5 |
| | | **TIA** | 5 (0.14) | 6 (0.11) | 5 (2.08) | < 5 | < 5 | < 5 | 8 (0.36) | < 5 |
| | | **MI** | 5 (0.14) | < 5 | 7 (2.92) | < 5 | < 5 | < 5 | 7 (0.31) | < 5 |
| | | **HF** | 37 (1.07) | 51 (0.92) | 126 (52.54) | 17 (6.36) | < 5 | < 5 | 86 (3.84) | 7 (0.78) |
| | | **HS** | < 5 | < 5 | < 5 | < 5 | < 5 | < 5 | < 5 | < 5 |
| | | **MP** | < 5 | < 5 | < 5 | < 5 | < 5 | < 5 | < 5 | < 5 |
| | **181 to 365 days** | **VTE** | 10 (0.29) | 8 (0.14) | 42 (17.51) | 5 (1.87) | < 5 | < 5 | 11 (0.49) | < 5 |
| | | **DVT** | 5 (0.14) | < 5 | 12 (5.00) | < 5 | < 5 | < 5 | 6 (0.27) | < 5 |
| | | **PE** | 5 (0.14) | < 5 | 35 (14.59) | < 5 | < 5 | < 5 | 5 (0.22) | < 5 |
| | | **ATE** | 10 (0.29) | 19 (0.34) | 44 (18.35) | < 5 | < 5 | < 5 | 42 (1.88) | 8 (0.89) |
| | | **IS** | < 5 | 6 (0.11) | 26 (10.84) | < 5 | < 5 | < 5 | 22 (0.98) | 5 (0.56) |
| | | **TIA** | 6 (0.17) | < 5 | 16 (6.67) | < 5 | < 5 | < 5 | 10 (0.45) | < 5 |
| | | **MI** | < 5 | 10 (0.18) | 10 (4.17) | < 5 | < 5 | < 5 | 10 (0.45) | < 5 |
| | | **HF** | 40 (1.15) | 53 (0.96) | 222 (92.57) | 38 (14.21) | < 5 | 6 (0.51) | 85 (3.80) | 20 (2.22) |
| | | **HS** | < 5 | < 5 | < 5 | < 5 | < 5 | < 5 | < 5 | < 5 |
| | | **MP** | < 5 | < 5 | < 5 | < 5 | < 5 | < 5 | < 5 | < 5 |
| **Cohort 2** | | | **N = 1,975,726** | **N = 1,563,569** | **N = 34,317** | **N = 4,572** | **N = 583,399** | **N = 486,619** | **N = 433,151** | **N = 819,590** |
| | **0 to 30 days** | **VTE** | 241 (1.22) | 54 (0.35) | 71 (20.69) | < 5 | 31 (0.53) | 7 (0.14) | 263 (6.07) | 27 (0.33) |
| | | **DVT** | 41 (0.21) | 19 (0.12) | 10 (2.91) | < 5 | 8 (0.14) | < 5 | 65 (1.50) | 9 (0.11) |
| | | **PE** | 204 (1.03) | 38 (0.24) | 62 (18.07) | < 5 | 24 (0.41) | 6 (0.12) | 217 (5.01) | 20 (0.24) |

| Cohort | Time window | Outcome | AURUM | | CORIVA | | GOLD | | SIDIAP | |
|---|---|---|---|---|---|---|---|---|---|---|
| | | | Unvaccinated | Vaccinated | Unvaccinated | Vaccinated | Unvaccinated | Vaccinated | Unvaccinated | Vaccinated |
| | | **ATE** | 41 (0.21) | 25 (0.16) | 90 (26.23) | < 5 | < 5 | < 5 | 171 (3.95) | 29 (0.35) |
| | | **IS** | 7 (0.04) | 5 (0.03) | 52 (15.15) | < 5 | < 5 | < 5 | 93 (2.15) | 12 (0.15) |
| | | **TIA** | 8 (0.04) | < 5 | 17 (4.95) | < 5 | < 5 | < 5 | 16 (0.37) | 6 (0.07) |
| | | **MI** | 26 (0.13) | 17 (0.11) | 27 (7.87) | < 5 | < 5 | < 5 | 66 (1.52) | 11 (0.13) |
| | | **HF** | 45 (0.23) | 47 (0.30) | 314 (91.50) | < 5 | 5 (0.09) | < 5 | 379 (8.75) | 76 (0.93) |
| | | **HS** | 6 (0.03) | < 5 | < 5 | < 5 | < 5 | < 5 | 18 (0.42) | < 5 |
| | | **MP** | 12 (0.06) | < 5 | < 5 | < 5 | < 5 | < 5 | 13 (0.30) | < 5 |
| | **31 to 90 days** | **VTE** | 43 (0.22) | 13 (0.08) | 25 (7.29) | < 5 | < 5 | < 5 | 62 (1.43) | 9 (0.11) |
| | | **DVT** | 24 (0.12) | 6 (0.04) | 14 (4.08) | < 5 | < 5 | < 5 | 34 (0.78) | 5 (0.06) |
| | | **PE** | 23 (0.12) | 8 (0.05) | 13 (3.79) | < 5 | < 5 | < 5 | 35 (0.81) | < 5 |
| | | **ATE** | 18 (0.09) | 18 (0.12) | 20 (5.83) | < 5 | < 5 | < 5 | 87 (2.01) | 14 (0.17) |
| | | **IS** | < 5 | < 5 | 13 (3.79) | < 5 | < 5 | < 5 | 44 (1.02) | 6 (0.07) |
| | | **TIA** | < 5 | 7 (0.04) | < 5 | < 5 | < 5 | < 5 | 23 (0.53) | 5 (0.06) |
| | | **MI** | 13 (0.07) | 10 (0.06) | 9 (2.62) | < 5 | < 5 | < 5 | 27 (0.62) | < 5 |
| | | **HF** | 27 (0.14) | 30 (0.19) | 108 (31.47) | < 5 | < 5 | < 5 | 140 (3.23) | 29 (0.35) |
| | | **HS** | < 5 | < 5 | < 5 | < 5 | < 5 | < 5 | 8 (0.18) | < 5 |
| | | **MP** | 6 (0.03) | < 5 | < 5 | < 5 | < 5 | < 5 | < 5 | < 5 |
| | **91 to 180 days** | **VTE** | 28 (0.14) | 15 (0.10) | 21 (6.12) | < 5 | 6 (0.10) | < 5 | 59 (1.36) | 6 (0.07) |
| | | **DVT** | 13 (0.07) | 9 (0.06) | 11 (3.21) | < 5 | < 5 | < 5 | 32 (0.74) | 6 (0.07) |
| | | **PE** | 15 (0.08) | 6 (0.04) | 11 (3.21) | < 5 | < 5 | < 5 | 30 (0.69) | < 5 |
| | | **ATE** | 17 (0.09) | 15 (0.10) | 18 (5.25) | < 5 | < 5 | < 5 | 88 (2.03) | 19 (0.23) |
| | | **IS** | < 5 | < 5 | 7 (2.04) | < 5 | < 5 | < 5 | 48 (1.11) | 9 (0.11) |
| | | **TIA** | 9 (0.05) | 5 (0.03) | 6 (1.75) | < 5 | < 5 | < 5 | 19 (0.44) | 5 (0.06) |
| | | **MI** | 9 (0.05) | 10 (0.06) | 7 (2.04) | < 5 | < 5 | < 5 | 24 (0.55) | 6 (0.07) |
| | | **HF** | 22 (0.11) | 27 (0.17) | 129 (37.59) | < 5 | < 5 | < 5 | 115 (2.65) | 41 (0.50) |
| | | **HS** | < 5 | < 5 | < 5 | < 5 | < 5 | < 5 | 7 (0.16) | < 5 |
| | | **MP** | < 5 | < 5 | < 5 | < 5 | < 5 | < 5 | 7 (0.16) | < 5 |
| | **181 to 365 days** | **VTE** | 9 (0.05) | 12 (0.08) | 38 (11.07) | < 5 | < 5 | < 5 | 17 (0.39) | 23 (0.28) |
| | | **DVT** | < 5 | 5 (0.03) | 14 (4.08) | < 5 | < 5 | < 5 | 11 (0.25) | 14 (0.17) |
| | | **PE** | 5 (0.03) | 7 (0.04) | 28 (8.16) | < 5 | < 5 | < 5 | 8 (0.18) | 10 (0.12) |
| | | **ATE** | 12 (0.06) | 16 (0.10) | 44 (12.82) | < 5 | < 5 | < 5 | 68 (1.57) | 33 (0.40) |
| | | **IS** | < 5 | < 5 | 29 (8.45) | < 5 | < 5 | < 5 | 38 (0.88) | 11 (0.13) |
| | | **TIA** | < 5 | < 5 | 12 (3.50) | < 5 | < 5 | < 5 | 19 (0.44) | 13 (0.16) |
| | | **MI** | 8 (0.04) | 10 (0.06) | 8 (2.33) | < 5 | < 5 | < 5 | 13 (0.30) | 10 (0.12) |
| | | **HF** | 20 (0.10) | 31 (0.20) | 214 (62.36) | < 5 | < 5 | < 5 | 77 (1.78) | 41 (0.50) |
| | | **HS** | < 5 | < 5 | < 5 | < 5 | < 5 | < 5 | < 5 | 5 (0.06) |
| | | **MP** | < 5 | < 5 | < 5 | < 5 | < 5 | < 5 | < 5 | < 5 |
| **Cohort 3** | | | **N = 1,510,401** | **N = 1,528,031** | **N = 96,423** | **N = 24,050** | **N = 417,996** | **N = 462,832** | **N = 869,497** | **N = 954,232** |
| | **0 to 30 days** | **VTE** | 245 (1.62) | 25 (0.16) | 103 (10.68) | < 5 | 27 (0.65) | < 5 | 328 (3.77) | 27 (0.28) |
| | | **DVT** | 45 (0.30) | < 5 | 19 (1.97) | < 5 | 6 (0.14) | < 5 | 93 (1.07) | 7 (0.07) |
| | | **PE** | 209 (1.38) | 22 (0.14) | 86 (8.92) | < 5 | 22 (0.53) | < 5 | 263 (3.02) | 22 (0.23) |
| | | **ATE** | 29 (0.19) | 10 (0.07) | 97 (10.06) | < 5 | < 5 | < 5 | 213 (2.45) | 14 (0.15) |
| | | **IS** | < 5 | < 5 | 53 (5.50) | < 5 | < 5 | < 5 | 103 (1.18) | 5 (0.05) |
| | | **TIA** | 5 (0.03) | < 5 | 18 (1.87) | < 5 | < 5 | < 5 | 20 (0.23) | < 5 |
| | | **MI** | 20 (0.13) | 8 (0.05) | 31 (3.22) | < 5 | < 5 | < 5 | 96 (1.10) | 7 (0.07) |
| | | **HF** | 31 (0.21) | 18 (0.12) | 326 (33.81) | 13 (5.41) | < 5 | < 5 | 365 (4.20) | 23 (0.24) |
| | | **HS** | < 5 | < 5 | 5 (0.52) | < 5 | < 5 | < 5 | 20 (0.23) | < 5 |
| | | **MP** | 9 (0.06) | < 5 | < 5 | < 5 | < 5 | < 5 | 19 (0.22) | < 5 |
| | **31 to 90 days** | **VTE** | 44 (0.29) | < 5 | 32 (3.32) | < 5 | < 5 | < 5 | 85 (0.98) | 9 (0.09) |

| Cohort | Time window | Outcome | AURUM | | CORIVA | | GOLD | | SIDIAP | |
|---|---|---|---|---|---|---|---|---|---|---|
| | | | Unvaccinated | Vaccinated | Unvaccinated | Vaccinated | Unvaccinated | Vaccinated | Unvaccinated | Vaccinated |
| | | **DVT** | 20 (0.13) | < 5 | 19 (1.97) | < 5 | < 5 | < 5 | 57 (0.66) | 5 (0.05) |
| | | **PE** | 27 (0.18) | < 5 | 16 (1.66) | < 5 | < 5 | < 5 | 38 (0.44) | < 5 |
| | | **ATE** | 11 (0.07) | 7 (0.05) | 33 (3.42) | < 5 | < 5 | < 5 | 107 (1.23) | 10 (0.10) |
| | | **IS** | < 5 | < 5 | 17 (1.76) | < 5 | < 5 | < 5 | 52 (0.60) | 6 (0.06) |
| | | **TIA** | < 5 | < 5 | 5 (0.52) | < 5 | < 5 | < 5 | 27 (0.31) | < 5 |
| | | **MI** | 9 (0.06) | < 5 | 15 (1.56) | < 5 | < 5 | < 5 | 36 (0.41) | < 5 |
| | | **HF** | 15 (0.10) | 8 (0.05) | 131 (13.59) | 5 (2.08) | < 5 | < 5 | 140 (1.61) | 13 (0.14) |
| | | **HS** | < 5 | < 5 | < 5 | < 5 | < 5 | < 5 | 7 (0.08) | < 5 |
| | | **MP** | 8 (0.05) | < 5 | 6 (0.62) | < 5 | < 5 | < 5 | 7 (0.08) | < 5 |
| | **91 to 180 days** | **VTE** | 24 (0.16) | 8 (0.05) | 36 (3.73) | < 5 | < 5 | < 5 | 63 (0.72) | 8 (0.08) |
| | | **DVT** | 11 (0.07) | 6 (0.04) | 22 (2.28) | < 5 | < 5 | < 5 | 35 (0.40) | 7 (0.07) |
| | | **PE** | 14 (0.09) | < 5 | 15 (1.56) | < 5 | < 5 | < 5 | 32 (0.37) | < 5 |
| | | **ATE** | < 5 | 7 (0.05) | 25 (2.59) | < 5 | < 5 | < 5 | 115 (1.32) | 20 (0.21) |
| | | **IS** | < 5 | < 5 | 10 (1.04) | < 5 | < 5 | < 5 | 58 (0.67) | 12 (0.13) |
| | | **TIA** | < 5 | < 5 | 9 (0.93) | < 5 | < 5 | < 5 | 22 (0.25) | < 5 |
| | | **MI** | < 5 | < 5 | 10 (1.04) | < 5 | < 5 | < 5 | 38 (0.44) | 6 (0.06) |
| | | **HF** | 11 (0.07) | 6 (0.04) | 162 (16.80) | < 5 | < 5 | < 5 | 121 (1.39) | 15 (0.16) |
| | | **HS** | < 5 | < 5 | < 5 | < 5 | < 5 | < 5 | 9 (0.10) | < 5 |
| | | **MP** | < 5 | < 5 | < 5 | < 5 | < 5 | < 5 | 7 (0.08) | < 5 |
| | **181 to 365 days** | **VTE** | < 5 | 9 (0.06) | 59 (6.12) | < 5 | < 5 | < 5 | 37 (0.43) | 5 (0.05) |
| | | **DVT** | < 5 | < 5 | 30 (3.11) | < 5 | < 5 | < 5 | 23 (0.26) | < 5 |
| | | **PE** | < 5 | 6 (0.04) | 32 (3.32) | < 5 | < 5 | < 5 | 16 (0.18) | < 5 |
| | | **ATE** | < 5 | < 5 | 60 (6.22) | < 5 | < 5 | < 5 | 53 (0.61) | 18 (0.19) |
| | | **IS** | < 5 | < 5 | 30 (3.11) | < 5 | < 5 | < 5 | 24 (0.28) | 9 (0.09) |
| | | **TIA** | < 5 | < 5 | 17 (1.76) | < 5 | < 5 | < 5 | 12 (0.14) | < 5 |
| | | **MI** | < 5 | < 5 | 20 (2.07) | < 5 | < 5 | < 5 | 18 (0.21) | 7 (0.07) |
| | | **HF** | 5 (0.03) | < 5 | 260 (26.96) | 6 (2.49) | < 5 | < 5 | 62 (0.71) | 13 (0.14) |
| | | **HS** | < 5 | < 5 | < 5 | < 5 | < 5 | < 5 | < 5 | < 5 |
| | | **MP** | < 5 | < 5 | < 5 | < 5 | < 5 | < 5 | < 5 | < 5 |
| **Cohort 4** | | | **N = 2,027,763** | **N = 2,085,598** | **N = 147,545** | **N = 22,245** | **N = 469,876** | **N = 550,437** | **N = 1,061,634** | **N = 880,950** |
| | **0 to 30 days** | **VTE** | 334 (1.65) | 19 (0.09) | 105 (7.12) | < 5 | 36 (0.77) | 7 (0.13) | 349 (3.29) | 38 (0.43) |
| | | **DVT** | 55 (0.27) | 6 (0.03) | 21 (1.42) | < 5 | 6 (0.13) | < 5 | 104 (0.98) | 14 (0.16) |
| | | **PE** | 291 (1.44) | 13 (0.06) | 86 (5.83) | < 5 | 31 (0.66) | 7 (0.13) | 270 (2.54) | 25 (0.28) |
| | | **ATE** | 26 (0.13) | 5 (0.02) | 93 (6.30) | < 5 | < 5 | < 5 | 225 (2.12) | 35 (0.40) |
| | | **IS** | < 5 | < 5 | 52 (3.52) | < 5 | < 5 | < 5 | 111 (1.05) | 17 (0.19) |
| | | **TIA** | 6 (0.03) | < 5 | 16 (1.08) | < 5 | < 5 | < 5 | 27 (0.25) | < 5 |
| | | **MI** | 16 (0.08) | < 5 | 30 (2.03) | < 5 | < 5 | < 5 | 93 (0.88) | 15 (0.17) |
| | | **HF** | 28 (0.14) | < 5 | 313 (21.21) | 6 (2.70) | < 5 | < 5 | 368 (3.47) | 28 (0.32) |
| | | **HS** | < 5 | < 5 | < 5 | < 5 | < 5 | < 5 | 24 (0.23) | < 5 |
| | | **MP** | 12 (0.06) | < 5 | 6 (0.41) | < 5 | < 5 | < 5 | 29 (0.27) | 9 (0.10) |
| | **31 to 90 days** | **VTE** | 58 (0.29) | 11 (0.05) | 34 (2.30) | < 5 | 5 (0.11) | < 5 | 90 (0.85) | 12 (0.14) |
| | | **DVT** | 21 (0.10) | < 5 | 21 (1.42) | < 5 | < 5 | < 5 | 61 (0.57) | 7 (0.08) |
| | | **PE** | 40 (0.20) | 7 (0.03) | 16 (1.08) | < 5 | < 5 | < 5 | 38 (0.36) | 6 (0.07) |
| | | **ATE** | 12 (0.06) | < 5 | 32 (2.17) | < 5 | < 5 | < 5 | 114 (1.07) | 18 (0.20) |
| | | **IS** | < 5 | < 5 | 16 (1.08) | < 5 | < 5 | < 5 | 50 (0.47) | 7 (0.08) |
| | | **TIA** | < 5 | < 5 | 5 (0.34) | < 5 | < 5 | < 5 | 32 (0.30) | 7 (0.08) |
| | | **MI** | 9 (0.04) | < 5 | 14 (0.95) | < 5 | < 5 | < 5 | 40 (0.38) | < 5 |
| | | **HF** | 14 (0.07) | 8 (0.04) | 126 (8.54) | < 5 | < 5 | < 5 | 138 (1.30) | 17 (0.19) |
| | | **HS** | < 5 | < 5 | < 5 | < 5 | < 5 | < 5 | 9 (0.08) | < 5 |

| Cohort | Time window | Outcome | AURUM | | CORIVA | | GOLD | | SIDIAP | |
|---|---|---|---|---|---|---|---|---|---|---|
| | | | Unvaccinated | Vaccinated | Unvaccinated | Vaccinated | Unvaccinated | Vaccinated | Unvaccinated | Vaccinated |
| | | MP | 7 (0.03) | < 5 | 6 (0.41) | < 5 | < 5 | < 5 | 11 (0.10) | < 5 |
| | 91 to 180 days | VTE | 26 (0.13) | 9 (0.04) | 41 (2.78) | < 5 | < 5 | < 5 | 72 (0.68) | 11 (0.12) |
| | | DVT | 10 (0.05) | 7 (0.03) | 27 (1.83) | < 5 | < 5 | < 5 | 42 (0.40) | 6 (0.07) |
| | | PE | 17 (0.08) | < 5 | 15 (1.02) | < 5 | < 5 | < 5 | 34 (0.32) | 6 (0.07) |
| | | ATE | < 5 | 5 (0.02) | 24 (1.63) | < 5 | < 5 | < 5 | 129 (1.22) | 21 (0.24) |
| | | IS | < 5 | < 5 | 10 (0.68) | < 5 | < 5 | < 5 | 62 (0.58) | 11 (0.12) |
| | | TIA | < 5 | < 5 | 9 (0.61) | < 5 | < 5 | < 5 | 30 (0.28) | < 5 |
| | | MI | < 5 | < 5 | 9 (0.61) | < 5 | < 5 | < 5 | 40 (0.38) | 7 (0.08) |
| | | HF | 10 (0.05) | < 5 | 152 (10.30) | < 5 | < 5 | < 5 | 138 (1.30) | 19 (0.22) |
| | | HS | < 5 | < 5 | < 5 | < 5 | < 5 | < 5 | 14 (0.13) | < 5 |
| | | MP | < 5 | < 5 | < 5 | < 5 | < 5 | < 5 | 12 (0.11) | < 5 |
| | 181 to 365 days | VTE | < 5 | < 5 | 64 (4.34) | 5 (2.25) | < 5 | < 5 | 48 (0.45) | 9 (0.10) |
| | | DVT | < 5 | < 5 | 36 (2.44) | < 5 | < 5 | < 5 | 31 (0.29) | 5 (0.06) |
| | | PE | < 5 | < 5 | 31 (2.10) | < 5 | < 5 | < 5 | 18 (0.17) | < 5 |
| | | ATE | < 5 | < 5 | 56 (3.80) | < 5 | < 5 | < 5 | 51 (0.48) | 22 (0.25) |
| | | IS | < 5 | < 5 | 28 (1.90) | < 5 | < 5 | < 5 | 22 (0.21) | 15 (0.17) |
| | | TIA | < 5 | < 5 | 15 (1.02) | < 5 | < 5 | < 5 | 11 (0.10) | < 5 |
| | | MI | < 5 | < 5 | 19 (1.29) | < 5 | < 5 | < 5 | 19 (0.18) | 7 (0.08) |
| | | HF | < 5 | < 5 | 243 (16.47) | < 5 | < 5 | < 5 | 59 (0.56) | 11 (0.12) |
| | | HS | < 5 | < 5 | 6 (0.41) | < 5 | < 5 | < 5 | 5 (0.05) | < 5 |
| | | MP | < 5 | < 5 | < 5 | < 5 | < 5 | < 5 | 6 (0.06) | < 5 |

VTE = Venous thromboembolism (DVT + PE), DVT = Deep vein thrombosis, PE = Pulmonary embolism, ATE = Arterial thromboembolism (IS + TIA + MI), IS = Ischemic stroke, TIA = Transient ischemic attack, MI = Myocardial infarction, HF = Heart failure, HS = Hemorrhagic stroke, MP = Myocarditis or Pericarditis

**Table S28: Number of records (and risk per 10,000 individuals) for post COVID-19 cardiac and thromboembolic complications,** across cohorts and databases, stratified by exposure status (any COVID-19 vaccine). Only first outcome after COVID-19 captured.

| Cohort | Time window | Outcome | AURUM | | CORIVA | | GOLD | | SIDIAP | |
|---|---|---|---|---|---|---|---|---|---|---|
| | | | Unvaccinated | Vaccinated | Unvaccinated | Vaccinated | Unvaccinated | Vaccinated | Unvaccinated | Vaccinated |
| Cohort 1 | | | N = 346,674 | N = 552,602 | N = 23,982 | N = 26,736 | N = 169,100 | N = 118,507 | N = 223,962 | N = 89,941 |
| | 0 to 30 days | VTE | 154 (4.44) | 117 (2.12) | 93 (38.78) | 36 (13.46) | 15 (0.89) | 8 (0.68) | 251 (11.21) | 96 (10.67) |
| | | DVT | 40 (1.15) | 27 (0.49) | 17 (7.09) | 8 (2.99) | < 5 | < 5 | 77 (3.44) | 29 (3.22) |
| | | PE | 123 (3.55) | 95 (1.72) | 79 (32.94) | 30 (11.22) | 13 (0.77) | 7 (0.59) | 186 (8.30) | 77 (8.56) |
| | | ATE | 57 (1.64) | 70 (1.27) | 128 (53.37) | 50 (18.70) | 8 (0.47) | 7 (0.59) | 350 (15.63) | 208 (23.13) |
| | | IS | 15 (0.43) | 5 (0.09) | 79 (32.94) | 23 (8.60) | < 5 | < 5 | 192 (8.57) | 116 (12.90) |
| | | TIA | 13 (0.37) | 18 (0.33) | 25 (10.42) | 9 (3.37) | < 5 | < 5 | 58 (2.59) | 41 (4.56) |
| | | MI | 31 (0.89) | 46 (0.83) | 36 (15.01) | 26 (9.72) | 6 (0.35) | 5 (0.42) | 116 (5.18) | 63 (7.00) |
| | | HF | 135 (3.89) | 198 (3.58) | 439 (183.05) | 199 (74.43) | 21 (1.24) | 9 (0.76) | 1,205 (53.80) | 640 (71.16) |
| | | HS | < 5 | 7 (0.13) | < 5 | < 5 | < 5 | < 5 | 46 (2.05) | 14 (1.56) |
| | | MP | < 5 | < 5 | < 5 | < 5 | < 5 | < 5 | 16 (0.71) | < 5 |
| | 31 to 90 days | VTE | 32 (0.92) | 39 (0.71) | 32 (13.34) | 12 (4.49) | < 5 | < 5 | 88 (3.93) | 45 (5.00) |
| | | DVT | 14 (0.40) | 15 (0.27) | 19 (7.92) | 5 (1.87) | < 5 | < 5 | 53 (2.37) | 29 (3.22) |
| | | PE | 18 (0.52) | 25 (0.45) | 16 (6.67) | 7 (2.62) | < 5 | < 5 | 41 (1.83) | 20 (2.22) |
| | | ATE | 15 (0.43) | 43 (0.78) | 30 (12.51) | 23 (8.60) | < 5 | < 5 | 216 (9.64) | 127 (14.12) |
| | | IS | < 5 | < 5 | 18 (7.51) | 11 (4.11) | < 5 | < 5 | 110 (4.91) | 70 (7.78) |
| | | TIA | 6 (0.17) | 19 (0.34) | 10 (4.17) | 8 (2.99) | < 5 | < 5 | 70 (3.13) | 32 (3.56) |
| | | MI | 10 (0.29) | 20 (0.36) | 10 (4.17) | 6 (2.24) | < 5 | < 5 | 49 (2.19) | 29 (3.22) |
| | | HF | 72 (2.08) | 109 (1.97) | 134 (55.88) | 83 (31.04) | 8 (0.47) | 8 (0.68) | 473 (21.12) | 290 (32.24) |
| | | HS | < 5 | < 5 | < 5 | < 5 | < 5 | < 5 | 17 (0.76) | 12 (1.33) |
| | | MP | < 5 | < 5 | < 5 | < 5 | < 5 | < 5 | < 5 | < 5 |
| | 91 to 180 days | VTE | 17 (0.49) | 18 (0.33) | 19 (7.92) | 14 (5.24) | 6 (0.35) | < 5 | 62 (2.77) | 35 (3.89) |
| | | DVT | 7 (0.20) | 8 (0.14) | 9 (3.75) | 5 (1.87) | < 5 | < 5 | 37 (1.65) | 19 (2.11) |
| | | PE | 10 (0.29) | 10 (0.18) | 12 (5.00) | 10 (3.74) | < 5 | < 5 | 29 (1.29) | 18 (2.00) |
| | | ATE | 23 (0.66) | 28 (0.51) | 30 (12.51) | 22 (8.23) | < 5 | 6 (0.51) | 184 (8.22) | 104 (11.56) |
| | | IS | < 5 | 8 (0.14) | 20 (8.34) | 9 (3.37) | < 5 | < 5 | 114 (5.09) | 57 (6.34) |
| | | TIA | 10 (0.29) | 12 (0.22) | 6 (2.50) | 5 (1.87) | < 5 | < 5 | 40 (1.79) | 34 (3.78) |
| | | MI | 12 (0.35) | 8 (0.14) | 7 (2.92) | 11 (4.11) | < 5 | 5 (0.42) | 38 (1.70) | 23 (2.56) |
| | | HF | 58 (1.67) | 89 (1.61) | 157 (65.47) | 108 (40.39) | < 5 | 5 (0.42) | 398 (17.77) | 228 (25.35) |
| | | HS | < 5 | < 5 | < 5 | < 5 | < 5 | < 5 | 21 (0.94) | < 5 |
| | | MP | < 5 | < 5 | < 5 | < 5 | < 5 | < 5 | < 5 | < 5 |
| | 181 to 365 days | VTE | 10 (0.29) | 11 (0.20) | 45 (18.76) | 22 (8.23) | < 5 | < 5 | 26 (1.16) | 11 (1.22) |
| | | DVT | 5 (0.14) | 6 (0.11) | 16 (6.67) | 13 (4.86) | < 5 | < 5 | 16 (0.71) | 7 (0.78) |
| | | PE | 5 (0.14) | 5 (0.09) | 34 (14.18) | 10 (3.74) | < 5 | < 5 | 12 (0.54) | < 5 |
| | | ATE | 14 (0.40) | 23 (0.42) | 51 (21.27) | 40 (14.96) | < 5 | < 5 | 104 (4.64) | 39 (4.34) |
| | | IS | < 5 | 6 (0.11) | 30 (12.51) | 19 (7.11) | < 5 | < 5 | 54 (2.41) | 24 (2.67) |
| | | TIA | 8 (0.23) | 7 (0.13) | 16 (6.67) | 17 (6.36) | < 5 | < 5 | 25 (1.12) | 9 (1.00) |
| | | MI | < 5 | 11 (0.20) | 12 (5.00) | 12 (4.49) | < 5 | < 5 | 29 (1.29) | 7 (0.78) |
| | | HF | 49 (1.41) | 53 (0.96) | 253 (105.50) | 191 (71.44) | < 5 | 6 (0.51) | 195 (8.71) | 117 (13.01) |
| | | HS | < 5 | < 5 | < 5 | < 5 | < 5 | < 5 | 5 (0.22) | 7 (0.78) |
| | | MP | < 5 | < 5 | < 5 | < 5 | < 5 | < 5 | < 5 | < 5 |
| Cohort 2 | | | N = 1,975,726 | N = 1,563,569 | N = 34,317 | N = 4,572 | N = 583,399 | N = 486,619 | N = 433,151 | N = 819,590 |
| | 0 to 30 days | VTE | 345 (1.75) | 220 (1.41) | 88 (25.64) | 8 (17.50) | 42 (0.72) | 24 (0.49) | 404 (9.33) | 400 (4.88) |
| | | DVT | 74 (0.37) | 65 (0.42) | 16 (4.66) | < 5 | 9 (0.15) | 5 (0.10) | 119 (2.75) | 165 (2.01) |
| | | PE | 278 (1.41) | 165 (1.06) | 73 (21.27) | 5 (10.94) | 34 (0.58) | 19 (0.39) | 315 (7.27) | 269 (3.28) |

| Cohort | Time window | Outcome | AURUM | | CORIVA | | GOLD | | SIDIAP | |
|---|---|---|---|---|---|---|---|---|---|---|
| | | | Unvaccinated | Vaccinated | Unvaccinated | Vaccinated | Unvaccinated | Vaccinated | Unvaccinated | Vaccinated |
| | | ATE | 74 (0.37) | 104 (0.67) | 129 (37.59) | 5 (10.94) | 8 (0.14) | 6 (0.12) | 345 (7.96) | 669 (8.16) |
| | | IS | 11 (0.06) | 14 (0.09) | 79 (23.02) | < 5 | < 5 | < 5 | 188 (4.34) | 346 (4.22) |
| | | TIA | 17 (0.09) | 24 (0.15) | 24 (6.99) | < 5 | < 5 | < 5 | 46 (1.06) | 122 (1.49) |
| | | MI | 46 (0.23) | 68 (0.43) | 36 (10.49) | < 5 | 6 (0.10) | 6 (0.12) | 128 (2.96) | 239 (2.92) |
| | | HF | 73 (0.37) | 146 (0.93) | 393 (114.52) | 20 (43.74) | 5 (0.09) | 13 (0.27) | 637 (14.71) | 1,331 (16.24) |
| | | HS | 6 (0.03) | 5 (0.03) | < 5 | < 5 | < 5 | < 5 | 39 (0.90) | 77 (0.94) |
| | | MP | 21 (0.11) | 7 (0.04) | < 5 | < 5 | < 5 | < 5 | 29 (0.67) | 37 (0.45) |
| | 31 to 90 days | VTE | 85 (0.43) | 75 (0.48) | 37 (10.78) | < 5 | 7 (0.12) | 9 (0.18) | 106 (2.45) | 187 (2.28) |
| | | DVT | 48 (0.24) | 32 (0.20) | 23 (6.70) | < 5 | < 5 | 5 (0.10) | 61 (1.41) | 119 (1.45) |
| | | PE | 41 (0.21) | 45 (0.29) | 17 (4.95) | < 5 | < 5 | < 5 | 55 (1.27) | 87 (1.06) |
| | | ATE | 39 (0.20) | 92 (0.59) | 31 (9.03) | < 5 | 7 (0.12) | 9 (0.18) | 201 (4.64) | 437 (5.33) |
| | | IS | 6 (0.03) | 11 (0.07) | 21 (6.12) | < 5 | < 5 | < 5 | 96 (2.22) | 231 (2.82) |
| | | TIA | 11 (0.06) | 33 (0.21) | 6 (1.75) | < 5 | < 5 | 6 (0.12) | 54 (1.25) | 115 (1.40) |
| | | MI | 23 (0.12) | 50 (0.32) | 10 (2.91) | < 5 | < 5 | < 5 | 64 (1.48) | 113 (1.38) |
| | | HF | 50 (0.25) | 99 (0.63) | 143 (41.67) | 10 (21.87) | 5 (0.09) | 7 (0.14) | 265 (6.12) | 622 (7.59) |
| | | HS | 6 (0.03) | < 5 | < 5 | < 5 | < 5 | < 5 | 19 (0.44) | 27 (0.33) |
| | | MP | 14 (0.07) | < 5 | < 5 | < 5 | < 5 | < 5 | 8 (0.18) | 8 (0.10) |
| | 91 to 180 days | VTE | 38 (0.19) | 37 (0.24) | 24 (6.99) | 5 (10.94) | 8 (0.14) | < 5 | 100 (2.31) | 109 (1.33) |
| | | DVT | 21 (0.11) | 21 (0.13) | 11 (3.21) | < 5 | < 5 | < 5 | 56 (1.29) | 71 (0.87) |
| | | PE | 17 (0.09) | 19 (0.12) | 14 (4.08) | < 5 | 7 (0.12) | < 5 | 51 (1.18) | 46 (0.56) |
| | | ATE | 31 (0.16) | 43 (0.28) | 28 (8.16) | < 5 | 5 (0.09) | < 5 | 171 (3.95) | 394 (4.81) |
| | | IS | < 5 | < 5 | 17 (4.95) | < 5 | < 5 | < 5 | 88 (2.03) | 202 (2.46) |
| | | TIA | 13 (0.07) | 18 (0.12) | 7 (2.04) | < 5 | < 5 | < 5 | 40 (0.92) | 109 (1.33) |
| | | MI | 17 (0.09) | 25 (0.16) | 7 (2.04) | < 5 | < 5 | < 5 | 49 (1.13) | 104 (1.27) |
| | | HF | 31 (0.16) | 65 (0.42) | 167 (48.66) | 14 (30.62) | < 5 | < 5 | 212 (4.89) | 534 (6.52) |
| | | HS | < 5 | < 5 | < 5 | < 5 | < 5 | < 5 | 18 (0.42) | 35 (0.43) |
| | | MP | 5 (0.03) | < 5 | < 5 | < 5 | < 5 | < 5 | 12 (0.28) | 9 (0.11) |
| | 181 to 365 days | VTE | 16 (0.08) | 12 (0.08) | 47 (13.70) | < 5 | < 5 | < 5 | 31 (0.72) | 52 (0.63) |
| | | DVT | 8 (0.04) | 5 (0.03) | 24 (6.99) | < 5 | < 5 | < 5 | 18 (0.42) | 36 (0.44) |
| | | PE | 9 (0.05) | 8 (0.05) | 28 (8.16) | < 5 | < 5 | < 5 | 16 (0.37) | 19 (0.23) |
| | | ATE | 12 (0.06) | 17 (0.11) | 57 (16.61) | 5 (10.94) | < 5 | < 5 | 97 (2.24) | 151 (1.84) |
| | | IS | < 5 | < 5 | 36 (10.49) | < 5 | < 5 | < 5 | 54 (1.25) | 71 (0.87) |
| | | TIA | < 5 | 5 (0.03) | 15 (4.37) | < 5 | < 5 | < 5 | 21 (0.48) | 46 (0.56) |
| | | MI | 7 (0.04) | 10 (0.06) | 11 (3.21) | < 5 | < 5 | < 5 | 27 (0.62) | 45 (0.55) |
| | | HF | 19 (0.10) | 34 (0.22) | 268 (78.10) | 17 (37.18) | < 5 | < 5 | 94 (2.17) | 209 (2.55) |
| | | HS | < 5 | < 5 | < 5 | < 5 | < 5 | < 5 | < 5 | 10 (0.12) |
| | | MP | < 5 | < 5 | < 5 | < 5 | < 5 | < 5 | < 5 | < 5 |
| Cohort 3 | | | N = 1,510,401 | N = 1,528,031 | N = 96,423 | N = 24,050 | N = 417,996 | N = 462,832 | N = 869,497 | N = 954,232 |
| | 0 to 30 days | VTE | 268 (1.77) | 142 (0.93) | 122 (12.65) | 8 (3.33) | 32 (0.77) | 17 (0.37) | 425 (4.89) | 180 (1.89) |
| | | DVT | 53 (0.35) | 41 (0.27) | 25 (2.59) | 6 (2.49) | 7 (0.17) | < 5 | 134 (1.54) | 78 (0.82) |
| | | PE | 224 (1.48) | 105 (0.69) | 98 (10.16) | < 5 | 26 (0.62) | 13 (0.28) | 327 (3.76) | 118 (1.24) |
| | | ATE | 34 (0.23) | 49 (0.32) | 133 (13.79) | 11 (4.57) | < 5 | 12 (0.26) | 315 (3.62) | 275 (2.88) |
| | | IS | 5 (0.03) | 5 (0.03) | 73 (7.57) | 8 (3.33) | < 5 | < 5 | 155 (1.78) | 130 (1.36) |
| | | TIA | 6 (0.04) | 17 (0.11) | 26 (2.70) | < 5 | < 5 | < 5 | 32 (0.37) | 53 (0.56) |
| | | MI | 23 (0.15) | 27 (0.18) | 42 (4.36) | < 5 | < 5 | 9 (0.19) | 137 (1.58) | 113 (1.18) |
| | | HF | 33 (0.22) | 38 (0.25) | 398 (41.28) | 34 (14.14) | < 5 | < 5 | 478 (5.50) | 256 (2.68) |
| | | HS | < 5 | < 5 | 6 (0.62) | < 5 | < 5 | < 5 | 34 (0.39) | 35 (0.37) |
| | | MP | 14 (0.09) | 6 (0.04) | < 5 | < 5 | < 5 | < 5 | 27 (0.31) | 20 (0.21) |
| | 31 to 90 days | VTE | 58 (0.38) | 46 (0.30) | 45 (4.67) | 8 (3.33) | < 5 | 6 (0.13) | 115 (1.32) | 90 (0.94) |

| Cohort | Time window | Outcome | AURUM | | CORIVA | | GOLD | | SIDIAP | |
|---|---|---|---|---|---|---|---|---|---|---|
| | | | Unvaccinated | Vaccinated | Unvaccinated | Vaccinated | Unvaccinated | Vaccinated | Unvaccinated | Vaccinated |
| | | **DVT** | 28 (0.19) | 27 (0.18) | 30 (3.11) | < 5 | < 5 | < 5 | 79 (0.91) | 60 (0.63) |
| | | **PE** | 33 (0.22) | 19 (0.12) | 18 (1.87) | 5 (2.08) | < 5 | < 5 | 47 (0.54) | 37 (0.39) |
| | | **ATE** | 14 (0.09) | 33 (0.22) | 41 (4.25) | 12 (4.99) | < 5 | 8 (0.17) | 169 (1.94) | 207 (2.17) |
| | | **IS** | < 5 | < 5 | 24 (2.49) | 5 (2.08) | < 5 | < 5 | 74 (0.85) | 87 (0.91) |
| | | **TIA** | < 5 | 14 (0.09) | 6 (0.62) | 6 (2.49) | < 5 | < 5 | 50 (0.58) | 35 (0.37) |
| | | **MI** | 11 (0.07) | 16 (0.10) | 16 (1.66) | < 5 | < 5 | 5 (0.11) | 57 (0.66) | 93 (0.97) |
| | | **HF** | 22 (0.15) | 26 (0.17) | 175 (18.15) | 15 (6.24) | < 5 | < 5 | 185 (2.13) | 154 (1.61) |
| | | **HS** | < 5 | < 5 | < 5 | < 5 | < 5 | < 5 | 13 (0.15) | 19 (0.20) |
| | | **MP** | 9 (0.06) | 8 (0.05) | 7 (0.73) | < 5 | < 5 | < 5 | 12 (0.14) | 11 (0.12) |
| | **91 to 180 days** | **VTE** | 28 (0.19) | 25 (0.16) | 41 (4.25) | 9 (3.74) | 5 (0.12) | < 5 | 90 (1.04) | 92 (0.96) |
| | | **DVT** | 14 (0.09) | 15 (0.10) | 25 (2.59) | 5 (2.08) | < 5 | < 5 | 54 (0.62) | 61 (0.64) |
| | | **PE** | 16 (0.11) | 13 (0.09) | 16 (1.66) | < 5 | < 5 | < 5 | 41 (0.47) | 34 (0.36) |
| | | **ATE** | 8 (0.05) | 28 (0.18) | 43 (4.46) | 8 (3.33) | < 5 | < 5 | 172 (1.98) | 199 (2.09) |
| | | **IS** | < 5 | < 5 | 19 (1.97) | 5 (2.08) | < 5 | < 5 | 78 (0.90) | 92 (0.96) |
| | | **TIA** | < 5 | 9 (0.06) | 13 (1.35) | < 5 | < 5 | < 5 | 30 (0.35) | 47 (0.49) |
| | | **MI** | < 5 | 18 (0.12) | 16 (1.66) | < 5 | < 5 | < 5 | 66 (0.76) | 69 (0.72) |
| | | **HF** | 12 (0.08) | 12 (0.08) | 206 (21.36) | 18 (7.48) | < 5 | < 5 | 168 (1.93) | 133 (1.39) |
| | | **HS** | < 5 | < 5 | < 5 | < 5 | < 5 | < 5 | 16 (0.18) | 13 (0.14) |
| | | **MP** | < 5 | < 5 | 6 (0.62) | < 5 | < 5 | < 5 | 11 (0.13) | 14 (0.15) |
| | **181 to 365 days** | **VTE** | < 5 | 9 (0.06) | 69 (7.16) | 15 (6.24) | < 5 | < 5 | 39 (0.45) | 19 (0.20) |
| | | **DVT** | < 5 | < 5 | 42 (4.36) | 12 (4.99) | < 5 | < 5 | 24 (0.28) | 14 (0.15) |
| | | **PE** | < 5 | 6 (0.04) | 30 (3.11) | 5 (2.08) | < 5 | < 5 | 18 (0.21) | 6 (0.06) |
| | | **ATE** | < 5 | < 5 | 80 (8.30) | 8 (3.33) | < 5 | < 5 | 56 (0.64) | 58 (0.61) |
| | | **IS** | < 5 | < 5 | 37 (3.84) | < 5 | < 5 | < 5 | 28 (0.32) | 22 (0.23) |
| | | **TIA** | < 5 | < 5 | 23 (2.39) | < 5 | < 5 | < 5 | 11 (0.13) | 7 (0.07) |
| | | **MI** | < 5 | < 5 | 27 (2.80) | < 5 | < 5 | < 5 | 18 (0.21) | 32 (0.34) |
| | | **HF** | 5 (0.03) | < 5 | 314 (32.56) | 35 (14.55) | < 5 | < 5 | 63 (0.72) | 36 (0.38) |
| | | **HS** | < 5 | < 5 | 6 (0.62) | < 5 | < 5 | < 5 | < 5 | < 5 |
| | | **MP** | < 5 | < 5 | 5 (0.52) | < 5 | < 5 | < 5 | < 5 | < 5 |
| **Cohort 4** | | | **N = 2,027,763** | **N = 2,085,598** | **N = 147,545** | **N = 22,245** | **N = 469,876** | **N = 550,437** | **N = 1,061,634** | **N = 880,950** |
| | **0 to 30 days** | **VTE** | 337 (1.66) | 50 (0.24) | 120 (8.13) | < 5 | 36 (0.77) | 11 (0.20) | 388 (3.65) | 98 (1.11) |
| | | **DVT** | 55 (0.27) | 17 (0.08) | 24 (1.63) | < 5 | 6 (0.13) | < 5 | 123 (1.16) | 48 (0.54) |
| | | **PE** | 294 (1.45) | 34 (0.16) | 97 (6.57) | < 5 | 31 (0.66) | 8 (0.15) | 297 (2.80) | 55 (0.62) |
| | | **ATE** | 26 (0.13) | 8 (0.04) | 121 (8.20) | 10 (4.50) | < 5 | < 5 | 268 (2.52) | 95 (1.08) |
| | | **IS** | < 5 | < 5 | 69 (4.68) | < 5 | < 5 | < 5 | 136 (1.28) | 47 (0.53) |
| | | **TIA** | 6 (0.03) | < 5 | 22 (1.49) | < 5 | < 5 | < 5 | 36 (0.34) | 12 (0.14) |
| | | **MI** | 16 (0.08) | < 5 | 38 (2.58) | < 5 | < 5 | < 5 | 103 (0.97) | 39 (0.44) |
| | | **HF** | 28 (0.14) | < 5 | 370 (25.08) | 18 (8.09) | < 5 | < 5 | 430 (4.05) | 75 (0.85) |
| | | **HS** | < 5 | < 5 | < 5 | < 5 | < 5 | < 5 | 33 (0.31) | 13 (0.15) |
| | | **MP** | 13 (0.06) | 8 (0.04) | 7 (0.47) | < 5 | < 5 | < 5 | 31 (0.29) | 17 (0.19) |
| | **31 to 90 days** | **VTE** | 56 (0.28) | 22 (0.11) | 45 (3.05) | < 5 | 6 (0.13) | < 5 | 106 (1.00) | 48 (0.54) |
| | | **DVT** | 21 (0.10) | 12 (0.06) | 30 (2.03) | < 5 | < 5 | < 5 | 73 (0.69) | 32 (0.36) |
| | | **PE** | 38 (0.19) | 10 (0.05) | 18 (1.22) | < 5 | 5 (0.11) | < 5 | 42 (0.40) | 17 (0.19) |
| | | **ATE** | 12 (0.06) | 9 (0.04) | 39 (2.64) | < 5 | < 5 | < 5 | 141 (1.33) | 75 (0.85) |
| | | **IS** | < 5 | < 5 | 22 (1.49) | < 5 | < 5 | < 5 | 62 (0.58) | 30 (0.34) |
| | | **TIA** | < 5 | < 5 | 7 (0.47) | < 5 | < 5 | < 5 | 42 (0.40) | 23 (0.26) |
| | | **MI** | 9 (0.04) | 7 (0.03) | 15 (1.02) | < 5 | < 5 | < 5 | 46 (0.43) | 25 (0.28) |
| | | **HF** | 14 (0.07) | 9 (0.04) | 160 (10.84) | 16 (7.19) | < 5 | < 5 | 153 (1.44) | 47 (0.53) |
| | | **HS** | 5 (0.02) | < 5 | < 5 | < 5 | < 5 | < 5 | 12 (0.11) | 5 (0.06) |

| Cohort | Time window | Outcome | AURUM | | CORIVA | | GOLD | | SIDIAP | |
|---|---|---|---|---|---|---|---|---|---|---|
| | | | Unvaccinated | Vaccinated | Unvaccinated | Vaccinated | Unvaccinated | Vaccinated | Unvaccinated | Vaccinated |
| | | MP | 8 (0.04) | < 5 | 8 (0.54) | < 5 | < 5 | < 5 | 14 (0.13) | 11 (0.12) |
| | 91 to 180 days | VTE | 25 (0.12) | 9 (0.04) | 42 (2.85) | 7 (3.15) | < 5 | < 5 | 85 (0.80) | 52 (0.59) |
| | | DVT | 9 (0.04) | 7 (0.03) | 28 (1.90) | 5 (2.25) | < 5 | < 5 | 54 (0.51) | 33 (0.37) |
| | | PE | 17 (0.08) | < 5 | 15 (1.02) | < 5 | < 5 | < 5 | 36 (0.34) | 22 (0.25) |
| | | ATE | < 5 | 6 (0.03) | 32 (2.17) | 12 (5.39) | < 5 | < 5 | 141 (1.33) | 86 (0.98) |
| | | IS | < 5 | < 5 | 14 (0.95) | 5 (2.25) | < 5 | < 5 | 64 (0.60) | 42 (0.48) |
| | | TIA | < 5 | < 5 | 10 (0.68) | < 5 | < 5 | < 5 | 29 (0.27) | 17 (0.19) |
| | | MI | < 5 | < 5 | 12 (0.81) | < 5 | < 5 | < 5 | 50 (0.47) | 27 (0.31) |
| | | HF | 10 (0.05) | < 5 | 182 (12.34) | 13 (5.84) | < 5 | < 5 | 159 (1.50) | 54 (0.61) |
| | | HS | < 5 | < 5 | < 5 | < 5 | < 5 | < 5 | 17 (0.16) | 6 (0.07) |
| | | MP | < 5 | < 5 | 6 (0.41) | < 5 | < 5 | < 5 | 14 (0.13) | 6 (0.07) |
| | 181 to 365 days | VTE | < 5 | < 5 | 72 (4.88) | 10 (4.50) | < 5 | < 5 | 41 (0.39) | 8 (0.09) |
| | | DVT | < 5 | < 5 | 45 (3.05) | 9 (4.05) | < 5 | < 5 | 26 (0.24) | 6 (0.07) |
| | | PE | < 5 | < 5 | 30 (2.03) | < 5 | < 5 | < 5 | 16 (0.15) | < 5 |
| | | ATE | < 5 | < 5 | 67 (4.54) | 10 (4.50) | < 5 | < 5 | 42 (0.40) | 16 (0.18) |
| | | IS | < 5 | < 5 | 32 (2.17) | < 5 | < 5 | < 5 | 18 (0.17) | 11 (0.12) |
| | | TIA | < 5 | < 5 | 19 (1.29) | < 5 | < 5 | < 5 | 8 (0.08) | < 5 |
| | | MI | < 5 | < 5 | 22 (1.49) | < 5 | < 5 | < 5 | 17 (0.16) | 5 (0.06) |
| | | HF | < 5 | < 5 | 289 (19.59) | 13 (5.84) | < 5 | < 5 | 51 (0.48) | 11 (0.12) |
| | | HS | < 5 | < 5 | 7 (0.47) | < 5 | < 5 | < 5 | < 5 | < 5 |
| | | MP | < 5 | < 5 | 7 (0.47) | < 5 | < 5 | < 5 | 6 (0.06) | < 5 |

VTE = Venous thromboembolism (DVT + PE), DVT = Deep vein thrombosis, PE = Pulmonary embolism, ATE = Arterial thromboembolism (IS + TIA + MI), IS = Ischemic stroke, TIA = Transient ischemic attack, MI = Myocardial infarction, HF = Heart failure, HS = Hemorrhagic stroke, MP = Myocarditis or Pericarditis

**Table S29: Number of records (and risk per 10,000 individuals) for post COVID-19 cardiac and thromboembolic complications,** across cohorts and databases, stratified by exposure status (ChAdOx1 vaccine).

| Cohort | Time window | Outcome | AURUM | | GOLD | | SIDIAP | |
|---|---|---|---|---|---|---|---|---|
| | | | Unvaccinated | Vaccinated | Unvaccinated | Vaccinated | Unvaccinated | Vaccinated |
| Cohort 1 | | | N = 344,687 | N = 219,804 | N = 168,972 | N = 82,406 | | |
| | 0 to 30 days | VTE | 138 (4.00) | 41 (1.87) | 14 (0.83) | < 5 | | |
| | | DVT | 34 (0.99) | 12 (0.55) | < 5 | < 5 | | |
| | | PE | 112 (3.25) | 31 (1.41) | 12 (0.71) | < 5 | | |
| | | ATE | 53 (1.54) | 25 (1.14) | 9 (0.53) | 6 (0.73) | | |
| | | IS | 13 (0.38) | < 5 | < 5 | < 5 | | |
| | | TIA | 12 (0.35) | 7 (0.32) | < 5 | < 5 | | |
| | | MI | 29 (0.84) | 17 (0.77) | 7 (0.41) | < 5 | | |
| | | HF | 119 (3.45) | 72 (3.28) | 20 (1.18) | 5 (0.61) | | |
| | | HS | < 5 | < 5 | < 5 | < 5 | | |
| | | MP | < 5 | < 5 | < 5 | < 5 | | |
| | 31 to 90 days | VTE | 25 (0.73) | 14 (0.64) | < 5 | < 5 | | |
| | | DVT | 9 (0.26) | 8 (0.36) | < 5 | < 5 | | |
| | | PE | 16 (0.46) | 7 (0.32) | < 5 | < 5 | | |
| | | ATE | 16 (0.46) | 20 (0.91) | < 5 | < 5 | | |
| | | IS | < 5 | < 5 | < 5 | < 5 | | |
| | | TIA | 6 (0.17) | 9 (0.41) | < 5 | < 5 | | |
| | | MI | 11 (0.32) | 9 (0.41) | < 5 | < 5 | | |
| | | HF | 69 (2.00) | 42 (1.91) | 6 (0.36) | 5 (0.61) | | |
| | | HS | < 5 | < 5 | < 5 | < 5 | | |
| | | MP | < 5 | < 5 | < 5 | < 5 | | |
| | 91 to 180 days | VTE | 13 (0.38) | 15 (0.68) | 6 (0.36) | < 5 | | |
| | | DVT | 5 (0.15) | 5 (0.23) | < 5 | < 5 | | |
| | | PE | 8 (0.23) | 10 (0.45) | < 5 | < 5 | | |
| | | ATE | 22 (0.64) | 10 (0.45) | < 5 | 6 (0.73) | | |
| | | IS | < 5 | < 5 | < 5 | < 5 | | |
| | | TIA | 10 (0.29) | 6 (0.27) | < 5 | < 5 | | |
| | | MI | 11 (0.32) | < 5 | < 5 | 5 (0.61) | | |
| | | HF | 54 (1.57) | 38 (1.73) | < 5 | < 5 | | |
| | | HS | < 5 | < 5 | < 5 | < 5 | | |
| | | MP | < 5 | < 5 | < 5 | < 5 | | |
| | 181 to 365 days | VTE | 6 (0.17) | < 5 | < 5 | < 5 | | |
| | | DVT | < 5 | < 5 | < 5 | < 5 | | |
| | | PE | < 5 | < 5 | < 5 | < 5 | | |
| | | ATE | 13 (0.38) | 9 (0.41) | < 5 | < 5 | | |
| | | IS | < 5 | < 5 | < 5 | < 5 | | |
| | | TIA | 6 (0.17) | < 5 | < 5 | < 5 | | |
| | | MI | < 5 | < 5 | < 5 | < 5 | | |
| | | HF | 49 (1.42) | 20 (0.91) | < 5 | 5 (0.61) | | |
| | | HS | < 5 | < 5 | < 5 | < 5 | | |
| | | MP | < 5 | < 5 | < 5 | < 5 | | |
| Cohort 2 | | | N = 1,975,770 | N = 969,262 | N = 582,791 | N = 302,999 | N = 433,636 | N = 323,204 |
| | 0 to 30 days | VTE | 347 (1.76) | 158 (1.63) | 44 (0.75) | 19 (0.63) | 403 (9.29) | 90 (2.78) |
| | | DVT | 75 (0.38) | 47 (0.48) | 9 (0.15) | < 5 | 122 (2.81) | 45 (1.39) |
| | | PE | 280 (1.42) | 120 (1.24) | 36 (0.62) | 15 (0.50) | 311 (7.17) | 55 (1.70) |

| Cohort | Time window | Outcome | AURUM | | GOLD | | SIDIAP | |
|---|---|---|---|---|---|---|---|---|
| | | | Unvaccinated | Vaccinated | Unvaccinated | Vaccinated | Unvaccinated | Vaccinated |
| | | **ATE** | 70 (0.35) | 75 (0.77) | 9 (0.15) | < 5 | 341 (7.86) | 131 (4.05) |
| | | **IS** | 10 (0.05) | 12 (0.12) | < 5 | < 5 | 186 (4.29) | 52 (1.61) |
| | | **TIA** | 15 (0.08) | 13 (0.13) | < 5 | < 5 | 45 (1.04) | 23 (0.71) |
| | | **MI** | 45 (0.23) | 52 (0.54) | 7 (0.12) | < 5 | 127 (2.93) | 62 (1.92) |
| | | **HF** | 74 (0.37) | 103 (1.06) | 5 (0.09) | 11 (0.36) | 642 (14.81) | 123 (3.81) |
| | | **HS** | 5 (0.03) | < 5 | < 5 | < 5 | 38 (0.88) | 7 (0.22) |
| | | **MP** | 20 (0.10) | 6 (0.06) | < 5 | < 5 | 29 (0.67) | 10 (0.31) |
| | **31 to 90 days** | **VTE** | 93 (0.47) | 55 (0.57) | 6 (0.10) | 9 (0.30) | 107 (2.47) | 36 (1.11) |
| | | **DVT** | 50 (0.25) | 25 (0.26) | < 5 | 5 (0.17) | 60 (1.38) | 25 (0.77) |
| | | **PE** | 46 (0.23) | 32 (0.33) | < 5 | < 5 | 57 (1.31) | 18 (0.56) |
| | | **ATE** | 37 (0.19) | 57 (0.59) | 8 (0.14) | 5 (0.17) | 204 (4.70) | 107 (3.31) |
| | | **IS** | 6 (0.03) | 6 (0.06) | < 5 | < 5 | 97 (2.24) | 58 (1.79) |
| | | **TIA** | 11 (0.06) | 25 (0.26) | < 5 | < 5 | 55 (1.27) | 24 (0.74) |
| | | **MI** | 21 (0.11) | 28 (0.29) | < 5 | < 5 | 65 (1.50) | 27 (0.84) |
| | | **HF** | 47 (0.24) | 68 (0.70) | 6 (0.10) | 7 (0.23) | 269 (6.20) | 62 (1.92) |
| | | **HS** | 6 (0.03) | < 5 | < 5 | < 5 | 18 (0.42) | 7 (0.22) |
| | | **MP** | 14 (0.07) | < 5 | < 5 | < 5 | 8 (0.18) | < 5 |
| | **91 to 180 days** | **VTE** | 39 (0.20) | 30 (0.31) | 6 (0.10) | < 5 | 118 (2.72) | 23 (0.71) |
| | | **DVT** | 20 (0.10) | 16 (0.17) | < 5 | < 5 | 66 (1.52) | 16 (0.50) |
| | | **PE** | 19 (0.10) | 16 (0.17) | 6 (0.10) | < 5 | 60 (1.38) | 8 (0.25) |
| | | **ATE** | 28 (0.14) | 28 (0.29) | 5 (0.09) | < 5 | 187 (4.31) | 106 (3.28) |
| | | **IS** | < 5 | < 5 | < 5 | < 5 | 96 (2.21) | 40 (1.24) |
| | | **TIA** | 12 (0.06) | 13 (0.13) | < 5 | < 5 | 42 (0.97) | 32 (0.99) |
| | | **MI** | 15 (0.08) | 15 (0.15) | < 5 | < 5 | 55 (1.27) | 37 (1.14) |
| | | **HF** | 26 (0.13) | 44 (0.45) | < 5 | < 5 | 231 (5.33) | 60 (1.86) |
| | | **HS** | < 5 | < 5 | < 5 | < 5 | 17 (0.39) | < 5 |
| | | **MP** | < 5 | < 5 | < 5 | < 5 | 12 (0.28) | < 5 |
| | **181 to 365 days** | **VTE** | 23 (0.12) | 5 (0.05) | < 5 | < 5 | 35 (0.81) | 16 (0.50) |
| | | **DVT** | 9 (0.05) | < 5 | < 5 | < 5 | 19 (0.44) | 13 (0.40) |
| | | **PE** | 15 (0.08) | < 5 | < 5 | < 5 | 18 (0.42) | < 5 |
| | | **ATE** | 13 (0.07) | 11 (0.11) | < 5 | < 5 | 110 (2.54) | 46 (1.42) |
| | | **IS** | < 5 | < 5 | < 5 | < 5 | 57 (1.31) | 21 (0.65) |
| | | **TIA** | < 5 | < 5 | < 5 | < 5 | 25 (0.58) | 13 (0.40) |
| | | **MI** | 9 (0.05) | 7 (0.07) | < 5 | < 5 | 33 (0.76) | 16 (0.50) |
| | | **HF** | 20 (0.10) | 23 (0.24) | < 5 | < 5 | 110 (2.54) | 26 (0.80) |
| | | **HS** | < 5 | < 5 | < 5 | < 5 | 6 (0.14) | < 5 |
| | | **MP** | < 5 | < 5 | < 5 | < 5 | < 5 | < 5 |
| **Cohort 3** | | | **N = 1,510,493** | **N = 1,473,602** | **N = 418,184** | **N = 423,876** | **N = 873,400** | **N = 84,204** |
| | **0 to 30 days** | **VTE** | 267 (1.77) | 139 (0.94) | 33 (0.79) | 16 (0.38) | 431 (4.93) | 27 (3.21) |
| | | **DVT** | 52 (0.34) | 41 (0.28) | 7 (0.17) | < 5 | 137 (1.57) | 9 (1.07) |
| | | **PE** | 224 (1.48) | 101 (0.69) | 27 (0.65) | 12 (0.28) | 331 (3.79) | 22 (2.61) |
| | | **ATE** | 33 (0.22) | 47 (0.32) | < 5 | 12 (0.28) | 319 (3.65) | 32 (3.80) |
| | | **IS** | 5 (0.03) | 5 (0.03) | < 5 | < 5 | 154 (1.76) | 13 (1.54) |
| | | **TIA** | 6 (0.04) | 16 (0.11) | < 5 | < 5 | 33 (0.38) | 5 (0.59) |
| | | **MI** | 22 (0.15) | 26 (0.18) | < 5 | 9 (0.21) | 141 (1.61) | 16 (1.90) |
| | | **HF** | 32 (0.21) | 28 (0.19) | < 5 | < 5 | 486 (5.56) | 51 (6.06) |
| | | **HS** | < 5 | < 5 | < 5 | < 5 | 34 (0.39) | < 5 |
| | | **MP** | 14 (0.09) | 6 (0.04) | < 5 | < 5 | 27 (0.31) | 6 (0.71) |
| | **31 to 90 days** | **VTE** | 63 (0.42) | 44 (0.30) | < 5 | 7 (0.17) | 121 (1.39) | 9 (1.07) |

| Cohort | Time window | Outcome | AURUM | | GOLD | | SIDIAP | |
|---|---|---|---|---|---|---|---|---|
| | | | Unvaccinated | Vaccinated | Unvaccinated | Vaccinated | Unvaccinated | Vaccinated |
| | | DVT | 30 (0.20) | 26 (0.18) | < 5 | < 5 | 82 (0.94) | < 5 |
| | | PE | 36 (0.24) | 18 (0.12) | < 5 | < 5 | 51 (0.58) | 6 (0.71) |
| | | ATE | 14 (0.09) | 31 (0.21) | < 5 | 7 (0.17) | 174 (1.99) | 25 (2.97) |
| | | IS | < 5 | < 5 | < 5 | < 5 | 78 (0.89) | 11 (1.31) |
| | | TIA | < 5 | 13 (0.09) | < 5 | < 5 | 50 (0.57) | < 5 |
| | | MI | 11 (0.07) | 16 (0.11) | < 5 | < 5 | 58 (0.66) | 10 (1.19) |
| | | HF | 21 (0.14) | 23 (0.16) | < 5 | < 5 | 194 (2.22) | 25 (2.97) |
| | | HS | 5 (0.03) | < 5 | < 5 | < 5 | 13 (0.15) | < 5 |
| | | MP | 9 (0.06) | 7 (0.05) | < 5 | < 5 | 14 (0.16) | < 5 |
| | 91 to 180 days | VTE | 29 (0.19) | 26 (0.18) | 5 (0.12) | < 5 | 103 (1.18) | 14 (1.66) |
| | | DVT | 15 (0.10) | 15 (0.10) | < 5 | < 5 | 60 (0.69) | 7 (0.83) |
| | | PE | 16 (0.11) | 14 (0.10) | < 5 | < 5 | 48 (0.55) | 8 (0.95) |
| | | ATE | 8 (0.05) | 26 (0.18) | < 5 | < 5 | 189 (2.16) | 30 (3.56) |
| | | IS | < 5 | < 5 | < 5 | < 5 | 88 (1.01) | 14 (1.66) |
| | | TIA | < 5 | 8 (0.05) | < 5 | < 5 | 33 (0.38) | 10 (1.19) |
| | | MI | < 5 | 17 (0.12) | < 5 | < 5 | 71 (0.81) | 8 (0.95) |
| | | HF | 11 (0.07) | 12 (0.08) | < 5 | < 5 | 181 (2.07) | 12 (1.43) |
| | | HS | < 5 | < 5 | < 5 | < 5 | 16 (0.18) | < 5 |
| | | MP | < 5 | < 5 | < 5 | < 5 | 12 (0.14) | < 5 |
| | 181 to 365 days | VTE | < 5 | 11 (0.07) | < 5 | < 5 | 45 (0.52) | 7 (0.83) |
| | | DVT | < 5 | 5 (0.03) | < 5 | < 5 | 27 (0.31) | < 5 |
| | | PE | < 5 | 7 (0.05) | < 5 | < 5 | 22 (0.25) | 5 (0.59) |
| | | ATE | < 5 | < 5 | < 5 | < 5 | 79 (0.90) | 15 (1.78) |
| | | IS | < 5 | < 5 | < 5 | < 5 | 37 (0.42) | 6 (0.71) |
| | | TIA | < 5 | < 5 | < 5 | < 5 | 15 (0.17) | < 5 |
| | | MI | < 5 | < 5 | < 5 | < 5 | 28 (0.32) | 8 (0.95) |
| | | HF | < 5 | < 5 | < 5 | < 5 | 78 (0.89) | 6 (0.71) |
| | | HS | < 5 | < 5 | < 5 | < 5 | 5 (0.06) | < 5 |
| | | MP | < 5 | < 5 | < 5 | < 5 | < 5 | < 5 |
| Cohort 4 | | | N = 2,066,318 | N = 542,670 | N = 485,154 | N = 147,744 | | |
| | 0 to 30 days | VTE | 349 (1.69) | 27 (0.50) | 38 (0.78) | 8 (0.54) | | |
| | | DVT | 56 (0.27) | 7 (0.13) | 6 (0.12) | < 5 | | |
| | | PE | 305 (1.48) | 21 (0.39) | 33 (0.68) | 6 (0.41) | | |
| | | ATE | 26 (0.13) | 5 (0.09) | < 5 | < 5 | | |
| | | IS | < 5 | < 5 | < 5 | < 5 | | |
| | | TIA | 5 (0.02) | < 5 | < 5 | < 5 | | |
| | | MI | 17 (0.08) | < 5 | < 5 | < 5 | | |
| | | HF | 28 (0.14) | < 5 | < 5 | < 5 | | |
| | | HS | < 5 | < 5 | < 5 | < 5 | | |
| | | MP | 15 (0.07) | < 5 | < 5 | < 5 | | |
| | 31 to 90 days | VTE | 61 (0.30) | 14 (0.26) | 5 (0.10) | < 5 | | |
| | | DVT | 24 (0.12) | 8 (0.15) | < 5 | < 5 | | |
| | | PE | 40 (0.19) | 6 (0.11) | < 5 | < 5 | | |
| | | ATE | 13 (0.06) | < 5 | < 5 | < 5 | | |
| | | IS | < 5 | < 5 | < 5 | < 5 | | |
| | | TIA | < 5 | < 5 | < 5 | < 5 | | |
| | | MI | 9 (0.04) | < 5 | < 5 | < 5 | | |
| | | HF | 14 (0.07) | < 5 | < 5 | < 5 | | |
| | | HS | 5 (0.02) | < 5 | < 5 | < 5 | | |

| Cohort | Time window | Outcome | AURUM | | GOLD | | SIDIAP | |
|---|---|---|---|---|---|---|---|---|
| | | | Unvaccinated | Vaccinated | Unvaccinated | Vaccinated | Unvaccinated | Vaccinated |
| | | **MP** | 8 (0.04) | < 5 | < 5 | < 5 | | |
| | **91 to 180 days** | **VTE** | 29 (0.14) | < 5 | < 5 | < 5 | | |
| | | **DVT** | 10 (0.05) | < 5 | < 5 | < 5 | | |
| | | **PE** | 20 (0.10) | < 5 | < 5 | < 5 | | |
| | | **ATE** | < 5 | < 5 | < 5 | < 5 | | |
| | | **IS** | < 5 | < 5 | < 5 | < 5 | | |
| | | **TIA** | < 5 | < 5 | < 5 | < 5 | | |
| | | **MI** | < 5 | < 5 | < 5 | < 5 | | |
| | | **HF** | 10 (0.05) | < 5 | < 5 | < 5 | | |
| | | **HS** | < 5 | < 5 | < 5 | < 5 | | |
| | | **MP** | < 5 | < 5 | < 5 | < 5 | | |
| | **181 to 365 days** | **VTE** | < 5 | < 5 | < 5 | < 5 | | |
| | | **DVT** | < 5 | < 5 | < 5 | < 5 | | |
| | | **PE** | < 5 | < 5 | < 5 | < 5 | | |
| | | **ATE** | < 5 | < 5 | < 5 | < 5 | | |
| | | **IS** | < 5 | < 5 | < 5 | < 5 | | |
| | | **TIA** | < 5 | < 5 | < 5 | < 5 | | |
| | | **MI** | < 5 | < 5 | < 5 | < 5 | | |
| | | **HF** | < 5 | < 5 | < 5 | < 5 | | |
| | | **HS** | < 5 | < 5 | < 5 | < 5 | | |
| | | **MP** | < 5 | < 5 | < 5 | < 5 | | |

VTE = Venous thromboembolism (DVT + PE), DVT = Deep vein thrombosis, PE = Pulmonary embolism, ATE = Arterial thromboembolism (IS + TIA + MI), IS = Ischemic stroke, TIA = Transient ischemic attack, MI = Myocardial infarction, HF = Heart failure, HS = Hemorrhagic stroke, MP = Myocarditis or Pericarditis

**Table S30: Number of records (and risk per 10,000 individuals) for post COVID-19 cardiac and thromboembolic complications,** across cohorts and databases, stratified by exposure status (ChAdOx1 vaccine). Follow-up ends at first vaccine dose after index date.

| Cohort | Time window | Outcome | AURUM | | GOLD | | SIDIAP | |
|---|---|---|---|---|---|---|---|---|
| | | | Unvaccinated | Vaccinated | Unvaccinated | Vaccinated | Unvaccinated | Vaccinated |
| Cohort 1 | | | N = 344,687 | N = 219,804 | N = 168,972 | N = 82,406 | | |
| | 0 to 30 days | VTE | 77 (2.23) | 14 (0.64) | 7 (0.41) | < 5 | | |
| | | DVT | 16 (0.46) | < 5 | < 5 | < 5 | | |
| | | PE | 64 (1.86) | 13 (0.59) | 6 (0.36) | < 5 | | |
| | | ATE | 18 (0.52) | 10 (0.45) | 7 (0.41) | < 5 | | |
| | | IS | 6 (0.17) | < 5 | < 5 | < 5 | | |
| | | TIA | < 5 | < 5 | < 5 | < 5 | | |
| | | MI | 9 (0.26) | 6 (0.27) | 5 (0.30) | < 5 | | |
| | | HF | 42 (1.22) | 28 (1.27) | 9 (0.53) | < 5 | | |
| | | HS | < 5 | < 5 | < 5 | < 5 | | |
| | | MP | < 5 | < 5 | < 5 | < 5 | | |
| | 31 to 90 days | VTE | 12 (0.35) | 5 (0.23) | < 5 | < 5 | | |
| | | DVT | 5 (0.15) | < 5 | < 5 | < 5 | | |
| | | PE | 7 (0.20) | < 5 | < 5 | < 5 | | |
| | | ATE | 6 (0.17) | 8 (0.36) | < 5 | < 5 | | |
| | | IS | < 5 | < 5 | < 5 | < 5 | | |
| | | TIA | < 5 | < 5 | < 5 | < 5 | | |
| | | MI | 5 (0.15) | < 5 | < 5 | < 5 | | |
| | | HF | 26 (0.75) | 20 (0.91) | < 5 | < 5 | | |
| | | HS | < 5 | < 5 | < 5 | < 5 | | |
| | | MP | < 5 | < 5 | < 5 | < 5 | | |
| | 91 to 180 days | VTE | 6 (0.17) | 5 (0.23) | < 5 | < 5 | | |
| | | DVT | < 5 | < 5 | < 5 | < 5 | | |
| | | PE | < 5 | < 5 | < 5 | < 5 | | |
| | | ATE | 10 (0.29) | < 5 | < 5 | < 5 | | |
| | | IS | < 5 | < 5 | < 5 | < 5 | | |
| | | TIA | 5 (0.15) | < 5 | < 5 | < 5 | | |
| | | MI | < 5 | < 5 | < 5 | < 5 | | |
| | | HF | 31 (0.90) | 19 (0.86) | < 5 | < 5 | | |
| | | HS | < 5 | < 5 | < 5 | < 5 | | |
| | | MP | < 5 | < 5 | < 5 | < 5 | | |
| | 181 to 365 days | VTE | 5 (0.15) | < 5 | < 5 | < 5 | | |
| | | DVT | < 5 | < 5 | < 5 | < 5 | | |
| | | PE | < 5 | < 5 | < 5 | < 5 | | |
| | | ATE | 9 (0.26) | 8 (0.36) | < 5 | < 5 | | |
| | | IS | < 5 | < 5 | < 5 | < 5 | | |
| | | TIA | < 5 | < 5 | < 5 | < 5 | | |
| | | MI | < 5 | < 5 | < 5 | < 5 | | |
| | | HF | 35 (1.02) | 18 (0.82) | < 5 | 5 (0.61) | | |
| | | HS | < 5 | < 5 | < 5 | < 5 | | |
| | | MP | < 5 | < 5 | < 5 | < 5 | | |
| Cohort 2 | | | N = 1,975,770 | N = 969,262 | N = 582,791 | N = 302,999 | N = 433,636 | N = 323,204 |
| | 0 to 30 days | VTE | 243 (1.23) | 40 (0.41) | 33 (0.57) | < 5 | 262 (6.04) | 9 (0.28) |
| | | DVT | 42 (0.21) | 13 (0.13) | 8 (0.14) | < 5 | 68 (1.57) | 5 (0.15) |
| | | PE | 206 (1.04) | 30 (0.31) | 26 (0.45) | < 5 | 213 (4.91) | 6 (0.19) |

| Cohort | Time window | Outcome | AURUM | | GOLD | | SIDIAP | |
|---|---|---|---|---|---|---|---|---|
| | | | Unvaccinated | Vaccinated | Unvaccinated | Vaccinated | Unvaccinated | Vaccinated |
| | | **ATE** | 38 (0.19) | 11 (0.11) | < 5 | < 5 | 168 (3.87) | 6 (0.19) |
| | | **IS** | 6 (0.03) | < 5 | < 5 | < 5 | 91 (2.10) | < 5 |
| | | **TIA** | 7 (0.04) | < 5 | < 5 | < 5 | 16 (0.37) | < 5 |
| | | **MI** | 25 (0.13) | 9 (0.09) | < 5 | < 5 | 65 (1.50) | < 5 |
| | | **HF** | 46 (0.23) | 32 (0.33) | 5 (0.09) | < 5 | 383 (8.83) | 12 (0.37) |
| | | **HS** | 5 (0.03) | < 5 | < 5 | < 5 | 17 (0.39) | < 5 |
| | | **MP** | 11 (0.06) | < 5 | < 5 | < 5 | 13 (0.30) | < 5 |
| | **31 to 90 days** | **VTE** | 49 (0.25) | 8 (0.08) | < 5 | < 5 | 59 (1.36) | < 5 |
| | | **DVT** | 25 (0.13) | 5 (0.05) | < 5 | < 5 | 31 (0.71) | < 5 |
| | | **PE** | 27 (0.14) | < 5 | < 5 | < 5 | 34 (0.78) | < 5 |
| | | **ATE** | 16 (0.08) | 9 (0.09) | < 5 | < 5 | 86 (1.98) | 6 (0.19) |
| | | **IS** | < 5 | < 5 | < 5 | < 5 | 43 (0.99) | < 5 |
| | | **TIA** | < 5 | < 5 | < 5 | < 5 | 23 (0.53) | < 5 |
| | | **MI** | 11 (0.06) | 6 (0.06) | < 5 | < 5 | 27 (0.62) | < 5 |
| | | **HF** | 24 (0.12) | 13 (0.13) | < 5 | < 5 | 135 (3.11) | 7 (0.22) |
| | | **HS** | < 5 | < 5 | < 5 | < 5 | 7 (0.16) | < 5 |
| | | **MP** | 6 (0.03) | < 5 | < 5 | < 5 | < 5 | < 5 |
| | **91 to 180 days** | **VTE** | 25 (0.13) | 12 (0.12) | < 5 | < 5 | 65 (1.50) | < 5 |
| | | **DVT** | 11 (0.06) | 7 (0.07) | < 5 | < 5 | 35 (0.81) | < 5 |
| | | **PE** | 14 (0.07) | 5 (0.05) | < 5 | < 5 | 34 (0.78) | < 5 |
| | | **ATE** | 12 (0.06) | 10 (0.10) | < 5 | < 5 | 91 (2.10) | 6 (0.19) |
| | | **IS** | < 5 | < 5 | < 5 | < 5 | 49 (1.13) | < 5 |
| | | **TIA** | 7 (0.04) | < 5 | < 5 | < 5 | 19 (0.44) | < 5 |
| | | **MI** | 6 (0.03) | 6 (0.06) | < 5 | < 5 | 26 (0.60) | < 5 |
| | | **HF** | 16 (0.08) | 18 (0.19) | < 5 | < 5 | 118 (2.72) | < 5 |
| | | **HS** | < 5 | < 5 | < 5 | < 5 | 7 (0.16) | < 5 |
| | | **MP** | < 5 | < 5 | < 5 | < 5 | 7 (0.16) | < 5 |
| | **181 to 365 days** | **VTE** | 14 (0.07) | 5 (0.05) | < 5 | < 5 | 15 (0.35) | 9 (0.28) |
| | | **DVT** | < 5 | < 5 | < 5 | < 5 | 9 (0.21) | 7 (0.22) |
| | | **PE** | 10 (0.05) | < 5 | < 5 | < 5 | 7 (0.16) | < 5 |
| | | **ATE** | 11 (0.06) | 10 (0.10) | < 5 | < 5 | 66 (1.52) | 10 (0.31) |
| | | **IS** | < 5 | < 5 | < 5 | < 5 | 33 (0.76) | < 5 |
| | | **TIA** | < 5 | < 5 | < 5 | < 5 | 20 (0.46) | < 5 |
| | | **MI** | 8 (0.04) | 6 (0.06) | < 5 | < 5 | 15 (0.35) | 5 (0.15) |
| | | **HF** | 18 (0.09) | 20 (0.21) | < 5 | < 5 | 76 (1.75) | 9 (0.28) |
| | | **HS** | < 5 | < 5 | < 5 | < 5 | < 5 | < 5 |
| | | **MP** | < 5 | < 5 | < 5 | < 5 | < 5 | < 5 |
| **Cohort 3** | | | **N = 1,510,493** | **N = 1,473,602** | **N = 418,184** | **N = 423,876** | **N = 873,400** | **N = 84,204** |
| | **0 to 30 days** | **VTE** | 244 (1.62) | 23 (0.16) | 28 (0.67) | < 5 | 334 (3.82) | 5 (0.59) |
| | | **DVT** | 44 (0.29) | < 5 | 6 (0.14) | < 5 | 96 (1.10) | < 5 |
| | | **PE** | 209 (1.38) | 19 (0.13) | 23 (0.55) | < 5 | 267 (3.06) | < 5 |
| | | **ATE** | 28 (0.19) | 8 (0.05) | < 5 | < 5 | 217 (2.48) | < 5 |
| | | **IS** | < 5 | < 5 | < 5 | < 5 | 102 (1.17) | < 5 |
| | | **TIA** | 5 (0.03) | < 5 | < 5 | < 5 | 21 (0.24) | < 5 |
| | | **MI** | 19 (0.13) | 7 (0.05) | < 5 | < 5 | 100 (1.14) | < 5 |
| | | **HF** | 30 (0.20) | 11 (0.07) | < 5 | < 5 | 374 (4.28) | 5 (0.59) |
| | | **HS** | < 5 | < 5 | < 5 | < 5 | 20 (0.23) | < 5 |
| | | **MP** | 9 (0.06) | < 5 | < 5 | < 5 | 19 (0.22) | < 5 |
| | **31 to 90 days** | **VTE** | 46 (0.30) | < 5 | < 5 | < 5 | 86 (0.98) | < 5 |

| Cohort | Time window | Outcome | AURUM | | GOLD | | SIDIAP | |
|---|---|---|---|---|---|---|---|---|
| | | | Unvaccinated | Vaccinated | Unvaccinated | Vaccinated | Unvaccinated | Vaccinated |
| | | DVT | 21 (0.14) | < 5 | < 5 | < 5 | 57 (0.65) | < 5 |
| | | PE | 28 (0.19) | < 5 | < 5 | < 5 | 39 (0.45) | < 5 |
| | | ATE | 11 (0.07) | 6 (0.04) | < 5 | < 5 | 110 (1.26) | < 5 |
| | | IS | < 5 | < 5 | < 5 | < 5 | 55 (0.63) | < 5 |
| | | TIA | < 5 | < 5 | < 5 | < 5 | 27 (0.31) | < 5 |
| | | MI | 9 (0.06) | < 5 | < 5 | < 5 | 36 (0.41) | < 5 |
| | | HF | 14 (0.09) | 5 (0.03) | < 5 | < 5 | 142 (1.63) | < 5 |
| | | HS | < 5 | < 5 | < 5 | < 5 | 7 (0.08) | < 5 |
| | | MP | 8 (0.05) | < 5 | < 5 | < 5 | 9 (0.10) | < 5 |
| | 91 to 180 days | VTE | 24 (0.16) | 8 (0.05) | < 5 | < 5 | 65 (0.74) | < 5 |
| | | DVT | 11 (0.07) | 6 (0.04) | < 5 | < 5 | 37 (0.42) | < 5 |
| | | PE | 14 (0.09) | < 5 | < 5 | < 5 | 32 (0.37) | < 5 |
| | | ATE | < 5 | 7 (0.05) | < 5 | < 5 | 116 (1.33) | 6 (0.71) |
| | | IS | < 5 | < 5 | < 5 | < 5 | 57 (0.65) | 5 (0.59) |
| | | TIA | < 5 | < 5 | < 5 | < 5 | 22 (0.25) | < 5 |
| | | MI | < 5 | < 5 | < 5 | < 5 | 40 (0.46) | < 5 |
| | | HF | 10 (0.07) | < 5 | < 5 | < 5 | 122 (1.40) | < 5 |
| | | HS | < 5 | < 5 | < 5 | < 5 | 9 (0.10) | < 5 |
| | | MP | < 5 | < 5 | < 5 | < 5 | 8 (0.09) | < 5 |
| | 181 to 365 days | VTE | < 5 | 9 (0.06) | < 5 | < 5 | 37 (0.42) | < 5 |
| | | DVT | < 5 | < 5 | < 5 | < 5 | 23 (0.26) | < 5 |
| | | PE | < 5 | 6 (0.04) | < 5 | < 5 | 16 (0.18) | < 5 |
| | | ATE | < 5 | < 5 | < 5 | < 5 | 60 (0.69) | 8 (0.95) |
| | | IS | < 5 | < 5 | < 5 | < 5 | 26 (0.30) | < 5 |
| | | TIA | < 5 | < 5 | < 5 | < 5 | 14 (0.16) | < 5 |
| | | MI | < 5 | < 5 | < 5 | < 5 | 21 (0.24) | < 5 |
| | | HF | < 5 | < 5 | < 5 | < 5 | 65 (0.74) | 5 (0.59) |
| | | HS | < 5 | < 5 | < 5 | < 5 | < 5 | < 5 |
| | | MP | < 5 | < 5 | < 5 | < 5 | < 5 | < 5 |
| **Cohort 4** | | | **N = 2,066,318** | **N = 542,670** | **N = 485,154** | **N = 147,744** | | |
| | 0 to 30 days | VTE | 346 (1.67) | 8 (0.15) | 38 (0.78) | 6 (0.41) | | |
| | | DVT | 56 (0.27) | < 5 | 6 (0.12) | < 5 | | |
| | | PE | 302 (1.46) | 7 (0.13) | 33 (0.68) | 6 (0.41) | | |
| | | ATE | 26 (0.13) | < 5 | < 5 | < 5 | | |
| | | IS | < 5 | < 5 | < 5 | < 5 | | |
| | | TIA | 5 (0.02) | < 5 | < 5 | < 5 | | |
| | | MI | 17 (0.08) | < 5 | < 5 | < 5 | | |
| | | HF | 28 (0.14) | < 5 | < 5 | < 5 | | |
| | | HS | < 5 | < 5 | < 5 | < 5 | | |
| | | MP | 14 (0.07) | < 5 | < 5 | < 5 | | |
| | 31 to 90 days | VTE | 60 (0.29) | 8 (0.15) | < 5 | < 5 | | |
| | | DVT | 23 (0.11) | < 5 | < 5 | < 5 | | |
| | | PE | 40 (0.19) | 5 (0.09) | < 5 | < 5 | | |
| | | ATE | 13 (0.06) | < 5 | < 5 | < 5 | | |
| | | IS | < 5 | < 5 | < 5 | < 5 | | |
| | | TIA | < 5 | < 5 | < 5 | < 5 | | |
| | | MI | 9 (0.04) | < 5 | < 5 | < 5 | | |
| | | HF | 14 (0.07) | < 5 | < 5 | < 5 | | |
| | | HS | < 5 | < 5 | < 5 | < 5 | | |

| Cohort | Time window | Outcome | AURUM | | GOLD | | SIDIAP | |
|---|---|---|---|---|---|---|---|---|
| | | | Unvaccinated | Vaccinated | Unvaccinated | Vaccinated | Unvaccinated | Vaccinated |
| | | **MP** | 7 (0.03) | < 5 | < 5 | < 5 | | |
| | **91 to 180 days** | **VTE** | 29 (0.14) | < 5 | < 5 | < 5 | | |
| | | **DVT** | 10 (0.05) | < 5 | < 5 | < 5 | | |
| | | **PE** | 20 (0.10) | < 5 | < 5 | < 5 | | |
| | | **ATE** | < 5 | < 5 | < 5 | < 5 | | |
| | | **IS** | < 5 | < 5 | < 5 | < 5 | | |
| | | **TIA** | < 5 | < 5 | < 5 | < 5 | | |
| | | **MI** | < 5 | < 5 | < 5 | < 5 | | |
| | | **HF** | 10 (0.05) | < 5 | < 5 | < 5 | | |
| | | **HS** | < 5 | < 5 | < 5 | < 5 | | |
| | | **MP** | < 5 | < 5 | < 5 | < 5 | | |
| | **181 to 365 days** | **VTE** | < 5 | < 5 | < 5 | < 5 | | |
| | | **DVT** | < 5 | < 5 | < 5 | < 5 | | |
| | | **PE** | < 5 | < 5 | < 5 | < 5 | | |
| | | **ATE** | < 5 | < 5 | < 5 | < 5 | | |
| | | **IS** | < 5 | < 5 | < 5 | < 5 | | |
| | | **TIA** | < 5 | < 5 | < 5 | < 5 | | |
| | | **MI** | < 5 | < 5 | < 5 | < 5 | | |
| | | **HF** | < 5 | < 5 | < 5 | < 5 | | |
| | | **HS** | < 5 | < 5 | < 5 | < 5 | | |
| | | **MP** | < 5 | < 5 | < 5 | < 5 | | |

VTE = Venous thromboembolism (DVT + PE), DVT = Deep vein thrombosis, PE = Pulmonary embolism, ATE = Arterial thromboembolism (IS + TIA + MI), IS = Ischemic stroke, TIA = Transient ischemic attack, MI = Myocardial infarction, HF = Heart failure, HS = Hemorrhagic stroke, MP = Myocarditis or Pericarditis

**Table S31: Number of records (and risk per 10,000 individuals) for post COVID-19 cardiac and thromboembolic complications,** across cohorts and databases, stratified by exposure status (ChAdOx1 vaccine). Only first outcome after COVID-19 captured.

| Cohort | Time window | Outcome | AURUM | | GOLD | | SIDIAP | |
|---|---|---|---|---|---|---|---|---|
| | | | Unvaccinated | Vaccinated | Unvaccinated | Vaccinated | Unvaccinated | Vaccinated |
| Cohort 1 | | | N = 344,687 | N = 219,804 | N = 168,972 | N = 82,406 | | |
| | 0 to 30 days | VTE | 138 (4.00) | 41 (1.87) | 14 (0.83) | < 5 | | |
| | | DVT | 34 (0.99) | 12 (0.55) | < 5 | < 5 | | |
| | | PE | 112 (3.25) | 31 (1.41) | 12 (0.71) | < 5 | | |
| | | ATE | 53 (1.54) | 25 (1.14) | 9 (0.53) | 6 (0.73) | | |
| | | IS | 13 (0.38) | < 5 | < 5 | < 5 | | |
| | | TIA | 12 (0.35) | 7 (0.32) | < 5 | < 5 | | |
| | | MI | 29 (0.84) | 17 (0.77) | 7 (0.41) | < 5 | | |
| | | HF | 119 (3.45) | 72 (3.28) | 20 (1.18) | 5 (0.61) | | |
| | | HS | < 5 | < 5 | < 5 | < 5 | | |
| | | MP | < 5 | < 5 | < 5 | < 5 | | |
| | 31 to 90 days | VTE | 25 (0.73) | 14 (0.64) | < 5 | < 5 | | |
| | | DVT | 9 (0.26) | 8 (0.36) | < 5 | < 5 | | |
| | | PE | 16 (0.46) | 7 (0.32) | < 5 | < 5 | | |
| | | ATE | 16 (0.46) | 20 (0.91) | < 5 | < 5 | | |
| | | IS | < 5 | < 5 | < 5 | < 5 | | |
| | | TIA | 6 (0.17) | 9 (0.41) | < 5 | < 5 | | |
| | | MI | 11 (0.32) | 9 (0.41) | < 5 | < 5 | | |
| | | HF | 69 (2.00) | 39 (1.77) | 6 (0.36) | 5 (0.61) | | |
| | | HS | < 5 | < 5 | < 5 | < 5 | | |
| | | MP | < 5 | < 5 | < 5 | < 5 | | |
| | 91 to 180 days | VTE | 13 (0.38) | 15 (0.68) | 6 (0.36) | < 5 | | |
| | | DVT | 5 (0.15) | 5 (0.23) | < 5 | < 5 | | |
| | | PE | 8 (0.23) | 10 (0.45) | < 5 | < 5 | | |
| | | ATE | 22 (0.64) | 10 (0.45) | < 5 | 6 (0.73) | | |
| | | IS | < 5 | < 5 | < 5 | < 5 | | |
| | | TIA | 10 (0.29) | 6 (0.27) | < 5 | < 5 | | |
| | | MI | 11 (0.32) | < 5 | < 5 | 5 (0.61) | | |
| | | HF | 50 (1.45) | 35 (1.59) | < 5 | < 5 | | |
| | | HS | < 5 | < 5 | < 5 | < 5 | | |
| | | MP | < 5 | < 5 | < 5 | < 5 | | |
| | 181 to 365 days | VTE | 5 (0.15) | < 5 | < 5 | < 5 | | |
| | | DVT | < 5 | < 5 | < 5 | < 5 | | |
| | | PE | < 5 | < 5 | < 5 | < 5 | | |
| | | ATE | 13 (0.38) | 9 (0.41) | < 5 | < 5 | | |
| | | IS | < 5 | < 5 | < 5 | < 5 | | |
| | | TIA | 6 (0.17) | < 5 | < 5 | < 5 | | |
| | | MI | < 5 | < 5 | < 5 | < 5 | | |
| | | HF | 45 (1.31) | 18 (0.82) | < 5 | 5 (0.61) | | |
| | | HS | < 5 | < 5 | < 5 | < 5 | | |
| | | MP | < 5 | < 5 | < 5 | < 5 | | |
| Cohort 2 | | | N = 1,975,770 | N = 969,262 | N = 582,791 | N = 302,999 | N = 433,636 | N = 323,204 |
| | 0 to 30 days | VTE | 347 (1.76) | 158 (1.63) | 44 (0.75) | 19 (0.63) | 403 (9.29) | 90 (2.78) |
| | | DVT | 75 (0.38) | 47 (0.48) | 9 (0.15) | < 5 | 122 (2.81) | 45 (1.39) |
| | | PE | 280 (1.42) | 120 (1.24) | 36 (0.62) | 15 (0.50) | 311 (7.17) | 55 (1.70) |

| Cohort | Time window | Outcome | AURUM | | GOLD | | SIDIAP | |
|---|---|---|---|---|---|---|---|---|
| | | | Unvaccinated | Vaccinated | Unvaccinated | Vaccinated | Unvaccinated | Vaccinated |
| | | ATE | 70 (0.35) | 75 (0.77) | 9 (0.15) | < 5 | 341 (7.86) | 131 (4.05) |
| | | IS | 10 (0.05) | 12 (0.12) | < 5 | < 5 | 186 (4.29) | 52 (1.61) |
| | | TIA | 15 (0.08) | 13 (0.13) | < 5 | < 5 | 45 (1.04) | 23 (0.71) |
| | | MI | 45 (0.23) | 52 (0.54) | 7 (0.12) | < 5 | 127 (2.93) | 62 (1.92) |
| | | HF | 74 (0.37) | 103 (1.06) | 5 (0.09) | 11 (0.36) | 642 (14.81) | 123 (3.81) |
| | | HS | 5 (0.03) | < 5 | < 5 | < 5 | 38 (0.88) | 7 (0.22) |
| | | MP | 20 (0.10) | 6 (0.06) | < 5 | < 5 | 29 (0.67) | 10 (0.31) |
| | 31 to 90 days | VTE | 90 (0.46) | 54 (0.56) | 6 (0.10) | 9 (0.30) | 103 (2.38) | 34 (1.05) |
| | | DVT | 49 (0.25) | 25 (0.26) | < 5 | 5 (0.17) | 58 (1.34) | 23 (0.71) |
| | | PE | 44 (0.22) | 31 (0.32) | < 5 | < 5 | 54 (1.25) | 16 (0.50) |
| | | ATE | 37 (0.19) | 56 (0.58) | 8 (0.14) | 5 (0.17) | 199 (4.59) | 106 (3.28) |
| | | IS | 6 (0.03) | 6 (0.06) | < 5 | < 5 | 95 (2.19) | 57 (1.76) |
| | | TIA | 11 (0.06) | 24 (0.25) | < 5 | < 5 | 54 (1.25) | 24 (0.74) |
| | | MI | 21 (0.11) | 28 (0.29) | < 5 | < 5 | 63 (1.45) | 27 (0.84) |
| | | HF | 47 (0.24) | 65 (0.67) | 5 (0.09) | 7 (0.23) | 260 (6.00) | 61 (1.89) |
| | | HS | 6 (0.03) | < 5 | < 5 | < 5 | 18 (0.42) | 7 (0.22) |
| | | MP | 14 (0.07) | < 5 | < 5 | < 5 | 8 (0.18) | < 5 |
| | 91 to 180 days | VTE | 35 (0.18) | 28 (0.29) | 6 (0.10) | < 5 | 104 (2.40) | 21 (0.65) |
| | | DVT | 19 (0.10) | 15 (0.15) | < 5 | < 5 | 57 (1.31) | 16 (0.50) |
| | | PE | 16 (0.08) | 15 (0.15) | 6 (0.10) | < 5 | 55 (1.27) | 6 (0.19) |
| | | ATE | 27 (0.14) | 28 (0.29) | 5 (0.09) | < 5 | 173 (3.99) | 106 (3.28) |
| | | IS | < 5 | < 5 | < 5 | < 5 | 88 (2.03) | 40 (1.24) |
| | | TIA | 12 (0.06) | 13 (0.13) | < 5 | < 5 | 40 (0.92) | 32 (0.99) |
| | | MI | 14 (0.07) | 15 (0.15) | < 5 | < 5 | 51 (1.18) | 37 (1.14) |
| | | HF | 25 (0.13) | 41 (0.42) | < 5 | < 5 | 215 (4.96) | 54 (1.67) |
| | | HS | < 5 | < 5 | < 5 | < 5 | 17 (0.39) | < 5 |
| | | MP | < 5 | < 5 | < 5 | < 5 | 12 (0.28) | < 5 |
| | 181 to 365 days | VTE | 22 (0.11) | < 5 | < 5 | < 5 | 29 (0.67) | 14 (0.43) |
| | | DVT | 8 (0.04) | < 5 | < 5 | < 5 | 16 (0.37) | 12 (0.37) |
| | | PE | 15 (0.08) | < 5 | < 5 | < 5 | 15 (0.35) | < 5 |
| | | ATE | 12 (0.06) | 10 (0.10) | < 5 | < 5 | 96 (2.21) | 40 (1.24) |
| | | IS | < 5 | < 5 | < 5 | < 5 | 50 (1.15) | 19 (0.59) |
| | | TIA | < 5 | < 5 | < 5 | < 5 | 22 (0.51) | 12 (0.37) |
| | | MI | 8 (0.04) | 6 (0.06) | < 5 | < 5 | 29 (0.67) | 13 (0.40) |
| | | HF | 17 (0.09) | 22 (0.23) | < 5 | < 5 | 93 (2.14) | 25 (0.77) |
| | | HS | < 5 | < 5 | < 5 | < 5 | 6 (0.14) | < 5 |
| | | MP | < 5 | < 5 | < 5 | < 5 | < 5 | < 5 |
| Cohort 3 | | | N = 1,510,493 | N = 1,473,602 | N = 418,184 | N = 423,876 | N = 873,400 | N = 84,204 |
| | 0 to 30 days | VTE | 267 (1.77) | 139 (0.94) | 33 (0.79) | 16 (0.38) | 431 (4.93) | 27 (3.21) |
| | | DVT | 52 (0.34) | 41 (0.28) | 7 (0.17) | < 5 | 137 (1.57) | 9 (1.07) |
| | | PE | 224 (1.48) | 101 (0.69) | 27 (0.65) | 12 (0.28) | 331 (3.79) | 22 (2.61) |
| | | ATE | 33 (0.22) | 47 (0.32) | < 5 | 12 (0.28) | 319 (3.65) | 32 (3.80) |
| | | IS | 5 (0.03) | 5 (0.03) | < 5 | < 5 | 154 (1.76) | 13 (1.54) |
| | | TIA | 6 (0.04) | 16 (0.11) | < 5 | < 5 | 33 (0.38) | 5 (0.59) |
| | | MI | 22 (0.15) | 26 (0.18) | < 5 | 9 (0.21) | 141 (1.61) | 16 (1.90) |
| | | HF | 32 (0.21) | 28 (0.19) | < 5 | < 5 | 486 (5.56) | 51 (6.06) |
| | | HS | < 5 | < 5 | < 5 | < 5 | 34 (0.39) | < 5 |
| | | MP | 14 (0.09) | 6 (0.04) | < 5 | < 5 | 27 (0.31) | 6 (0.71) |
| | 31 to 90 days | VTE | 60 (0.40) | 44 (0.30) | < 5 | 6 (0.14) | 115 (1.32) | 9 (1.07) |

| Cohort | Time window | Outcome | AURUM | | GOLD | | SIDIAP | |
|---|---|---|---|---|---|---|---|---|
| | | | Unvaccinated | Vaccinated | Unvaccinated | Vaccinated | Unvaccinated | Vaccinated |
| | | **DVT** | 29 (0.19) | 26 (0.18) | < 5 | < 5 | 79 (0.90) | < 5 |
| | | **PE** | 34 (0.23) | 18 (0.12) | < 5 | < 5 | 47 (0.54) | 6 (0.71) |
| | | **ATE** | 14 (0.09) | 31 (0.21) | < 5 | 7 (0.17) | 172 (1.97) | 25 (2.97) |
| | | **IS** | < 5 | < 5 | < 5 | < 5 | 77 (0.88) | 11 (1.31) |
| | | **TIA** | < 5 | 13 (0.09) | < 5 | < 5 | 50 (0.57) | < 5 |
| | | **MI** | 11 (0.07) | 16 (0.11) | < 5 | < 5 | 57 (0.65) | 10 (1.19) |
| | | **HF** | 21 (0.14) | 23 (0.16) | < 5 | < 5 | 187 (2.14) | 25 (2.97) |
| | | **HS** | 5 (0.03) | < 5 | < 5 | < 5 | 13 (0.15) | < 5 |
| | | **MP** | 9 (0.06) | 7 (0.05) | < 5 | < 5 | 14 (0.16) | < 5 |
| | **91 to 180 days** | **VTE** | 28 (0.19) | 25 (0.17) | 5 (0.12) | < 5 | 92 (1.05) | 13 (1.54) |
| | | **DVT** | 14 (0.09) | 15 (0.10) | < 5 | < 5 | 56 (0.64) | 7 (0.83) |
| | | **PE** | 16 (0.11) | 13 (0.09) | < 5 | < 5 | 41 (0.47) | 7 (0.83) |
| | | **ATE** | 8 (0.05) | 26 (0.18) | < 5 | < 5 | 174 (1.99) | 28 (3.33) |
| | | **IS** | < 5 | < 5 | < 5 | < 5 | 77 (0.88) | 12 (1.43) |
| | | **TIA** | < 5 | 8 (0.05) | < 5 | < 5 | 30 (0.34) | 10 (1.19) |
| | | **MI** | < 5 | 17 (0.12) | < 5 | < 5 | 69 (0.79) | 8 (0.95) |
| | | **HF** | 11 (0.07) | 10 (0.07) | < 5 | < 5 | 169 (1.93) | 12 (1.43) |
| | | **HS** | < 5 | < 5 | < 5 | < 5 | 16 (0.18) | < 5 |
| | | **MP** | < 5 | < 5 | < 5 | < 5 | 12 (0.14) | < 5 |
| | **181 to 365 days** | **VTE** | < 5 | 9 (0.06) | < 5 | < 5 | 39 (0.45) | < 5 |
| | | **DVT** | < 5 | < 5 | < 5 | < 5 | 24 (0.27) | < 5 |
| | | **PE** | < 5 | 6 (0.04) | < 5 | < 5 | 18 (0.21) | < 5 |
| | | **ATE** | < 5 | < 5 | < 5 | < 5 | 62 (0.71) | 15 (1.78) |
| | | **IS** | < 5 | < 5 | < 5 | < 5 | 30 (0.34) | 6 (0.71) |
| | | **TIA** | < 5 | < 5 | < 5 | < 5 | 13 (0.15) | < 5 |
| | | **MI** | < 5 | < 5 | < 5 | < 5 | 20 (0.23) | 8 (0.95) |
| | | **HF** | < 5 | < 5 | < 5 | < 5 | 65 (0.74) | 6 (0.71) |
| | | **HS** | < 5 | < 5 | < 5 | < 5 | < 5 | < 5 |
| | | **MP** | < 5 | < 5 | < 5 | < 5 | < 5 | < 5 |
| **Cohort 4** | | | **N = 2,066,318** | **N = 542,670** | **N = 485,154** | **N = 147,744** | | |
| | **0 to 30 days** | **VTE** | 349 (1.69) | 27 (0.50) | 38 (0.78) | 8 (0.54) | | |
| | | **DVT** | 56 (0.27) | 7 (0.13) | 6 (0.12) | < 5 | | |
| | | **PE** | 305 (1.48) | 21 (0.39) | 33 (0.68) | 6 (0.41) | | |
| | | **ATE** | 26 (0.13) | 5 (0.09) | < 5 | < 5 | | |
| | | **IS** | < 5 | < 5 | < 5 | < 5 | | |
| | | **TIA** | 5 (0.02) | < 5 | < 5 | < 5 | | |
| | | **MI** | 17 (0.08) | < 5 | < 5 | < 5 | | |
| | | **HF** | 28 (0.14) | < 5 | < 5 | < 5 | | |
| | | **HS** | < 5 | < 5 | < 5 | < 5 | | |
| | | **MP** | 15 (0.07) | < 5 | < 5 | < 5 | | |
| | **31 to 90 days** | **VTE** | 58 (0.28) | 14 (0.26) | 5 (0.10) | < 5 | | |
| | | **DVT** | 23 (0.11) | 8 (0.15) | < 5 | < 5 | | |
| | | **PE** | 38 (0.18) | 6 (0.11) | < 5 | < 5 | | |
| | | **ATE** | 13 (0.06) | < 5 | < 5 | < 5 | | |
| | | **IS** | < 5 | < 5 | < 5 | < 5 | | |
| | | **TIA** | < 5 | < 5 | < 5 | < 5 | | |
| | | **MI** | 9 (0.04) | < 5 | < 5 | < 5 | | |
| | | **HF** | 14 (0.07) | < 5 | < 5 | < 5 | | |
| | | **HS** | 5 (0.02) | < 5 | < 5 | < 5 | | |

| Cohort | Time window | Outcome | AURUM | | GOLD | | SIDIAP | |
|---|---|---|---|---|---|---|---|---|
| | | | Unvaccinated | Vaccinated | Unvaccinated | Vaccinated | Unvaccinated | Vaccinated |
| | | MP | 8 (0.04) | < 5 | < 5 | < 5 | | |
| | 91 to 180 days | VTE | 28 (0.14) | < 5 | < 5 | < 5 | | |
| | | DVT | 9 (0.04) | < 5 | < 5 | < 5 | | |
| | | PE | 20 (0.10) | < 5 | < 5 | < 5 | | |
| | | ATE | < 5 | < 5 | < 5 | < 5 | | |
| | | IS | < 5 | < 5 | < 5 | < 5 | | |
| | | TIA | < 5 | < 5 | < 5 | < 5 | | |
| | | MI | < 5 | < 5 | < 5 | < 5 | | |
| | | HF | 10 (0.05) | < 5 | < 5 | < 5 | | |
| | | HS | < 5 | < 5 | < 5 | < 5 | | |
| | | MP | < 5 | < 5 | < 5 | < 5 | | |
| | 181 to 365 days | VTE | < 5 | < 5 | < 5 | < 5 | | |
| | | DVT | < 5 | < 5 | < 5 | < 5 | | |
| | | PE | < 5 | < 5 | < 5 | < 5 | | |
| | | ATE | < 5 | < 5 | < 5 | < 5 | | |
| | | IS | < 5 | < 5 | < 5 | < 5 | | |
| | | TIA | < 5 | < 5 | < 5 | < 5 | | |
| | | MI | < 5 | < 5 | < 5 | < 5 | | |
| | | HF | < 5 | < 5 | < 5 | < 5 | | |
| | | HS | < 5 | < 5 | < 5 | < 5 | | |
| | | MP | < 5 | < 5 | < 5 | < 5 | | |

VTE = Venous thromboembolism (DVT + PE), DVT = Deep vein thrombosis, PE = Pulmonary embolism, ATE = Arterial thromboembolism (IS + TIA + MI), IS = Ischemic stroke, TIA = Transient ischemic attack, MI = Myocardial infarction, HF = Heart failure, HS = Hemorrhagic stroke, MP = Myocarditis or Pericarditis

**Table S32: Number of records (and risk per 10,000 individuals) for post COVID-19 cardiac and thromboembolic complications,** across cohorts and databases, stratified by exposure status (BNT162b2 vaccine).

| Cohort | Time window | Outcome | AURUM | | CORIVA | | GOLD | | SIDIAP | |
|---|---|---|---|---|---|---|---|---|---|---|
| | | | Unvaccinated | Vaccinated | Unvaccinated | Vaccinated | Unvaccinated | Vaccinated | Unvaccinated | Vaccinated |
| **Cohort 1** | | | **N = 348,052** | **N = 332,790** | **N = 24,073** | **N = 19,686** | **N = 169,459** | **N = 32,755** | **N = 223,960** | **N = 88,896** |
| | **0 to 30 days** | **VTE** | 167 (4.80) | 76 (2.28) | 97 (40.29) | 31 (15.75) | 15 (0.89) | < 5 | 251 (11.21) | 94 (10.57) |
| | | **DVT** | 39 (1.12) | 15 (0.45) | 18 (7.48) | 8 (4.06) | < 5 | < 5 | 76 (3.39) | 29 (3.26) |
| | | **PE** | 138 (3.96) | 64 (1.92) | 81 (33.65) | 24 (12.19) | 12 (0.71) | < 5 | 187 (8.35) | 75 (8.44) |
| | | **ATE** | 62 (1.78) | 45 (1.35) | 125 (51.93) | 37 (18.80) | 8 (0.47) | < 5 | 353 (15.76) | 206 (23.17) |
| | | **IS** | 16 (0.46) | 5 (0.15) | 77 (31.99) | 18 (9.14) | < 5 | < 5 | 196 (8.75) | 116 (13.05) |
| | | **TIA** | 14 (0.40) | 11 (0.33) | 23 (9.55) | 7 (3.56) | < 5 | < 5 | 58 (2.59) | 41 (4.61) |
| | | **MI** | 34 (0.98) | 29 (0.87) | 37 (15.37) | 19 (9.65) | 6 (0.35) | < 5 | 115 (5.13) | 61 (6.86) |
| | | **HF** | 136 (3.91) | 126 (3.79) | 438 (181.95) | 154 (78.23) | 20 (1.18) | < 5 | 1,203 (53.71) | 634 (71.32) |
| | | **HS** | < 5 | < 5 | < 5 | < 5 | < 5 | < 5 | 46 (2.05) | 14 (1.57) |
| | | **MP** | < 5 | < 5 | < 5 | < 5 | < 5 | < 5 | 16 (0.71) | < 5 |
| | **31 to 90 days** | **VTE** | 31 (0.89) | 26 (0.78) | 38 (15.79) | 8 (4.06) | 5 (0.30) | < 5 | 91 (4.06) | 44 (4.95) |
| | | **DVT** | 13 (0.37) | 7 (0.21) | 23 (9.55) | < 5 | < 5 | < 5 | 55 (2.46) | 29 (3.26) |
| | | **PE** | 18 (0.52) | 19 (0.57) | 17 (7.06) | < 5 | < 5 | < 5 | 42 (1.88) | 19 (2.14) |
| | | **ATE** | 20 (0.57) | 23 (0.69) | 33 (13.71) | 20 (10.16) | < 5 | < 5 | 222 (9.91) | 128 (14.40) |
| | | **IS** | < 5 | < 5 | 21 (8.72) | 10 (5.08) | < 5 | < 5 | 112 (5.00) | 71 (7.99) |
| | | **TIA** | 10 (0.29) | 10 (0.30) | 11 (4.57) | 7 (3.56) | < 5 | < 5 | 72 (3.21) | 32 (3.60) |
| | | **MI** | 11 (0.32) | 11 (0.33) | 10 (4.15) | < 5 | < 5 | < 5 | 52 (2.32) | 29 (3.26) |
| | | **HF** | 75 (2.15) | 71 (2.13) | 149 (61.90) | 67 (34.03) | 6 (0.35) | < 5 | 493 (22.01) | 296 (33.30) |
| | | **HS** | < 5 | < 5 | < 5 | < 5 | < 5 | < 5 | 16 (0.71) | 13 (1.46) |
| | | **MP** | < 5 | < 5 | < 5 | < 5 | < 5 | < 5 | < 5 | < 5 |
| | **91 to 180 days** | **VTE** | 17 (0.49) | 6 (0.18) | 25 (10.39) | 13 (6.60) | 6 (0.35) | < 5 | 70 (3.13) | 40 (4.50) |
| | | **DVT** | 6 (0.17) | < 5 | 13 (5.40) | < 5 | < 5 | < 5 | 41 (1.83) | 23 (2.59) |
| | | **PE** | 11 (0.32) | < 5 | 14 (5.82) | 10 (5.08) | < 5 | < 5 | 34 (1.52) | 20 (2.25) |
| | | **ATE** | 27 (0.78) | 18 (0.54) | 32 (13.29) | 18 (9.14) | < 5 | < 5 | 201 (8.97) | 110 (12.37) |
| | | **IS** | < 5 | 6 (0.18) | 19 (7.89) | 6 (3.05) | < 5 | < 5 | 126 (5.63) | 59 (6.64) |
| | | **TIA** | 13 (0.37) | 6 (0.18) | 8 (3.32) | < 5 | < 5 | < 5 | 45 (2.01) | 34 (3.82) |
| | | **MI** | 12 (0.34) | 6 (0.18) | 9 (3.74) | 11 (5.59) | < 5 | < 5 | 39 (1.74) | 26 (2.92) |
| | | **HF** | 68 (1.95) | 57 (1.71) | 172 (71.45) | 95 (48.26) | < 5 | < 5 | 432 (19.29) | 249 (28.01) |
| | | **HS** | < 5 | < 5 | < 5 | < 5 | < 5 | < 5 | 23 (1.03) | < 5 |
| | | **MP** | < 5 | < 5 | < 5 | < 5 | < 5 | < 5 | < 5 | < 5 |
| | **181 to 365 days** | **VTE** | 15 (0.43) | 10 (0.30) | 59 (24.51) | 21 (10.67) | < 5 | < 5 | 35 (1.56) | 12 (1.35) |
| | | **DVT** | < 5 | 6 (0.18) | 19 (7.89) | 13 (6.60) | < 5 | < 5 | 22 (0.98) | 8 (0.90) |
| | | **PE** | 11 (0.32) | < 5 | 45 (18.69) | 9 (4.57) | < 5 | < 5 | 15 (0.67) | < 5 |
| | | **ATE** | 19 (0.55) | 14 (0.42) | 57 (23.68) | 31 (15.75) | < 5 | < 5 | 124 (5.54) | 52 (5.85) |
| | | **IS** | < 5 | < 5 | 33 (13.71) | 15 (7.62) | < 5 | < 5 | 58 (2.59) | 33 (3.71) |
| | | **TIA** | 12 (0.34) | < 5 | 15 (6.23) | 13 (6.60) | < 5 | < 5 | 35 (1.56) | 13 (1.46) |
| | | **MI** | < 5 | 7 (0.21) | 15 (6.23) | 9 (4.57) | < 5 | < 5 | 35 (1.56) | 8 (0.90) |
| | | **HF** | 66 (1.90) | 38 (1.14) | 276 (114.65) | 146 (74.16) | < 5 | < 5 | 234 (10.45) | 149 (16.76) |
| | | **HS** | < 5 | < 5 | < 5 | < 5 | < 5 | < 5 | 8 (0.36) | 9 (1.01) |
| | | **MP** | < 5 | < 5 | < 5 | < 5 | < 5 | < 5 | < 5 | < 5 |
| **Cohort 2** | | | **N = 1,976,163** | **N = 594,262** | **N = 34,320** | **N = 4,067** | **N = 584,309** | **N = 180,670** | **N = 433,111** | **N = 445,581** |
| | **0 to 30 days** | **VTE** | 344 (1.74) | 62 (1.04) | 89 (25.93) | 8 (19.67) | 40 (0.68) | 5 (0.28) | 396 (9.14) | 265 (5.95) |
| | | **DVT** | 72 (0.36) | 18 (0.30) | 16 (4.66) | < 5 | 8 (0.14) | < 5 | 117 (2.70) | 96 (2.15) |
| | | **PE** | 279 (1.41) | 45 (0.76) | 74 (21.56) | 5 (12.29) | 33 (0.56) | < 5 | 309 (7.13) | 189 (4.24) |

| Cohort | Time window | Outcome | AURUM | | CORIVA | | GOLD | | SIDIAP | |
|---|---|---|---|---|---|---|---|---|---|---|
| | | | Unvaccinated | Vaccinated | Unvaccinated | Vaccinated | Unvaccinated | Vaccinated | Unvaccinated | Vaccinated |
| | | ATE | 71 (0.36) | 29 (0.49) | 128 (37.30) | < 5 | 9 (0.15) | < 5 | 347 (8.01) | 489 (10.97) |
| | | IS | 11 (0.06) | < 5 | 79 (23.02) | < 5 | < 5 | < 5 | 192 (4.43) | 267 (5.99) |
| | | TIA | 14 (0.07) | 11 (0.19) | 23 (6.70) | < 5 | < 5 | < 5 | 45 (1.04) | 90 (2.02) |
| | | MI | 46 (0.23) | 16 (0.27) | 36 (10.49) | < 5 | 6 (0.10) | < 5 | 127 (2.93) | 163 (3.66) |
| | | HF | 78 (0.39) | 43 (0.72) | 392 (114.22) | 17 (41.80) | 6 (0.10) | < 5 | 639 (14.75) | 1,137 (25.52) |
| | | HS | 6 (0.03) | < 5 | < 5 | < 5 | < 5 | < 5 | 38 (0.88) | 61 (1.37) |
| | | MP | 20 (0.10) | < 5 | < 5 | < 5 | < 5 | < 5 | 30 (0.69) | 22 (0.49) |
| | 31 to 90 days | VTE | 92 (0.47) | 21 (0.35) | 36 (10.49) | < 5 | 7 (0.12) | < 5 | 108 (2.49) | 128 (2.87) |
| | | DVT | 51 (0.26) | 7 (0.12) | 22 (6.41) | < 5 | < 5 | < 5 | 62 (1.43) | 81 (1.82) |
| | | PE | 44 (0.22) | 14 (0.24) | 17 (4.95) | < 5 | < 5 | < 5 | 56 (1.29) | 56 (1.26) |
| | | ATE | 38 (0.19) | 36 (0.61) | 33 (9.62) | < 5 | 8 (0.14) | < 5 | 201 (4.64) | 313 (7.02) |
| | | IS | 7 (0.04) | 5 (0.08) | 23 (6.70) | < 5 | < 5 | < 5 | 97 (2.24) | 167 (3.75) |
| | | TIA | 11 (0.06) | 9 (0.15) | 7 (2.04) | < 5 | < 5 | < 5 | 54 (1.25) | 86 (1.93) |
| | | MI | 21 (0.11) | 22 (0.37) | 10 (2.91) | < 5 | < 5 | < 5 | 63 (1.45) | 80 (1.80) |
| | | HF | 50 (0.25) | 35 (0.59) | 151 (44.00) | 10 (24.59) | 5 (0.09) | < 5 | 269 (6.21) | 554 (12.43) |
| | | HS | 5 (0.03) | < 5 | < 5 | < 5 | < 5 | < 5 | 18 (0.42) | 19 (0.43) |
| | | MP | 14 (0.07) | < 5 | < 5 | < 5 | < 5 | < 5 | 8 (0.18) | < 5 |
| | 91 to 180 days | VTE | 42 (0.21) | 10 (0.17) | 28 (8.16) | 5 (12.29) | 7 (0.12) | < 5 | 112 (2.59) | 96 (2.15) |
| | | DVT | 21 (0.11) | 6 (0.10) | 14 (4.08) | < 5 | < 5 | < 5 | 62 (1.43) | 59 (1.32) |
| | | PE | 21 (0.11) | 5 (0.08) | 15 (4.37) | < 5 | 7 (0.12) | < 5 | 56 (1.29) | 45 (1.01) |
| | | ATE | 31 (0.16) | 15 (0.25) | 29 (8.45) | < 5 | 5 (0.09) | < 5 | 184 (4.25) | 287 (6.44) |
| | | IS | < 5 | < 5 | 17 (4.95) | < 5 | < 5 | < 5 | 96 (2.22) | 168 (3.77) |
| | | TIA | 13 (0.07) | 5 (0.08) | 7 (2.04) | < 5 | < 5 | < 5 | 42 (0.97) | 76 (1.71) |
| | | MI | 17 (0.09) | 10 (0.17) | 8 (2.33) | < 5 | < 5 | < 5 | 52 (1.20) | 63 (1.41) |
| | | HF | 29 (0.15) | 25 (0.42) | 172 (50.12) | 13 (31.96) | < 5 | < 5 | 227 (5.24) | 472 (10.59) |
| | | HS | < 5 | < 5 | < 5 | < 5 | < 5 | < 5 | 18 (0.42) | 34 (0.76) |
| | | MP | < 5 | < 5 | < 5 | < 5 | < 5 | < 5 | 12 (0.28) | < 5 |
| | 181 to 365 days | VTE | 19 (0.10) | 8 (0.13) | 55 (16.03) | < 5 | < 5 | < 5 | 36 (0.83) | 40 (0.90) |
| | | DVT | 7 (0.04) | < 5 | 27 (7.87) | < 5 | < 5 | < 5 | 21 (0.48) | 26 (0.58) |
| | | PE | 13 (0.07) | 7 (0.12) | 33 (9.62) | < 5 | < 5 | < 5 | 18 (0.42) | 16 (0.36) |
| | | ATE | 14 (0.07) | 7 (0.12) | 62 (18.07) | < 5 | < 5 | < 5 | 110 (2.54) | 123 (2.76) |
| | | IS | < 5 | < 5 | 40 (11.66) | < 5 | < 5 | < 5 | 59 (1.36) | 58 (1.30) |
| | | TIA | < 5 | < 5 | 16 (4.66) | < 5 | < 5 | < 5 | 24 (0.55) | 39 (0.88) |
| | | MI | 10 (0.05) | < 5 | 12 (3.50) | < 5 | < 5 | < 5 | 32 (0.74) | 32 (0.72) |
| | | HF | 25 (0.13) | 12 (0.20) | 285 (83.04) | 15 (36.88) | < 5 | < 5 | 107 (2.47) | 212 (4.76) |
| | | HS | < 5 | < 5 | < 5 | < 5 | < 5 | < 5 | 7 (0.16) | 12 (0.27) |
| | | MP | < 5 | < 5 | < 5 | < 5 | < 5 | < 5 | < 5 | < 5 |
| Cohort 3 | | | N = 1,510,323 | N = 54,102 | N = 96,471 | N = 18,645 | N = 416,549 | N = 36,748 | N = 869,109 | N = 706,435 |
| | 0 to 30 days | VTE | 267 (1.77) | < 5 | 124 (12.85) | 7 (3.75) | 33 (0.79) | < 5 | 422 (4.86) | 114 (1.61) |
| | | DVT | 52 (0.34) | < 5 | 25 (2.59) | 5 (2.68) | 7 (0.17) | < 5 | 132 (1.52) | 51 (0.72) |
| | | PE | 224 (1.48) | < 5 | 100 (10.37) | < 5 | 27 (0.65) | < 5 | 326 (3.75) | 71 (1.01) |
| | | ATE | 34 (0.23) | < 5 | 131 (13.58) | 8 (4.29) | < 5 | < 5 | 314 (3.61) | 197 (2.79) |
| | | IS | 5 (0.03) | < 5 | 73 (7.57) | 6 (3.22) | < 5 | < 5 | 154 (1.77) | 95 (1.34) |
| | | TIA | 6 (0.04) | < 5 | 24 (2.49) | < 5 | < 5 | < 5 | 32 (0.37) | 42 (0.59) |
| | | MI | 23 (0.15) | < 5 | 42 (4.35) | < 5 | < 5 | < 5 | 137 (1.58) | 78 (1.10) |
| | | HF | 32 (0.21) | 10 (1.85) | 401 (41.57) | 16 (8.58) | < 5 | < 5 | 476 (5.48) | 163 (2.31) |
| | | HS | < 5 | < 5 | 5 (0.52) | < 5 | < 5 | < 5 | 34 (0.39) | 26 (0.37) |
| | | MP | 14 (0.09) | < 5 | < 5 | < 5 | < 5 | < 5 | 27 (0.31) | 13 (0.18) |
| | 31 to 90 days | VTE | 61 (0.40) | < 5 | 46 (4.77) | 5 (2.68) | < 5 | < 5 | 120 (1.38) | 65 (0.92) |

| Cohort | Time window | Outcome | AURUM | | CORIVA | | GOLD | | SIDIAP | |
|---|---|---|---|---|---|---|---|---|---|---|
| | | | Unvaccinated | Vaccinated | Unvaccinated | Vaccinated | Unvaccinated | Vaccinated | Unvaccinated | Vaccinated |
| | | **DVT** | 29 (0.19) | < 5 | 31 (3.21) | < 5 | < 5 | < 5 | 81 (0.93) | 47 (0.67) |
| | | **PE** | 35 (0.23) | < 5 | 19 (1.97) | < 5 | < 5 | < 5 | 51 (0.59) | 23 (0.33) |
| | | **ATE** | 14 (0.09) | < 5 | 43 (4.46) | 10 (5.36) | < 5 | < 5 | 171 (1.97) | 144 (2.04) |
| | | **IS** | < 5 | < 5 | 24 (2.49) | < 5 | < 5 | < 5 | 75 (0.86) | 55 (0.78) |
| | | **TIA** | < 5 | < 5 | 8 (0.83) | 6 (3.22) | < 5 | < 5 | 50 (0.58) | 28 (0.40) |
| | | **MI** | 11 (0.07) | < 5 | 16 (1.66) | < 5 | < 5 | < 5 | 58 (0.67) | 69 (0.98) |
| | | **HF** | 22 (0.15) | < 5 | 184 (19.07) | 11 (5.90) | < 5 | < 5 | 193 (2.22) | 111 (1.57) |
| | | **HS** | 5 (0.03) | < 5 | < 5 | < 5 | < 5 | < 5 | 13 (0.15) | 14 (0.20) |
| | | **MP** | 9 (0.06) | < 5 | 7 (0.73) | < 5 | < 5 | < 5 | 13 (0.15) | 9 (0.13) |
| | **91 to 180 days** | **VTE** | 29 (0.19) | < 5 | 49 (5.08) | 8 (4.29) | 5 (0.12) | < 5 | 103 (1.19) | 71 (1.01) |
| | | **DVT** | 15 (0.10) | < 5 | 31 (3.21) | 5 (2.68) | < 5 | < 5 | 60 (0.69) | 45 (0.64) |
| | | **PE** | 16 (0.11) | < 5 | 18 (1.87) | < 5 | < 5 | < 5 | 48 (0.55) | 27 (0.38) |
| | | **ATE** | 8 (0.05) | < 5 | 45 (4.66) | 7 (3.75) | < 5 | < 5 | 185 (2.13) | 141 (2.00) |
| | | **IS** | < 5 | < 5 | 20 (2.07) | < 5 | < 5 | < 5 | 88 (1.01) | 65 (0.92) |
| | | **TIA** | < 5 | < 5 | 13 (1.35) | < 5 | < 5 | < 5 | 33 (0.38) | 29 (0.41) |
| | | **MI** | < 5 | < 5 | 17 (1.76) | < 5 | < 5 | < 5 | 67 (0.77) | 53 (0.75) |
| | | **HF** | 11 (0.07) | < 5 | 214 (22.18) | 12 (6.44) | < 5 | < 5 | 181 (2.08) | 100 (1.42) |
| | | **HS** | < 5 | < 5 | < 5 | < 5 | < 5 | < 5 | 16 (0.18) | 10 (0.14) |
| | | **MP** | < 5 | < 5 | 6 (0.62) | < 5 | < 5 | < 5 | 11 (0.13) | 9 (0.13) |
| | **181 to 365 days** | **VTE** | < 5 | < 5 | 82 (8.50) | 14 (7.51) | < 5 | < 5 | 42 (0.48) | 17 (0.24) |
| | | **DVT** | < 5 | < 5 | 47 (4.87) | 12 (6.44) | < 5 | < 5 | 24 (0.28) | 14 (0.20) |
| | | **PE** | < 5 | < 5 | 38 (3.94) | < 5 | < 5 | < 5 | 22 (0.25) | < 5 |
| | | **ATE** | < 5 | < 5 | 86 (8.91) | 6 (3.22) | < 5 | < 5 | 70 (0.81) | 45 (0.64) |
| | | **IS** | < 5 | < 5 | 42 (4.35) | < 5 | < 5 | < 5 | 35 (0.40) | 17 (0.24) |
| | | **TIA** | < 5 | < 5 | 22 (2.28) | < 5 | < 5 | < 5 | 11 (0.13) | 7 (0.10) |
| | | **MI** | < 5 | < 5 | 30 (3.11) | < 5 | < 5 | < 5 | 25 (0.29) | 24 (0.34) |
| | | **HF** | 5 (0.03) | < 5 | 350 (36.28) | 26 (13.94) | < 5 | < 5 | 73 (0.84) | 31 (0.44) |
| | | **HS** | < 5 | < 5 | 6 (0.62) | < 5 | < 5 | < 5 | 5 (0.06) | < 5 |
| | | **MP** | < 5 | < 5 | 5 (0.52) | < 5 | < 5 | < 5 | < 5 | < 5 |
| **Cohort 4** | | | **N = 2,014,161** | **N = 1,335,671** | **N = 147,553** | **N = 15,683** | **N = 465,326** | **N = 365,096** | **N = 1,068,043** | **N = 580,329** |
| | **0 to 30 days** | **VTE** | 331 (1.64) | 20 (0.15) | 121 (8.20) | < 5 | 36 (0.77) | < 5 | 386 (3.61) | 66 (1.14) |
| | | **DVT** | 52 (0.26) | 9 (0.07) | 24 (1.63) | < 5 | 6 (0.13) | < 5 | 124 (1.16) | 35 (0.60) |
| | | **PE** | 290 (1.44) | 11 (0.08) | 98 (6.64) | < 5 | 31 (0.67) | < 5 | 298 (2.79) | 35 (0.60) |
| | | **ATE** | 25 (0.12) | < 5 | 122 (8.27) | 8 (5.10) | < 5 | < 5 | 277 (2.59) | 55 (0.95) |
| | | **IS** | < 5 | < 5 | 70 (4.74) | < 5 | < 5 | < 5 | 139 (1.30) | 29 (0.50) |
| | | **TIA** | 6 (0.03) | < 5 | 22 (1.49) | < 5 | < 5 | < 5 | 37 (0.35) | 7 (0.12) |
| | | **MI** | 15 (0.07) | < 5 | 38 (2.58) | < 5 | < 5 | < 5 | 108 (1.01) | 20 (0.34) |
| | | **HF** | 26 (0.13) | < 5 | 373 (25.28) | 13 (8.29) | < 5 | < 5 | 425 (3.98) | 44 (0.76) |
| | | **HS** | < 5 | < 5 | < 5 | < 5 | < 5 | < 5 | 32 (0.30) | 7 (0.12) |
| | | **MP** | 14 (0.07) | < 5 | 7 (0.47) | < 5 | < 5 | < 5 | 33 (0.31) | 11 (0.19) |
| | **31 to 90 days** | **VTE** | 57 (0.28) | 7 (0.05) | 46 (3.12) | < 5 | < 5 | < 5 | 114 (1.07) | 33 (0.57) |
| | | **DVT** | 21 (0.10) | < 5 | 31 (2.10) | < 5 | < 5 | < 5 | 80 (0.75) | 23 (0.40) |
| | | **PE** | 39 (0.19) | < 5 | 19 (1.29) | < 5 | < 5 | < 5 | 44 (0.41) | 11 (0.19) |
| | | **ATE** | 13 (0.06) | < 5 | 40 (2.71) | < 5 | < 5 | < 5 | 146 (1.37) | 51 (0.88) |
| | | **IS** | < 5 | < 5 | 23 (1.56) | < 5 | < 5 | < 5 | 64 (0.60) | 21 (0.36) |
| | | **TIA** | < 5 | < 5 | 8 (0.54) | < 5 | < 5 | < 5 | 41 (0.38) | 12 (0.21) |
| | | **MI** | 9 (0.04) | < 5 | 15 (1.02) | < 5 | < 5 | < 5 | 50 (0.47) | 21 (0.36) |
| | | **HF** | 14 (0.07) | 6 (0.04) | 167 (11.32) | 11 (7.01) | < 5 | < 5 | 162 (1.52) | 28 (0.48) |
| | | **HS** | 5 (0.02) | < 5 | < 5 | < 5 | < 5 | < 5 | 12 (0.11) | 5 (0.09) |

| Cohort | Time window | Outcome | AURUM | | CORIVA | | GOLD | | SIDIAP | |
|---|---|---|---|---|---|---|---|---|---|---|
| | | | Unvaccinated | Vaccinated | Unvaccinated | Vaccinated | Unvaccinated | Vaccinated | Unvaccinated | Vaccinated |
| | | **MP** | 8 (0.04) | < 5 | 8 (0.54) | < 5 | < 5 | < 5 | 14 (0.13) | 8 (0.14) |
| | **91 to 180 days** | **VTE** | 24 (0.12) | 9 (0.07) | 46 (3.12) | 7 (4.46) | < 5 | < 5 | 88 (0.82) | 45 (0.78) |
| | | **DVT** | 11 (0.05) | 7 (0.05) | 31 (2.10) | 6 (3.83) | < 5 | < 5 | 53 (0.50) | 28 (0.48) |
| | | **PE** | 14 (0.07) | < 5 | 16 (1.08) | < 5 | < 5 | < 5 | 40 (0.37) | 20 (0.34) |
| | | **ATE** | < 5 | < 5 | 33 (2.24) | 8 (5.10) | < 5 | < 5 | 150 (1.40) | 71 (1.22) |
| | | **IS** | < 5 | < 5 | 14 (0.95) | < 5 | < 5 | < 5 | 72 (0.67) | 37 (0.64) |
| | | **TIA** | < 5 | < 5 | 10 (0.68) | < 5 | < 5 | < 5 | 29 (0.27) | 14 (0.24) |
| | | **MI** | < 5 | < 5 | 13 (0.88) | < 5 | < 5 | < 5 | 52 (0.49) | 20 (0.34) |
| | | **HF** | 9 (0.04) | < 5 | 185 (12.54) | 9 (5.74) | < 5 | < 5 | 176 (1.65) | 35 (0.60) |
| | | **HS** | < 5 | < 5 | < 5 | < 5 | < 5 | < 5 | 20 (0.19) | < 5 |
| | | **MP** | < 5 | < 5 | 6 (0.41) | < 5 | < 5 | < 5 | 14 (0.13) | < 5 |
| | **181 to 365 days** | **VTE** | < 5 | < 5 | 80 (5.42) | 6 (3.83) | < 5 | < 5 | 51 (0.48) | 10 (0.17) |
| | | **DVT** | < 5 | < 5 | 48 (3.25) | 6 (3.83) | < 5 | < 5 | 33 (0.31) | 8 (0.14) |
| | | **PE** | < 5 | < 5 | 35 (2.37) | < 5 | < 5 | < 5 | 19 (0.18) | < 5 |
| | | **ATE** | < 5 | < 5 | 72 (4.88) | 6 (3.83) | < 5 | < 5 | 56 (0.52) | 21 (0.36) |
| | | **IS** | < 5 | < 5 | 35 (2.37) | < 5 | < 5 | < 5 | 28 (0.26) | 11 (0.19) |
| | | **TIA** | < 5 | < 5 | 19 (1.29) | < 5 | < 5 | < 5 | 10 (0.09) | < 5 |
| | | **MI** | < 5 | < 5 | 25 (1.69) | < 5 | < 5 | < 5 | 19 (0.18) | 9 (0.16) |
| | | **HF** | < 5 | < 5 | 311 (21.08) | 10 (6.38) | < 5 | < 5 | 63 (0.59) | 8 (0.14) |
| | | **HS** | < 5 | < 5 | 7 (0.47) | < 5 | < 5 | < 5 | 6 (0.06) | < 5 |
| | | **MP** | < 5 | < 5 | 7 (0.47) | < 5 | < 5 | < 5 | 6 (0.06) | < 5 |

VTE = Venous thromboembolism (DVT + PE), DVT = Deep vein thrombosis, PE = Pulmonary embolism, ATE = Arterial thromboembolism (IS + TIA + MI), IS = Ischemic stroke, TIA = Transient ischemic attack, MI = Myocardial infarction, HF = Heart failure, HS = Hemorrhagic stroke, MP = Myocarditis or Pericarditis

**Table S33: Number of records (and risk per 10,000 individuals) for post COVID-19 cardiac and thromboembolic complications,** across cohorts and databases, stratified by exposure status (BNT162b2 vaccine). Follow-up ends at first vaccine dose after index date.

| Cohort | Time window | Outcome | AURUM | | CORIVA | | GOLD | | SIDIAP | |
|---|---|---|---|---|---|---|---|---|---|---|
| | | | Unvaccinated | Vaccinated | Unvaccinated | Vaccinated | Unvaccinated | Vaccinated | Unvaccinated | Vaccinated |
| **Cohort 1** | | | **N = 348,052** | **N = 332,790** | **N = 24,073** | **N = 19,686** | **N = 169,459** | **N = 32,755** | **N = 223,960** | **N = 88,896** |
| | **0 to 30 days** | **VTE** | 106 (3.05) | 31 (0.93) | 78 (32.40) | < 5 | 8 (0.47) | < 5 | 76 (3.39) | < 5 |
| | | **DVT** | 21 (0.60) | 8 (0.24) | 12 (4.98) | < 5 | < 5 | < 5 | 19 (0.85) | < 5 |
| | | **PE** | 90 (2.59) | 24 (0.72) | 68 (28.25) | < 5 | 6 (0.35) | < 5 | 61 (2.72) | < 5 |
| | | **ATE** | 27 (0.78) | 18 (0.54) | 89 (36.97) | < 5 | 6 (0.35) | < 5 | 78 (3.48) | 5 (0.56) |
| | | **IS** | 9 (0.26) | < 5 | 50 (20.77) | < 5 | < 5 | < 5 | 46 (2.05) | < 5 |
| | | **TIA** | 5 (0.14) | < 5 | 17 (7.06) | < 5 | < 5 | < 5 | 7 (0.31) | < 5 |
| | | **MI** | 14 (0.40) | 14 (0.42) | 30 (12.46) | < 5 | < 5 | < 5 | 27 (1.21) | < 5 |
| | | **HF** | 59 (1.70) | 45 (1.35) | 352 (146.22) | 17 (8.64) | 9 (0.53) | < 5 | 301 (13.44) | 23 (2.59) |
| | | **HS** | < 5 | < 5 | < 5 | < 5 | < 5 | < 5 | 7 (0.31) | < 5 |
| | | **MP** | < 5 | < 5 | < 5 | < 5 | < 5 | < 5 | < 5 | < 5 |
| | **31 to 90 days** | **VTE** | 18 (0.52) | 15 (0.45) | 29 (12.05) | < 5 | < 5 | < 5 | 15 (0.67) | < 5 |
| | | **DVT** | 9 (0.26) | < 5 | 16 (6.65) | < 5 | < 5 | < 5 | 9 (0.40) | < 5 |
| | | **PE** | 9 (0.26) | 11 (0.33) | 15 (6.23) | < 5 | < 5 | < 5 | 6 (0.27) | < 5 |
| | | **ATE** | 9 (0.26) | 8 (0.24) | 20 (8.31) | < 5 | < 5 | < 5 | 42 (1.88) | 5 (0.56) |
| | | **IS** | < 5 | < 5 | 11 (4.57) | < 5 | < 5 | < 5 | 20 (0.89) | < 5 |
| | | **TIA** | 5 (0.14) | < 5 | 6 (2.49) | < 5 | < 5 | < 5 | 14 (0.63) | < 5 |
| | | **MI** | 5 (0.14) | < 5 | 9 (3.74) | < 5 | < 5 | < 5 | 11 (0.49) | < 5 |
| | | **HF** | 32 (0.92) | 36 (1.08) | 115 (47.77) | 13 (6.60) | < 5 | < 5 | 87 (3.88) | 8 (0.90) |
| | | **HS** | < 5 | < 5 | < 5 | < 5 | < 5 | < 5 | < 5 | < 5 |
| | | **MP** | < 5 | < 5 | < 5 | < 5 | < 5 | < 5 | < 5 | < 5 |
| | **91 to 180 days** | **VTE** | 10 (0.29) | < 5 | 19 (7.89) | < 5 | < 5 | < 5 | 20 (0.89) | < 5 |
| | | **DVT** | < 5 | < 5 | 11 (4.57) | < 5 | < 5 | < 5 | 9 (0.40) | < 5 |
| | | **PE** | 6 (0.17) | < 5 | 10 (4.15) | < 5 | < 5 | < 5 | 11 (0.49) | < 5 |
| | | **ATE** | 15 (0.43) | 10 (0.30) | 22 (9.14) | < 5 | < 5 | < 5 | 31 (1.38) | < 5 |
| | | **IS** | < 5 | < 5 | 10 (4.15) | < 5 | < 5 | < 5 | 16 (0.71) | < 5 |
| | | **TIA** | 8 (0.23) | < 5 | 7 (2.91) | < 5 | < 5 | < 5 | 9 (0.40) | < 5 |
| | | **MI** | 5 (0.14) | < 5 | 8 (3.32) | < 5 | < 5 | < 5 | 7 (0.31) | < 5 |
| | | **HF** | 43 (1.24) | 32 (0.96) | 134 (55.66) | 12 (6.10) | < 5 | < 5 | 86 (3.84) | 6 (0.67) |
| | | **HS** | < 5 | < 5 | < 5 | < 5 | < 5 | < 5 | < 5 | < 5 |
| | | **MP** | < 5 | < 5 | < 5 | < 5 | < 5 | < 5 | < 5 | < 5 |
| | **181 to 365 days** | **VTE** | 14 (0.40) | 7 (0.21) | 48 (19.94) | < 5 | < 5 | < 5 | 10 (0.45) | < 5 |
| | | **DVT** | < 5 | < 5 | 13 (5.40) | < 5 | < 5 | < 5 | 5 (0.22) | < 5 |
| | | **PE** | 11 (0.32) | < 5 | 40 (16.62) | < 5 | < 5 | < 5 | 5 (0.22) | < 5 |
| | | **ATE** | 15 (0.43) | 11 (0.33) | 45 (18.69) | < 5 | < 5 | < 5 | 42 (1.88) | 8 (0.90) |
| | | **IS** | < 5 | < 5 | 25 (10.39) | < 5 | < 5 | < 5 | 18 (0.80) | 5 (0.56) |
| | | **TIA** | 10 (0.29) | < 5 | 13 (5.40) | < 5 | < 5 | < 5 | 13 (0.58) | < 5 |
| | | **MI** | < 5 | 6 (0.18) | 12 (4.98) | < 5 | < 5 | < 5 | 11 (0.49) | < 5 |
| | | **HF** | 52 (1.49) | 35 (1.05) | 218 (90.56) | 29 (14.73) | < 5 | < 5 | 84 (3.75) | 20 (2.25) |
| | | **HS** | < 5 | < 5 | < 5 | < 5 | < 5 | < 5 | < 5 | < 5 |
| | | **MP** | < 5 | < 5 | < 5 | < 5 | < 5 | < 5 | < 5 | < 5 |
| **Cohort 2** | | | **N = 1,976,163** | **N = 594,262** | **N = 34,320** | **N = 4,067** | **N = 584,309** | **N = 180,670** | **N = 433,111** | **N = 445,581** |
| | **0 to 30 days** | **VTE** | 240 (1.21) | 14 (0.24) | 71 (20.69) | < 5 | 29 (0.50) | < 5 | 255 (5.89) | 14 (0.31) |
| | | **DVT** | 39 (0.20) | 6 (0.10) | 10 (2.91) | < 5 | 7 (0.12) | < 5 | 63 (1.45) | < 5 |
| | | **PE** | 205 (1.04) | 8 (0.13) | 62 (18.07) | < 5 | 23 (0.39) | < 5 | 211 (4.87) | 12 (0.27) |

| Cohort | Time window | Outcome | AURUM | | CORIVA | | GOLD | | SIDIAP | |
|---|---|---|---|---|---|---|---|---|---|---|
| | | | Unvaccinated | Vaccinated | Unvaccinated | Vaccinated | Unvaccinated | Vaccinated | Unvaccinated | Vaccinated |
| | | **ATE** | 39 (0.20) | 14 (0.24) | 89 (25.93) | < 5 | < 5 | < 5 | 175 (4.04) | 16 (0.36) |
| | | **IS** | 7 (0.04) | < 5 | 52 (15.15) | < 5 | < 5 | < 5 | 97 (2.24) | 8 (0.18) |
| | | **TIA** | 6 (0.03) | < 5 | 16 (4.66) | < 5 | < 5 | < 5 | 17 (0.39) | < 5 |
| | | **MI** | 26 (0.13) | 8 (0.13) | 27 (7.87) | < 5 | < 5 | < 5 | 65 (1.50) | 5 (0.11) |
| | | **HF** | 50 (0.25) | 15 (0.25) | 312 (90.91) | < 5 | 6 (0.10) | < 5 | 380 (8.77) | 56 (1.26) |
| | | **HS** | 6 (0.03) | < 5 | < 5 | < 5 | < 5 | < 5 | 17 (0.39) | < 5 |
| | | **MP** | 11 (0.06) | < 5 | < 5 | < 5 | < 5 | < 5 | 14 (0.32) | < 5 |
| | **31 to 90 days** | **VTE** | 49 (0.25) | 5 (0.08) | 24 (6.99) | < 5 | < 5 | < 5 | 60 (1.39) | 6 (0.13) |
| | | **DVT** | 26 (0.13) | < 5 | 13 (3.79) | < 5 | < 5 | < 5 | 33 (0.76) | < 5 |
| | | **PE** | 26 (0.13) | < 5 | 13 (3.79) | < 5 | < 5 | < 5 | 33 (0.76) | < 5 |
| | | **ATE** | 17 (0.09) | 9 (0.15) | 20 (5.83) | < 5 | < 5 | < 5 | 85 (1.96) | 7 (0.16) |
| | | **IS** | < 5 | < 5 | 13 (3.79) | < 5 | < 5 | < 5 | 43 (0.99) | < 5 |
| | | **TIA** | < 5 | < 5 | < 5 | < 5 | < 5 | < 5 | 23 (0.53) | < 5 |
| | | **MI** | 11 (0.06) | < 5 | 9 (2.62) | < 5 | < 5 | < 5 | 26 (0.60) | < 5 |
| | | **HF** | 27 (0.14) | 17 (0.29) | 107 (31.18) | < 5 | < 5 | < 5 | 135 (3.12) | 19 (0.43) |
| | | **HS** | < 5 | < 5 | < 5 | < 5 | < 5 | < 5 | 7 (0.16) | < 5 |
| | | **MP** | 6 (0.03) | < 5 | < 5 | < 5 | < 5 | < 5 | < 5 | < 5 |
| | **91 to 180 days** | **VTE** | 28 (0.14) | < 5 | 22 (6.41) | < 5 | 5 (0.09) | < 5 | 59 (1.36) | < 5 |
| | | **DVT** | 12 (0.06) | < 5 | 11 (3.21) | < 5 | < 5 | < 5 | 31 (0.72) | < 5 |
| | | **PE** | 16 (0.08) | < 5 | 12 (3.50) | < 5 | < 5 | < 5 | 30 (0.69) | < 5 |
| | | **ATE** | 14 (0.07) | 5 (0.08) | 18 (5.24) | < 5 | < 5 | < 5 | 87 (2.01) | 10 (0.22) |
| | | **IS** | < 5 | < 5 | 7 (2.04) | < 5 | < 5 | < 5 | 48 (1.11) | 5 (0.11) |
| | | **TIA** | 7 (0.04) | < 5 | 6 (1.75) | < 5 | < 5 | < 5 | 19 (0.44) | < 5 |
| | | **MI** | 8 (0.04) | < 5 | 7 (2.04) | < 5 | < 5 | < 5 | 23 (0.53) | < 5 |
| | | **HF** | 19 (0.10) | 9 (0.15) | 130 (37.88) | < 5 | < 5 | < 5 | 114 (2.63) | 25 (0.56) |
| | | **HS** | < 5 | < 5 | < 5 | < 5 | < 5 | < 5 | 7 (0.16) | < 5 |
| | | **MP** | < 5 | < 5 | < 5 | < 5 | < 5 | < 5 | 7 (0.16) | < 5 |
| | **181 to 365 days** | **VTE** | 10 (0.05) | 7 (0.12) | 39 (11.36) | < 5 | < 5 | < 5 | 16 (0.37) | 10 (0.22) |
| | | **DVT** | < 5 | < 5 | 15 (4.37) | < 5 | < 5 | < 5 | 11 (0.25) | 6 (0.13) |
| | | **PE** | 8 (0.04) | 5 (0.08) | 28 (8.16) | < 5 | < 5 | < 5 | 7 (0.16) | < 5 |
| | | **ATE** | 12 (0.06) | 6 (0.10) | 44 (12.82) | < 5 | < 5 | < 5 | 65 (1.50) | 19 (0.43) |
| | | **IS** | < 5 | < 5 | 29 (8.45) | < 5 | < 5 | < 5 | 35 (0.81) | 9 (0.20) |
| | | **TIA** | < 5 | < 5 | 12 (3.50) | < 5 | < 5 | < 5 | 19 (0.44) | 5 (0.11) |
| | | **MI** | 9 (0.05) | < 5 | 8 (2.33) | < 5 | < 5 | < 5 | 13 (0.30) | 5 (0.11) |
| | | **HF** | 23 (0.12) | 11 (0.19) | 213 (62.06) | < 5 | < 5 | < 5 | 73 (1.69) | 27 (0.61) |
| | | **HS** | < 5 | < 5 | < 5 | < 5 | < 5 | < 5 | 5 (0.12) | < 5 |
| | | **MP** | < 5 | < 5 | < 5 | < 5 | < 5 | < 5 | < 5 | < 5 |
| **Cohort 3** | | | **N = 1,510,323** | **N = 54,102** | **N = 96,471** | **N = 18,645** | **N = 416,549** | **N = 36,748** | **N = 869,109** | **N = 706,435** |
| | **0 to 30 days** | **VTE** | 244 (1.62) | < 5 | 105 (10.88) | < 5 | 28 (0.67) | < 5 | 326 (3.75) | < 5 |
| | | **DVT** | 44 (0.29) | < 5 | 19 (1.97) | < 5 | 6 (0.14) | < 5 | 92 (1.06) | < 5 |
| | | **PE** | 209 (1.38) | < 5 | 88 (9.12) | < 5 | 23 (0.55) | < 5 | 262 (3.01) | < 5 |
| | | **ATE** | 29 (0.19) | < 5 | 95 (9.85) | < 5 | < 5 | < 5 | 212 (2.44) | < 5 |
| | | **IS** | < 5 | < 5 | 53 (5.49) | < 5 | < 5 | < 5 | 102 (1.17) | < 5 |
| | | **TIA** | 5 (0.03) | < 5 | 16 (1.66) | < 5 | < 5 | < 5 | 20 (0.23) | < 5 |
| | | **MI** | 20 (0.13) | < 5 | 31 (3.21) | < 5 | < 5 | < 5 | 96 (1.10) | < 5 |
| | | **HF** | 30 (0.20) | 7 (1.29) | 329 (34.10) | < 5 | < 5 | < 5 | 364 (4.19) | 5 (0.07) |
| | | **HS** | < 5 | < 5 | < 5 | < 5 | < 5 | < 5 | 20 (0.23) | < 5 |
| | | **MP** | 9 (0.06) | < 5 | < 5 | < 5 | < 5 | < 5 | 19 (0.22) | < 5 |
| | **31 to 90 days** | **VTE** | 44 (0.29) | < 5 | 32 (3.32) | < 5 | < 5 | < 5 | 85 (0.98) | < 5 |

| Cohort | Time window | Outcome | AURUM | | CORIVA | | GOLD | | SIDIAP | |
|---|---|---|---|---|---|---|---|---|---|---|
| | | | Unvaccinated | Vaccinated | Unvaccinated | Vaccinated | Unvaccinated | Vaccinated | Unvaccinated | Vaccinated |
| | | DVT | 20 (0.13) | < 5 | 19 (1.97) | < 5 | < 5 | < 5 | 56 (0.64) | < 5 |
| | | PE | 27 (0.18) | < 5 | 16 (1.66) | < 5 | < 5 | < 5 | 39 (0.45) | < 5 |
| | | ATE | 11 (0.07) | < 5 | 34 (3.52) | < 5 | < 5 | < 5 | 107 (1.23) | < 5 |
| | | IS | < 5 | < 5 | 16 (1.66) | < 5 | < 5 | < 5 | 52 (0.60) | < 5 |
| | | TIA | < 5 | < 5 | 6 (0.62) | < 5 | < 5 | < 5 | 27 (0.31) | < 5 |
| | | MI | 9 (0.06) | < 5 | 15 (1.55) | < 5 | < 5 | < 5 | 36 (0.41) | < 5 |
| | | HF | 15 (0.10) | < 5 | 133 (13.79) | < 5 | < 5 | < 5 | 141 (1.62) | < 5 |
| | | HS | < 5 | < 5 | < 5 | < 5 | < 5 | < 5 | 7 (0.08) | < 5 |
| | | MP | 8 (0.05) | < 5 | 6 (0.62) | < 5 | < 5 | < 5 | 8 (0.09) | < 5 |
| | 91 to 180 days | VTE | 24 (0.16) | < 5 | 38 (3.94) | < 5 | < 5 | < 5 | 65 (0.75) | < 5 |
| | | DVT | 11 (0.07) | < 5 | 23 (2.38) | < 5 | < 5 | < 5 | 37 (0.43) | < 5 |
| | | PE | 14 (0.09) | < 5 | 16 (1.66) | < 5 | < 5 | < 5 | 32 (0.37) | < 5 |
| | | ATE | < 5 | < 5 | 26 (2.70) | < 5 | < 5 | < 5 | 113 (1.30) | 6 (0.08) |
| | | IS | < 5 | < 5 | 11 (1.14) | < 5 | < 5 | < 5 | 57 (0.66) | < 5 |
| | | TIA | < 5 | < 5 | 9 (0.93) | < 5 | < 5 | < 5 | 22 (0.25) | < 5 |
| | | MI | < 5 | < 5 | 10 (1.04) | < 5 | < 5 | < 5 | 37 (0.43) | < 5 |
| | | HF | 10 (0.07) | < 5 | 166 (17.21) | < 5 | < 5 | < 5 | 122 (1.40) | < 5 |
| | | HS | < 5 | < 5 | < 5 | < 5 | < 5 | < 5 | 9 (0.10) | < 5 |
| | | MP | < 5 | < 5 | < 5 | < 5 | < 5 | < 5 | 7 (0.08) | < 5 |
| | 181 to 365 days | VTE | < 5 | < 5 | 62 (6.43) | < 5 | < 5 | < 5 | 34 (0.39) | < 5 |
| | | DVT | < 5 | < 5 | 32 (3.32) | < 5 | < 5 | < 5 | 20 (0.23) | < 5 |
| | | PE | < 5 | < 5 | 33 (3.42) | < 5 | < 5 | < 5 | 16 (0.18) | < 5 |
| | | ATE | < 5 | < 5 | 60 (6.22) | < 5 | < 5 | < 5 | 51 (0.59) | 6 (0.08) |
| | | IS | < 5 | < 5 | 31 (3.21) | < 5 | < 5 | < 5 | 24 (0.28) | < 5 |
| | | TIA | < 5 | < 5 | 16 (1.66) | < 5 | < 5 | < 5 | 10 (0.12) | < 5 |
| | | MI | < 5 | < 5 | 20 (2.07) | < 5 | < 5 | < 5 | 18 (0.21) | < 5 |
| | | HF | 5 (0.03) | < 5 | 270 (27.99) | < 5 | < 5 | < 5 | 60 (0.69) | < 5 |
| | | HS | < 5 | < 5 | < 5 | < 5 | < 5 | < 5 | < 5 | < 5 |
| | | MP | < 5 | < 5 | < 5 | < 5 | < 5 | < 5 | < 5 | < 5 |
| Cohort 4 | | | N = 2,014,161 | N = 1,335,671 | N = 147,553 | N = 15,683 | N = 465,326 | N = 365,096 | N = 1,068,043 | N = 580,329 |
| | 0 to 30 days | VTE | 328 (1.63) | 9 (0.07) | 106 (7.18) | < 5 | 36 (0.77) | < 5 | 347 (3.25) | 18 (0.31) |
| | | DVT | 52 (0.26) | 5 (0.04) | 21 (1.42) | < 5 | 6 (0.13) | < 5 | 105 (0.98) | 8 (0.14) |
| | | PE | 287 (1.42) | < 5 | 87 (5.90) | < 5 | 31 (0.67) | < 5 | 271 (2.54) | 10 (0.17) |
| | | ATE | 25 (0.12) | < 5 | 94 (6.37) | < 5 | < 5 | < 5 | 234 (2.19) | 9 (0.16) |
| | | IS | < 5 | < 5 | 53 (3.59) | < 5 | < 5 | < 5 | 114 (1.07) | 6 (0.10) |
| | | TIA | 6 (0.03) | < 5 | 16 (1.08) | < 5 | < 5 | < 5 | 28 (0.26) | < 5 |
| | | MI | 15 (0.07) | < 5 | 30 (2.03) | < 5 | < 5 | < 5 | 98 (0.92) | < 5 |
| | | HF | 26 (0.13) | < 5 | 316 (21.42) | < 5 | < 5 | < 5 | 363 (3.40) | 7 (0.12) |
| | | HS | < 5 | < 5 | < 5 | < 5 | < 5 | < 5 | 23 (0.22) | < 5 |
| | | MP | 13 (0.06) | < 5 | 6 (0.41) | < 5 | < 5 | < 5 | 30 (0.28) | < 5 |
| | 31 to 90 days | VTE | 56 (0.28) | < 5 | 34 (2.30) | < 5 | < 5 | < 5 | 93 (0.87) | 6 (0.10) |
| | | DVT | 20 (0.10) | < 5 | 21 (1.42) | < 5 | < 5 | < 5 | 64 (0.60) | < 5 |
| | | PE | 39 (0.19) | < 5 | 16 (1.08) | < 5 | < 5 | < 5 | 38 (0.36) | < 5 |
| | | ATE | 13 (0.06) | < 5 | 32 (2.17) | < 5 | < 5 | < 5 | 117 (1.10) | < 5 |
| | | IS | < 5 | < 5 | 16 (1.08) | < 5 | < 5 | < 5 | 51 (0.48) | < 5 |
| | | TIA | < 5 | < 5 | 5 (0.34) | < 5 | < 5 | < 5 | 31 (0.29) | < 5 |
| | | MI | 9 (0.04) | < 5 | 14 (0.95) | < 5 | < 5 | < 5 | 43 (0.40) | < 5 |
| | | HF | 14 (0.07) | 6 (0.04) | 126 (8.54) | < 5 | < 5 | < 5 | 140 (1.31) | < 5 |
| | | HS | < 5 | < 5 | < 5 | < 5 | < 5 | < 5 | 9 (0.08) | < 5 |

| Cohort | Time window | Outcome | AURUM | | CORIVA | | GOLD | | SIDIAP | |
|---|---|---|---|---|---|---|---|---|---|---|
| | | | Unvaccinated | Vaccinated | Unvaccinated | Vaccinated | Unvaccinated | Vaccinated | Unvaccinated | Vaccinated |
| | | MP | 7 (0.03) | < 5 | 6 (0.41) | < 5 | < 5 | < 5 | 11 (0.10) | < 5 |
| | 91 to 180 days | VTE | 24 (0.12) | 8 (0.06) | 41 (2.78) | < 5 | < 5 | < 5 | 69 (0.65) | < 5 |
| | | DVT | 11 (0.05) | 7 (0.05) | 27 (1.83) | < 5 | < 5 | < 5 | 38 (0.36) | < 5 |
| | | PE | 14 (0.07) | < 5 | 15 (1.02) | < 5 | < 5 | < 5 | 35 (0.33) | < 5 |
| | | ATE | < 5 | < 5 | 24 (1.63) | < 5 | < 5 | < 5 | 124 (1.16) | 9 (0.16) |
| | | IS | < 5 | < 5 | 10 (0.68) | < 5 | < 5 | < 5 | 61 (0.57) | 7 (0.12) |
| | | TIA | < 5 | < 5 | 9 (0.61) | < 5 | < 5 | < 5 | 27 (0.25) | < 5 |
| | | MI | < 5 | < 5 | 9 (0.61) | < 5 | < 5 | < 5 | 39 (0.37) | < 5 |
| | | HF | 9 (0.04) | < 5 | 149 (10.10) | < 5 | < 5 | < 5 | 142 (1.33) | 9 (0.16) |
| | | HS | < 5 | < 5 | < 5 | < 5 | < 5 | < 5 | 16 (0.15) | < 5 |
| | | MP | < 5 | < 5 | < 5 | < 5 | < 5 | < 5 | 12 (0.11) | < 5 |
| | 181 to 365 days | VTE | < 5 | < 5 | 64 (4.34) | < 5 | < 5 | < 5 | 51 (0.48) | 7 (0.12) |
| | | DVT | < 5 | < 5 | 36 (2.44) | < 5 | < 5 | < 5 | 33 (0.31) | 5 (0.09) |
| | | PE | < 5 | < 5 | 31 (2.10) | < 5 | < 5 | < 5 | 19 (0.18) | < 5 |
| | | ATE | < 5 | < 5 | 56 (3.80) | < 5 | < 5 | < 5 | 54 (0.51) | 15 (0.26) |
| | | IS | < 5 | < 5 | 28 (1.90) | < 5 | < 5 | < 5 | 26 (0.24) | 10 (0.17) |
| | | TIA | < 5 | < 5 | 15 (1.02) | < 5 | < 5 | < 5 | 10 (0.09) | < 5 |
| | | MI | < 5 | < 5 | 19 (1.29) | < 5 | < 5 | < 5 | 19 (0.18) | 5 (0.09) |
| | | HF | < 5 | < 5 | 242 (16.40) | < 5 | < 5 | < 5 | 61 (0.57) | < 5 |
| | | HS | < 5 | < 5 | 6 (0.41) | < 5 | < 5 | < 5 | 5 (0.05) | < 5 |
| | | MP | < 5 | < 5 | < 5 | < 5 | < 5 | < 5 | 6 (0.06) | < 5 |

VTE = Venous thromboembolism (DVT + PE), DVT = Deep vein thrombosis, PE = Pulmonary embolism, ATE = Arterial thromboembolism (IS + TIA + MI), IS = Ischemic stroke, TIA = Transient ischemic attack, MI = Myocardial infarction, HF = Heart failure, HS = Hemorrhagic stroke, MP = Myocarditis or Pericarditis

**Table S34: Number of records (and risk per 10,000 individuals) for post COVID-19 cardiac and thromboembolic complications,** across cohorts and databases, stratified by exposure status (BNT162b2 vaccine). Only first outcome after COVID-19 captured.

| Cohort | Time window | Outcome | AURUM | | CORIVA | | GOLD | | SIDIAP | |
|---|---|---|---|---|---|---|---|---|---|---|
| | | | Unvaccinated | Vaccinated | Unvaccinated | Vaccinated | Unvaccinated | Vaccinated | Unvaccinated | Vaccinated |
| **Cohort 1** | | | **N = 348,052** | **N = 332,790** | **N = 24,073** | **N = 19,686** | **N = 169,459** | **N = 32,755** | **N = 223,960** | **N = 88,896** |
| | **0 to 30 days** | **VTE** | 167 (4.80) | 76 (2.28) | 97 (40.29) | 31 (15.75) | 15 (0.89) | < 5 | 251 (11.21) | 94 (10.57) |
| | | **DVT** | 39 (1.12) | 15 (0.45) | 18 (7.48) | 8 (4.06) | < 5 | < 5 | 76 (3.39) | 29 (3.26) |
| | | **PE** | 138 (3.96) | 64 (1.92) | 81 (33.65) | 24 (12.19) | 12 (0.71) | < 5 | 187 (8.35) | 75 (8.44) |
| | | **ATE** | 62 (1.78) | 45 (1.35) | 125 (51.93) | 37 (18.80) | 8 (0.47) | < 5 | 353 (15.76) | 206 (23.17) |
| | | **IS** | 16 (0.46) | 5 (0.15) | 77 (31.99) | 18 (9.14) | < 5 | < 5 | 196 (8.75) | 116 (13.05) |
| | | **TIA** | 14 (0.40) | 11 (0.33) | 23 (9.55) | 7 (3.56) | < 5 | < 5 | 58 (2.59) | 41 (4.61) |
| | | **MI** | 34 (0.98) | 29 (0.87) | 37 (15.37) | 19 (9.65) | 6 (0.35) | < 5 | 115 (5.13) | 61 (6.86) |
| | | **HF** | 136 (3.91) | 126 (3.79) | 438 (181.95) | 154 (78.23) | 20 (1.18) | < 5 | 1,203 (53.71) | 634 (71.32) |
| | | **HS** | < 5 | < 5 | < 5 | < 5 | < 5 | < 5 | 46 (2.05) | 14 (1.57) |
| | | **MP** | < 5 | < 5 | < 5 | < 5 | < 5 | < 5 | 16 (0.71) | < 5 |
| | **31 to 90 days** | **VTE** | 31 (0.89) | 25 (0.75) | 37 (15.37) | 8 (4.06) | 5 (0.30) | < 5 | 88 (3.93) | 43 (4.84) |
| | | **DVT** | 13 (0.37) | 7 (0.21) | 22 (9.14) | < 5 | < 5 | < 5 | 53 (2.37) | 28 (3.15) |
| | | **PE** | 18 (0.52) | 18 (0.54) | 17 (7.06) | < 5 | < 5 | < 5 | 41 (1.83) | 19 (2.14) |
| | | **ATE** | 20 (0.57) | 23 (0.69) | 30 (12.46) | 20 (10.16) | < 5 | < 5 | 216 (9.64) | 125 (14.06) |
| | | **IS** | < 5 | < 5 | 18 (7.48) | 10 (5.08) | < 5 | < 5 | 110 (4.91) | 69 (7.76) |
| | | **TIA** | 10 (0.29) | 10 (0.30) | 10 (4.15) | 7 (3.56) | < 5 | < 5 | 70 (3.13) | 32 (3.60) |
| | | **MI** | 11 (0.32) | 11 (0.33) | 10 (4.15) | < 5 | < 5 | < 5 | 49 (2.19) | 28 (3.15) |
| | | **HF** | 75 (2.15) | 70 (2.10) | 142 (58.99) | 65 (33.02) | 6 (0.35) | < 5 | 473 (21.12) | 288 (32.40) |
| | | **HS** | < 5 | < 5 | < 5 | < 5 | < 5 | < 5 | 16 (0.71) | 12 (1.35) |
| | | **MP** | < 5 | < 5 | < 5 | < 5 | < 5 | < 5 | < 5 | < 5 |
| | **91 to 180 days** | **VTE** | 17 (0.49) | < 5 | 22 (9.14) | 12 (6.10) | 6 (0.35) | < 5 | 62 (2.77) | 35 (3.94) |
| | | **DVT** | 6 (0.17) | < 5 | 10 (4.15) | < 5 | < 5 | < 5 | 37 (1.65) | 19 (2.14) |
| | | **PE** | 11 (0.32) | < 5 | 14 (5.82) | 9 (4.57) | < 5 | < 5 | 29 (1.29) | 18 (2.02) |
| | | **ATE** | 27 (0.78) | 18 (0.54) | 31 (12.88) | 15 (7.62) | < 5 | < 5 | 185 (8.26) | 102 (11.47) |
| | | **IS** | < 5 | 6 (0.18) | 19 (7.89) | 5 (2.54) | < 5 | < 5 | 114 (5.09) | 56 (6.30) |
| | | **TIA** | 13 (0.37) | 6 (0.18) | 8 (3.32) | < 5 | < 5 | < 5 | 41 (1.83) | 33 (3.71) |
| | | **MI** | 12 (0.34) | 6 (0.18) | 8 (3.32) | 10 (5.08) | < 5 | < 5 | 38 (1.70) | 22 (2.47) |
| | | **HF** | 64 (1.84) | 54 (1.62) | 166 (68.96) | 91 (46.23) | < 5 | < 5 | 398 (17.77) | 226 (25.42) |
| | | **HS** | < 5 | < 5 | < 5 | < 5 | < 5 | < 5 | 21 (0.94) | < 5 |
| | | **MP** | < 5 | < 5 | < 5 | < 5 | < 5 | < 5 | < 5 | < 5 |
| | **181 to 365 days** | **VTE** | 12 (0.34) | 10 (0.30) | 50 (20.77) | 21 (10.67) | < 5 | < 5 | 26 (1.16) | 10 (1.12) |
| | | **DVT** | < 5 | 6 (0.18) | 16 (6.65) | 13 (6.60) | < 5 | < 5 | 16 (0.71) | 7 (0.79) |
| | | **PE** | 9 (0.26) | < 5 | 39 (16.20) | 9 (4.57) | < 5 | < 5 | 12 (0.54) | < 5 |
| | | **ATE** | 18 (0.52) | 14 (0.42) | 51 (21.19) | 31 (15.75) | < 5 | < 5 | 104 (4.64) | 38 (4.27) |
| | | **IS** | < 5 | < 5 | 28 (11.63) | 15 (7.62) | < 5 | < 5 | 50 (2.23) | 23 (2.59) |
| | | **TIA** | 11 (0.32) | < 5 | 14 (5.82) | 13 (6.60) | < 5 | < 5 | 28 (1.25) | 9 (1.01) |
| | | **MI** | < 5 | 7 (0.21) | 14 (5.82) | 9 (4.57) | < 5 | < 5 | 30 (1.34) | 7 (0.79) |
| | | **HF** | 60 (1.72) | 35 (1.05) | 257 (106.76) | 139 (70.61) | < 5 | < 5 | 192 (8.57) | 117 (13.16) |
| | | **HS** | < 5 | < 5 | < 5 | < 5 | < 5 | < 5 | 6 (0.27) | 7 (0.79) |
| | | **MP** | < 5 | < 5 | < 5 | < 5 | < 5 | < 5 | < 5 | < 5 |
| **Cohort 2** | | | **N = 1,976,163** | **N = 594,262** | **N = 34,320** | **N = 4,067** | **N = 584,309** | **N = 180,670** | **N = 433,111** | **N = 445,581** |
| | **0 to 30 days** | **VTE** | 344 (1.74) | 62 (1.04) | 89 (25.93) | 8 (19.67) | 40 (0.68) | 5 (0.28) | 396 (9.14) | 265 (5.95) |
| | | **DVT** | 72 (0.36) | 18 (0.30) | 16 (4.66) | < 5 | 8 (0.14) | < 5 | 117 (2.70) | 96 (2.15) |
| | | **PE** | 279 (1.41) | 45 (0.76) | 74 (21.56) | 5 (12.29) | 33 (0.56) | < 5 | 309 (7.13) | 189 (4.24) |

| Cohort | Time window | Outcome | AURUM | | CORIVA | | GOLD | | SIDIAP | |
|---|---|---|---|---|---|---|---|---|---|---|
| | | | Unvaccinated | Vaccinated | Unvaccinated | Vaccinated | Unvaccinated | Vaccinated | Unvaccinated | Vaccinated |
| | | **ATE** | 71 (0.36) | 29 (0.49) | 128 (37.30) | < 5 | 9 (0.15) | < 5 | 347 (8.01) | 489 (10.97) |
| | | **IS** | 11 (0.06) | < 5 | 79 (23.02) | < 5 | < 5 | < 5 | 192 (4.43) | 267 (5.99) |
| | | **TIA** | 14 (0.07) | 11 (0.19) | 23 (6.70) | < 5 | < 5 | < 5 | 45 (1.04) | 90 (2.02) |
| | | **MI** | 46 (0.23) | 16 (0.27) | 36 (10.49) | < 5 | 6 (0.10) | < 5 | 127 (2.93) | 163 (3.66) |
| | | **HF** | 78 (0.39) | 43 (0.72) | 392 (114.22) | 17 (41.80) | 6 (0.10) | < 5 | 639 (14.75) | 1,137 (25.52) |
| | | **HS** | 6 (0.03) | < 5 | < 5 | < 5 | < 5 | < 5 | 38 (0.88) | 61 (1.37) |
| | | **MP** | 20 (0.10) | < 5 | < 5 | < 5 | < 5 | < 5 | 30 (0.69) | 22 (0.49) |
| | **31 to 90 days** | **VTE** | 89 (0.45) | 21 (0.35) | 36 (10.49) | < 5 | 7 (0.12) | < 5 | 104 (2.40) | 123 (2.76) |
| | | **DVT** | 50 (0.25) | 7 (0.12) | 22 (6.41) | < 5 | < 5 | < 5 | 60 (1.39) | 77 (1.73) |
| | | **PE** | 42 (0.21) | 14 (0.24) | 17 (4.95) | < 5 | < 5 | < 5 | 53 (1.22) | 55 (1.23) |
| | | **ATE** | 38 (0.19) | 36 (0.61) | 31 (9.03) | < 5 | 8 (0.14) | < 5 | 196 (4.53) | 307 (6.89) |
| | | **IS** | 7 (0.04) | 5 (0.08) | 21 (6.12) | < 5 | < 5 | < 5 | 95 (2.19) | 162 (3.64) |
| | | **TIA** | 11 (0.06) | 9 (0.15) | 6 (1.75) | < 5 | < 5 | < 5 | 53 (1.22) | 85 (1.91) |
| | | **MI** | 21 (0.11) | 22 (0.37) | 10 (2.91) | < 5 | < 5 | < 5 | 61 (1.41) | 79 (1.77) |
| | | **HF** | 50 (0.25) | 34 (0.57) | 143 (41.67) | 10 (24.59) | 5 (0.09) | < 5 | 260 (6.00) | 534 (11.98) |
| | | **HS** | 5 (0.03) | < 5 | < 5 | < 5 | < 5 | < 5 | 18 (0.42) | 19 (0.43) |
| | | **MP** | 14 (0.07) | < 5 | < 5 | < 5 | < 5 | < 5 | 8 (0.18) | < 5 |
| | **91 to 180 days** | **VTE** | 38 (0.19) | 9 (0.15) | 24 (6.99) | 5 (12.29) | 7 (0.12) | < 5 | 100 (2.31) | 84 (1.89) |
| | | **DVT** | 20 (0.10) | 6 (0.10) | 11 (3.21) | < 5 | < 5 | < 5 | 55 (1.27) | 52 (1.17) |
| | | **PE** | 18 (0.09) | < 5 | 14 (4.08) | < 5 | 7 (0.12) | < 5 | 51 (1.18) | 39 (0.88) |
| | | **ATE** | 30 (0.15) | 15 (0.25) | 28 (8.16) | < 5 | 5 (0.09) | < 5 | 171 (3.95) | 265 (5.95) |
| | | **IS** | < 5 | < 5 | 17 (4.95) | < 5 | < 5 | < 5 | 88 (2.03) | 148 (3.32) |
| | | **TIA** | 13 (0.07) | 5 (0.08) | 7 (2.04) | < 5 | < 5 | < 5 | 40 (0.92) | 72 (1.62) |
| | | **MI** | 16 (0.08) | 10 (0.17) | 7 (2.04) | < 5 | < 5 | < 5 | 49 (1.13) | 62 (1.39) |
| | | **HF** | 28 (0.14) | 24 (0.40) | 167 (48.66) | 13 (31.96) | < 5 | < 5 | 211 (4.87) | 438 (9.83) |
| | | **HS** | < 5 | < 5 | < 5 | < 5 | < 5 | < 5 | 18 (0.42) | 33 (0.74) |
| | | **MP** | < 5 | < 5 | < 5 | < 5 | < 5 | < 5 | 12 (0.28) | < 5 |
| | **181 to 365 days** | **VTE** | 17 (0.09) | 8 (0.13) | 48 (13.99) | < 5 | < 5 | < 5 | 30 (0.69) | 30 (0.67) |
| | | **DVT** | 6 (0.03) | < 5 | 25 (7.28) | < 5 | < 5 | < 5 | 18 (0.42) | 20 (0.45) |
| | | **PE** | 12 (0.06) | 7 (0.12) | 28 (8.16) | < 5 | < 5 | < 5 | 15 (0.35) | 12 (0.27) |
| | | **ATE** | 11 (0.06) | 7 (0.12) | 57 (16.61) | < 5 | < 5 | < 5 | 96 (2.22) | 103 (2.31) |
| | | **IS** | < 5 | < 5 | 36 (10.49) | < 5 | < 5 | < 5 | 51 (1.18) | 51 (1.14) |
| | | **TIA** | < 5 | < 5 | 15 (4.37) | < 5 | < 5 | < 5 | 22 (0.51) | 29 (0.65) |
| | | **MI** | 8 (0.04) | < 5 | 11 (3.21) | < 5 | < 5 | < 5 | 28 (0.65) | 29 (0.65) |
| | | **HF** | 22 (0.11) | 12 (0.20) | 267 (77.80) | 15 (36.88) | < 5 | < 5 | 90 (2.08) | 178 (3.99) |
| | | **HS** | < 5 | < 5 | < 5 | < 5 | < 5 | < 5 | 6 (0.14) | 9 (0.20) |
| | | **MP** | < 5 | < 5 | < 5 | < 5 | < 5 | < 5 | < 5 | < 5 |
| **Cohort 3** | | | **N = 1,510,323** | **N = 54,102** | **N = 96,471** | **N = 18,645** | **N = 416,549** | **N = 36,748** | **N = 869,109** | **N = 706,435** |
| | **0 to 30 days** | **VTE** | 267 (1.77) | < 5 | 124 (12.85) | 7 (3.75) | 33 (0.79) | < 5 | 422 (4.86) | 114 (1.61) |
| | | **DVT** | 52 (0.34) | < 5 | 25 (2.59) | 5 (2.68) | 7 (0.17) | < 5 | 132 (1.52) | 51 (0.72) |
| | | **PE** | 224 (1.48) | < 5 | 100 (10.37) | < 5 | 27 (0.65) | < 5 | 326 (3.75) | 71 (1.01) |
| | | **ATE** | 34 (0.23) | < 5 | 131 (13.58) | 8 (4.29) | < 5 | < 5 | 314 (3.61) | 197 (2.79) |
| | | **IS** | 5 (0.03) | < 5 | 73 (7.57) | 6 (3.22) | < 5 | < 5 | 154 (1.77) | 95 (1.34) |
| | | **TIA** | 6 (0.04) | < 5 | 24 (2.49) | < 5 | < 5 | < 5 | 32 (0.37) | 42 (0.59) |
| | | **MI** | 23 (0.15) | < 5 | 42 (4.35) | < 5 | < 5 | < 5 | 137 (1.58) | 78 (1.10) |
| | | **HF** | 32 (0.21) | 10 (1.85) | 401 (41.57) | 16 (8.58) | < 5 | < 5 | 476 (5.48) | 163 (2.31) |
| | | **HS** | < 5 | < 5 | 5 (0.52) | < 5 | < 5 | < 5 | 34 (0.39) | 26 (0.37) |
| | | **MP** | 14 (0.09) | < 5 | < 5 | < 5 | < 5 | < 5 | 27 (0.31) | 13 (0.18) |
| | **31 to 90 days** | **VTE** | 58 (0.38) | < 5 | 45 (4.66) | 5 (2.68) | < 5 | < 5 | 114 (1.31) | 63 (0.89) |

| Cohort | Time window | Outcome | AURUM | | CORIVA | | GOLD | | SIDIAP | |
|---|---|---|---|---|---|---|---|---|---|---|
| | | | Unvaccinated | Vaccinated | Unvaccinated | Vaccinated | Unvaccinated | Vaccinated | Unvaccinated | Vaccinated |
| | | DVT | 28 (0.19) | < 5 | 30 (3.11) | < 5 | < 5 | < 5 | 78 (0.90) | 47 (0.67) |
| | | PE | 33 (0.22) | < 5 | 18 (1.87) | < 5 | < 5 | < 5 | 47 (0.54) | 21 (0.30) |
| | | ATE | 14 (0.09) | < 5 | 42 (4.35) | 10 (5.36) | < 5 | < 5 | 169 (1.94) | 142 (2.01) |
| | | IS | < 5 | < 5 | 23 (2.38) | < 5 | < 5 | < 5 | 74 (0.85) | 54 (0.76) |
| | | TIA | < 5 | < 5 | 7 (0.73) | 6 (3.22) | < 5 | < 5 | 50 (0.58) | 27 (0.38) |
| | | MI | 11 (0.07) | < 5 | 16 (1.66) | < 5 | < 5 | < 5 | 57 (0.66) | 69 (0.98) |
| | | HF | 22 (0.15) | < 5 | 177 (18.35) | 11 (5.90) | < 5 | < 5 | 186 (2.14) | 108 (1.53) |
| | | HS | 5 (0.03) | < 5 | < 5 | < 5 | < 5 | < 5 | 13 (0.15) | 14 (0.20) |
| | | MP | 9 (0.06) | < 5 | 7 (0.73) | < 5 | < 5 | < 5 | 13 (0.15) | 9 (0.13) |
| | **91 to 180 days** | VTE | 28 (0.19) | < 5 | 44 (4.56) | 8 (4.29) | 5 (0.12) | < 5 | 92 (1.06) | 67 (0.95) |
| | | DVT | 14 (0.09) | < 5 | 27 (2.80) | 5 (2.68) | < 5 | < 5 | 56 (0.64) | 43 (0.61) |
| | | PE | 16 (0.11) | < 5 | 17 (1.76) | < 5 | < 5 | < 5 | 41 (0.47) | 25 (0.35) |
| | | ATE | 8 (0.05) | < 5 | 44 (4.56) | 7 (3.75) | < 5 | < 5 | 170 (1.96) | 137 (1.94) |
| | | IS | < 5 | < 5 | 20 (2.07) | < 5 | < 5 | < 5 | 77 (0.89) | 62 (0.88) |
| | | TIA | < 5 | < 5 | 13 (1.35) | < 5 | < 5 | < 5 | 30 (0.35) | 28 (0.40) |
| | | MI | < 5 | < 5 | 16 (1.66) | < 5 | < 5 | < 5 | 65 (0.75) | 53 (0.75) |
| | | HF | 11 (0.07) | < 5 | 210 (21.77) | 12 (6.44) | < 5 | < 5 | 169 (1.94) | 97 (1.37) |
| | | HS | < 5 | < 5 | < 5 | < 5 | < 5 | < 5 | 16 (0.18) | 10 (0.14) |
| | | MP | < 5 | < 5 | 6 (0.62) | < 5 | < 5 | < 5 | 11 (0.13) | 9 (0.13) |
| | **181 to 365 days** | VTE | < 5 | < 5 | 72 (7.46) | 13 (6.97) | < 5 | < 5 | 37 (0.43) | 13 (0.18) |
| | | DVT | < 5 | < 5 | 44 (4.56) | 11 (5.90) | < 5 | < 5 | 22 (0.25) | 11 (0.16) |
| | | PE | < 5 | < 5 | 31 (3.21) | < 5 | < 5 | < 5 | 18 (0.21) | < 5 |
| | | ATE | < 5 | < 5 | 80 (8.29) | 6 (3.22) | < 5 | < 5 | 54 (0.62) | 36 (0.51) |
| | | IS | < 5 | < 5 | 38 (3.94) | < 5 | < 5 | < 5 | 28 (0.32) | 13 (0.18) |
| | | TIA | < 5 | < 5 | 22 (2.28) | < 5 | < 5 | < 5 | 9 (0.10) | 5 (0.07) |
| | | MI | < 5 | < 5 | 27 (2.80) | < 5 | < 5 | < 5 | 18 (0.21) | 21 (0.30) |
| | | HF | 5 (0.03) | < 5 | 324 (33.59) | 25 (13.41) | < 5 | < 5 | 60 (0.69) | 25 (0.35) |
| | | HS | < 5 | < 5 | 6 (0.62) | < 5 | < 5 | < 5 | < 5 | < 5 |
| | | MP | < 5 | < 5 | 5 (0.52) | < 5 | < 5 | < 5 | < 5 | < 5 |
| **Cohort 4** | | | **N = 2,014,161** | **N = 1,335,671** | **N = 147,553** | **N = 15,683** | **N = 465,326** | **N = 365,096** | **N = 1,068,043** | **N = 580,329** |
| | **0 to 30 days** | VTE | 331 (1.64) | 20 (0.15) | 121 (8.20) | < 5 | 36 (0.77) | < 5 | 386 (3.61) | 66 (1.14) |
| | | DVT | 52 (0.26) | 9 (0.07) | 24 (1.63) | < 5 | 6 (0.13) | < 5 | 124 (1.16) | 35 (0.60) |
| | | PE | 290 (1.44) | 11 (0.08) | 98 (6.64) | < 5 | 31 (0.67) | < 5 | 298 (2.79) | 35 (0.60) |
| | | ATE | 25 (0.12) | < 5 | 122 (8.27) | 8 (5.10) | < 5 | < 5 | 277 (2.59) | 55 (0.95) |
| | | IS | < 5 | < 5 | 70 (4.74) | < 5 | < 5 | < 5 | 139 (1.30) | 29 (0.50) |
| | | TIA | 6 (0.03) | < 5 | 22 (1.49) | < 5 | < 5 | < 5 | 37 (0.35) | 7 (0.12) |
| | | MI | 15 (0.07) | < 5 | 38 (2.58) | < 5 | < 5 | < 5 | 108 (1.01) | 20 (0.34) |
| | | HF | 26 (0.13) | < 5 | 373 (25.28) | 13 (8.29) | < 5 | < 5 | 425 (3.98) | 44 (0.76) |
| | | HS | < 5 | < 5 | < 5 | < 5 | < 5 | < 5 | 32 (0.30) | 7 (0.12) |
| | | MP | 14 (0.07) | < 5 | 7 (0.47) | < 5 | < 5 | < 5 | 33 (0.31) | 11 (0.19) |
| | **31 to 90 days** | VTE | 54 (0.27) | 7 (0.05) | 45 (3.05) | < 5 | < 5 | < 5 | 109 (1.02) | 32 (0.55) |
| | | DVT | 20 (0.10) | < 5 | 30 (2.03) | < 5 | < 5 | < 5 | 76 (0.71) | 23 (0.40) |
| | | PE | 37 (0.18) | < 5 | 18 (1.22) | < 5 | < 5 | < 5 | 42 (0.39) | 10 (0.17) |
| | | ATE | 13 (0.06) | < 5 | 39 (2.64) | < 5 | < 5 | < 5 | 145 (1.36) | 51 (0.88) |
| | | IS | < 5 | < 5 | 22 (1.49) | < 5 | < 5 | < 5 | 64 (0.60) | 21 (0.36) |
| | | TIA | < 5 | < 5 | 7 (0.47) | < 5 | < 5 | < 5 | 41 (0.38) | 12 (0.21) |
| | | MI | 9 (0.04) | < 5 | 15 (1.02) | < 5 | < 5 | < 5 | 49 (0.46) | 21 (0.36) |
| | | HF | 14 (0.07) | 6 (0.04) | 160 (10.84) | 11 (7.01) | < 5 | < 5 | 155 (1.45) | 28 (0.48) |
| | | HS | 5 (0.02) | < 5 | < 5 | < 5 | < 5 | < 5 | 12 (0.11) | 5 (0.09) |

| Cohort | Time window | Outcome | AURUM | | CORIVA | | GOLD | | SIDIAP | |
|---|---|---|---|---|---|---|---|---|---|---|
| | | | Unvaccinated | Vaccinated | Unvaccinated | Vaccinated | Unvaccinated | Vaccinated | Unvaccinated | Vaccinated |
| | | **MP** | 8 (0.04) | < 5 | 8 (0.54) | < 5 | < 5 | < 5 | 14 (0.13) | 8 (0.14) |
| | **91 to 180 days** | **VTE** | 23 (0.11) | 9 (0.07) | 42 (2.85) | 5 (3.19) | < 5 | < 5 | 82 (0.77) | 40 (0.69) |
| | | **DVT** | 10 (0.05) | 7 (0.05) | 28 (1.90) | < 5 | < 5 | < 5 | 50 (0.47) | 25 (0.43) |
| | | **PE** | 14 (0.07) | < 5 | 15 (1.02) | < 5 | < 5 | < 5 | 37 (0.35) | 18 (0.31) |
| | | **ATE** | < 5 | < 5 | 32 (2.17) | 8 (5.10) | < 5 | < 5 | 139 (1.30) | 68 (1.17) |
| | | **IS** | < 5 | < 5 | 14 (0.95) | < 5 | < 5 | < 5 | 64 (0.60) | 34 (0.59) |
| | | **TIA** | < 5 | < 5 | 10 (0.68) | < 5 | < 5 | < 5 | 27 (0.25) | 14 (0.24) |
| | | **MI** | < 5 | < 5 | 12 (0.81) | < 5 | < 5 | < 5 | 50 (0.47) | 20 (0.34) |
| | | **HF** | 9 (0.04) | < 5 | 180 (12.20) | 9 (5.74) | < 5 | < 5 | 163 (1.53) | 35 (0.60) |
| | | **HS** | < 5 | < 5 | < 5 | < 5 | < 5 | < 5 | 19 (0.18) | < 5 |
| | | **MP** | < 5 | < 5 | 6 (0.41) | < 5 | < 5 | < 5 | 14 (0.13) | < 5 |
| | **181 to 365 days** | **VTE** | < 5 | < 5 | 72 (4.88) | 6 (3.83) | < 5 | < 5 | 44 (0.41) | 7 (0.12) |
| | | **DVT** | < 5 | < 5 | 45 (3.05) | 6 (3.83) | < 5 | < 5 | 28 (0.26) | 6 (0.10) |
| | | **PE** | < 5 | < 5 | 30 (2.03) | < 5 | < 5 | < 5 | 17 (0.16) | < 5 |
| | | **ATE** | < 5 | < 5 | 67 (4.54) | 6 (3.83) | < 5 | < 5 | 44 (0.41) | 9 (0.16) |
| | | **IS** | < 5 | < 5 | 32 (2.17) | < 5 | < 5 | < 5 | 21 (0.20) | 6 (0.10) |
| | | **TIA** | < 5 | < 5 | 19 (1.29) | < 5 | < 5 | < 5 | 8 (0.07) | < 5 |
| | | **MI** | < 5 | < 5 | 22 (1.49) | < 5 | < 5 | < 5 | 16 (0.15) | < 5 |
| | | **HF** | < 5 | < 5 | 288 (19.52) | 9 (5.74) | < 5 | < 5 | 53 (0.50) | 5 (0.09) |
| | | **HS** | < 5 | < 5 | 7 (0.47) | < 5 | < 5 | < 5 | 5 (0.05) | < 5 |
| | | **MP** | < 5 | < 5 | 7 (0.47) | < 5 | < 5 | < 5 | 6 (0.06) | < 5 |

VTE = Venous thromboembolism (DVT + PE), DVT = Deep vein thrombosis, PE = Pulmonary embolism, ATE = Arterial thromboembolism (IS + TIA + MI), IS = Ischemic stroke, TIA = Transient ischemic attack, MI = Myocardial infarction, HF = Heart failure, HS = Hemorrhagic stroke, MP = Myocarditis or Pericarditis

**Table S35: Number of records (and risk per 10,000 individuals) for post COVID-19 cardiac and thromboembolic complications,** across cohorts and databases, stratified by vaccine brand.

| Cohort | Time window | Outcome | AURUM | | GOLD | |
|---|---|---|---|---|---|---|
| | | | BNT162b2 | ChAdOx1 | BNT162b2 | ChAdOx1 |
| Cohort 1 | | | N = 332,790 | N = 219,804 | N = 32,755 | N = 82,406 |
| | 0 to 30 days | VTE | 76 (2.28) | 41 (1.87) | < 5 | < 5 |
| | | DVT | 15 (0.45) | 12 (0.55) | < 5 | < 5 |
| | | PE | 64 (1.92) | 31 (1.41) | < 5 | < 5 |
| | | ATE | 45 (1.35) | 25 (1.14) | < 5 | 6 (0.73) |
| | | IS | 5 (0.15) | < 5 | < 5 | < 5 |
| | | TIA | 11 (0.33) | 7 (0.32) | < 5 | < 5 |
| | | MI | 29 (0.87) | 17 (0.77) | < 5 | < 5 |
| | | HF | 126 (3.79) | 72 (3.28) | < 5 | 5 (0.61) |
| | | HS | < 5 | < 5 | < 5 | < 5 |
| | | MP | < 5 | < 5 | < 5 | < 5 |
| | 31 to 90 days | VTE | 26 (0.78) | 14 (0.64) | < 5 | < 5 |
| | | DVT | 7 (0.21) | 8 (0.36) | < 5 | < 5 |
| | | PE | 19 (0.57) | 7 (0.32) | < 5 | < 5 |
| | | ATE | 23 (0.69) | 20 (0.91) | < 5 | < 5 |
| | | IS | < 5 | < 5 | < 5 | < 5 |
| | | TIA | 10 (0.30) | 9 (0.41) | < 5 | < 5 |
| | | MI | 11 (0.33) | 9 (0.41) | < 5 | < 5 |
| | | HF | 71 (2.13) | 42 (1.91) | < 5 | 5 (0.61) |
| | | HS | < 5 | < 5 | < 5 | < 5 |
| | | MP | < 5 | < 5 | < 5 | < 5 |
| | 91 to 180 days | VTE | 6 (0.18) | 15 (0.68) | < 5 | < 5 |
| | | DVT | < 5 | 5 (0.23) | < 5 | < 5 |
| | | PE | < 5 | 10 (0.45) | < 5 | < 5 |
| | | ATE | 18 (0.54) | 10 (0.45) | < 5 | 6 (0.73) |
| | | IS | 6 (0.18) | < 5 | < 5 | < 5 |
| | | TIA | 6 (0.18) | 6 (0.27) | < 5 | < 5 |
| | | MI | 6 (0.18) | < 5 | < 5 | 5 (0.61) |
| | | HF | 57 (1.71) | 38 (1.73) | < 5 | < 5 |
| | | HS | < 5 | < 5 | < 5 | < 5 |
| | | MP | < 5 | < 5 | < 5 | < 5 |
| | 181 to 365 days | VTE | 10 (0.30) | < 5 | < 5 | < 5 |
| | | DVT | 6 (0.18) | < 5 | < 5 | < 5 |
| | | PE | < 5 | < 5 | < 5 | < 5 |
| | | ATE | 14 (0.42) | 9 (0.41) | < 5 | < 5 |
| | | IS | < 5 | < 5 | < 5 | < 5 |
| | | TIA | < 5 | < 5 | < 5 | < 5 |
| | | MI | 7 (0.21) | < 5 | < 5 | < 5 |
| | | HF | 38 (1.14) | 20 (0.91) | < 5 | 5 (0.61) |
| | | HS | < 5 | < 5 | < 5 | < 5 |
| | | MP | < 5 | < 5 | < 5 | < 5 |
| Cohort 2 | | | N = 594,262 | N = 969,262 | N = 180,670 | N = 302,999 |
| | 0 to 30 days | VTE | 62 (1.04) | 158 (1.63) | 5 (0.28) | 19 (0.63) |
| | | DVT | 18 (0.30) | 47 (0.48) | < 5 | < 5 |
| | | PE | 45 (0.76) | 120 (1.24) | < 5 | 15 (0.50) |
| | | ATE | 29 (0.49) | 75 (0.77) | < 5 | < 5 |

| Cohort | Time window | Outcome | AURUM | | GOLD | |
|---|---|---|---|---|---|---|
| | | | BNT162b2 | ChAdOx1 | BNT162b2 | ChAdOx1 |
| | | **IS** | < 5 | 12 (0.12) | < 5 | < 5 |
| | | **TIA** | 11 (0.19) | 13 (0.13) | < 5 | < 5 |
| | | **MI** | 16 (0.27) | 52 (0.54) | < 5 | < 5 |
| | | **HF** | 43 (0.72) | 103 (1.06) | < 5 | 11 (0.36) |
| | | **HS** | < 5 | < 5 | < 5 | < 5 |
| | | **MP** | < 5 | 6 (0.06) | < 5 | < 5 |
| | **31 to 90 days** | **VTE** | 21 (0.35) | 55 (0.57) | < 5 | 9 (0.30) |
| | | **DVT** | 7 (0.12) | 25 (0.26) | < 5 | 5 (0.17) |
| | | **PE** | 14 (0.24) | 32 (0.33) | < 5 | < 5 |
| | | **ATE** | 36 (0.61) | 57 (0.59) | < 5 | 5 (0.17) |
| | | **IS** | 5 (0.08) | 6 (0.06) | < 5 | < 5 |
| | | **TIA** | 9 (0.15) | 25 (0.26) | < 5 | < 5 |
| | | **MI** | 22 (0.37) | 28 (0.29) | < 5 | < 5 |
| | | **HF** | 35 (0.59) | 68 (0.70) | < 5 | 7 (0.23) |
| | | **HS** | < 5 | < 5 | < 5 | < 5 |
| | | **MP** | < 5 | < 5 | < 5 | < 5 |
| | **91 to 180 days** | **VTE** | 10 (0.17) | 30 (0.31) | < 5 | < 5 |
| | | **DVT** | 6 (0.10) | 16 (0.17) | < 5 | < 5 |
| | | **PE** | 5 (0.08) | 16 (0.17) | < 5 | < 5 |
| | | **ATE** | 15 (0.25) | 28 (0.29) | < 5 | < 5 |
| | | **IS** | < 5 | < 5 | < 5 | < 5 |
| | | **TIA** | 5 (0.08) | 13 (0.13) | < 5 | < 5 |
| | | **MI** | 10 (0.17) | 15 (0.15) | < 5 | < 5 |
| | | **HF** | 25 (0.42) | 44 (0.45) | < 5 | < 5 |
| | | **HS** | < 5 | < 5 | < 5 | < 5 |
| | | **MP** | < 5 | < 5 | < 5 | < 5 |
| | **181 to 365 days** | **VTE** | 8 (0.13) | 5 (0.05) | < 5 | < 5 |
| | | **DVT** | < 5 | < 5 | < 5 | < 5 |
| | | **PE** | 7 (0.12) | < 5 | < 5 | < 5 |
| | | **ATE** | 7 (0.12) | 11 (0.11) | < 5 | < 5 |
| | | **IS** | < 5 | < 5 | < 5 | < 5 |
| | | **TIA** | < 5 | < 5 | < 5 | < 5 |
| | | **MI** | < 5 | 7 (0.07) | < 5 | < 5 |
| | | **HF** | 12 (0.20) | 23 (0.24) | < 5 | < 5 |
| | | **HS** | < 5 | < 5 | < 5 | < 5 |
| | | **MP** | < 5 | < 5 | < 5 | < 5 |
| **Cohort 3** | | | **N = 54,102** | **N = 1,473,602** | **N = 36,748** | **N = 423,876** |
| | **0 to 30 days** | **VTE** | < 5 | 139 (0.94) | < 5 | 16 (0.38) |
| | | **DVT** | < 5 | 41 (0.28) | < 5 | < 5 |
| | | **PE** | < 5 | 101 (0.69) | < 5 | 12 (0.28) |
| | | **ATE** | < 5 | 47 (0.32) | < 5 | 12 (0.28) |
| | | **IS** | < 5 | 5 (0.03) | < 5 | < 5 |
| | | **TIA** | < 5 | 16 (0.11) | < 5 | < 5 |
| | | **MI** | < 5 | 26 (0.18) | < 5 | 9 (0.21) |
| | | **HF** | 10 (1.85) | 28 (0.19) | < 5 | < 5 |
| | | **HS** | < 5 | < 5 | < 5 | < 5 |
| | | **MP** | < 5 | 6 (0.04) | < 5 | < 5 |
| | **31 to 90 days** | **VTE** | < 5 | 44 (0.30) | < 5 | 7 (0.17) |
| | | **DVT** | < 5 | 26 (0.18) | < 5 | < 5 |

| Cohort | Time window | Outcome | AURUM | | GOLD | |
|---|---|---|---|---|---|---|
| | | | BNT162b2 | ChAdOx1 | BNT162b2 | ChAdOx1 |
| | | PE | < 5 | 18 (0.12) | < 5 | < 5 |
| | | ATE | < 5 | 31 (0.21) | < 5 | 7 (0.17) |
| | | IS | < 5 | < 5 | < 5 | < 5 |
| | | TIA | < 5 | 13 (0.09) | < 5 | < 5 |
| | | MI | < 5 | 16 (0.11) | < 5 | < 5 |
| | | HF | < 5 | 23 (0.16) | < 5 | < 5 |
| | | HS | < 5 | < 5 | < 5 | < 5 |
| | | MP | < 5 | 7 (0.05) | < 5 | < 5 |
| | 91 to 180 days | VTE | < 5 | 26 (0.18) | < 5 | < 5 |
| | | DVT | < 5 | 15 (0.10) | < 5 | < 5 |
| | | PE | < 5 | 14 (0.10) | < 5 | < 5 |
| | | ATE | < 5 | 26 (0.18) | < 5 | < 5 |
| | | IS | < 5 | < 5 | < 5 | < 5 |
| | | TIA | < 5 | 8 (0.05) | < 5 | < 5 |
| | | MI | < 5 | 17 (0.12) | < 5 | < 5 |
| | | HF | < 5 | 12 (0.08) | < 5 | < 5 |
| | | HS | < 5 | < 5 | < 5 | < 5 |
| | | MP | < 5 | < 5 | < 5 | < 5 |
| | 181 to 365 days | VTE | < 5 | 11 (0.07) | < 5 | < 5 |
| | | DVT | < 5 | 5 (0.03) | < 5 | < 5 |
| | | PE | < 5 | 7 (0.05) | < 5 | < 5 |
| | | ATE | < 5 | < 5 | < 5 | < 5 |
| | | IS | < 5 | < 5 | < 5 | < 5 |
| | | TIA | < 5 | < 5 | < 5 | < 5 |
| | | MI | < 5 | < 5 | < 5 | < 5 |
| | | HF | < 5 | < 5 | < 5 | < 5 |
| | | HS | < 5 | < 5 | < 5 | < 5 |
| | | MP | < 5 | < 5 | < 5 | < 5 |
| Cohort 4 | | | N = 1,335,671 | N = 542,670 | N = 365,096 | N = 147,744 |
| | 0 to 30 days | VTE | 20 (0.15) | 27 (0.50) | < 5 | 8 (0.54) |
| | | DVT | 9 (0.07) | 7 (0.13) | < 5 | < 5 |
| | | PE | 11 (0.08) | 21 (0.39) | < 5 | 6 (0.41) |
| | | ATE | < 5 | 5 (0.09) | < 5 | < 5 |
| | | IS | < 5 | < 5 | < 5 | < 5 |
| | | TIA | < 5 | < 5 | < 5 | < 5 |
| | | MI | < 5 | < 5 | < 5 | < 5 |
| | | HF | < 5 | < 5 | < 5 | < 5 |
| | | HS | < 5 | < 5 | < 5 | < 5 |
| | | MP | < 5 | < 5 | < 5 | < 5 |
| | 31 to 90 days | VTE | 7 (0.05) | 14 (0.26) | < 5 | < 5 |
| | | DVT | < 5 | 8 (0.15) | < 5 | < 5 |
| | | PE | < 5 | 6 (0.11) | < 5 | < 5 |
| | | ATE | < 5 | < 5 | < 5 | < 5 |
| | | IS | < 5 | < 5 | < 5 | < 5 |
| | | TIA | < 5 | < 5 | < 5 | < 5 |
| | | MI | < 5 | < 5 | < 5 | < 5 |
| | | HF | 6 (0.04) | < 5 | < 5 | < 5 |
| | | HS | < 5 | < 5 | < 5 | < 5 |
| | | MP | < 5 | < 5 | < 5 | < 5 |

| Cohort | Time window | Outcome | AURUM | | GOLD | |
|---|---|---|---|---|---|---|
| | | | BNT162b2 | ChAdOx1 | BNT162b2 | ChAdOx1 |
| | 91 to 180 days | VTE | 9 (0.07) | < 5 | < 5 | < 5 |
| | | DVT | 7 (0.05) | < 5 | < 5 | < 5 |
| | | PE | < 5 | < 5 | < 5 | < 5 |
| | | ATE | < 5 | < 5 | < 5 | < 5 |
| | | IS | < 5 | < 5 | < 5 | < 5 |
| | | TIA | < 5 | < 5 | < 5 | < 5 |
| | | MI | < 5 | < 5 | < 5 | < 5 |
| | | HF | < 5 | < 5 | < 5 | < 5 |
| | | HS | < 5 | < 5 | < 5 | < 5 |
| | | MP | < 5 | < 5 | < 5 | < 5 |
| | 181 to 365 days | VTE | < 5 | < 5 | < 5 | < 5 |
| | | DVT | < 5 | < 5 | < 5 | < 5 |
| | | PE | < 5 | < 5 | < 5 | < 5 |
| | | ATE | < 5 | < 5 | < 5 | < 5 |
| | | IS | < 5 | < 5 | < 5 | < 5 |
| | | TIA | < 5 | < 5 | < 5 | < 5 |
| | | MI | < 5 | < 5 | < 5 | < 5 |
| | | HF | < 5 | < 5 | < 5 | < 5 |
| | | HS | < 5 | < 5 | < 5 | < 5 |
| | | MP | < 5 | < 5 | < 5 | < 5 |

VTE = Venous thromboembolism (DVT + PE), DVT = Deep vein thrombosis, PE = Pulmonary embolism, ATE = Arterial thromboembolism (IS + TIA + MI), IS = Ischemic stroke, TIA = Transient ischemic attack, MI = Myocardial infarction, HF = Heart failure, HS = Hemorrhagic stroke, MP = Myocarditis or Pericarditis

**Table S36: Number of records (and risk per 10,000 individuals) for post COVID-19 cardiac and thromboembolic complications,** across cohorts and databases, stratified by vaccine brand. Follow-up ends at first vaccine dose after index date.

| Cohort | Time window | Outcome | AURUM | | GOLD | |
|---|---|---|---|---|---|---|
| | | | BNT162b2 | ChAdOx1 | BNT162b2 | ChAdOx1 |
| **Cohort 1** | | | **N = 332,790** | **N = 219,804** | **N = 32,755** | **N = 82,406** |
| | **0 to 30 days** | **VTE** | 31 (0.93) | 14 (0.64) | < 5 | < 5 |
| | | **DVT** | 8 (0.24) | < 5 | < 5 | < 5 |
| | | **PE** | 24 (0.72) | 13 (0.59) | < 5 | < 5 |
| | | **ATE** | 18 (0.54) | 10 (0.45) | < 5 | < 5 |
| | | **IS** | < 5 | < 5 | < 5 | < 5 |
| | | **TIA** | < 5 | < 5 | < 5 | < 5 |
| | | **MI** | 14 (0.42) | 6 (0.27) | < 5 | < 5 |
| | | **HF** | 45 (1.35) | 28 (1.27) | < 5 | < 5 |
| | | **HS** | < 5 | < 5 | < 5 | < 5 |
| | | **MP** | < 5 | < 5 | < 5 | < 5 |
| | **31 to 90 days** | **VTE** | 15 (0.45) | 5 (0.23) | < 5 | < 5 |
| | | **DVT** | < 5 | < 5 | < 5 | < 5 |
| | | **PE** | 11 (0.33) | < 5 | < 5 | < 5 |
| | | **ATE** | 8 (0.24) | 8 (0.36) | < 5 | < 5 |
| | | **IS** | < 5 | < 5 | < 5 | < 5 |
| | | **TIA** | < 5 | < 5 | < 5 | < 5 |
| | | **MI** | < 5 | < 5 | < 5 | < 5 |
| | | **HF** | 36 (1.08) | 20 (0.91) | < 5 | < 5 |
| | | **HS** | < 5 | < 5 | < 5 | < 5 |
| | | **MP** | < 5 | < 5 | < 5 | < 5 |
| | **91 to 180 days** | **VTE** | < 5 | 5 (0.23) | < 5 | < 5 |
| | | **DVT** | < 5 | < 5 | < 5 | < 5 |
| | | **PE** | < 5 | < 5 | < 5 | < 5 |
| | | **ATE** | 10 (0.30) | < 5 | < 5 | < 5 |
| | | **IS** | < 5 | < 5 | < 5 | < 5 |
| | | **TIA** | < 5 | < 5 | < 5 | < 5 |
| | | **MI** | < 5 | < 5 | < 5 | < 5 |
| | | **HF** | 32 (0.96) | 19 (0.86) | < 5 | < 5 |
| | | **HS** | < 5 | < 5 | < 5 | < 5 |
| | | **MP** | < 5 | < 5 | < 5 | < 5 |
| | **181 to 365 days** | **VTE** | 7 (0.21) | < 5 | < 5 | < 5 |
| | | **DVT** | < 5 | < 5 | < 5 | < 5 |
| | | **PE** | < 5 | < 5 | < 5 | < 5 |
| | | **ATE** | 11 (0.33) | 8 (0.36) | < 5 | < 5 |
| | | **IS** | < 5 | < 5 | < 5 | < 5 |
| | | **TIA** | < 5 | < 5 | < 5 | < 5 |
| | | **MI** | 6 (0.18) | < 5 | < 5 | < 5 |
| | | **HF** | 35 (1.05) | 18 (0.82) | < 5 | 5 (0.61) |
| | | **HS** | < 5 | < 5 | < 5 | < 5 |
| | | **MP** | < 5 | < 5 | < 5 | < 5 |
| **Cohort 2** | | | **N = 594,262** | **N = 969,262** | **N = 180,670** | **N = 302,999** |
| | **0 to 30 days** | **VTE** | 14 (0.24) | 40 (0.41) | < 5 | < 5 |
| | | **DVT** | 6 (0.10) | 13 (0.13) | < 5 | < 5 |
| | | **PE** | 8 (0.13) | 30 (0.31) | < 5 | < 5 |

| Cohort | Time window | Outcome | AURUM | | GOLD | |
|---|---|---|---|---|---|---|
| | | | BNT162b2 | ChAdOx1 | BNT162b2 | ChAdOx1 |
| | | ATE | 14 (0.24) | 11 (0.11) | < 5 | < 5 |
| | | IS | < 5 | < 5 | < 5 | < 5 |
| | | TIA | < 5 | < 5 | < 5 | < 5 |
| | | MI | 8 (0.13) | 9 (0.09) | < 5 | < 5 |
| | | HF | 15 (0.25) | 32 (0.33) | < 5 | < 5 |
| | | HS | < 5 | < 5 | < 5 | < 5 |
| | | MP | < 5 | < 5 | < 5 | < 5 |
| | 31 to 90 days | VTE | 5 (0.08) | 8 (0.08) | < 5 | < 5 |
| | | DVT | < 5 | 5 (0.05) | < 5 | < 5 |
| | | PE | < 5 | < 5 | < 5 | < 5 |
| | | ATE | 9 (0.15) | 9 (0.09) | < 5 | < 5 |
| | | IS | < 5 | < 5 | < 5 | < 5 |
| | | TIA | < 5 | < 5 | < 5 | < 5 |
| | | MI | < 5 | 6 (0.06) | < 5 | < 5 |
| | | HF | 17 (0.29) | 13 (0.13) | < 5 | < 5 |
| | | HS | < 5 | < 5 | < 5 | < 5 |
| | | MP | < 5 | < 5 | < 5 | < 5 |
| | 91 to 180 days | VTE | < 5 | 12 (0.12) | < 5 | < 5 |
| | | DVT | < 5 | 7 (0.07) | < 5 | < 5 |
| | | PE | < 5 | 5 (0.05) | < 5 | < 5 |
| | | ATE | 5 (0.08) | 10 (0.10) | < 5 | < 5 |
| | | IS | < 5 | < 5 | < 5 | < 5 |
| | | TIA | < 5 | < 5 | < 5 | < 5 |
| | | MI | < 5 | 6 (0.06) | < 5 | < 5 |
| | | HF | 9 (0.15) | 18 (0.19) | < 5 | < 5 |
| | | HS | < 5 | < 5 | < 5 | < 5 |
| | | MP | < 5 | < 5 | < 5 | < 5 |
| | 181 to 365 days | VTE | 7 (0.12) | 5 (0.05) | < 5 | < 5 |
| | | DVT | < 5 | < 5 | < 5 | < 5 |
| | | PE | 5 (0.08) | < 5 | < 5 | < 5 |
| | | ATE | 6 (0.10) | 10 (0.10) | < 5 | < 5 |
| | | IS | < 5 | < 5 | < 5 | < 5 |
| | | TIA | < 5 | < 5 | < 5 | < 5 |
| | | MI | < 5 | 6 (0.06) | < 5 | < 5 |
| | | HF | 11 (0.19) | 20 (0.21) | < 5 | < 5 |
| | | HS | < 5 | < 5 | < 5 | < 5 |
| | | MP | < 5 | < 5 | < 5 | < 5 |
| Cohort 3 | | | N = 54,102 | N = 1,473,602 | N = 36,748 | N = 423,876 |
| | 0 to 30 days | VTE | < 5 | 23 (0.16) | < 5 | < 5 |
| | | DVT | < 5 | < 5 | < 5 | < 5 |
| | | PE | < 5 | 19 (0.13) | < 5 | < 5 |
| | | ATE | < 5 | 8 (0.05) | < 5 | < 5 |
| | | IS | < 5 | < 5 | < 5 | < 5 |
| | | TIA | < 5 | < 5 | < 5 | < 5 |
| | | MI | < 5 | 7 (0.05) | < 5 | < 5 |
| | | HF | 7 (1.29) | 11 (0.07) | < 5 | < 5 |
| | | HS | < 5 | < 5 | < 5 | < 5 |
| | | MP | < 5 | < 5 | < 5 | < 5 |
| | 31 to 90 days | VTE | < 5 | < 5 | < 5 | < 5 |

| Cohort | Time window | Outcome | AURUM | | GOLD | |
|---|---|---|---|---|---|---|
| | | | BNT162b2 | ChAdOx1 | BNT162b2 | ChAdOx1 |
| | | DVT | < 5 | < 5 | < 5 | < 5 |
| | | PE | < 5 | < 5 | < 5 | < 5 |
| | | ATE | < 5 | 6 (0.04) | < 5 | < 5 |
| | | IS | < 5 | < 5 | < 5 | < 5 |
| | | TIA | < 5 | < 5 | < 5 | < 5 |
| | | MI | < 5 | < 5 | < 5 | < 5 |
| | | HF | < 5 | 5 (0.03) | < 5 | < 5 |
| | | HS | < 5 | < 5 | < 5 | < 5 |
| | | MP | < 5 | < 5 | < 5 | < 5 |
| | 91 to 180 days | VTE | < 5 | 8 (0.05) | < 5 | < 5 |
| | | DVT | < 5 | 6 (0.04) | < 5 | < 5 |
| | | PE | < 5 | < 5 | < 5 | < 5 |
| | | ATE | < 5 | 7 (0.05) | < 5 | < 5 |
| | | IS | < 5 | < 5 | < 5 | < 5 |
| | | TIA | < 5 | < 5 | < 5 | < 5 |
| | | MI | < 5 | < 5 | < 5 | < 5 |
| | | HF | < 5 | < 5 | < 5 | < 5 |
| | | HS | < 5 | < 5 | < 5 | < 5 |
| | | MP | < 5 | < 5 | < 5 | < 5 |
| | 181 to 365 days | VTE | < 5 | 9 (0.06) | < 5 | < 5 |
| | | DVT | < 5 | < 5 | < 5 | < 5 |
| | | PE | < 5 | 6 (0.04) | < 5 | < 5 |
| | | ATE | < 5 | < 5 | < 5 | < 5 |
| | | IS | < 5 | < 5 | < 5 | < 5 |
| | | TIA | < 5 | < 5 | < 5 | < 5 |
| | | MI | < 5 | < 5 | < 5 | < 5 |
| | | HF | < 5 | < 5 | < 5 | < 5 |
| | | HS | < 5 | < 5 | < 5 | < 5 |
| | | MP | < 5 | < 5 | < 5 | < 5 |
| Cohort 4 | | | N = 1,335,671 | N = 542,670 | N = 365,096 | N = 147,744 |
| | 0 to 30 days | VTE | 9 (0.07) | 8 (0.15) | < 5 | 6 (0.41) |
| | | DVT | 5 (0.04) | < 5 | < 5 | < 5 |
| | | PE | < 5 | 7 (0.13) | < 5 | 6 (0.41) |
| | | ATE | < 5 | < 5 | < 5 | < 5 |
| | | IS | < 5 | < 5 | < 5 | < 5 |
| | | TIA | < 5 | < 5 | < 5 | < 5 |
| | | MI | < 5 | < 5 | < 5 | < 5 |
| | | HF | < 5 | < 5 | < 5 | < 5 |
| | | HS | < 5 | < 5 | < 5 | < 5 |
| | | MP | < 5 | < 5 | < 5 | < 5 |
| | 31 to 90 days | VTE | < 5 | 8 (0.15) | < 5 | < 5 |
| | | DVT | < 5 | < 5 | < 5 | < 5 |
| | | PE | < 5 | 5 (0.09) | < 5 | < 5 |
| | | ATE | < 5 | < 5 | < 5 | < 5 |
| | | IS | < 5 | < 5 | < 5 | < 5 |
| | | TIA | < 5 | < 5 | < 5 | < 5 |
| | | MI | < 5 | < 5 | < 5 | < 5 |
| | | HF | 6 (0.04) | < 5 | < 5 | < 5 |
| | | HS | < 5 | < 5 | < 5 | < 5 |

| Cohort | Time window | Outcome | AURUM | | GOLD | |
|---|---|---|---|---|---|---|
| | | | BNT162b2 | ChAdOx1 | BNT162b2 | ChAdOx1 |
| | | MP | < 5 | < 5 | < 5 | < 5 |
| | 91 to 180 days | VTE | 8 (0.06) | < 5 | < 5 | < 5 |
| | | DVT | 7 (0.05) | < 5 | < 5 | < 5 |
| | | PE | < 5 | < 5 | < 5 | < 5 |
| | | ATE | < 5 | < 5 | < 5 | < 5 |
| | | IS | < 5 | < 5 | < 5 | < 5 |
| | | TIA | < 5 | < 5 | < 5 | < 5 |
| | | MI | < 5 | < 5 | < 5 | < 5 |
| | | HF | < 5 | < 5 | < 5 | < 5 |
| | | HS | < 5 | < 5 | < 5 | < 5 |
| | | MP | < 5 | < 5 | < 5 | < 5 |
| | 181 to 365 days | VTE | < 5 | < 5 | < 5 | < 5 |
| | | DVT | < 5 | < 5 | < 5 | < 5 |
| | | PE | < 5 | < 5 | < 5 | < 5 |
| | | ATE | < 5 | < 5 | < 5 | < 5 |
| | | IS | < 5 | < 5 | < 5 | < 5 |
| | | TIA | < 5 | < 5 | < 5 | < 5 |
| | | MI | < 5 | < 5 | < 5 | < 5 |
| | | HF | < 5 | < 5 | < 5 | < 5 |
| | | HS | < 5 | < 5 | < 5 | < 5 |
| | | MP | < 5 | < 5 | < 5 | < 5 |

VTE = Venous thromboembolism (DVT + PE), DVT = Deep vein thrombosis, PE = Pulmonary embolism, ATE = Arterial thromboembolism (IS + TIA + MI), IS = Ischemic stroke, TIA = Transient ischemic attack, MI = Myocardial infarction, HF = Heart failure, HS = Hemorrhagic stroke, MP = Myocarditis or Pericarditis

**Table S37: Number of records (and risk per 10,000 individuals) for post COVID-19 cardiac and thromboembolic complications,** across cohorts and databases, stratified by vaccine brand. Only first outcome after COVID-19 captured.

| Cohort | Time window | Outcome | AURUM | | GOLD | |
|---|---|---|---|---|---|---|
| | | | BNT162b2 | ChAdOx1 | BNT162b2 | ChAdOx1 |
| Cohort 1 | | | N = 332,790 | N = 219,804 | N = 32,755 | N = 82,406 |
| | 0 to 30 days | VTE | 76 (2.28) | 41 (1.87) | < 5 | < 5 |
| | | DVT | 15 (0.45) | 12 (0.55) | < 5 | < 5 |
| | | PE | 64 (1.92) | 31 (1.41) | < 5 | < 5 |
| | | ATE | 45 (1.35) | 25 (1.14) | < 5 | 6 (0.73) |
| | | IS | 5 (0.15) | < 5 | < 5 | < 5 |
| | | TIA | 11 (0.33) | 7 (0.32) | < 5 | < 5 |
| | | MI | 29 (0.87) | 17 (0.77) | < 5 | < 5 |
| | | HF | 126 (3.79) | 72 (3.28) | < 5 | 5 (0.61) |
| | | HS | < 5 | < 5 | < 5 | < 5 |
| | | MP | < 5 | < 5 | < 5 | < 5 |
| | 31 to 90 days | VTE | 25 (0.75) | 14 (0.64) | < 5 | < 5 |
| | | DVT | 7 (0.21) | 8 (0.36) | < 5 | < 5 |
| | | PE | 18 (0.54) | 7 (0.32) | < 5 | < 5 |
| | | ATE | 23 (0.69) | 20 (0.91) | < 5 | < 5 |
| | | IS | < 5 | < 5 | < 5 | < 5 |
| | | TIA | 10 (0.30) | 9 (0.41) | < 5 | < 5 |
| | | MI | 11 (0.33) | 9 (0.41) | < 5 | < 5 |
| | | HF | 70 (2.10) | 39 (1.77) | < 5 | 5 (0.61) |
| | | HS | < 5 | < 5 | < 5 | < 5 |
| | | MP | < 5 | < 5 | < 5 | < 5 |
| | 91 to 180 days | VTE | < 5 | 15 (0.68) | < 5 | < 5 |
| | | DVT | < 5 | 5 (0.23) | < 5 | < 5 |
| | | PE | < 5 | 10 (0.45) | < 5 | < 5 |
| | | ATE | 18 (0.54) | 10 (0.45) | < 5 | 6 (0.73) |
| | | IS | 6 (0.18) | < 5 | < 5 | < 5 |
| | | TIA | 6 (0.18) | 6 (0.27) | < 5 | < 5 |
| | | MI | 6 (0.18) | < 5 | < 5 | 5 (0.61) |
| | | HF | 54 (1.62) | 35 (1.59) | < 5 | < 5 |
| | | HS | < 5 | < 5 | < 5 | < 5 |
| | | MP | < 5 | < 5 | < 5 | < 5 |
| | 181 to 365 days | VTE | 10 (0.30) | < 5 | < 5 | < 5 |
| | | DVT | 6 (0.18) | < 5 | < 5 | < 5 |
| | | PE | < 5 | < 5 | < 5 | < 5 |
| | | ATE | 14 (0.42) | 9 (0.41) | < 5 | < 5 |
| | | IS | < 5 | < 5 | < 5 | < 5 |
| | | TIA | < 5 | < 5 | < 5 | < 5 |
| | | MI | 7 (0.21) | < 5 | < 5 | < 5 |
| | | HF | 35 (1.05) | 18 (0.82) | < 5 | 5 (0.61) |
| | | HS | < 5 | < 5 | < 5 | < 5 |
| | | MP | < 5 | < 5 | < 5 | < 5 |
| Cohort 2 | | | N = 594,262 | N = 969,262 | N = 180,670 | N = 302,999 |
| | 0 to 30 days | VTE | 62 (1.04) | 158 (1.63) | 5 (0.28) | 19 (0.63) |
| | | DVT | 18 (0.30) | 47 (0.48) | < 5 | < 5 |
| | | PE | 45 (0.76) | 120 (1.24) | < 5 | 15 (0.50) |

| Cohort | Time window | Outcome | AURUM | | GOLD | |
|---|---|---|---|---|---|---|
| | | | BNT162b2 | ChAdOx1 | BNT162b2 | ChAdOx1 |
| | | ATE | 29 (0.49) | 75 (0.77) | < 5 | < 5 |
| | | IS | < 5 | 12 (0.12) | < 5 | < 5 |
| | | TIA | 11 (0.19) | 13 (0.13) | < 5 | < 5 |
| | | MI | 16 (0.27) | 52 (0.54) | < 5 | < 5 |
| | | HF | 43 (0.72) | 103 (1.06) | < 5 | 11 (0.36) |
| | | HS | < 5 | < 5 | < 5 | < 5 |
| | | MP | < 5 | 6 (0.06) | < 5 | < 5 |
| | 31 to 90 days | VTE | 21 (0.35) | 54 (0.56) | < 5 | 9 (0.30) |
| | | DVT | 7 (0.12) | 25 (0.26) | < 5 | 5 (0.17) |
| | | PE | 14 (0.24) | 31 (0.32) | < 5 | < 5 |
| | | ATE | 36 (0.61) | 56 (0.58) | < 5 | 5 (0.17) |
| | | IS | 5 (0.08) | 6 (0.06) | < 5 | < 5 |
| | | TIA | 9 (0.15) | 24 (0.25) | < 5 | < 5 |
| | | MI | 22 (0.37) | 28 (0.29) | < 5 | < 5 |
| | | HF | 34 (0.57) | 65 (0.67) | < 5 | 7 (0.23) |
| | | HS | < 5 | < 5 | < 5 | < 5 |
| | | MP | < 5 | < 5 | < 5 | < 5 |
| | 91 to 180 days | VTE | 9 (0.15) | 28 (0.29) | < 5 | < 5 |
| | | DVT | 6 (0.10) | 15 (0.15) | < 5 | < 5 |
| | | PE | < 5 | 15 (0.15) | < 5 | < 5 |
| | | ATE | 15 (0.25) | 28 (0.29) | < 5 | < 5 |
| | | IS | < 5 | < 5 | < 5 | < 5 |
| | | TIA | 5 (0.08) | 13 (0.13) | < 5 | < 5 |
| | | MI | 10 (0.17) | 15 (0.15) | < 5 | < 5 |
| | | HF | 24 (0.40) | 41 (0.42) | < 5 | < 5 |
| | | HS | < 5 | < 5 | < 5 | < 5 |
| | | MP | < 5 | < 5 | < 5 | < 5 |
| | 181 to 365 days | VTE | 8 (0.13) | < 5 | < 5 | < 5 |
| | | DVT | < 5 | < 5 | < 5 | < 5 |
| | | PE | 7 (0.12) | < 5 | < 5 | < 5 |
| | | ATE | 7 (0.12) | 10 (0.10) | < 5 | < 5 |
| | | IS | < 5 | < 5 | < 5 | < 5 |
| | | TIA | < 5 | < 5 | < 5 | < 5 |
| | | MI | < 5 | 6 (0.06) | < 5 | < 5 |
| | | HF | 12 (0.20) | 22 (0.23) | < 5 | < 5 |
| | | HS | < 5 | < 5 | < 5 | < 5 |
| | | MP | < 5 | < 5 | < 5 | < 5 |
| Cohort 3 | | | N = 54,102 | N = 1,473,602 | N = 36,748 | N = 423,876 |
| | 0 to 30 days | VTE | < 5 | 139 (0.94) | < 5 | 16 (0.38) |
| | | DVT | < 5 | 41 (0.28) | < 5 | < 5 |
| | | PE | < 5 | 101 (0.69) | < 5 | 12 (0.28) |
| | | ATE | < 5 | 47 (0.32) | < 5 | 12 (0.28) |
| | | IS | < 5 | 5 (0.03) | < 5 | < 5 |
| | | TIA | < 5 | 16 (0.11) | < 5 | < 5 |
| | | MI | < 5 | 26 (0.18) | < 5 | 9 (0.21) |
| | | HF | 10 (1.85) | 28 (0.19) | < 5 | < 5 |
| | | HS | < 5 | < 5 | < 5 | < 5 |
| | | MP | < 5 | 6 (0.04) | < 5 | < 5 |
| | 31 to 90 days | VTE | < 5 | 44 (0.30) | < 5 | 6 (0.14) |

| Cohort | Time window | Outcome | AURUM | | GOLD | |
|---|---|---|---|---|---|---|
| | | | BNT162b2 | ChAdOx1 | BNT162b2 | ChAdOx1 |
| | | DVT | < 5 | 26 (0.18) | < 5 | < 5 |
| | | PE | < 5 | 18 (0.12) | < 5 | < 5 |
| | | ATE | < 5 | 31 (0.21) | < 5 | 7 (0.17) |
| | | IS | < 5 | < 5 | < 5 | < 5 |
| | | TIA | < 5 | 13 (0.09) | < 5 | < 5 |
| | | MI | < 5 | 16 (0.11) | < 5 | < 5 |
| | | HF | < 5 | 23 (0.16) | < 5 | < 5 |
| | | HS | < 5 | < 5 | < 5 | < 5 |
| | | MP | < 5 | 7 (0.05) | < 5 | < 5 |
| | 91 to 180 days | VTE | < 5 | 25 (0.17) | < 5 | < 5 |
| | | DVT | < 5 | 15 (0.10) | < 5 | < 5 |
| | | PE | < 5 | 13 (0.09) | < 5 | < 5 |
| | | ATE | < 5 | 26 (0.18) | < 5 | < 5 |
| | | IS | < 5 | < 5 | < 5 | < 5 |
| | | TIA | < 5 | 8 (0.05) | < 5 | < 5 |
| | | MI | < 5 | 17 (0.12) | < 5 | < 5 |
| | | HF | < 5 | 10 (0.07) | < 5 | < 5 |
| | | HS | < 5 | < 5 | < 5 | < 5 |
| | | MP | < 5 | < 5 | < 5 | < 5 |
| | 181 to 365 days | VTE | < 5 | 9 (0.06) | < 5 | < 5 |
| | | DVT | < 5 | < 5 | < 5 | < 5 |
| | | PE | < 5 | 6 (0.04) | < 5 | < 5 |
| | | ATE | < 5 | < 5 | < 5 | < 5 |
| | | IS | < 5 | < 5 | < 5 | < 5 |
| | | TIA | < 5 | < 5 | < 5 | < 5 |
| | | MI | < 5 | < 5 | < 5 | < 5 |
| | | HF | < 5 | < 5 | < 5 | < 5 |
| | | HS | < 5 | < 5 | < 5 | < 5 |
| | | MP | < 5 | < 5 | < 5 | < 5 |
| Cohort 4 | | | N = 1,335,671 | N = 542,670 | N = 365,096 | N = 147,744 |
| | 0 to 30 days | VTE | 20 (0.15) | 27 (0.50) | < 5 | 8 (0.54) |
| | | DVT | 9 (0.07) | 7 (0.13) | < 5 | < 5 |
| | | PE | 11 (0.08) | 21 (0.39) | < 5 | 6 (0.41) |
| | | ATE | < 5 | 5 (0.09) | < 5 | < 5 |
| | | IS | < 5 | < 5 | < 5 | < 5 |
| | | TIA | < 5 | < 5 | < 5 | < 5 |
| | | MI | < 5 | < 5 | < 5 | < 5 |
| | | HF | < 5 | < 5 | < 5 | < 5 |
| | | HS | < 5 | < 5 | < 5 | < 5 |
| | | MP | < 5 | < 5 | < 5 | < 5 |
| | 31 to 90 days | VTE | 7 (0.05) | 14 (0.26) | < 5 | < 5 |
| | | DVT | < 5 | 8 (0.15) | < 5 | < 5 |
| | | PE | < 5 | 6 (0.11) | < 5 | < 5 |
| | | ATE | < 5 | < 5 | < 5 | < 5 |
| | | IS | < 5 | < 5 | < 5 | < 5 |
| | | TIA | < 5 | < 5 | < 5 | < 5 |
| | | MI | < 5 | < 5 | < 5 | < 5 |
| | | HF | 6 (0.04) | < 5 | < 5 | < 5 |
| | | HS | < 5 | < 5 | < 5 | < 5 |

| Cohort | Time window | Outcome | AURUM | | GOLD | |
|---|---|---|---|---|---|---|
| | | | BNT162b2 | ChAdOx1 | BNT162b2 | ChAdOx1 |
| | | MP | < 5 | < 5 | < 5 | < 5 |
| | 91 to 180 days | VTE | 9 (0.07) | < 5 | < 5 | < 5 |
| | | DVT | 7 (0.05) | < 5 | < 5 | < 5 |
| | | PE | < 5 | < 5 | < 5 | < 5 |
| | | ATE | < 5 | < 5 | < 5 | < 5 |
| | | IS | < 5 | < 5 | < 5 | < 5 |
| | | TIA | < 5 | < 5 | < 5 | < 5 |
| | | MI | < 5 | < 5 | < 5 | < 5 |
| | | HF | < 5 | < 5 | < 5 | < 5 |
| | | HS | < 5 | < 5 | < 5 | < 5 |
| | | MP | < 5 | < 5 | < 5 | < 5 |
| | 181 to 365 days | VTE | < 5 | < 5 | < 5 | < 5 |
| | | DVT | < 5 | < 5 | < 5 | < 5 |
| | | PE | < 5 | < 5 | < 5 | < 5 |
| | | ATE | < 5 | < 5 | < 5 | < 5 |
| | | IS | < 5 | < 5 | < 5 | < 5 |
| | | TIA | < 5 | < 5 | < 5 | < 5 |
| | | MI | < 5 | < 5 | < 5 | < 5 |
| | | HF | < 5 | < 5 | < 5 | < 5 |
| | | HS | < 5 | < 5 | < 5 | < 5 |
| | | MP | < 5 | < 5 | < 5 | < 5 |

VTE = Venous thromboembolism (DVT + PE), DVT = Deep vein thrombosis, PE = Pulmonary embolism, ATE = Arterial thromboembolism (IS + TIA + MI), IS = Ischemic stroke, TIA = Transient ischemic attack, MI = Myocardial infarction, HF = Heart failure, HS = Hemorrhagic stroke, MP = Myocarditis or Pericarditis

**Figure S8: Forest plots for vaccine effectiveness (any COVID-19 vaccine),** meta-analysis across cohorts and databases. Follow-up ends at first vaccine dose after index date. Dashed line represents a level of hetereogeneity I2 > 0.4.

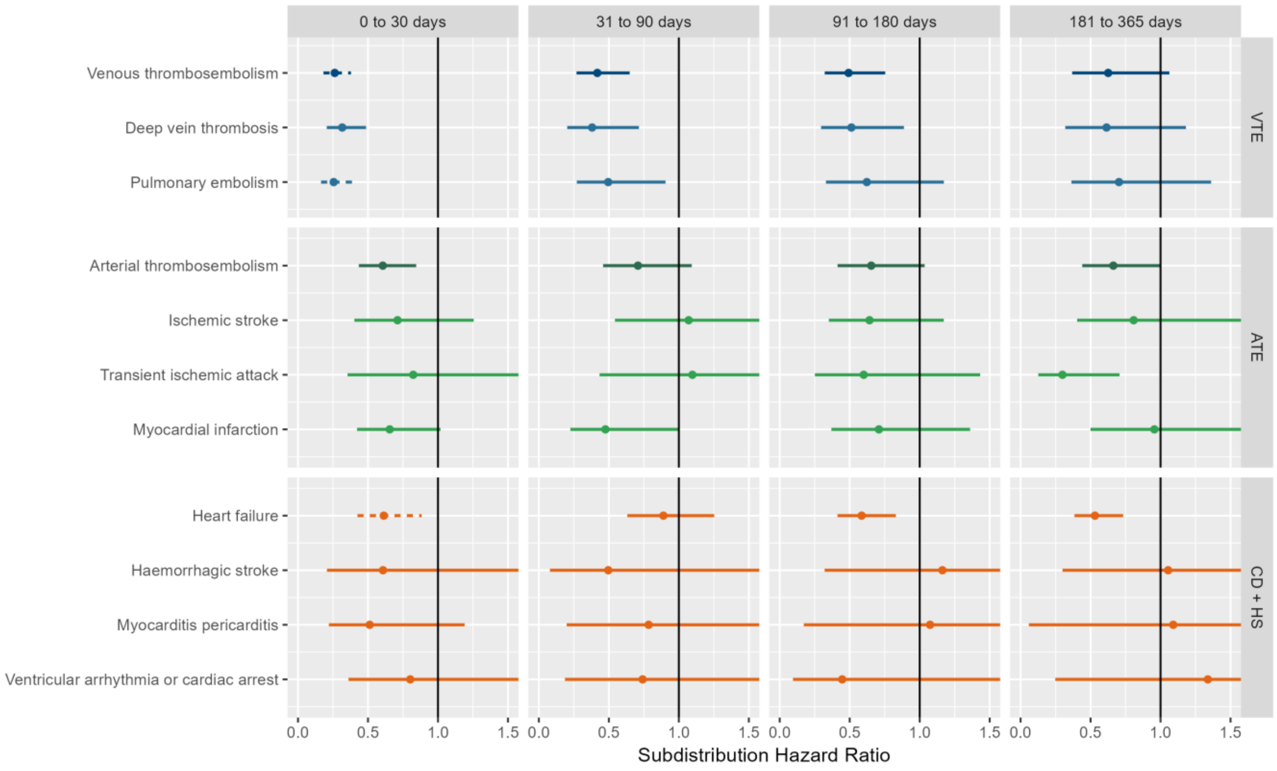

VTE = Venous thromboembolism, ATE = Arterial thromboembolism, CD + HS = Cardiac diseases and Hemorrhagic stroke

**Figure S9: Forest plots for vaccine effectiveness (any COVID-19 vaccine),** meta-analysis across cohorts and databases. Only first outcome after COVID-19 captured. Dashed line represents a level of hetereogeneity I2 > 0.4.

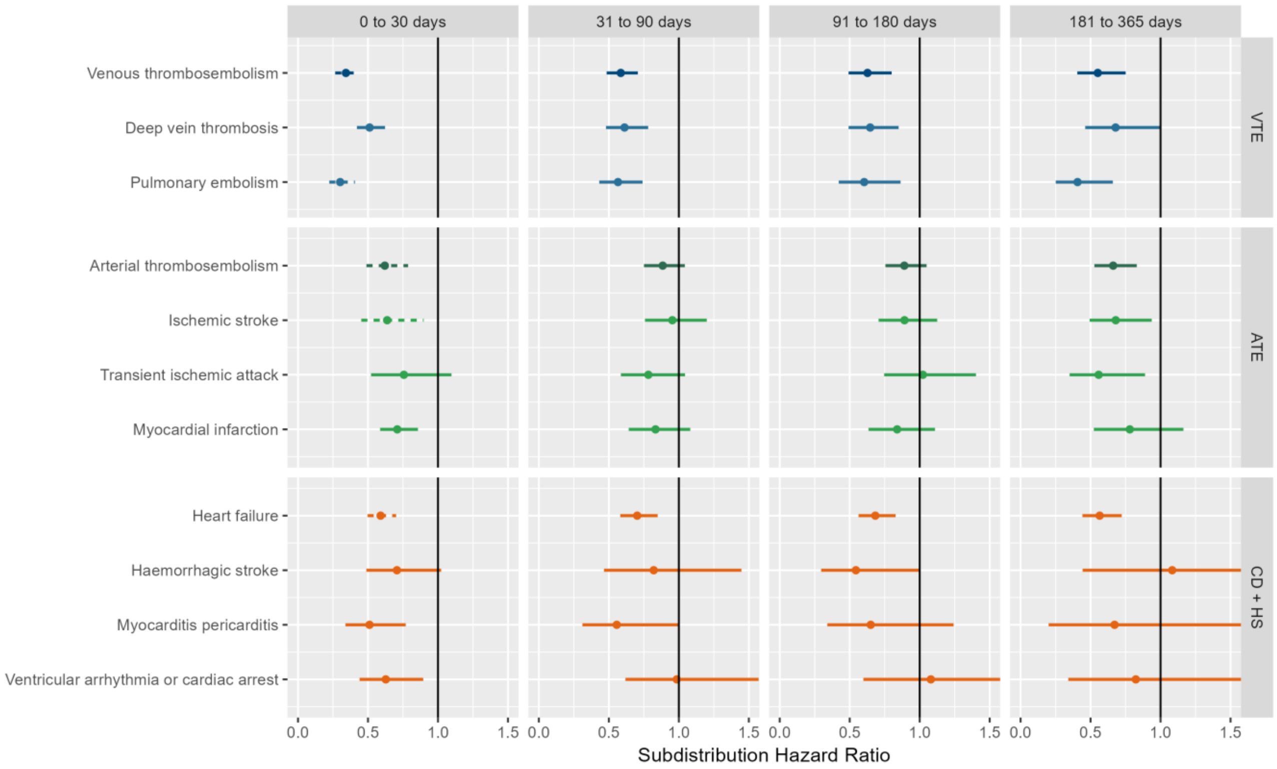

VTE = Venous thromboembolism, ATE = Arterial thromboembolism, CD + HS = Cardiac diseases and Hemorrhagic stroke

**Figure S10: Forest plots for vaccine effectiveness (ChAdOx1 vaccine),** meta-analysis across cohorts and databases. Dashed line represents a level of hetereogeneity I2 > 0.4.

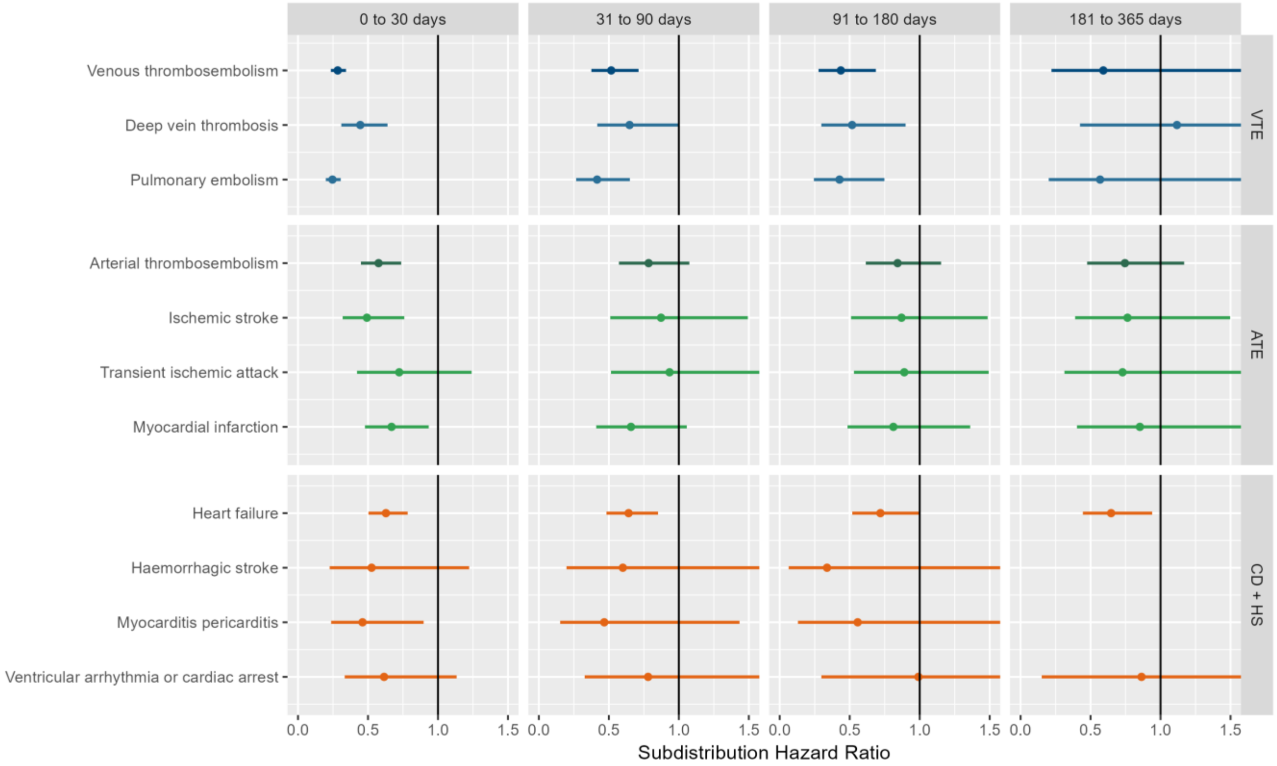

VTE = Venous thromboembolism, ATE = Arterial thromboembolism, CD + HS = Cardiac diseases and Hemorrhagic stroke

**Figure S11: Forest plots for vaccine effectiveness (ChAdOx1 vaccine),** meta-analysis across cohorts and databases. Follow-up ends at first vaccine dose after index date. Dashed line represents a level of hetereogeneity I2 > 0.4.

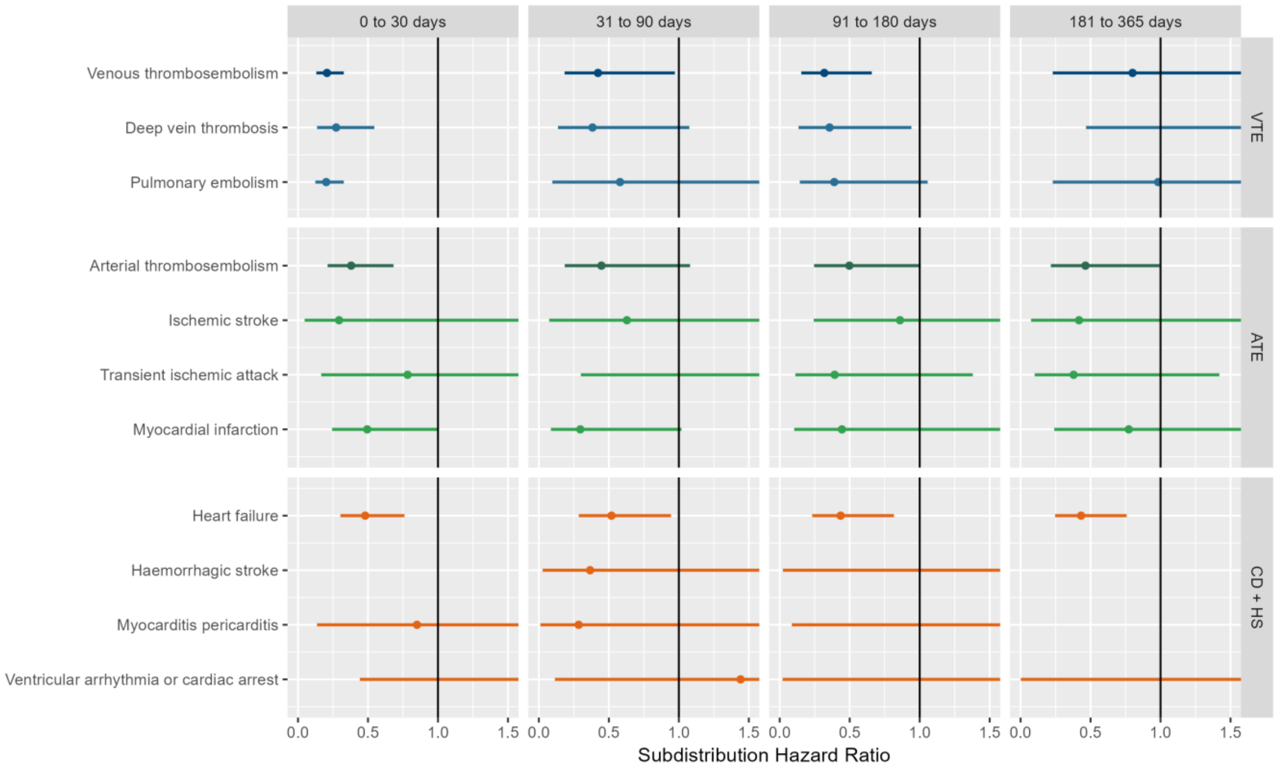

VTE = Venous thromboembolism, ATE = Arterial thromboembolism, CD + HS = Cardiac diseases and Hemorrhagic stroke

**Figure S12: Forest plots for vaccine effectiveness (ChAdOx1 vaccine),** meta-analysis across cohorts and databases. Only first outcome after COVID-19 captured. Dashed line represents a level of hetereogeneity I2 > 0.4.

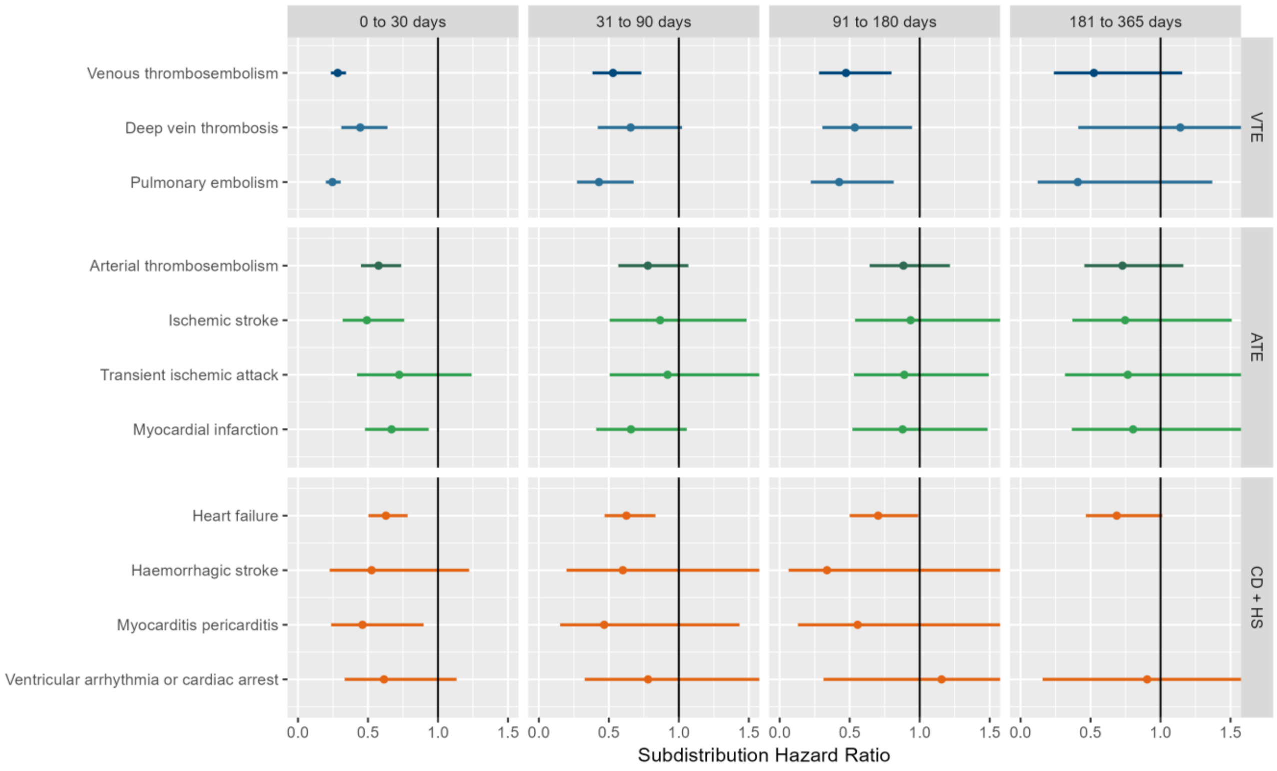

VTE = Venous thromboembolism, ATE = Arterial thromboembolism, CD + HS = Cardiac diseases and Hemorrhagic stroke

**Figure S13: Forest plots for vaccine effectiveness (BNT162b2 vaccine),** meta-analysis across cohorts and databases. Dashed line represents a level of hetereogeneity I2 > 0.4.

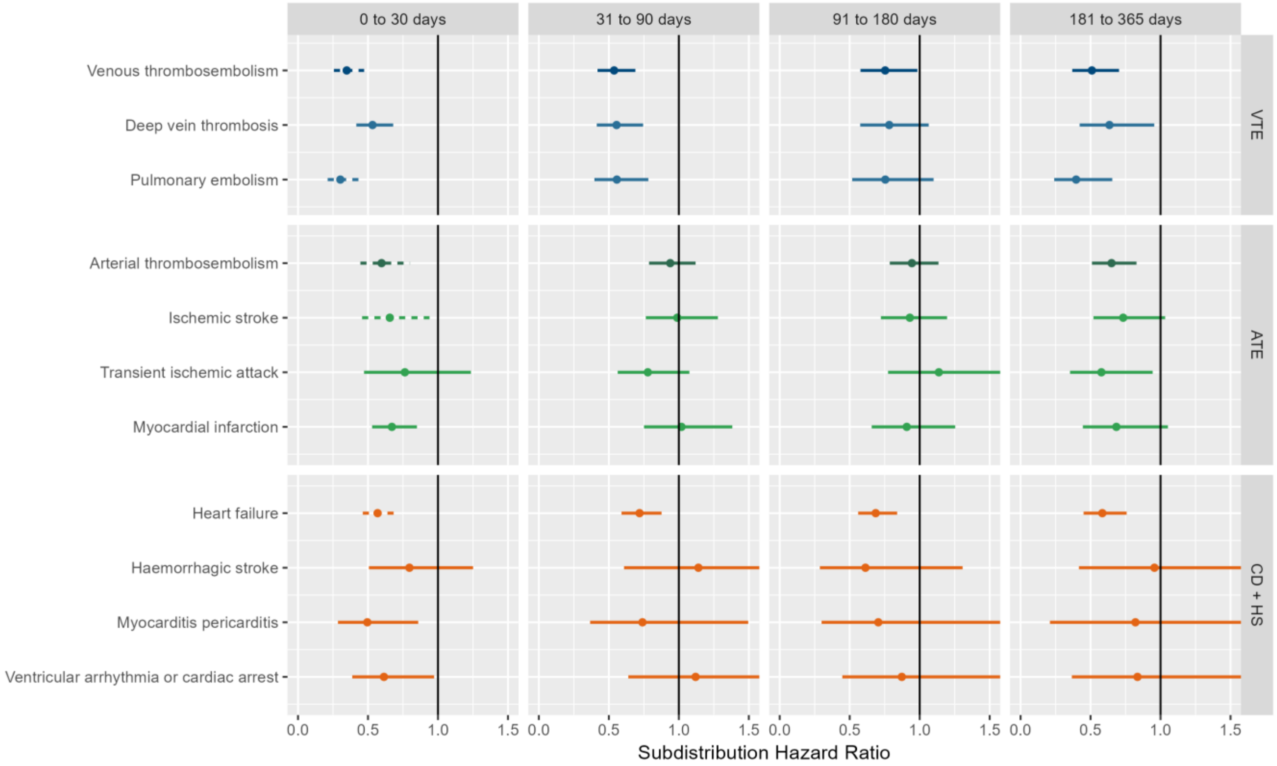

VTE = Venous thromboembolism, ATE = Arterial thromboembolism, CD + HS = Cardiac diseases and Hemorrhagic stroke

**Figure S14: Forest plots for vaccine effectiveness (BNT162b2 vaccine),** meta-analysis across cohorts and databases. Follow-up ends at first vaccine dose after index date. Dashed line represents a level of hetereogeneity I2 > 0.4.

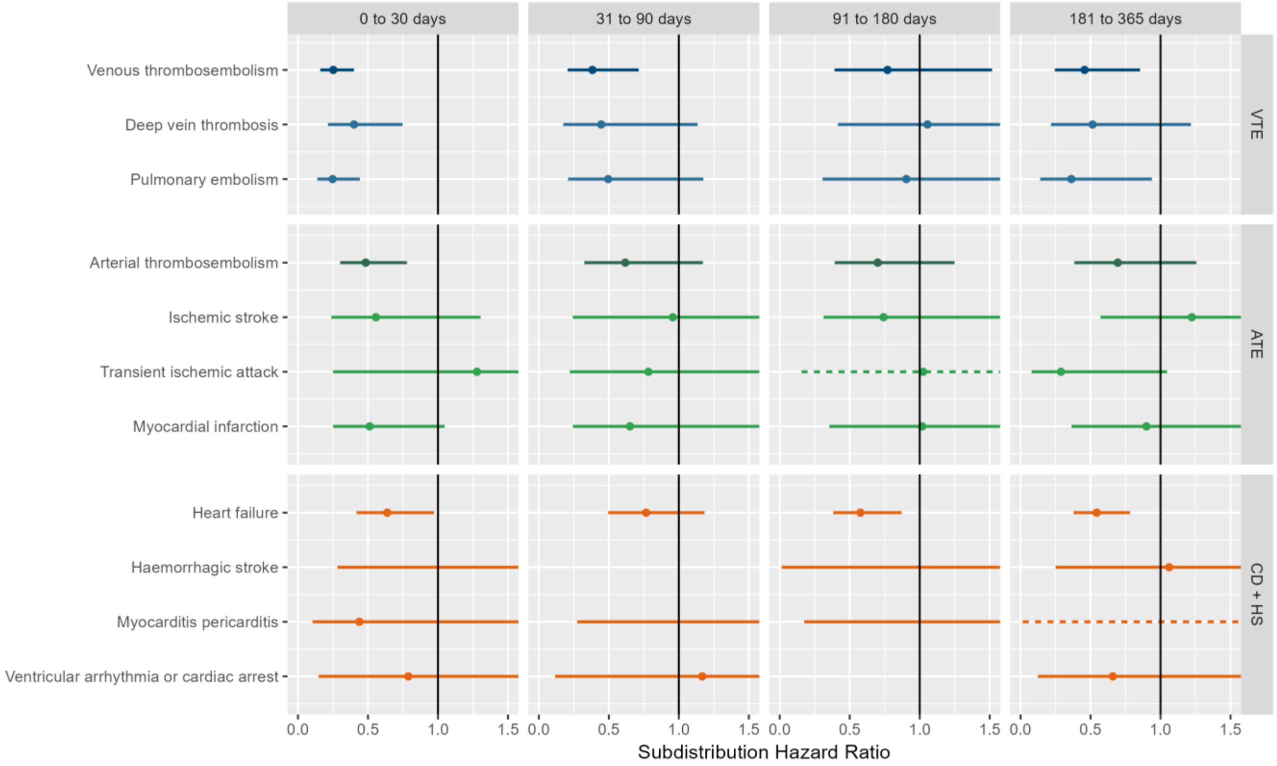

VTE = Venous thromboembolism, ATE = Arterial thromboembolism, CD + HS = Cardiac diseases and Hemorrhagic stroke

**Figure S15: Forest plots for vaccine effectiveness (BNT162b2 vaccine),** meta-analysis across cohorts and databases. Only first outcome after COVID-19 captured. Dashed line represents a level of hetereogeneity I2 > 0.4.

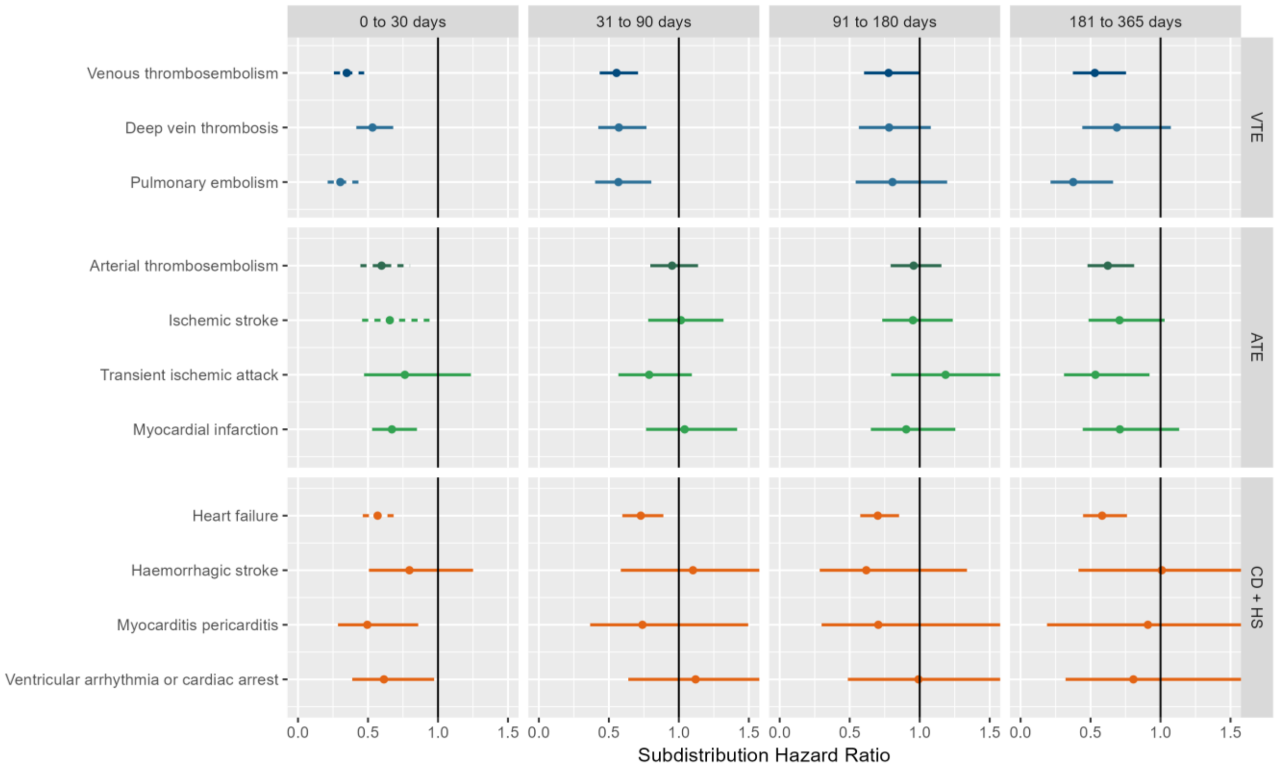

VTE = Venous thromboembolism, ATE = Arterial thromboembolism, CD + HS = Cardiac diseases and Hemorrhagic stroke

**Figure S16: Forest plots for vaccine effectiveness (any COVID-19 vaccine) on preventing venous thromboembolism complications,** for each cohort and database (meta-analysis estimates across cohorts in the last panel). Dashed line represents a level of hetereogeneity I2 > 0.4.

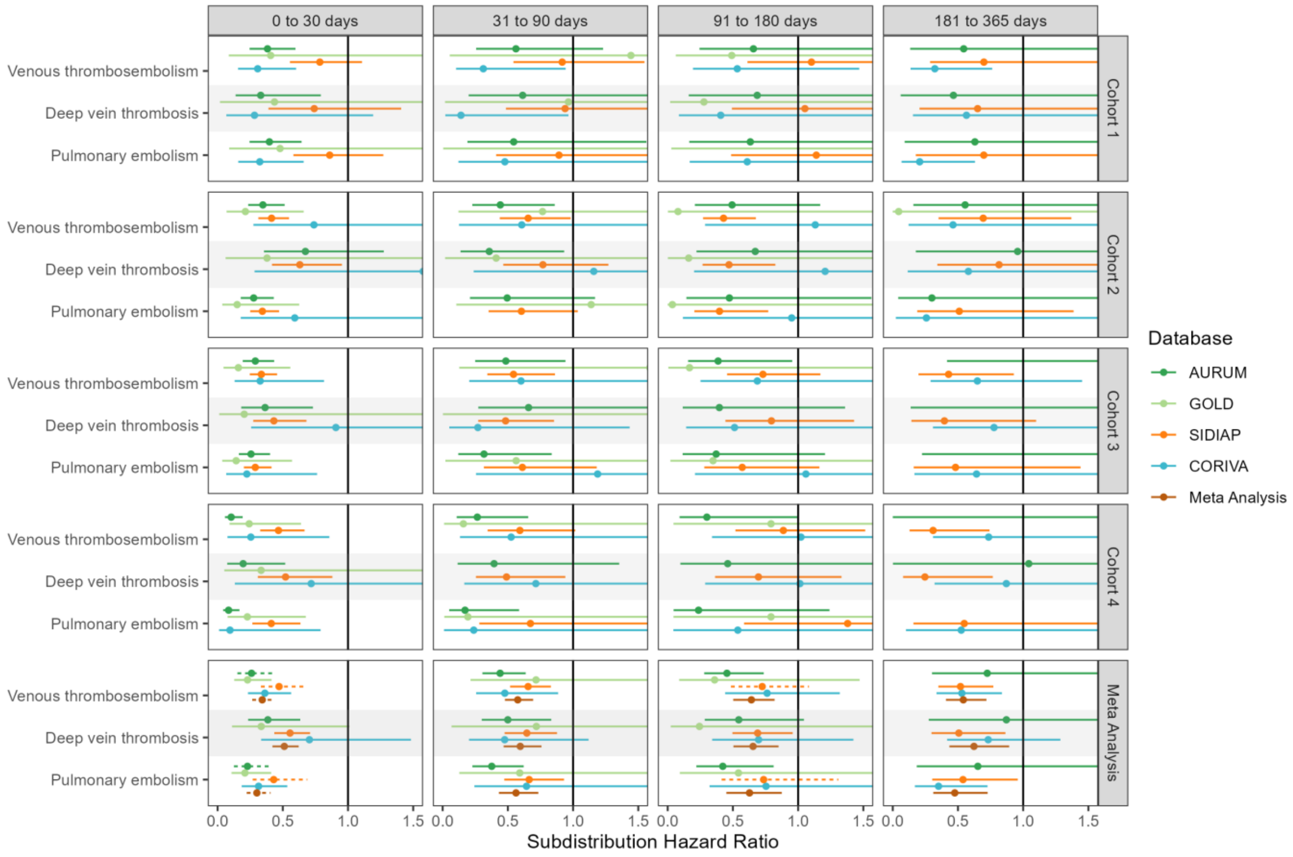

**Figure S17: Forest plots for vaccine effectiveness (any COVID-19 vaccine) on preventing venous thromboembolism complications,** for each cohort and database (meta-analysis estimates across cohorts in the last panel).Follow-up ends at first vaccine dose after index date. Dashed line represents a level of hetereogeneity I2 > 0.4.

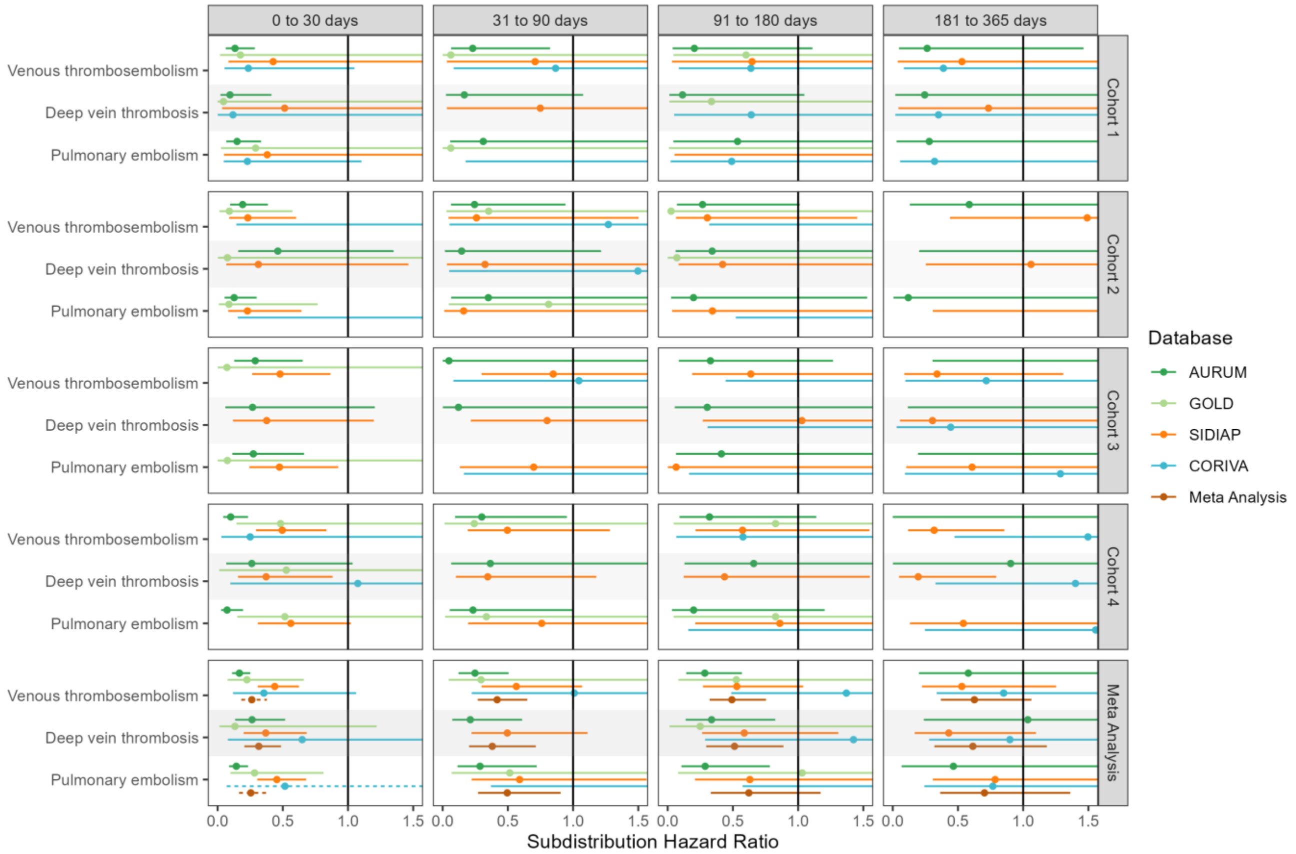

**Figure S18: Forest plots for vaccine effectiveness (any COVID-19 vaccine) on preventing venous thromboembolism complications,** for each cohort and database (meta-analysis estimates across cohorts in the last panel).Only first outcome after COVID-19 captured. Dashed line represents a level of hetereogeneity I2 > 0.4.

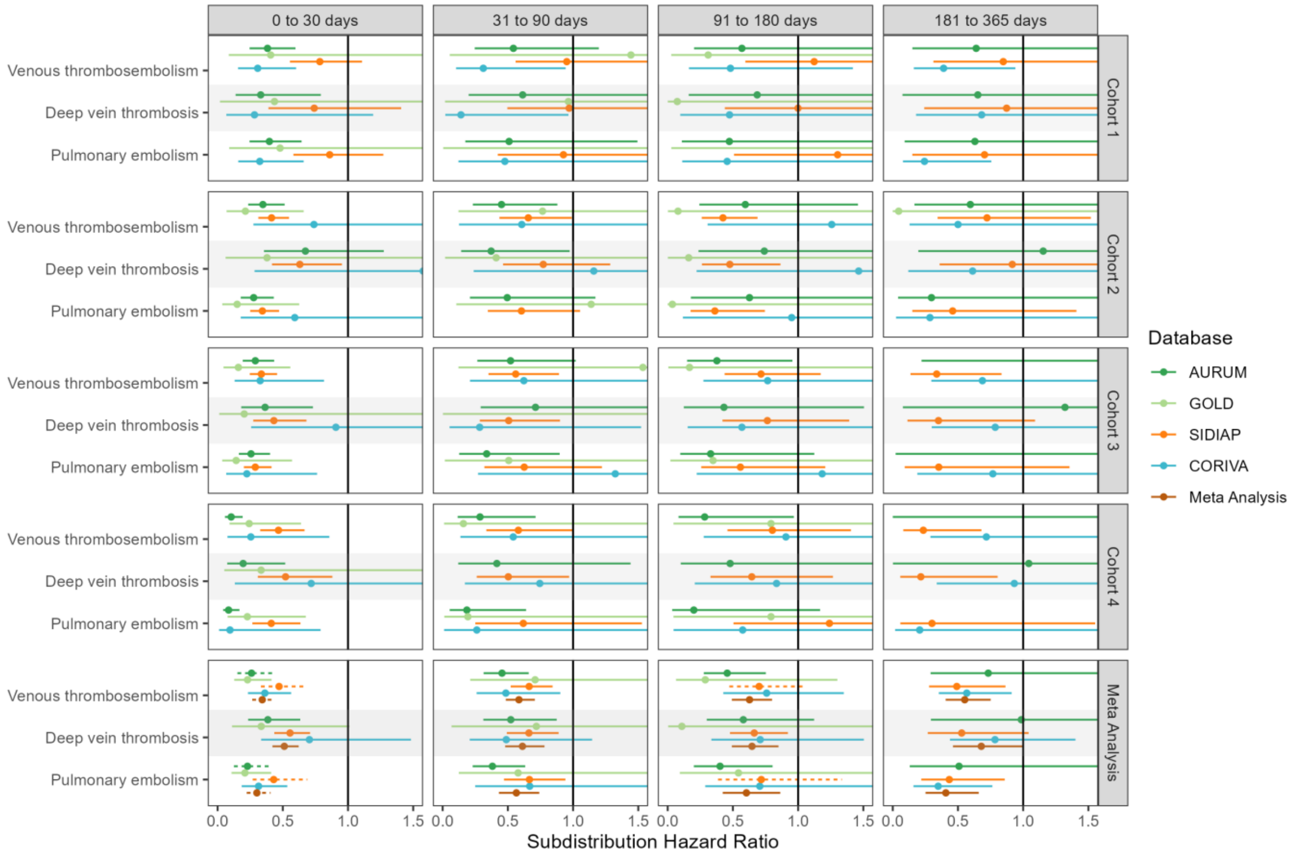

**Figure S19: Forest plots for vaccine effectiveness (ChAdOx1 vaccine) on preventing venous thromboembolism complications,** for each cohort and database (meta-analysis estimates across cohorts in the last panel). Dashed line represents a level of hetereogeneity I2 > 0.4.

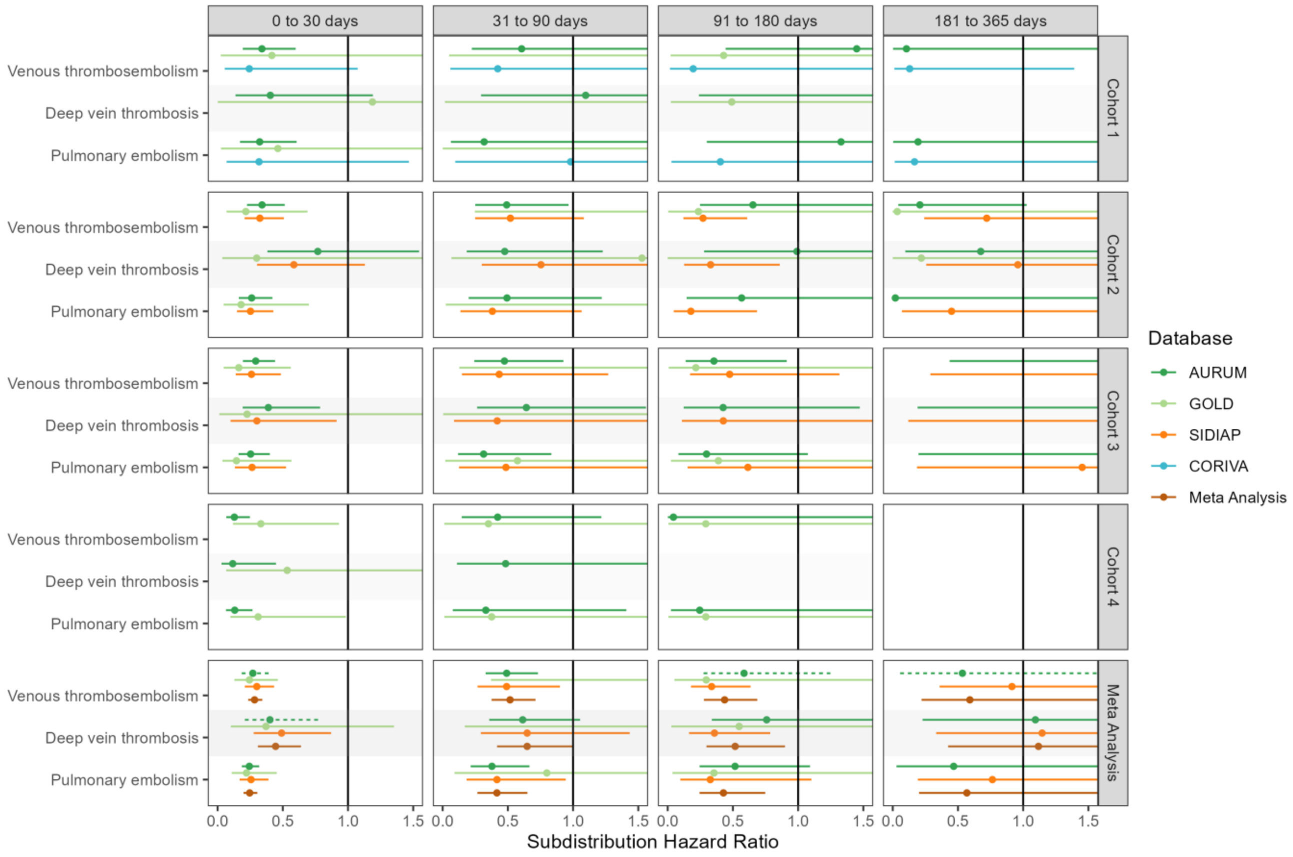

**Figure S20: Forest plots for vaccine effectiveness (ChAdOx1 vaccine) on preventing venous thromboembolism complications,** for each cohort and database (meta-analysis estimates across cohorts in the last panel).Follow-up ends at first vaccine dose after index date. Dashed line represents a level of hetereogeneity I2 > 0.4.

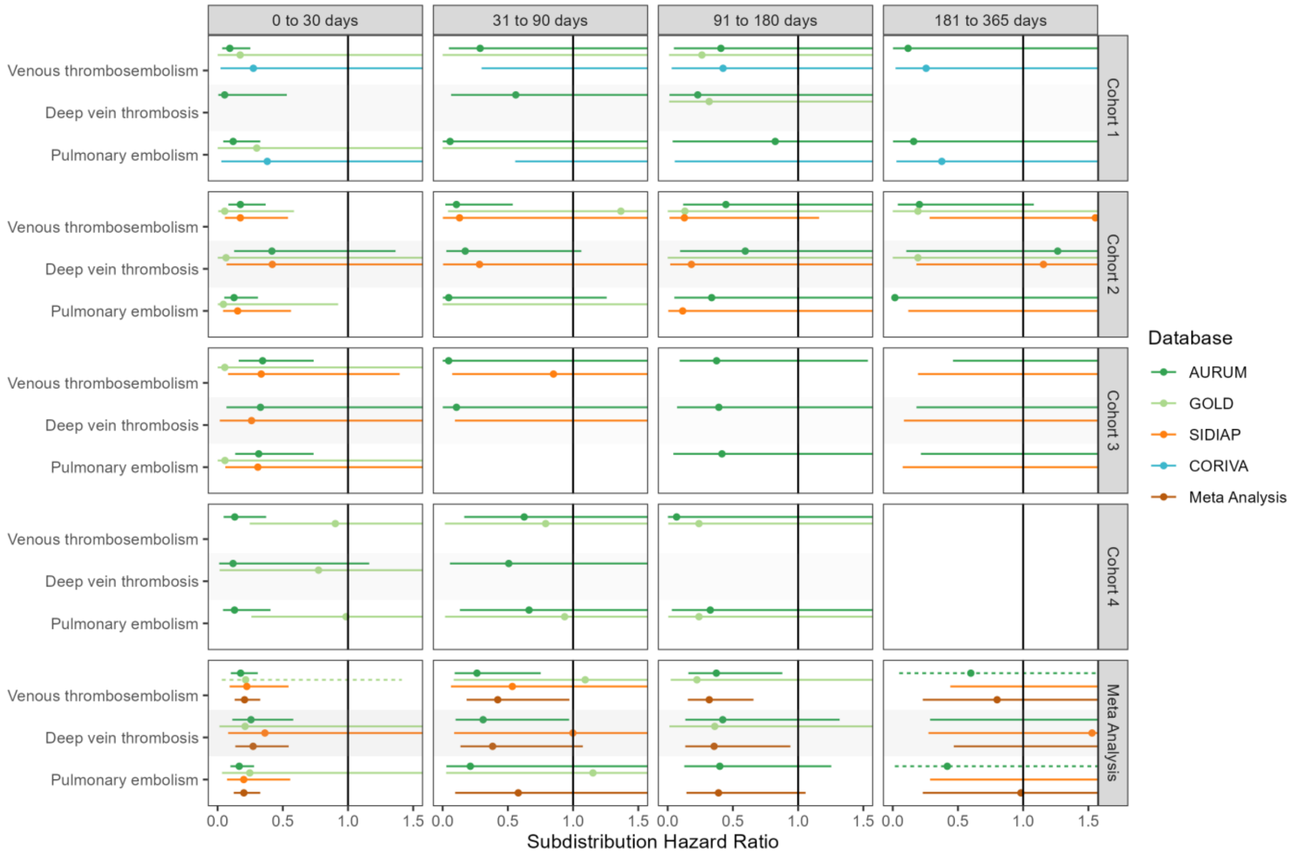

**Figure S21: Forest plots for vaccine effectiveness (ChAdOx1 vaccine) on preventing venous thromboembolism complications,** for each cohort and database (meta-analysis estimates across cohorts in the last panel).Only first outcome after COVID-19 captured. Dashed line represents a level of hetereogeneity I2 > 0.4.

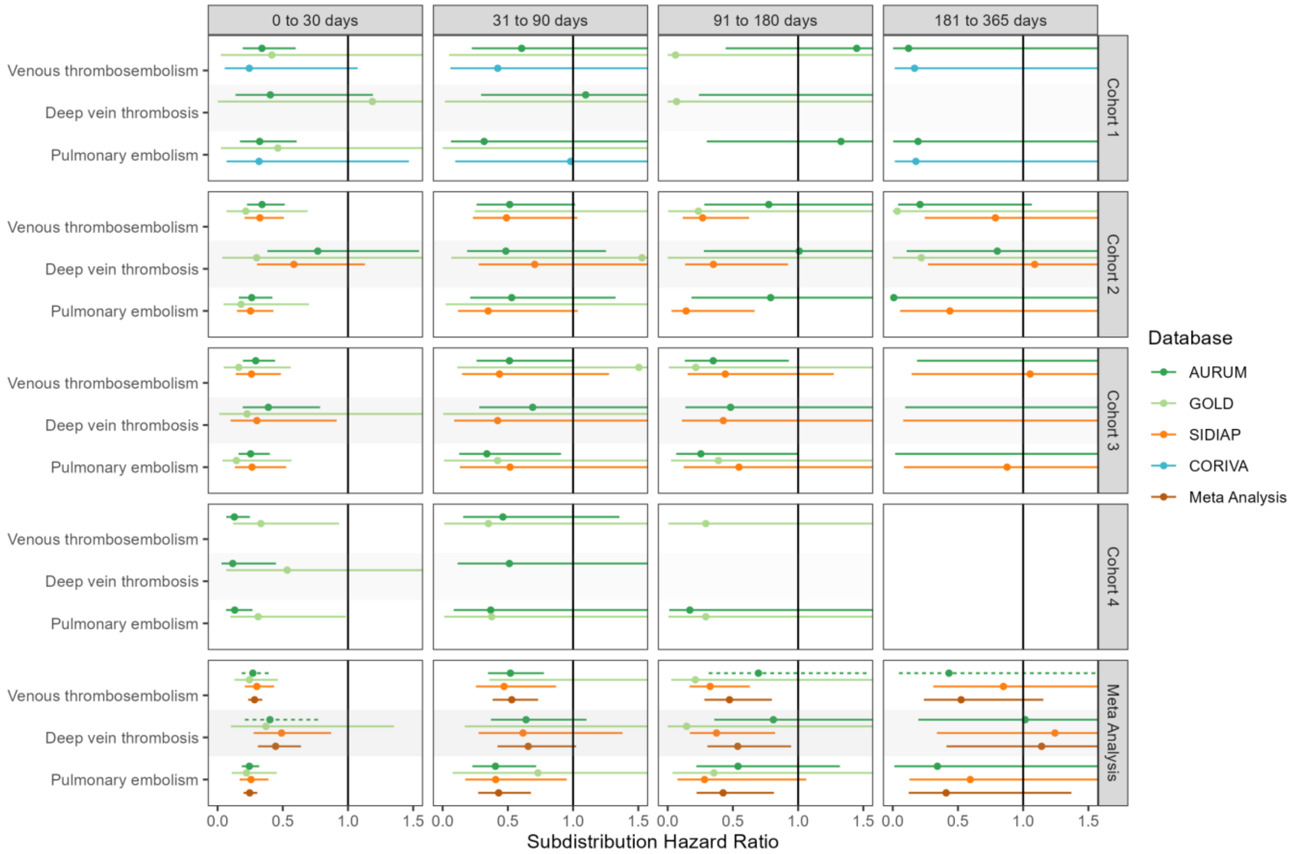

**Figure S22: Forest plots for vaccine effectiveness (BNT162b2 vaccine) on preventing venous thromboembolism complications,** for each cohort and database (meta-analysis estimates across cohorts in the last panel). Dashed line represents a level of hetereogeneity I2 > 0.4.

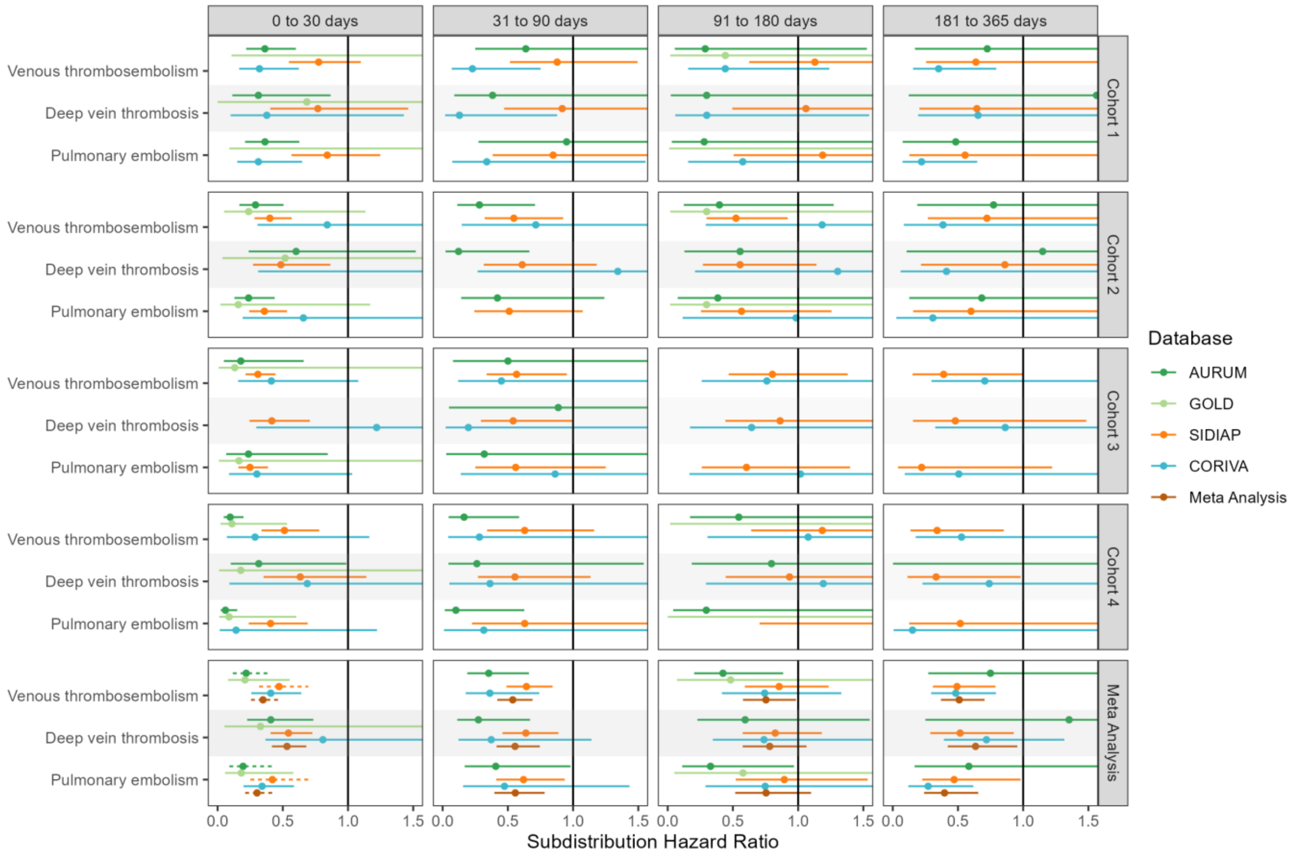

**Figure S23: Forest plots for vaccine effectiveness (BNT162b2 vaccine) on preventing venous thromboembolism complications,** for each cohort and database (meta-analysis estimates across cohorts in the last panel).Follow-up ends at first vaccine dose after index date. Dashed line represents a level of hetereogeneity I2 > 0.4.

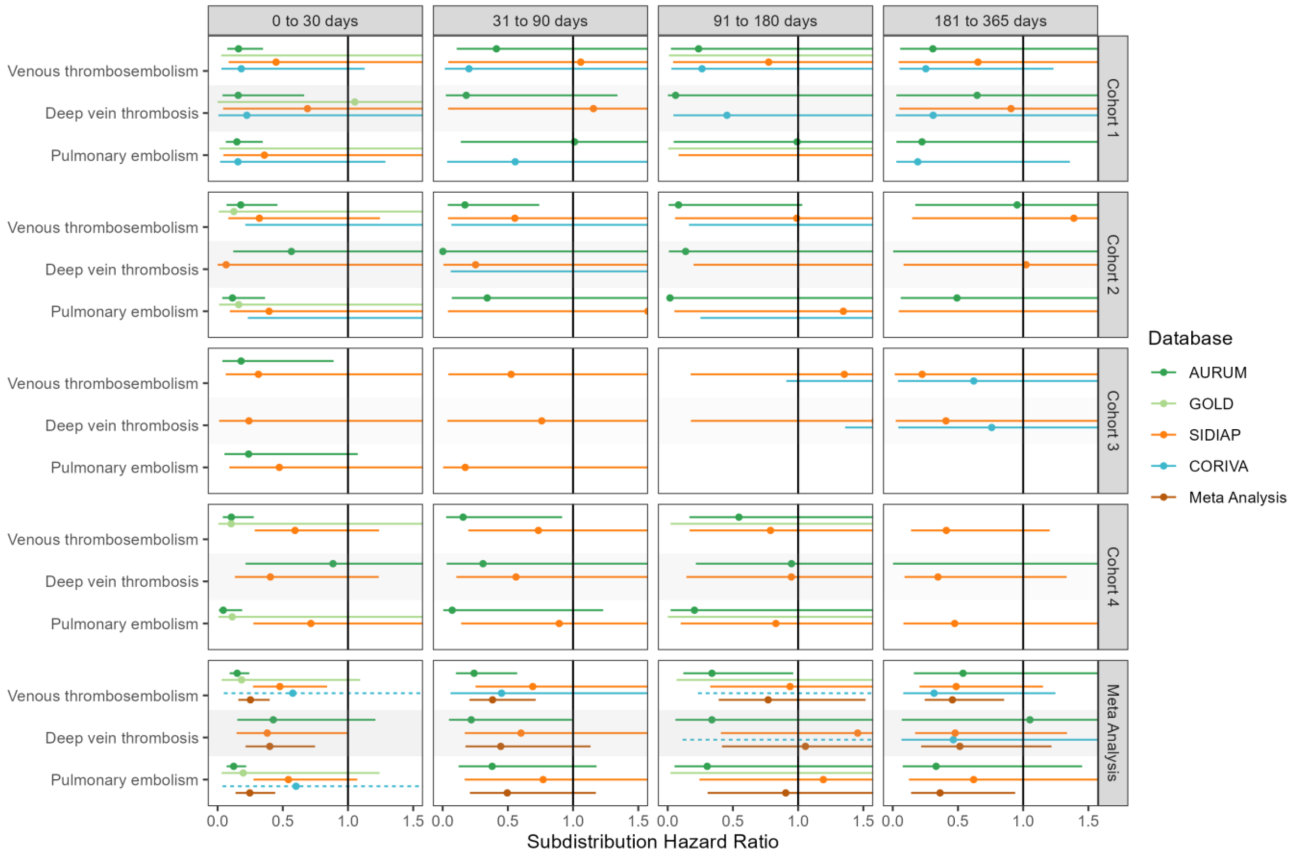

**Figure S24: Forest plots for vaccine effectiveness (BNT162b2 vaccine) on preventing venous thromboembolism complications,** for each cohort and database (meta-analysis estimates across cohorts in the last panel).Only first outcome after COVID-19 captured. Dashed line represents a level of hetereogeneity I2 > 0.4.

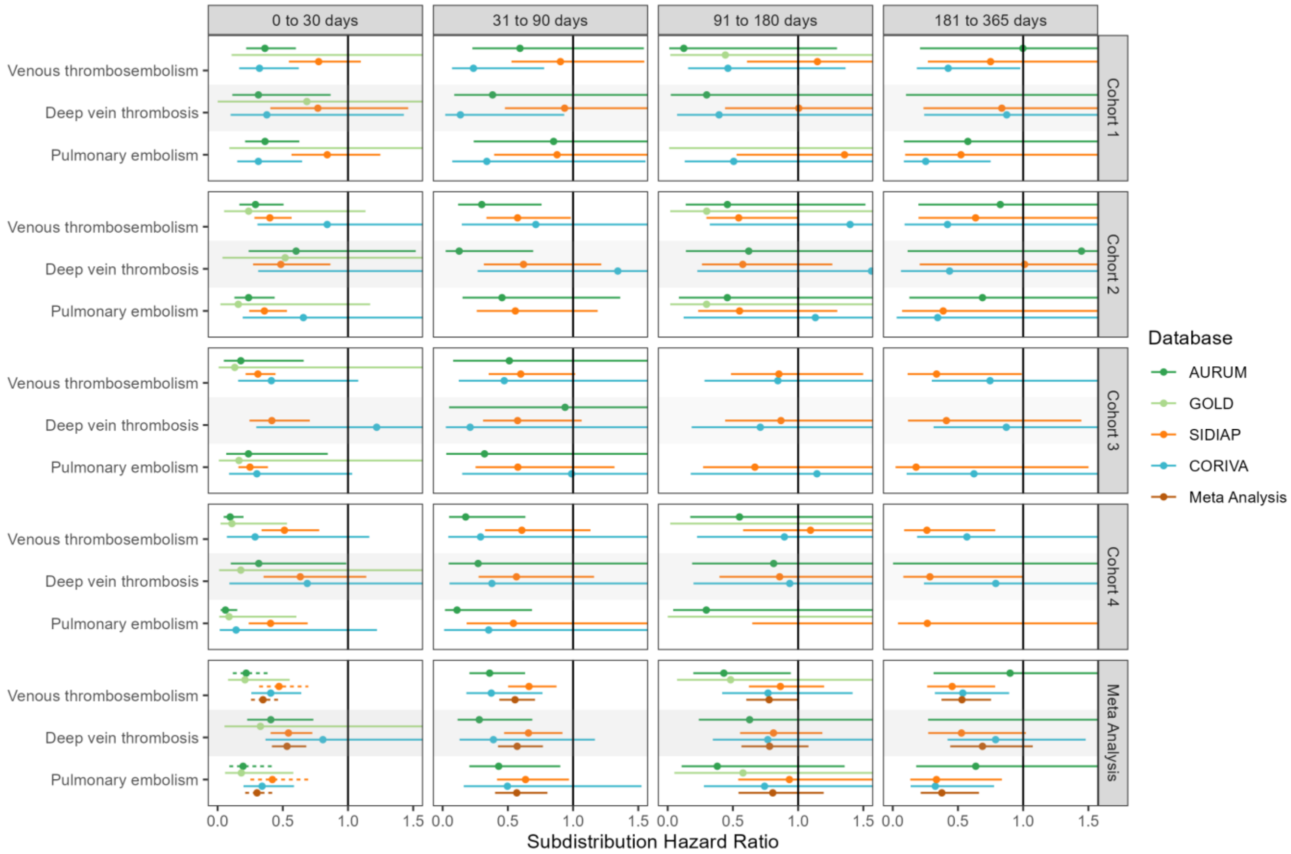

**Figure S25: Forest plots for vaccine effectiveness (any COVID-19 vaccine) on preventing arterial thromboembolism complications,** for each cohort and database (meta-analysis estimates across cohorts in the last panel). Dashed line represents a level of hetereogeneity I2 > 0.4.

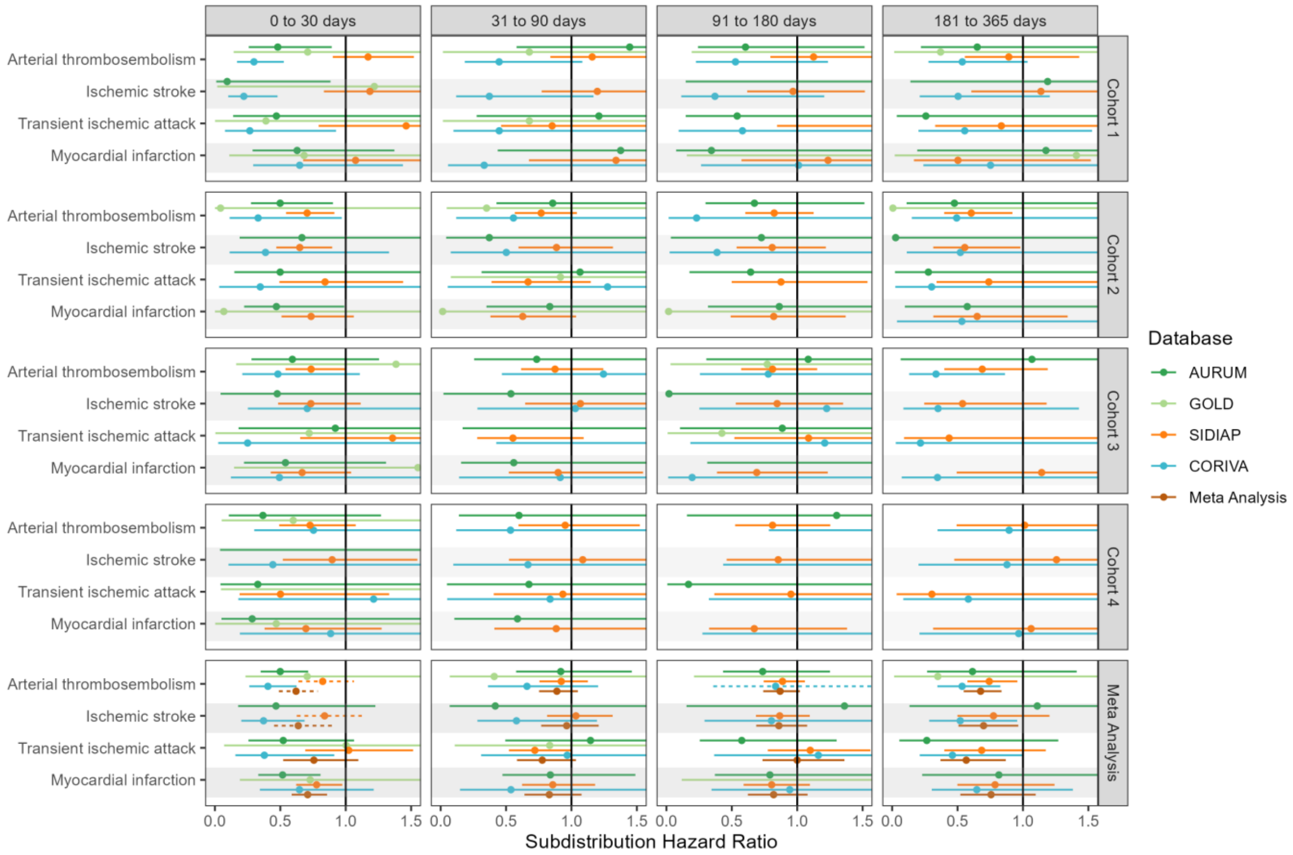

**Figure S26: Forest plots for vaccine effectiveness (any COVID-19 vaccine) on preventing arterial thromboembolism complications,** for each cohort and database (meta-analysis estimates across cohorts in the last panel).Follow-up ends at first vaccine dose after index date. Dashed line represents a level of hetereogeneity I2 > 0.4.

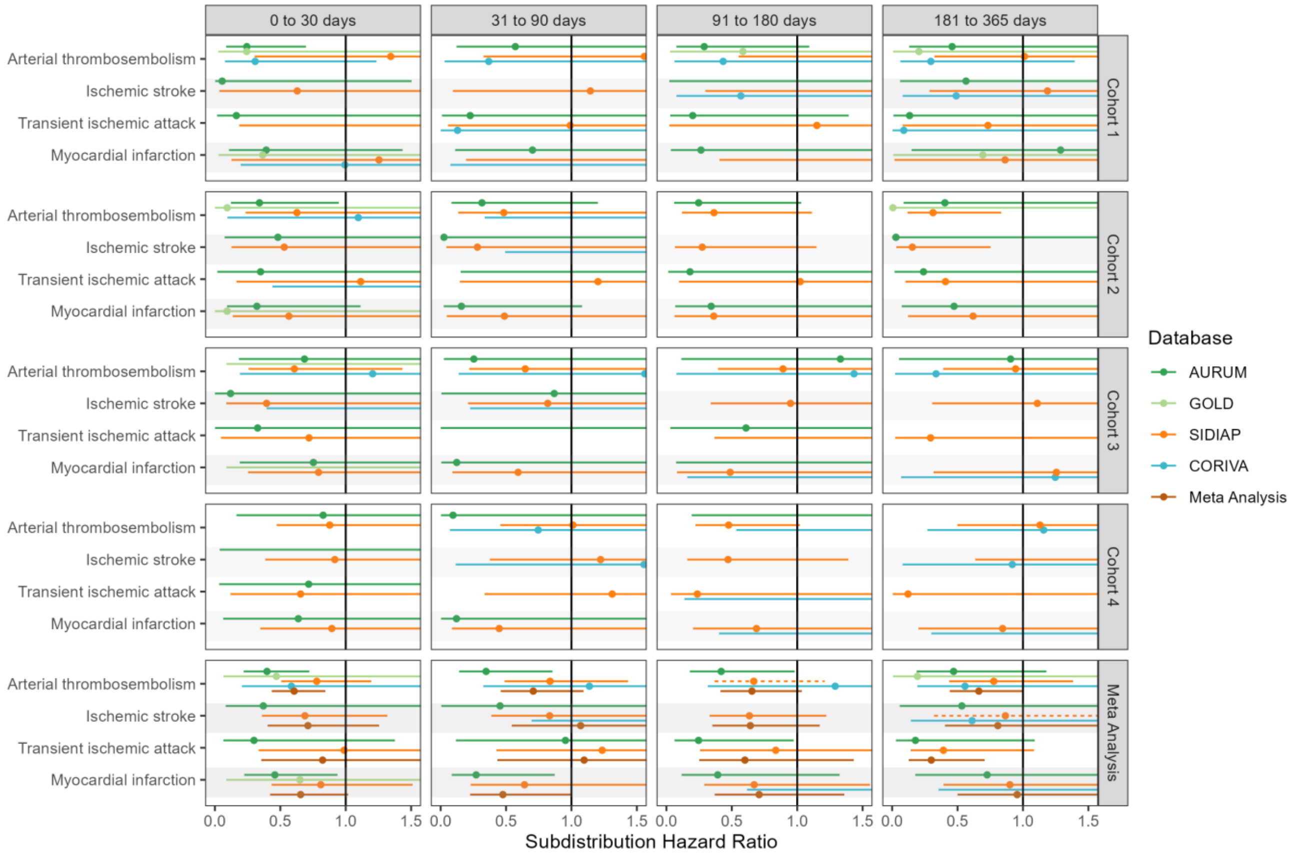

**Figure S27: Forest plots for vaccine effectiveness (any COVID-19 vaccine) on preventing arterial thromboembolism complications,** for each cohort and database (meta-analysis estimates across cohorts in the last panel).Only first outcome after COVID-19 captured. Dashed line represents a level of hetereogeneity I2 > 0.4.

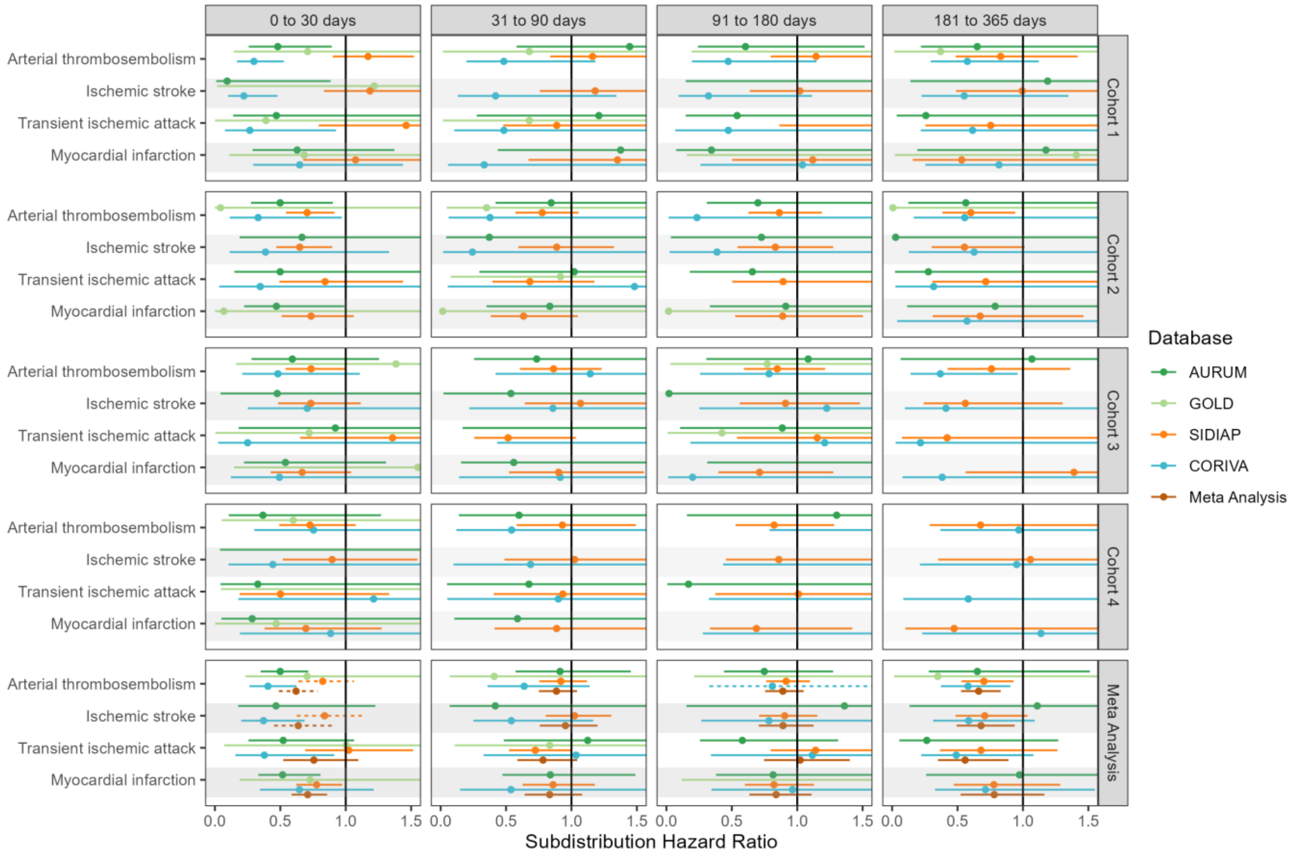

**Figure S28: Forest plots for vaccine effectiveness (ChAdOx1 vaccine) on preventing arterial thromboembolism complications,** for each cohort and database (meta-analysis estimates across cohorts in the last panel). Dashed line represents a level of hetereogeneity I2 > 0.4.

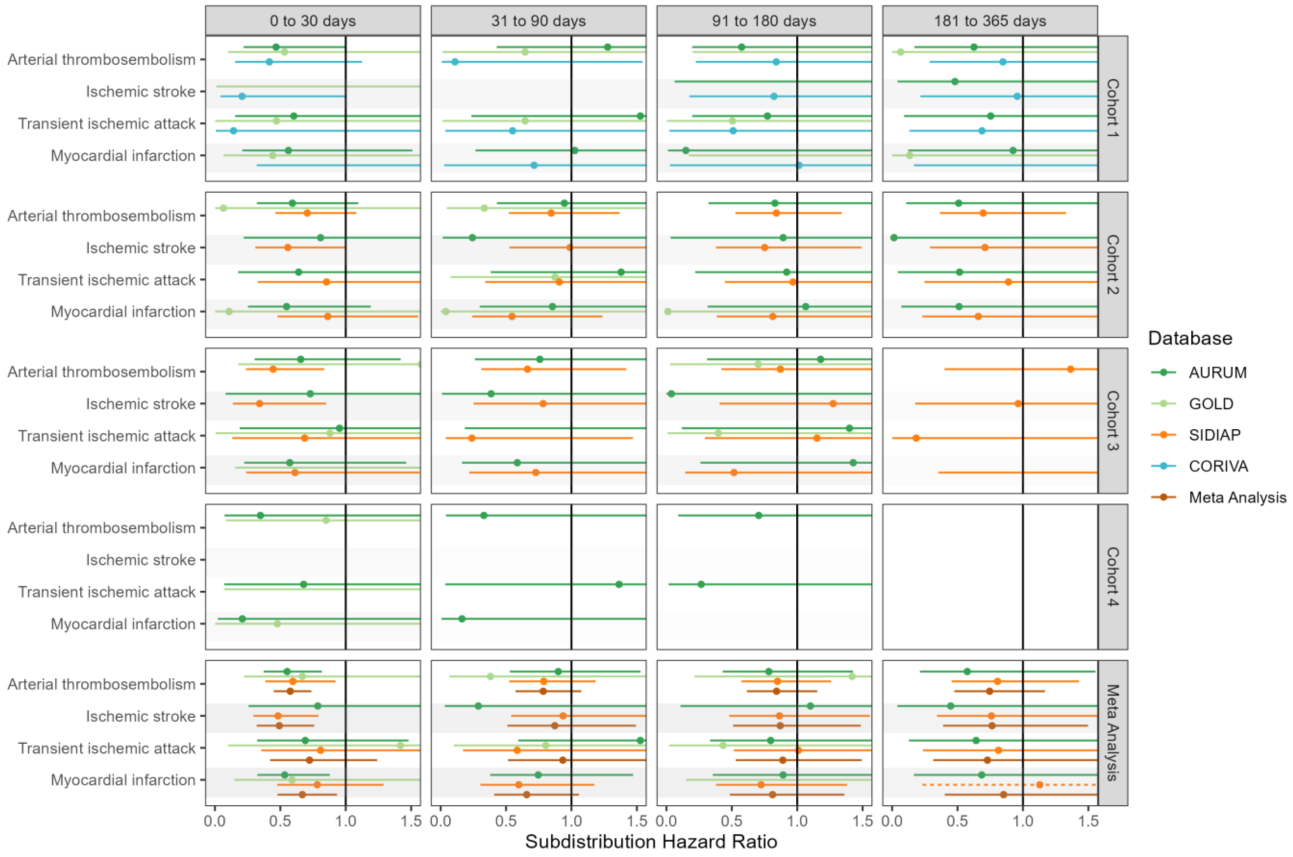

**Figure S29: Forest plots for vaccine effectiveness (ChAdOx1 vaccine) on preventing arterial thromboembolism complications,** for each cohort and database (meta-analysis estimates across cohorts in the last panel).Follow-up ends at first vaccine dose after index date. Dashed line represents a level of hetereogeneity I2 > 0.4.

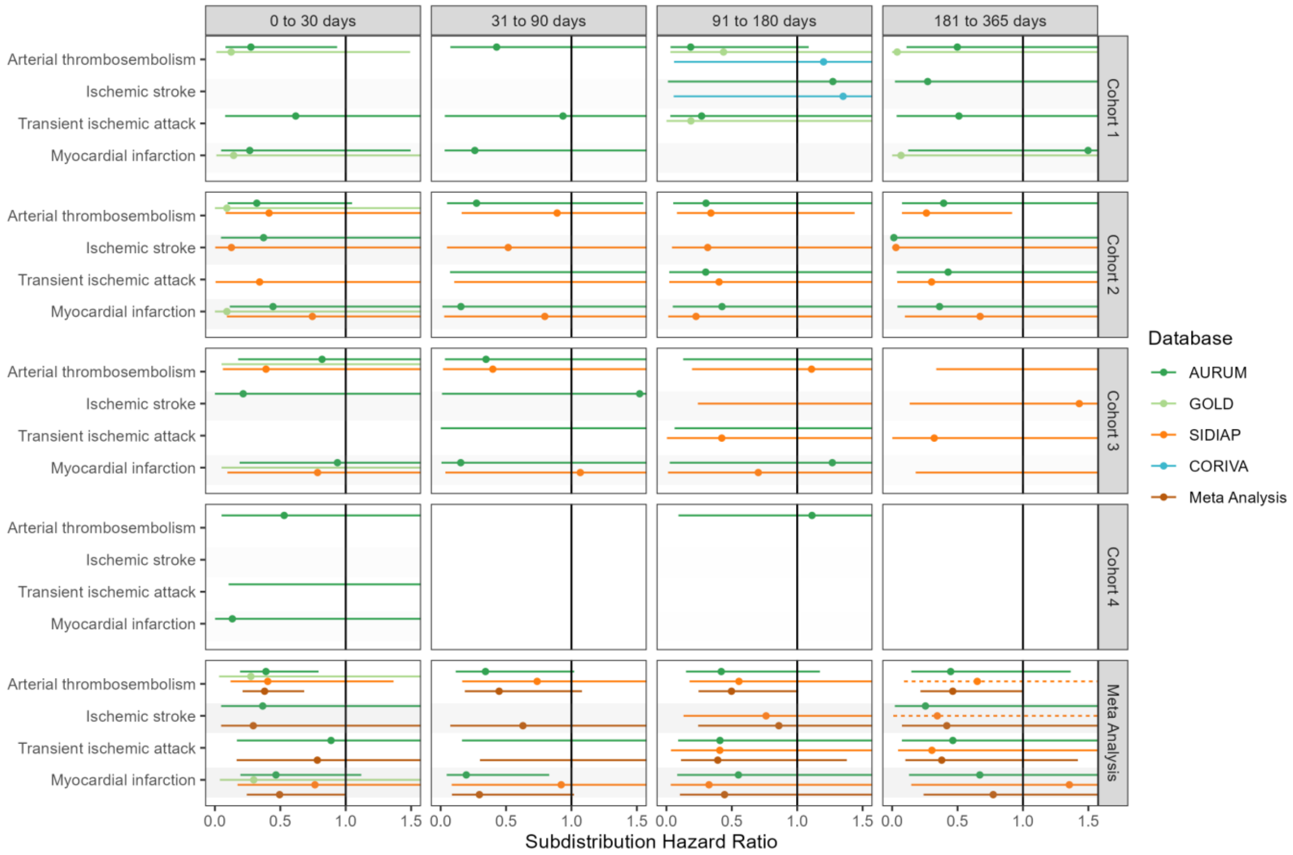

**Figure S30: Forest plots for vaccine effectiveness (ChAdOx1 vaccine) on preventing arterial thromboembolism complications,** for each cohort and database (meta-analysis estimates across cohorts in the last panel).Only first outcome after COVID-19 captured. Dashed line represents a level of hetereogeneity I2 > 0.4.

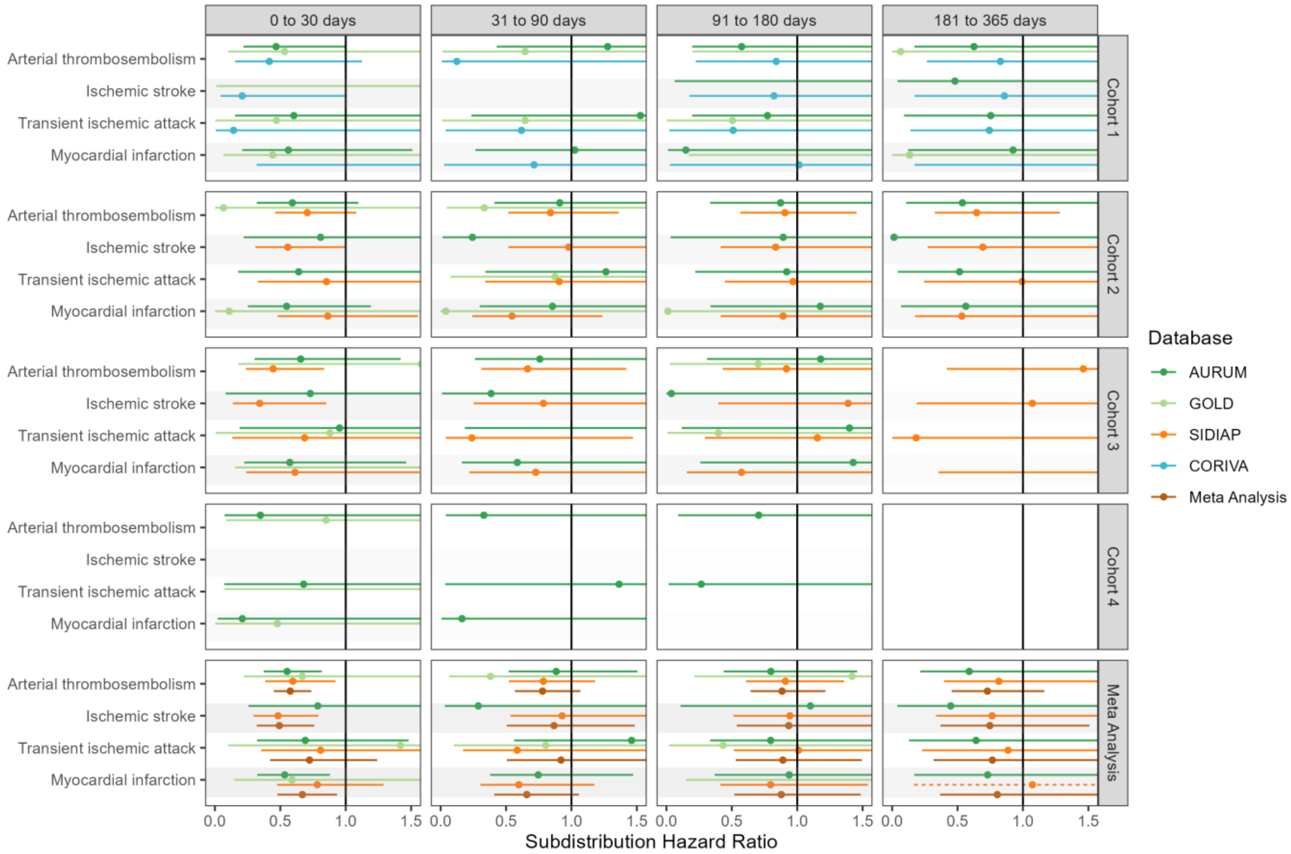

**Figure S31: Forest plots for vaccine effectiveness (BNT162b2 vaccine) on preventing arterial thromboembolism complications,** for each cohort and database (meta-analysis estimates across cohorts in the last panel). Dashed line represents a level of hetereogeneity I2 > 0.4.

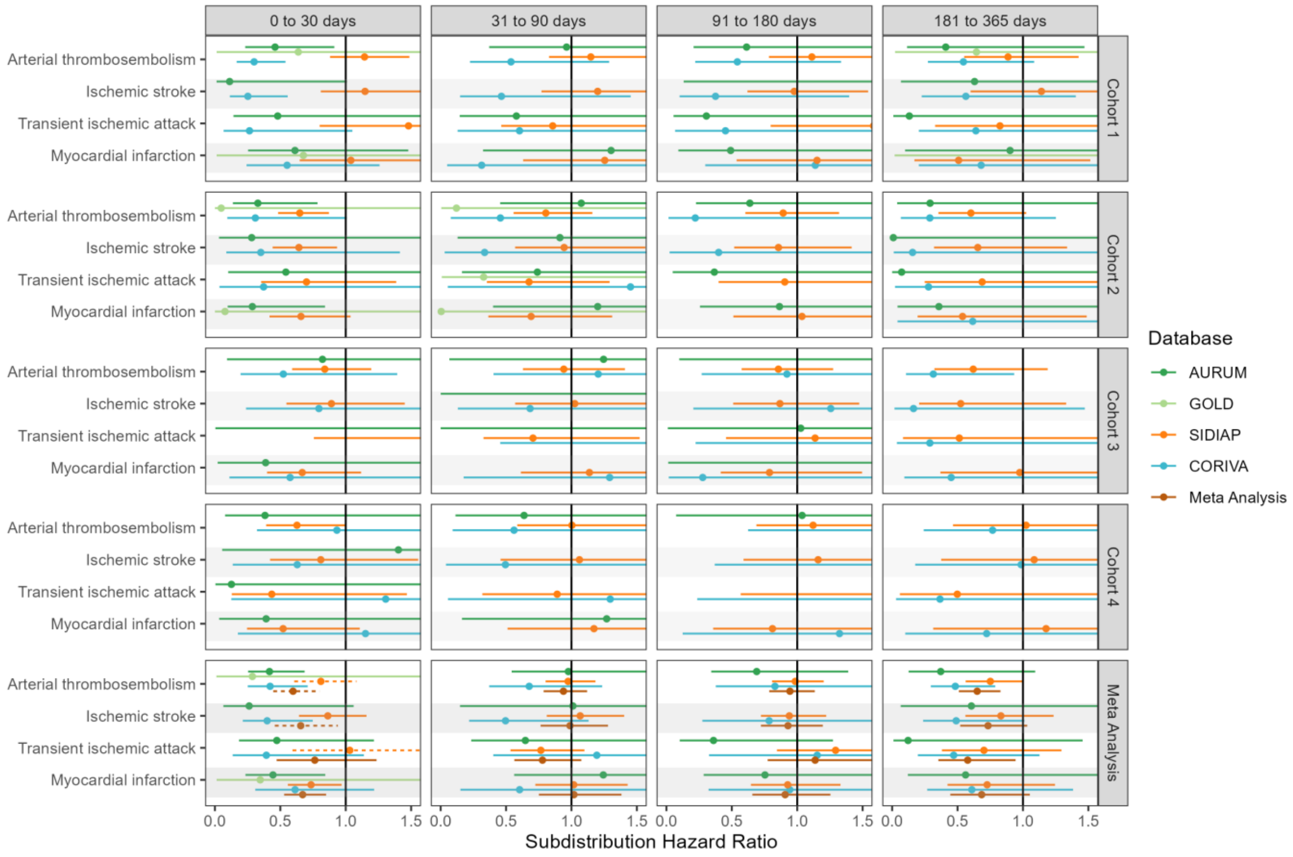

**Figure S32: Forest plots for vaccine effectiveness (BNT162b2 vaccine) on preventing arterial thromboembolism complications,** for each cohort and database (meta-analysis estimates across cohorts in the last panel).Follow-up ends at first vaccine dose after index date. Dashed line represents a level of hetereogeneity I2 > 0.4.

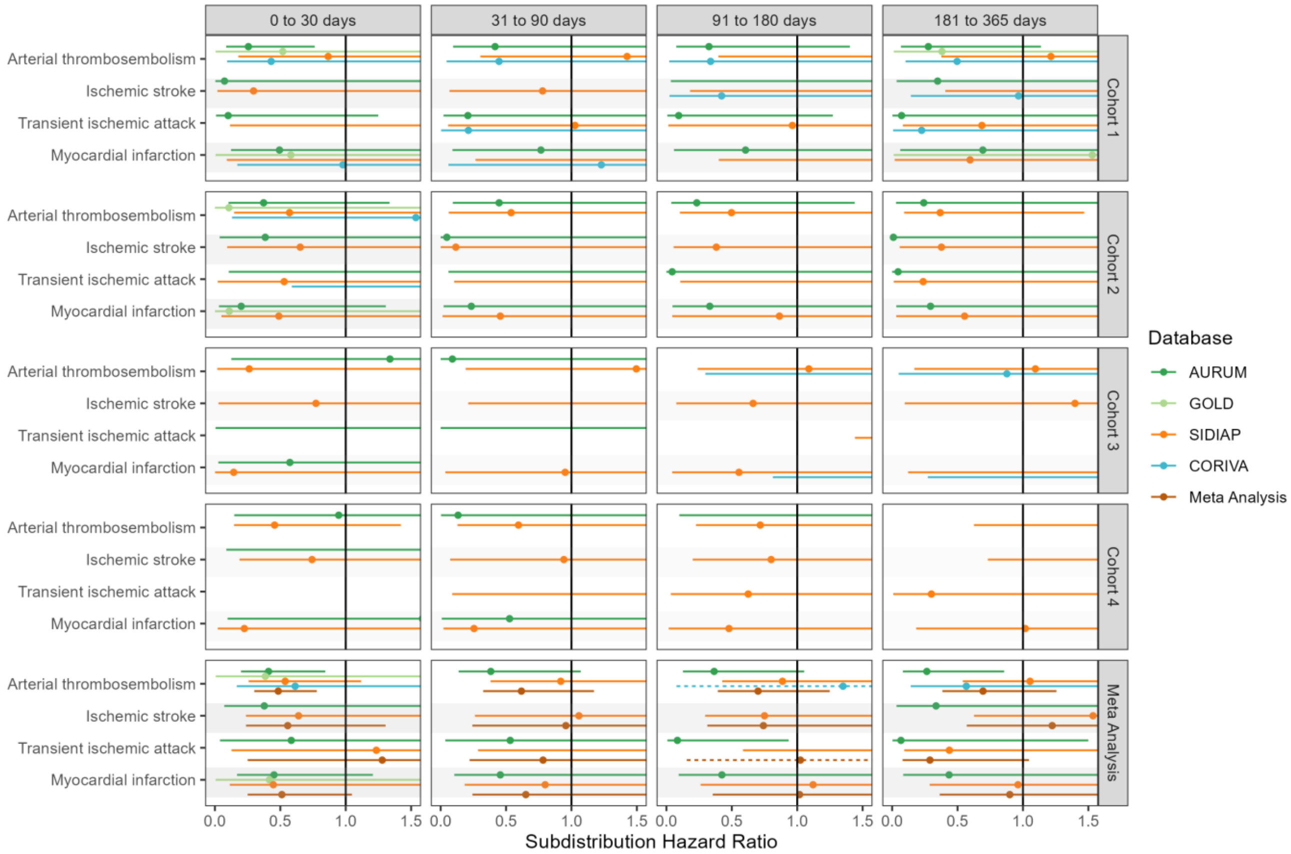

**Figure S33: Forest plots for vaccine effectiveness (BNT162b2 vaccine) on preventing arterial thromboembolism complications,** for each cohort and database (meta-analysis estimates across cohorts in the last panel).Only first outcome after COVID-19 captured. Dashed line represents a level of hetereogeneity I2 > 0.4.

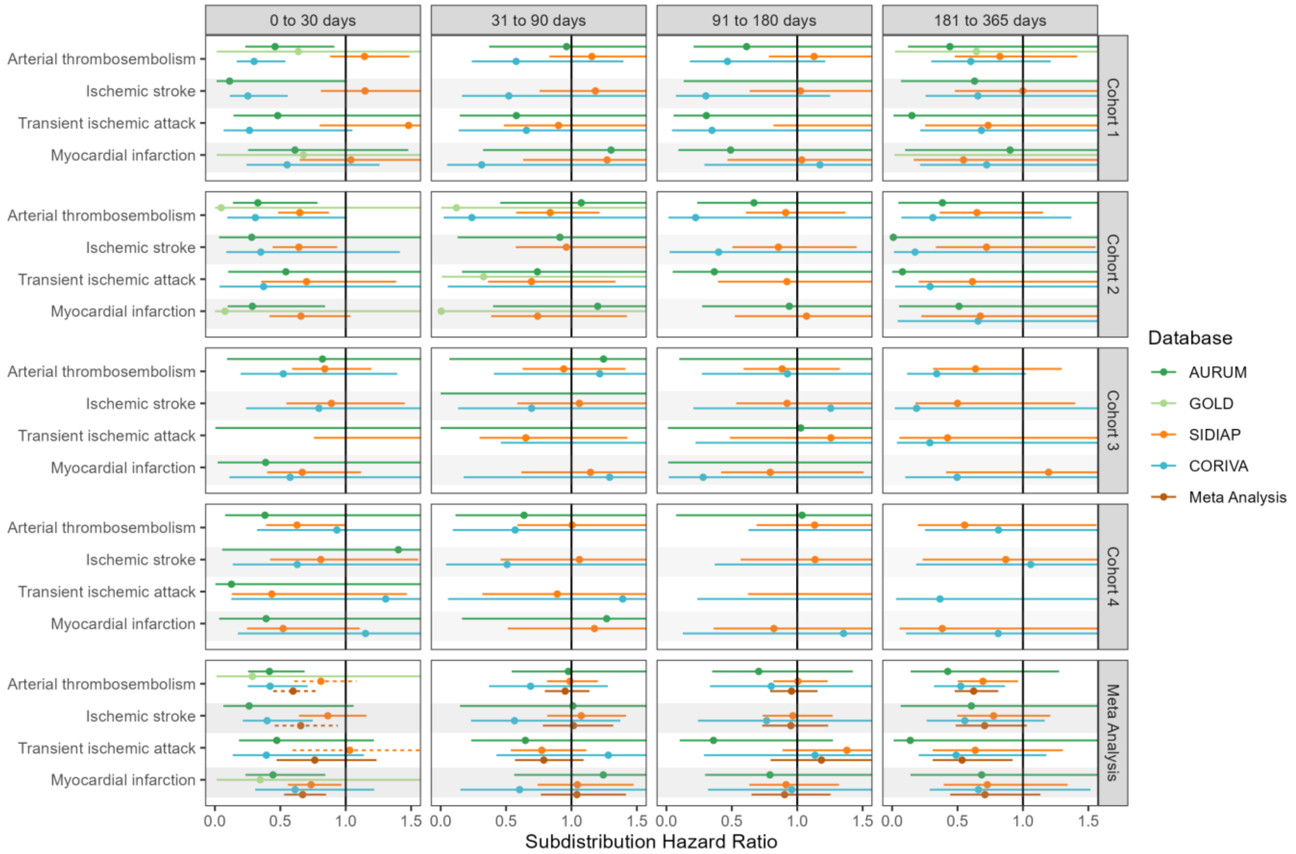

**Figure S34: Forest plots for vaccine effectiveness (any COVID-19 vaccine) on preventing cardiac diseases and hemorrhagic stroke,** for each cohort and database (meta-analysis estimates across cohorts in the last panel). Dashed line represents a level of hetereogeneity I2 > 0.4.

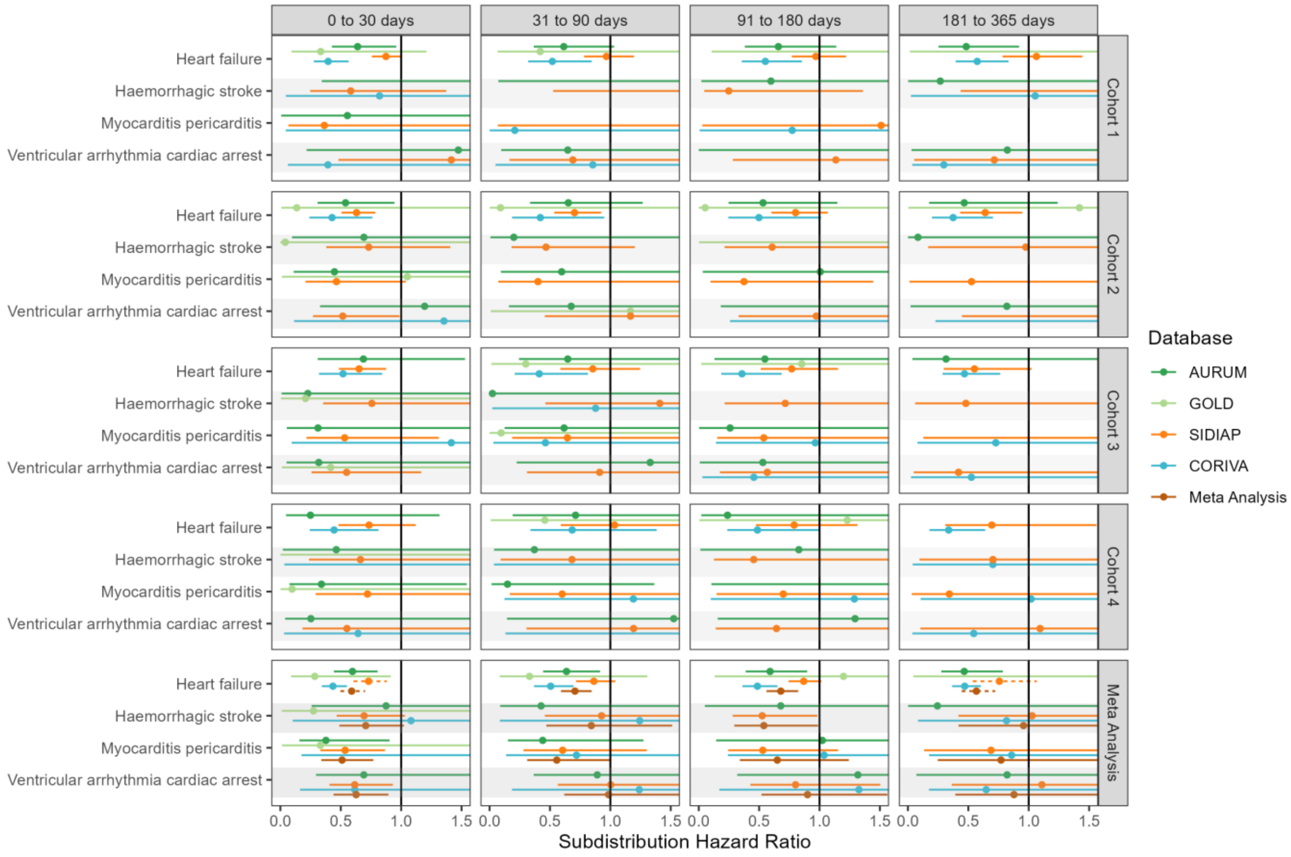

**Figure S35: Forest plots for vaccine effectiveness (any COVID-19 vaccine) on preventing cardiac diseases and hemorrhagic stroke,** for each cohort and database (meta-analysis estimates across cohorts in the last panel).Follow-up ends at first vaccine dose after index date. Dashed line represents a level of hetereogeneity I2 > 0.4.

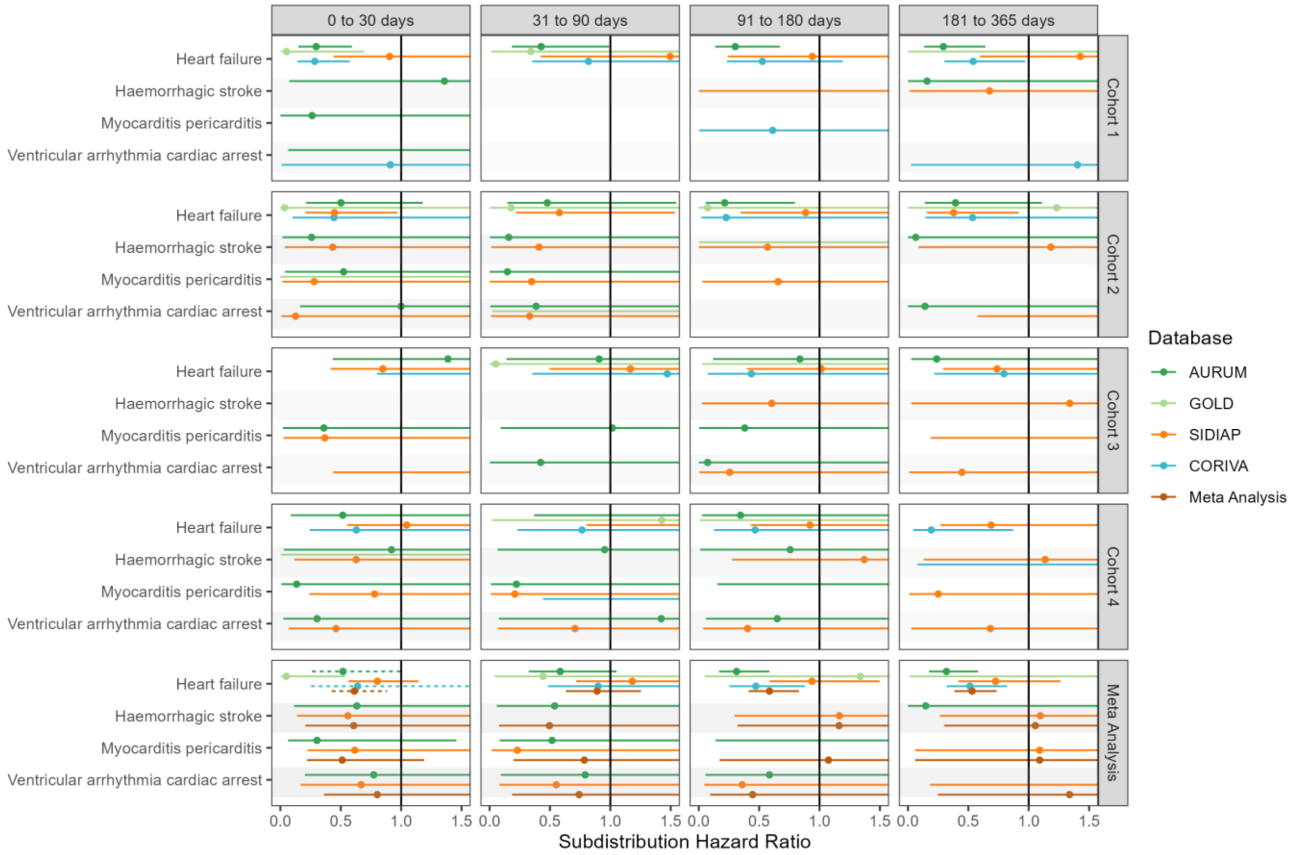

**Figure S36: Forest plots for vaccine effectiveness (any COVID-19 vaccine) on preventing cardiac diseases and hemorrhagic stroke,** for each cohort and database (meta-analysis estimates across cohorts in the last panel).Only first outcome after COVID-19 captured. Dashed line represents a level of hetereogeneity I2 > 0.4.

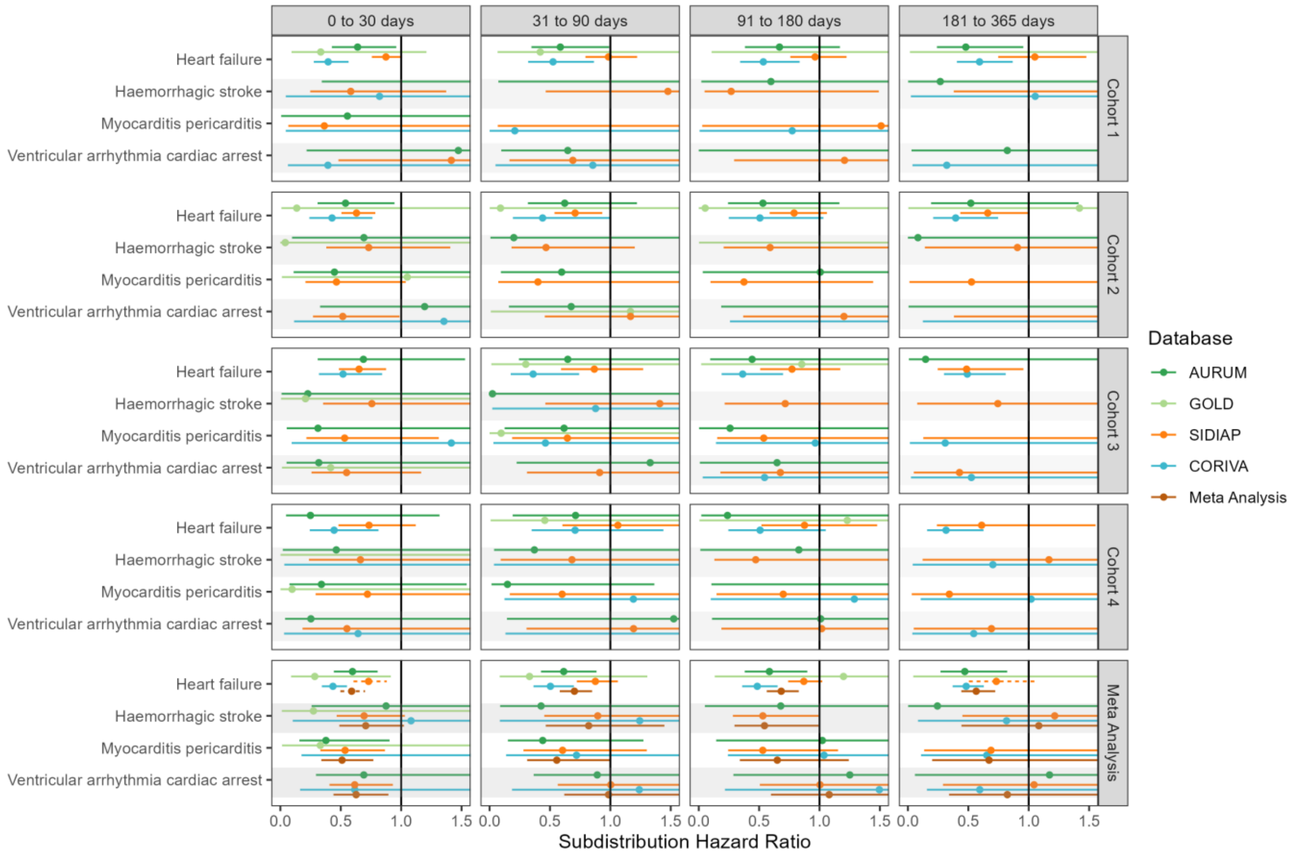

**Figure S37: Forest plots for vaccine effectiveness (ChAdOx1 vaccine) on preventing cardiac diseases and hemorrhagic stroke,** for each cohort and database (meta-analysis estimates across cohorts in the last panel). Dashed line represents a level of hetereogeneity I2 > 0.4.

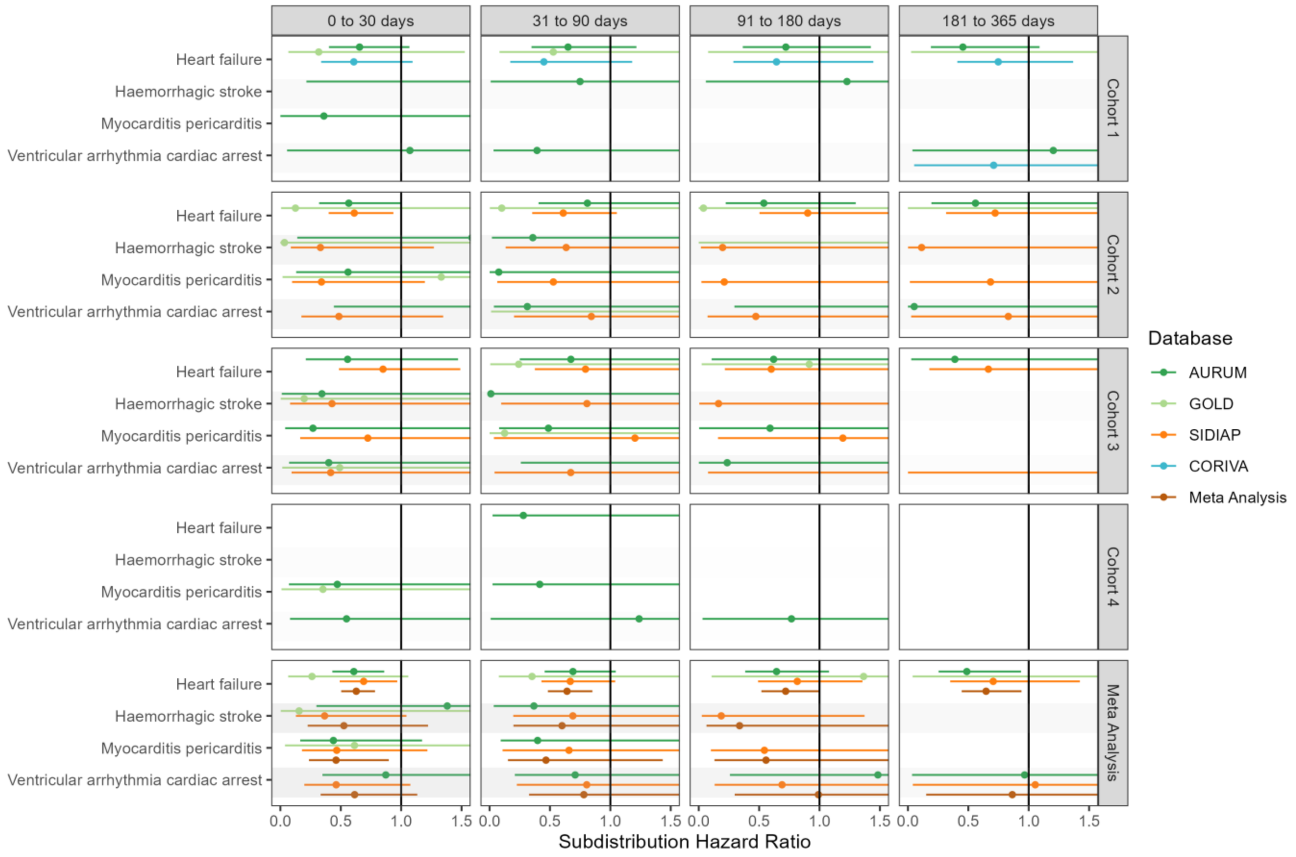

**Figure S38: Forest plots for vaccine effectiveness (ChAdOx1 vaccine) on preventing cardiac diseases and hemorrhagic stroke,** for each cohort and database (meta-analysis estimates across cohorts in the last panel).Follow-up ends at first vaccine dose after index date. Dashed line represents a level of hetereogeneity I2 > 0.4.

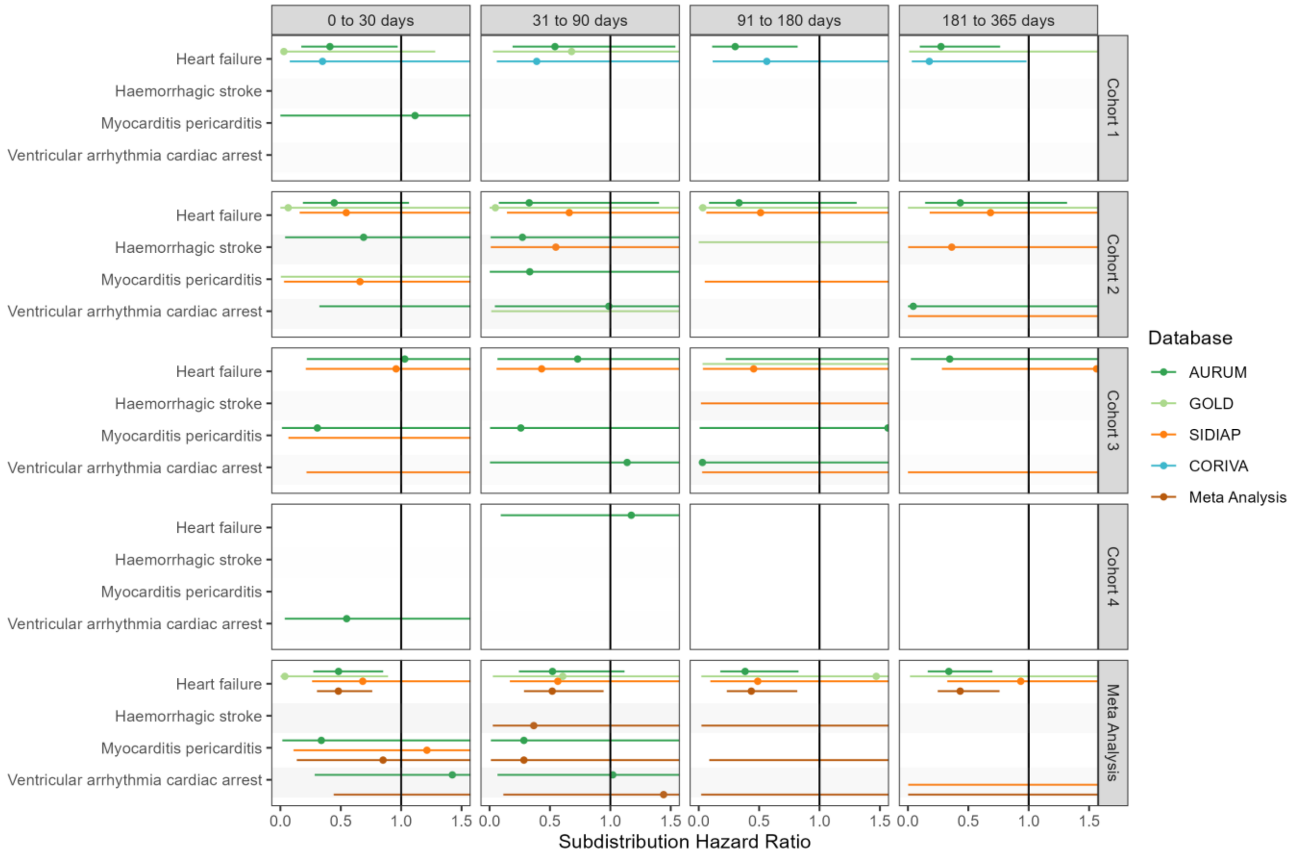

**Figure S39: Forest plots for vaccine effectiveness (ChAdOx1 vaccine) on preventing cardiac diseases and hemorrhagic stroke,** for each cohort and database (meta-analysis estimates across cohorts in the last panel).Only first outcome after COVID-19 captured. Dashed line represents a level of hetereogeneity I2 > 0.4.

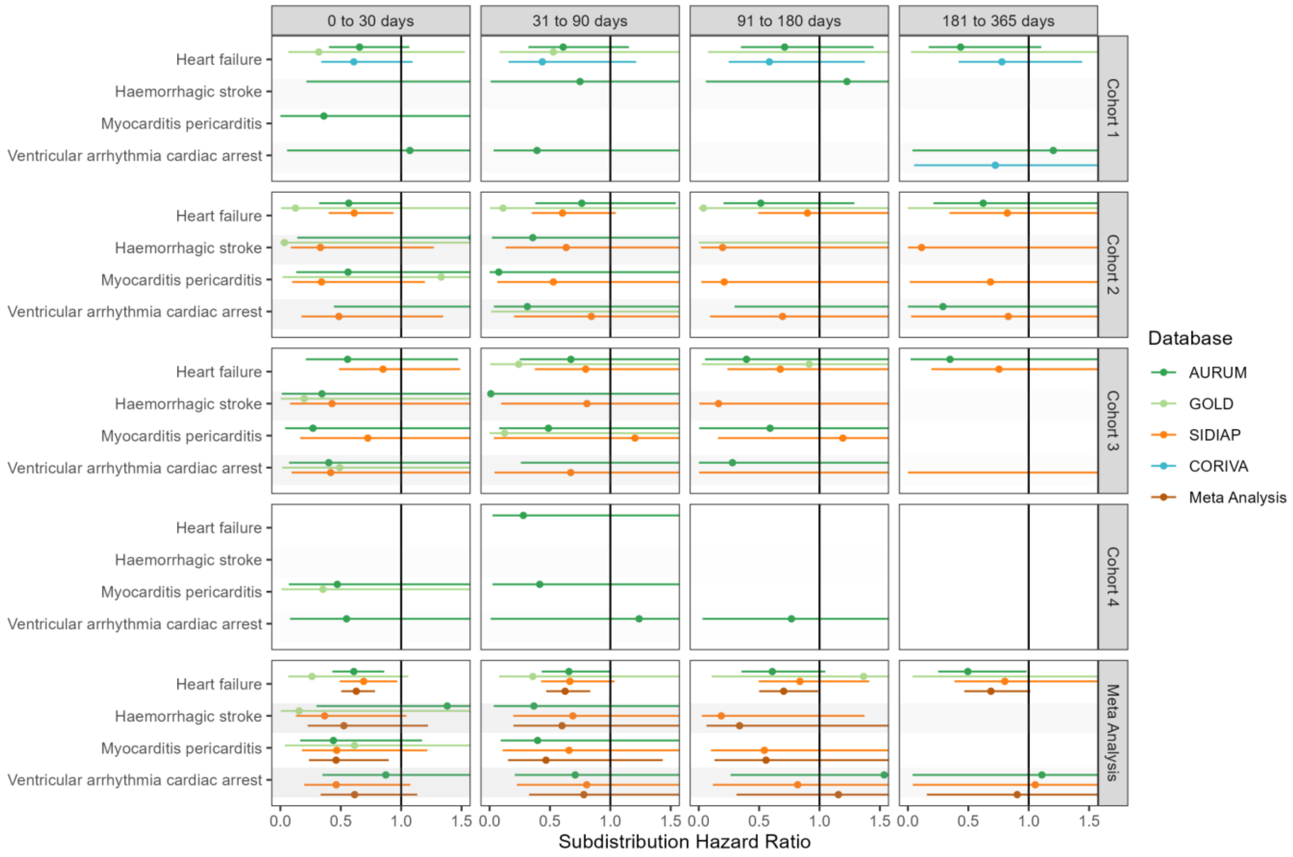

**Figure S40: Forest plots for vaccine effectiveness (BNT162b2 vaccine) on preventing cardiac diseases and hemorrhagic stroke,** for each cohort and database (meta-analysis estimates across cohorts in the last panel). Dashed line represents a level of hetereogeneity I2 > 0.4.

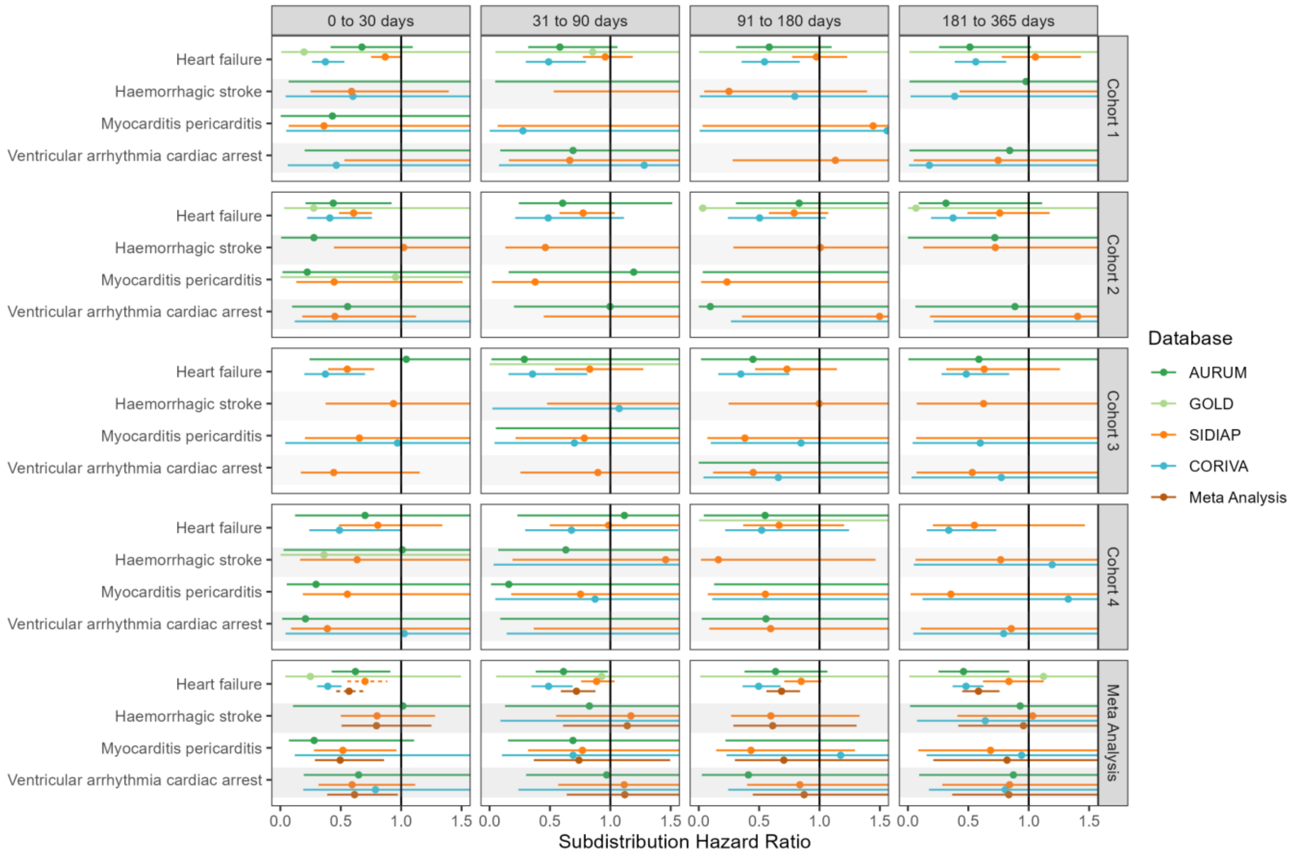

**Figure S41: Forest plots for vaccine effectiveness (BNT162b2 vaccine) on preventing cardiac diseases and hemorrhagic stroke,** for each cohort and database (meta-analysis estimates across cohorts in the last panel).Follow-up ends at first vaccine dose after index date. Dashed line represents a level of hetereogeneity I2 > 0.4.

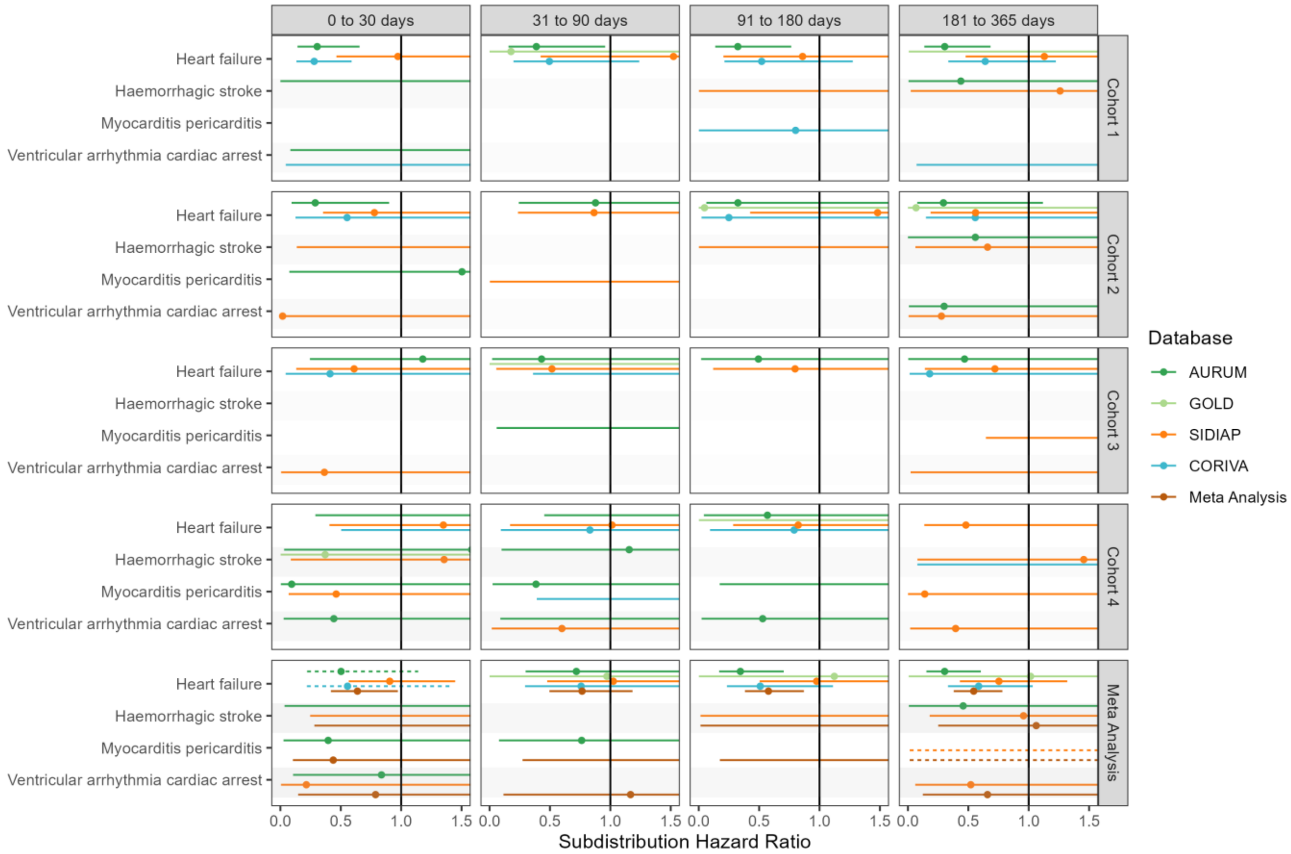

**Figure S42: Forest plots for vaccine effectiveness (BNT162b2 vaccine) on preventing cardiac diseases and hemorrhagic stroke,** for each cohort and database (meta-analysis estimates across cohorts in the last panel).Only first outcome after COVID-19 captured. Dashed line represents a level of hetereogeneity I2 > 0.4.

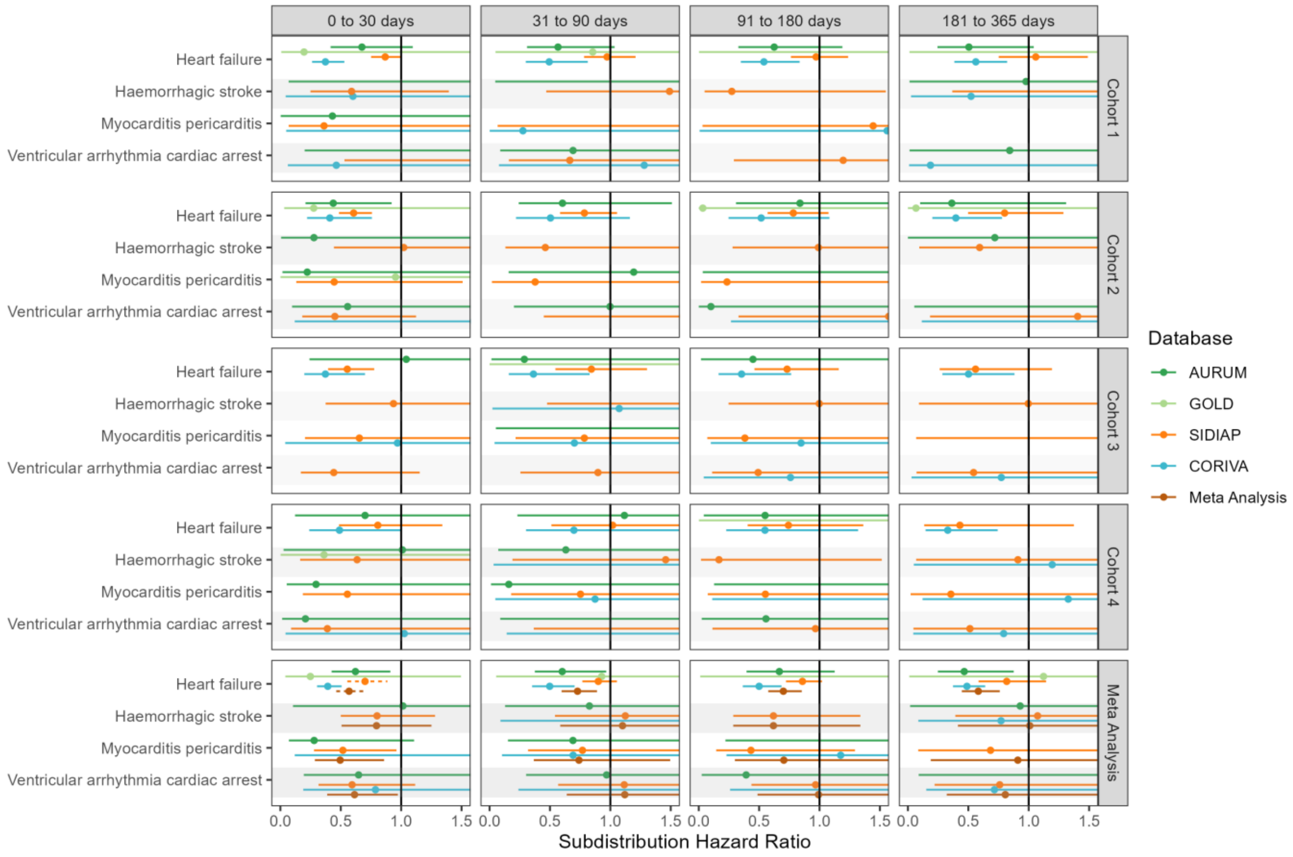

**Figure S43: Forest plots for comparative effectiveness,** meta-analysis across cohorts and databases. Follow-up ends at first vaccine dose after index date. Dashed line represents a level of hetereogeneity I2 > 0.4.

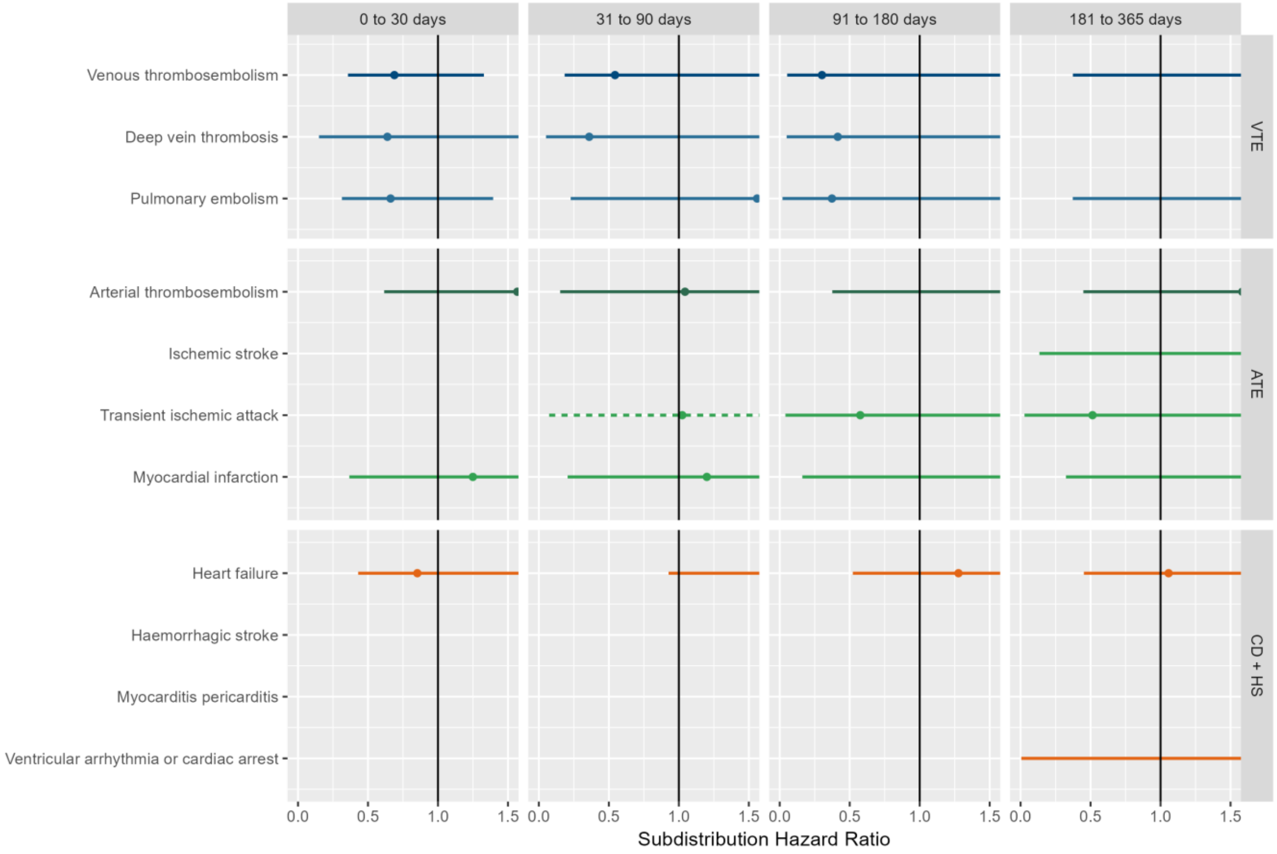

VTE = Venous thromboembolism, ATE = Arterial thromboembolism, CD + HS = Cardiac diseases and Hemorrhagic stroke

**Figure S44: Forest plots for comparative effectiveness,** meta-analysis across cohorts and databases. Only first outcome after COVID-19 captured. Dashed line represents a level of hetereogeneity I2 > 0.4.

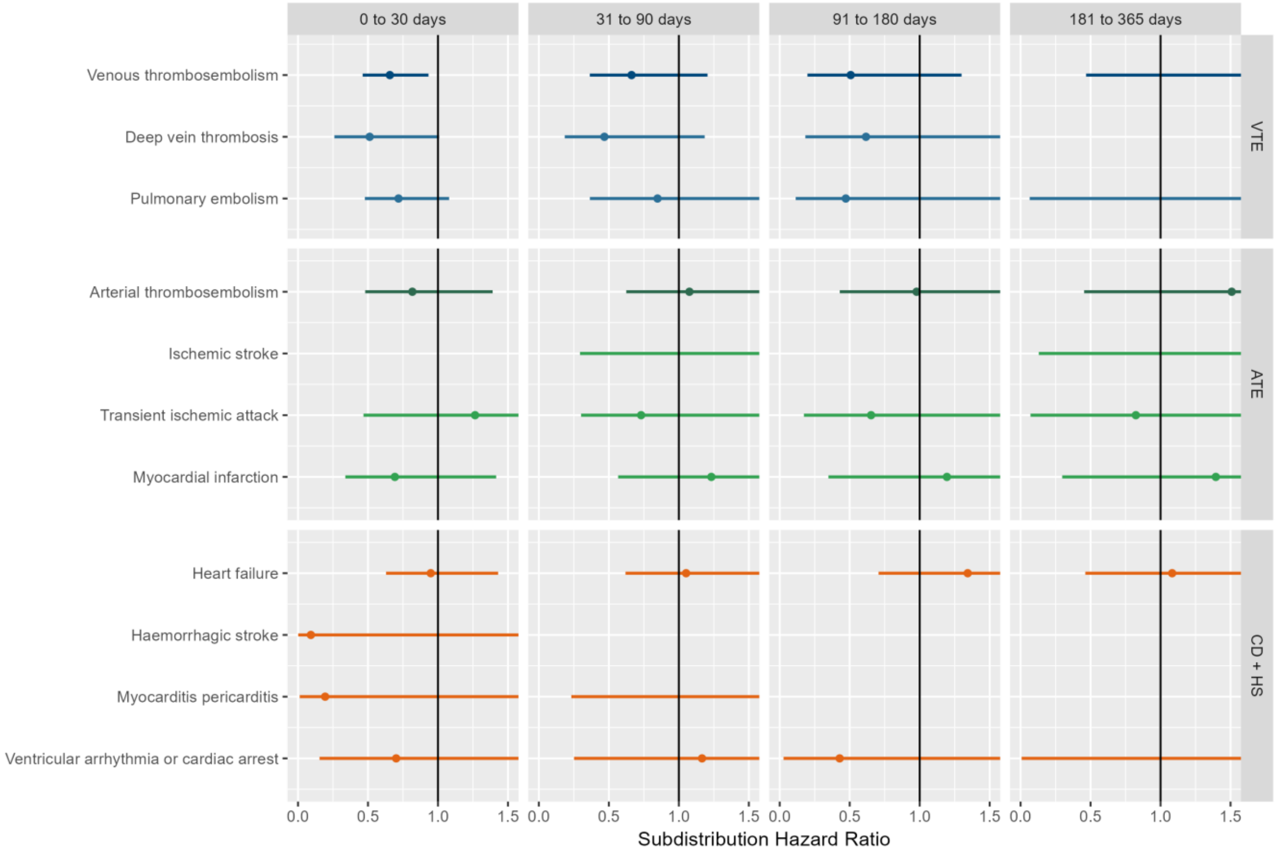

VTE = Venous thromboembolism, ATE = Arterial thromboembolism, CD + HS = Cardiac diseases and Hemorrhagic stroke

**Figure S45: Forest plots for comparative effectiveness on preventing venous thromboembolism complications,** for each cohort and database (meta-analysis estimates across cohorts in the last panel). Dashed line represents a level of hetereogeneity I2 > 0.4.

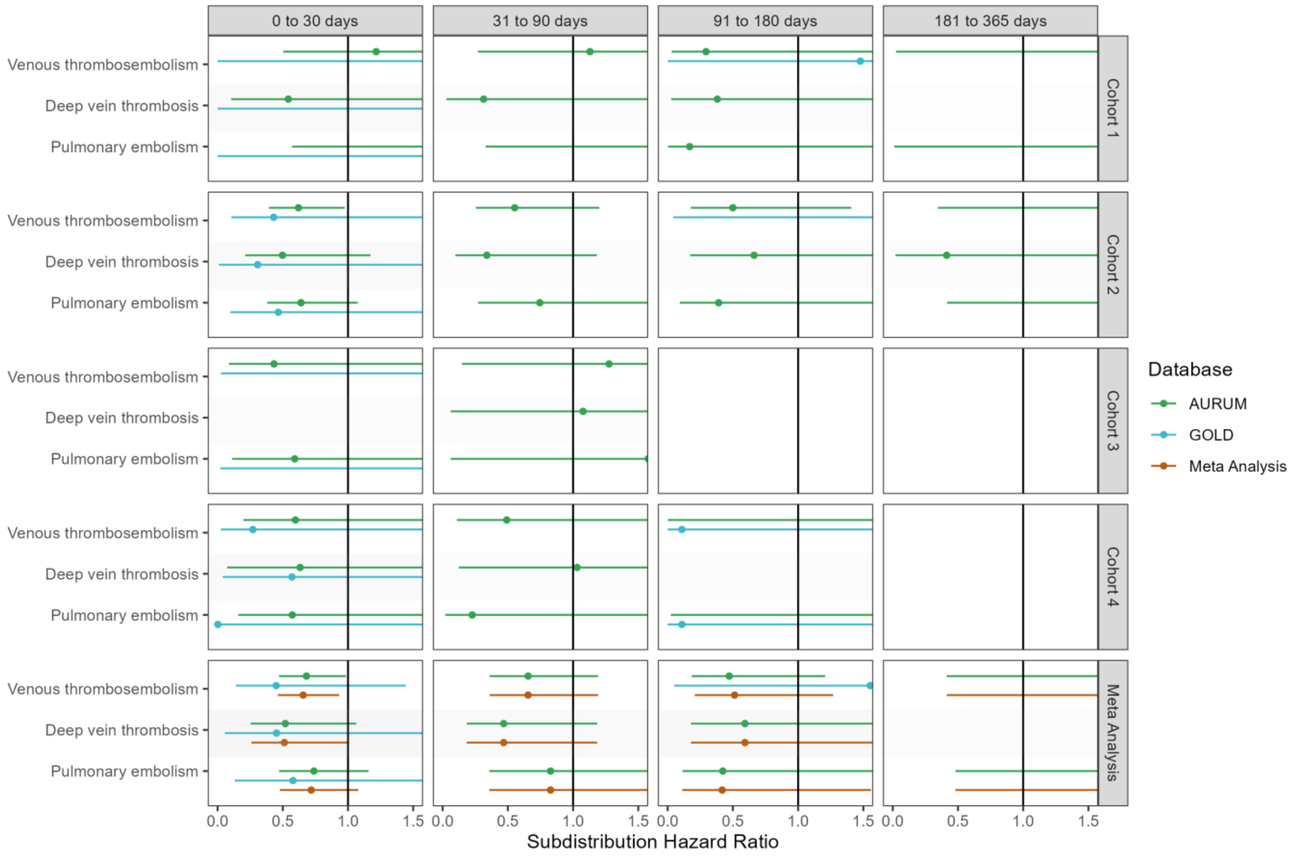

**Figure S46: Forest plots for comparative effectiveness on preventing venous thromboembolism complications,** for each cohort and database (meta-analysis estimates across cohorts in the last panel).Follow-up ends at first vaccine dose after index date. Dashed line represents a level of hetereogeneity I2 > 0.4.

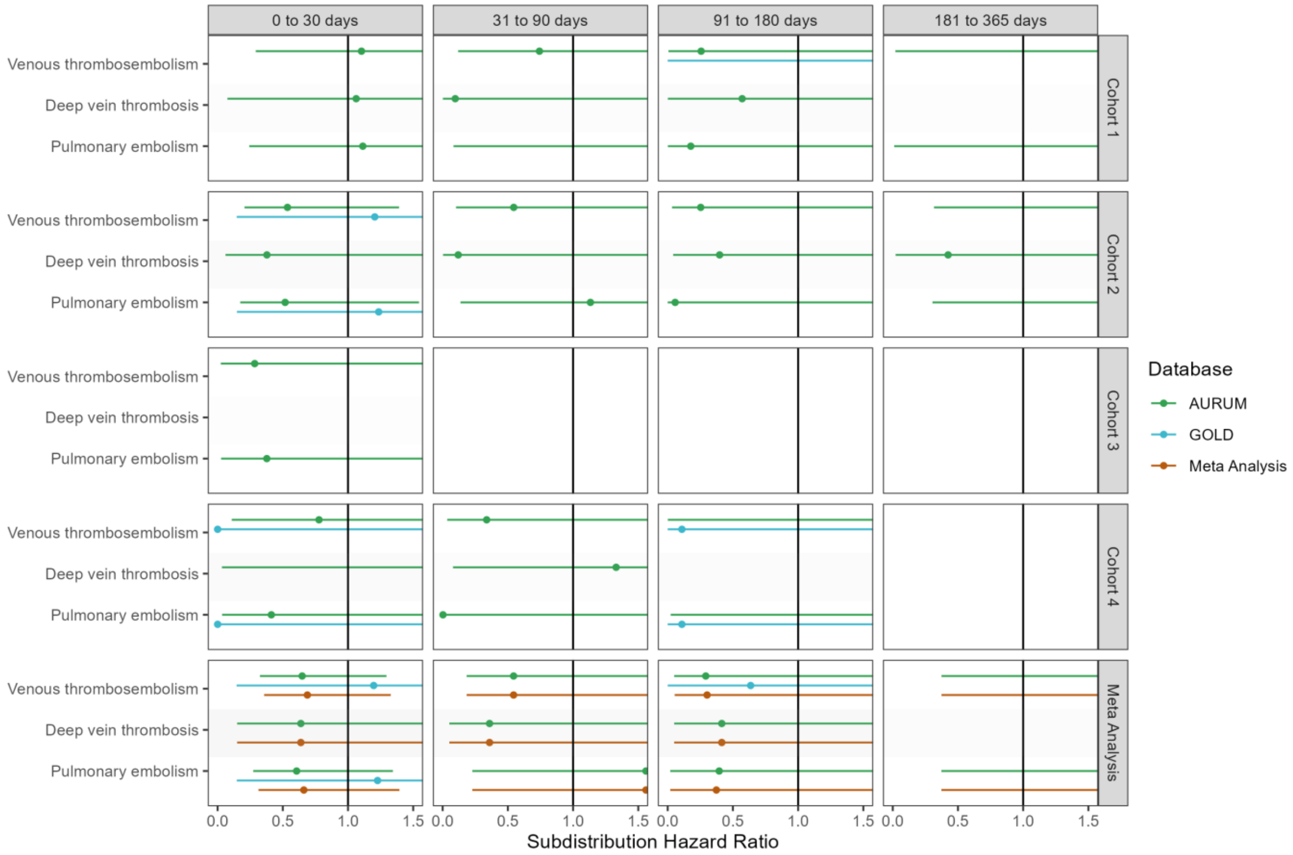

**Figure S47: Forest plots for comparative effectiveness on preventing venous thromboembolism complications,** for each cohort and database (meta-analysis estimates across cohorts in the last panel).Only first outcome after COVID-19 captured. Dashed line represents a level of hetereogeneity I2 > 0.4.

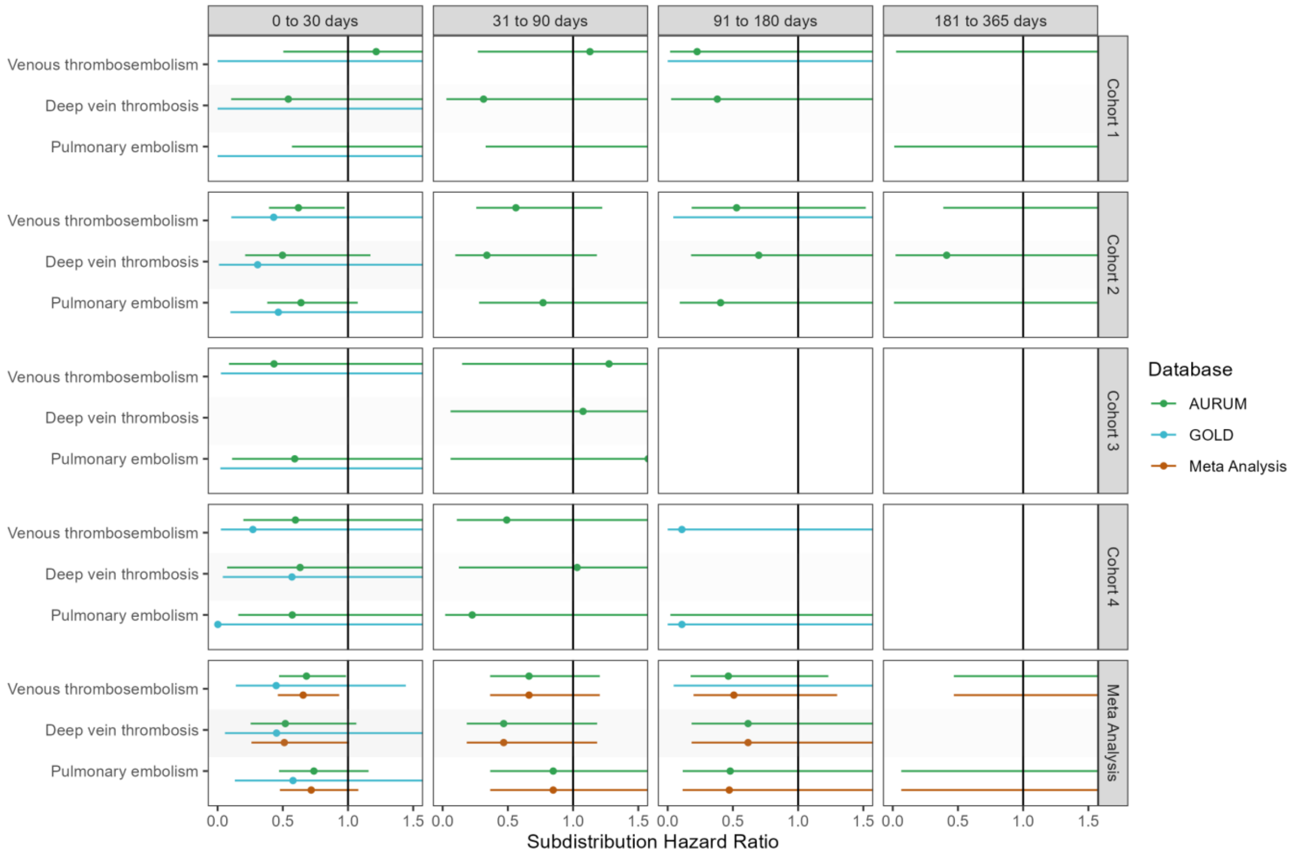

**Figure S48: Forest plots for comparative effectiveness on preventing arterial thromboembolism complications,** for each cohort and database (meta-analysis estimates across cohorts in the last panel). Dashed line represents a level of hetereogeneity I2 > 0.4.

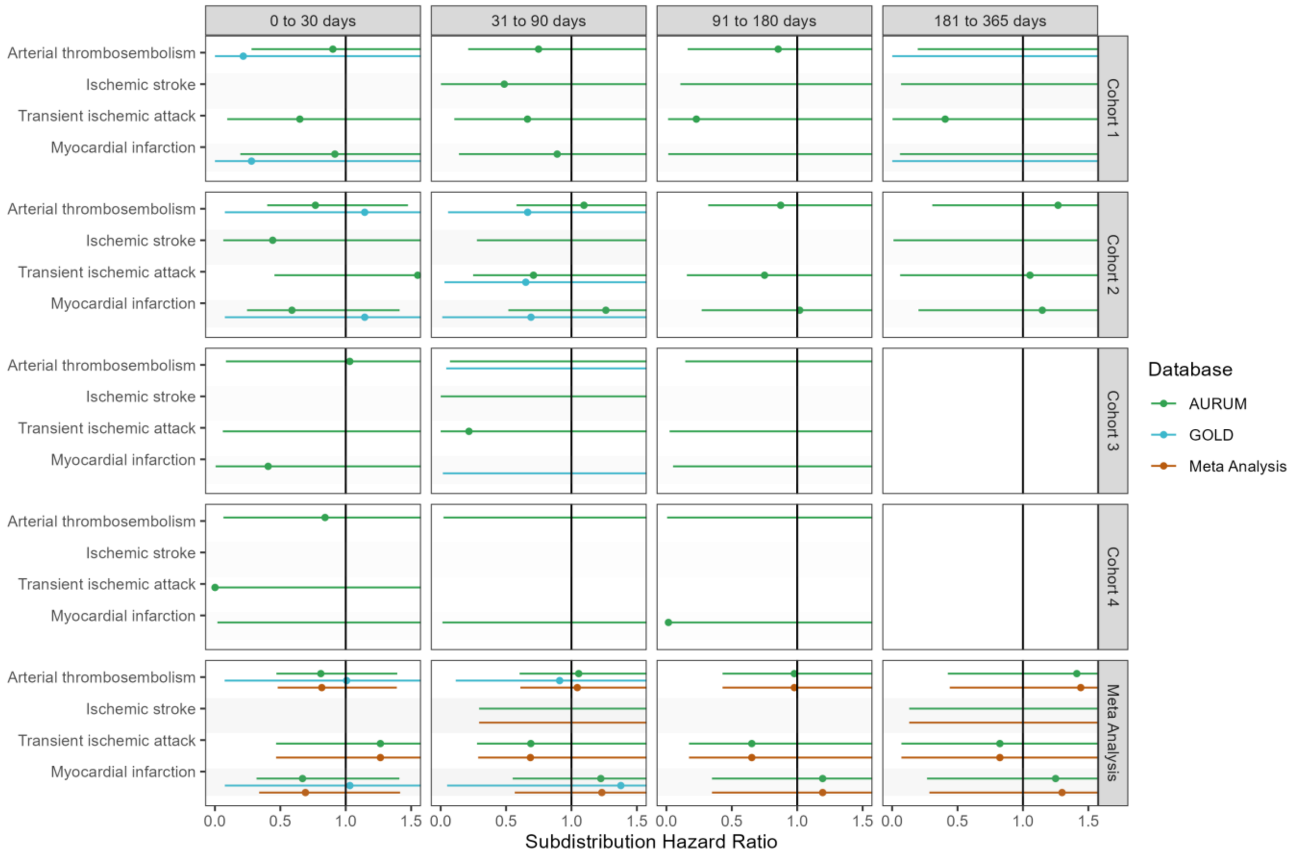

**Figure S49: Forest plots for comparative effectiveness on preventing arterial thromboembolism complications,** for each cohort and database (meta-analysis estimates across cohorts in the last panel).Follow-up ends at first vaccine dose after index date. Dashed line represents a level of hetereogeneity I2 > 0.4.

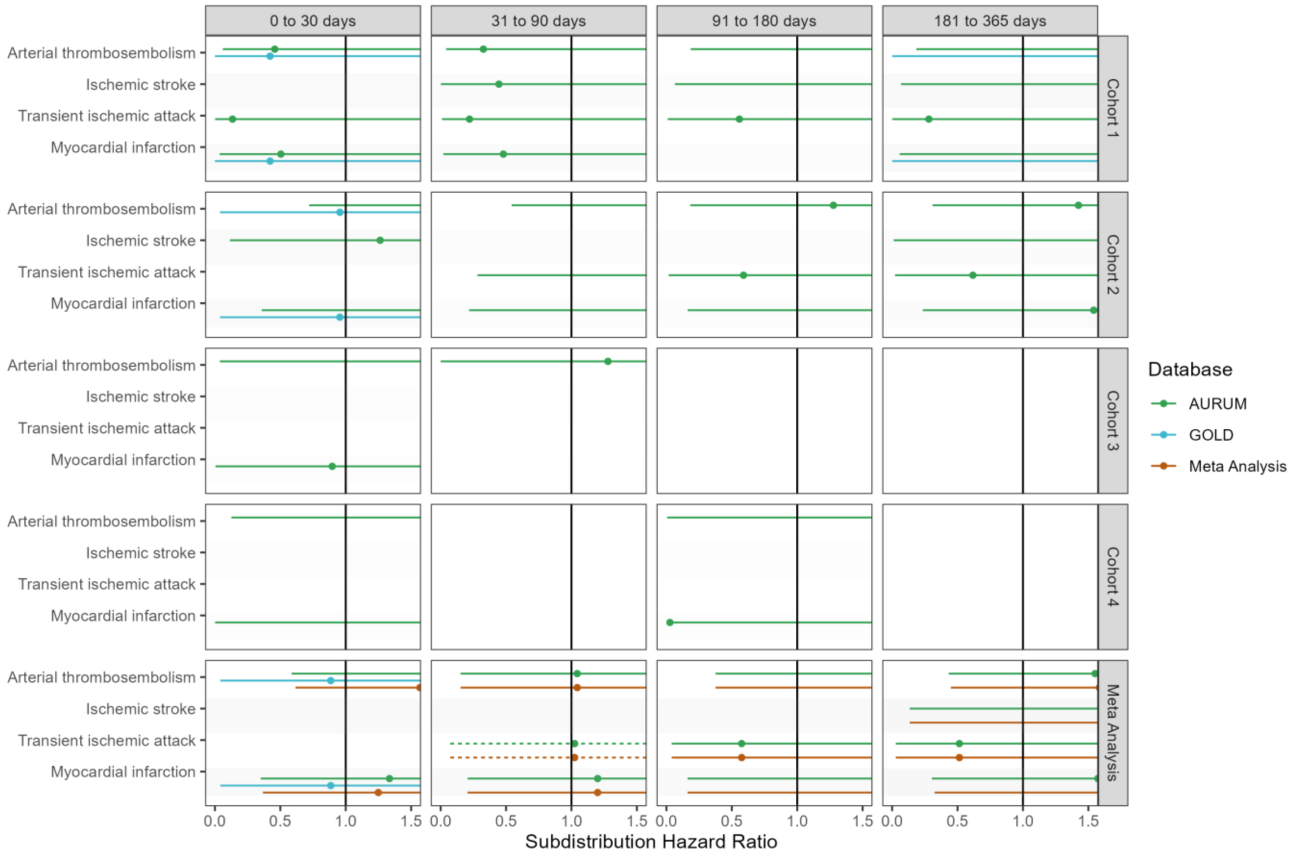

**Figure S50: Forest plots for comparative effectiveness on preventing arterial thromboembolism complications,** for each cohort and database (meta-analysis estimates across cohorts in the last panel).Only first outcome after COVID-19 captured. Dashed line represents a level of hetereogeneity I2 > 0.4.

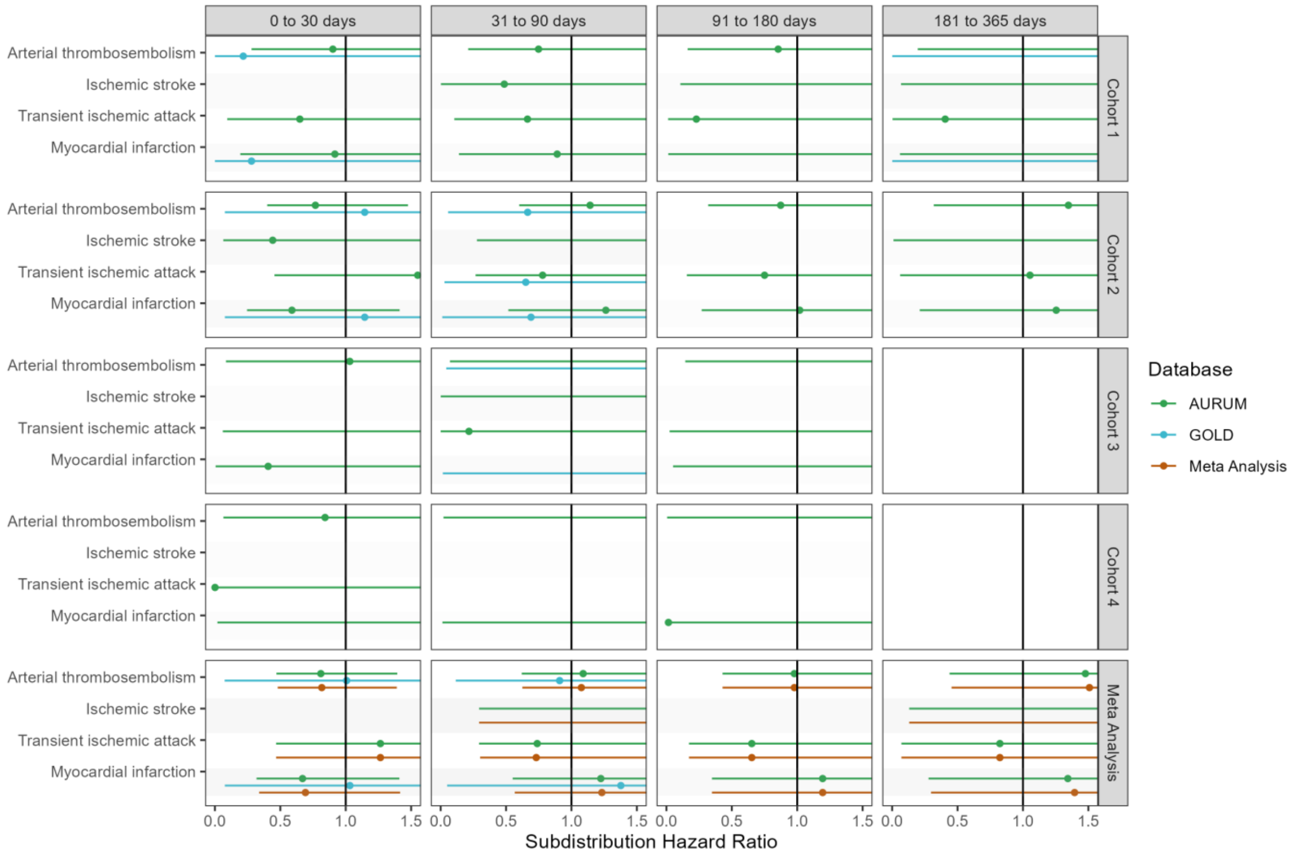

**Figure S51: Forest plots for comparative effectiveness on preventing cardiac diseases and hemorrhagic stroke,** for each cohort and database (meta-analysis estimates across cohorts in the last panel). Dashed line represents a level of hetereogeneity I2 > 0.4.

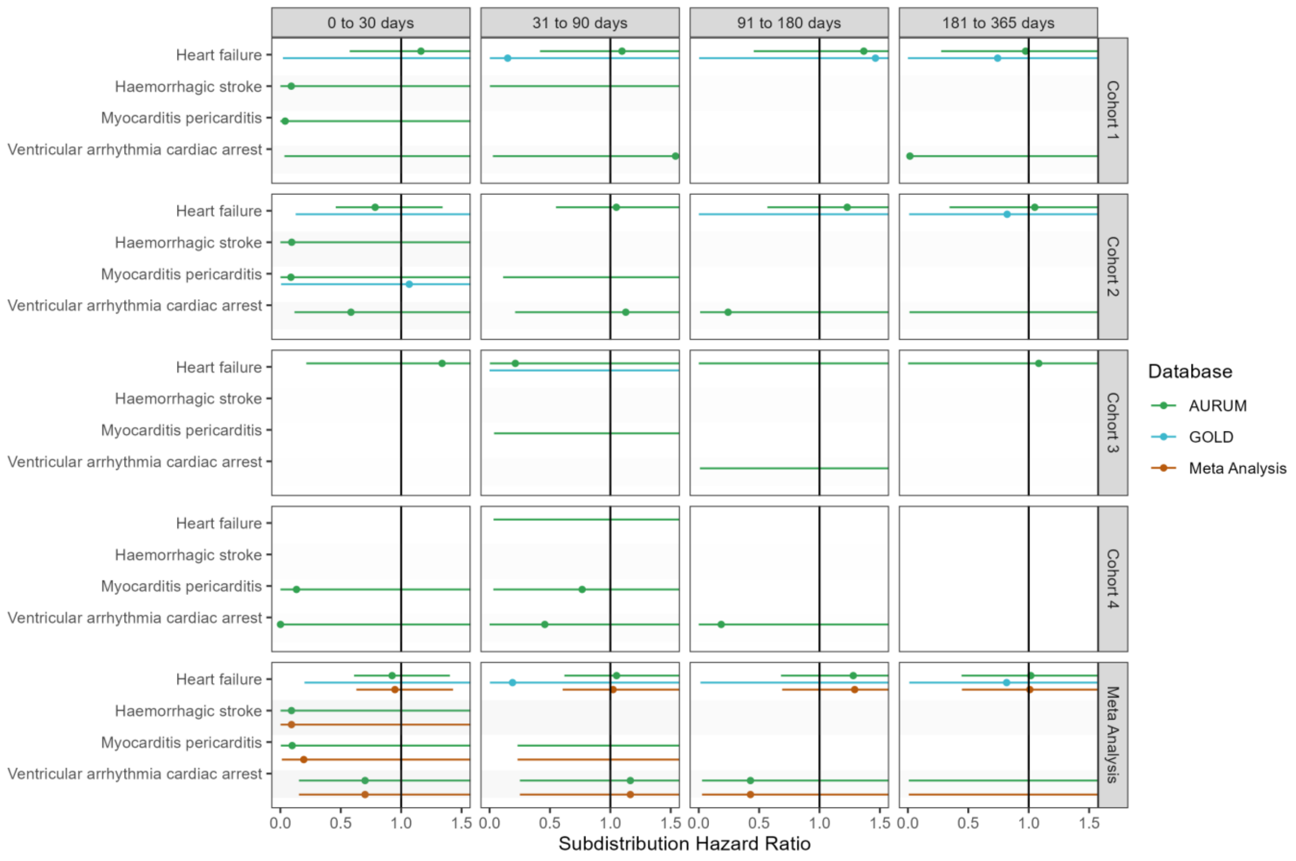

**Figure S52: Forest plots for comparative effectiveness on preventing cardiac diseases and hemorrhagic stroke,** for each cohort and database (meta-analysis estimates across cohorts in the last panel).Follow-up ends at first vaccine dose after index date. Dashed line represents a level of hetereogeneity I2 > 0.4.

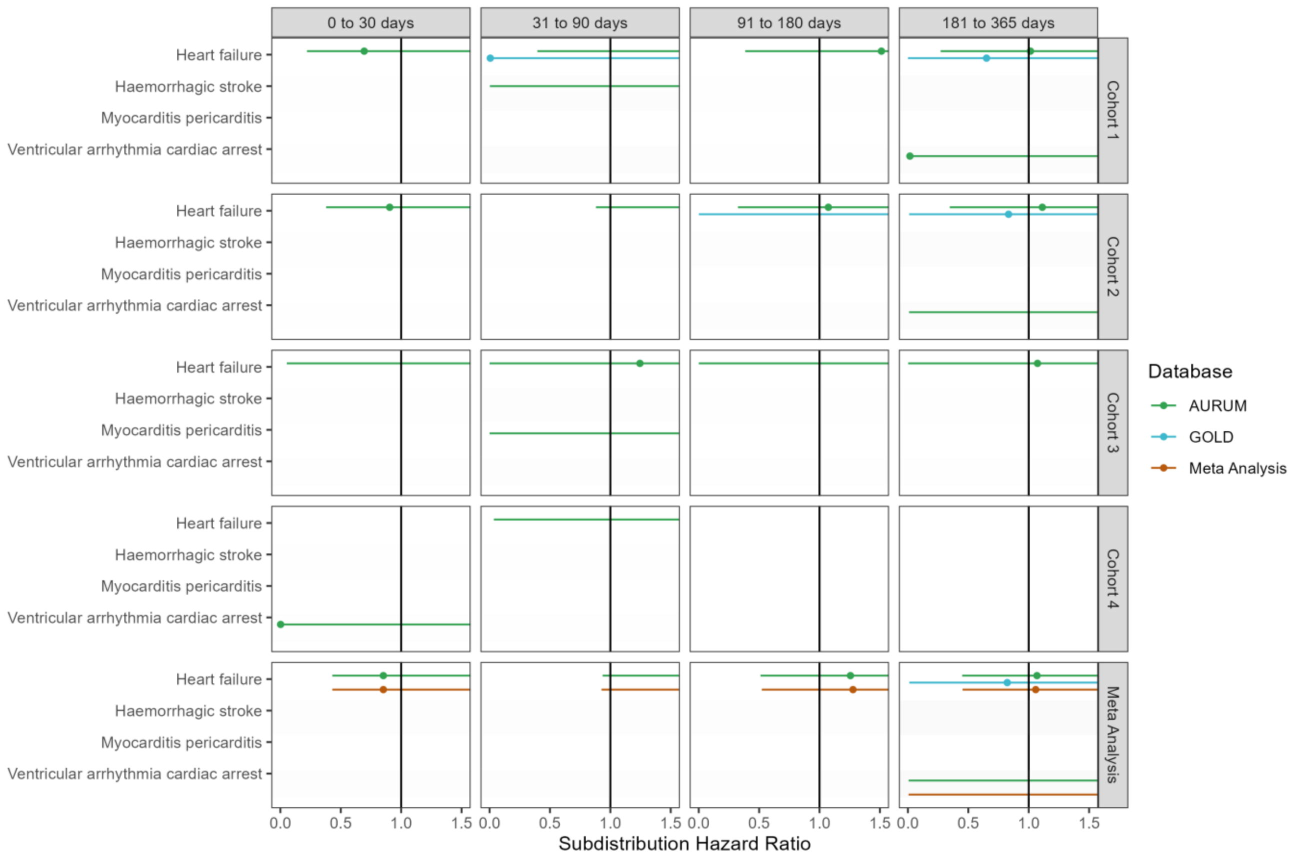

**Figure S53: Forest plots for comparative effectiveness on preventing cardiac diseases and hemorrhagic stroke,** for each cohort and database (meta-analysis estimates across cohorts in the last panel).Only first outcome after COVID-19 captured. Dashed line represents a level of hetereogeneity I2 > 0.4.

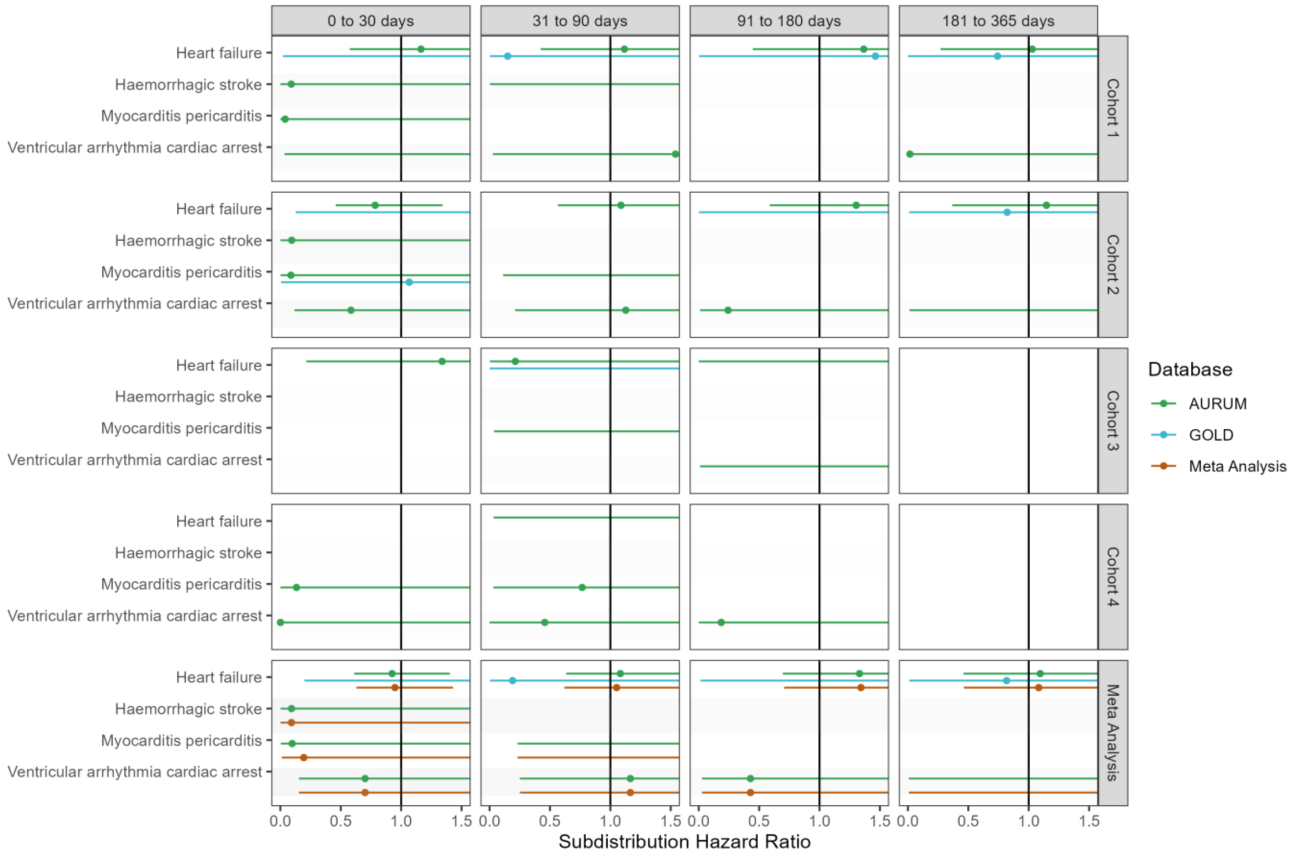
